# Current state of the evidence on community treatments for people with complex emotional needs: a scoping review

**DOI:** 10.1101/2021.12.07.21267399

**Authors:** Sarah Ledden, Luke Sheridan Rains, Merle Schlief, Phoebe Barnett, Brian Chi Fung Ching, Brendan Hallam, Mia Maria Günak, Thomas Steare, Jennie Parker, Sarah Labovitch, Sian Oram, Steve Pilling, Sonia Johnson, CEN Mental Health Policy Research Group

## Abstract

**Background:** Improving the quality of care in community settings for people with ‘Complex Emotional Needs’ (CEN - our preferred working term for services for people with a “personality disorder” diagnosis or comparable needs) is recognised internationally as a priority. Plans to improve care should be rooted as far as possible in evidence. We aimed to take stock of the current state of such evidence, and identify significant gaps through a scoping review of published investigations of outcomes of community-based psychosocial interventions designed for CEN.

**Methods:** We conducted a scoping review with systematic searches. We searched six bibliographic databases, including forward and backward citation searching, and reference searching of relevant systematic reviews. We included studies using quantitative methods to test for effects on any clinical, social, and functioning outcomes from community-based interventions for people with CEN.

**Results:** We included 226 papers in all (209 studies). Little relevant literature was published before 2000. Since then, publications per year and sample sizes have gradually increased, but most studies are relatively small, including many pilot or uncontrolled studies. Most studies focus on symptom and self-harm outcomes of various forms of specialist psychotherapy: most result in outcomes better than from inactive controls and similar to other specialist psychotherapies. We found large evidence gaps.

Adaptation and testing of therapies for significant groups (e.g. people with comorbid psychosis, bipolar disorder, post-traumatic stress disorder or substance misuse; older and younger groups; parents) have for the most part only reached a feasibility testing stage. We found little evidence regarding interventions to improve social aspects of people’s lives, peer support or ways of designing effective services.

**Conclusions:** Compared with other longer term mental health problems that significantly impair functioning, the evidence base on how to provide high quality care for people with CEN is very limited. There is good evidence that people with CEN can be effectively helped when specialist therapies are available and they are able to engage with them. However, a much more methodologically robust and substantial literature addressing a much wider range of research questions is urgently needed to optimise treatment and support across this group.

## Introduction

People who have received a diagnosis of “personality disorder” are reported to experience a range of difficulties with social functioning, mental and physical health [1, 2]. Substantial economic burdens are associated, especially due to treatment costs and productivity losses [3, 4]. Historically a “personality disorder” diagnosis was seen as indicating a lack of treatability [5]. More recently, there has been greater recognition of the needs for support and the provision of effective treatment for this group, and improving care has been identified as a priority in a variety of countries [6–9].

A heavy burden of stigma is associated with a “personality disorder” diagnosis, with negative views and discriminatory behaviour from some health professionals having especially immediate impacts [10–14]. We are sympathetic to the critique that the therapeutic nihilism and stigma accompanying a “personality disorder” diagnosis, and the lack of progress in delivering care that consistently helps rather than harms, are such that this diagnostic label - also criticised on grounds of validity - is now best left behind. Further work is needed on assessing and describing the difficulties that people who may receive this diagnostic label experience in more useful and acceptable ways: pending this, we prefer the term complex emotional needs (CEN) as a working description of the difficulties experienced by people who may receive a “personality disorder” diagnosis, and therefore use it as our headline description in this paper, as in our other publications on this topic. We are guided especially by members of our research team who have relevant lived experience in making this choice. However, the literature we have reviewed for the most part is based on “personality disorder” diagnoses of various types: thus, below we use this term where it is used in the papers included in our review.

Mental health services and mental health research are widely acknowledged not to have achieved parity in terms of resources and status with physical health care, and services for people with a “personality disorder” diagnosis are doubly disadvantaged as they appear to significantly lag behind services for people with other long-term mental health conditions [6, 15–17]. Recurrently reported difficulties include large variations in accessibility and quality of services, difficulty accessing specialist “personality disorder” services and lack of therapeutic interventions outside them, a tendency for interventions to focus narrowly on self-harm rather than on the broader range of psychological and social outcomes that service users and carers identify as important, lack of focus on trauma experiences even though are very frequent, and exclusion from care of people with common comorbidities such as substance misuse or bipolar disorder, or at the younger or older end of the age range [10, 17–20].

Internationally, service user activists, professional bodies and policy makers have advocated for better quality services for people with CEN [15–17]. Ideally, service improvement should be rooted in evidence-based practice [21, 22]. A number of systematic reviews have reported on the trial literature on psychological interventions for people with a borderline personality disorder, including Dialectal Behaviour Therapy (DBT), Mentalisation Based Therapy (MBT), Cognitive Behavioural Therapy (CBT), and psychodynamic therapies, amongst others [23–25]. Reviews tend to conclude that these specialist treatments are all more effective than treatment as usual in achieving clinical improvements in self-harm and “borderline symptoms”, although no single intervention type has emerged as dominant [26].

However, these relatively narrowly focused systematic reviews have left unanswered a range of questions that are key to improving care holistically for the full spectrum of people who have received a “personality disorder” diagnosis, or have comparable needs [26]. Questions not addressed include how to improve important social outcomes including employment, social inclusion, relationships and parenting, and the impact the interventions have on the trauma histories that are very frequent among this group should be addressed. These previous reviews have also not focused on the needs of important groups, such as older adults and younger people, people with comorbidities such as substance misuse, psychosis or bipolar disorder, and people who may have received “personality disorder” diagnoses other than borderline or emotionally unstable, or who have received multiple diagnoses. The key question of service design, and what kinds of teams and networks of services most effectively meet needs and deliver continuity of care also remains largely unanswered.

Given these crucial gaps in the evidence to underpin improvement of care, our intention in the current scoping paper was to cast the net widely, seeking any quantitative evidence that may have potential as building block for future intervention and service design and research in this area. Our aim was to conduct a scoping review of the evidence on the effectiveness of community-based psychological interventions designed for people with CEN. In order to capture a broad range of relevant evidence, we aimed to include in our searches a broad range of diagnoses and related difficulties, interventions focused not only on self-harm and symptoms but also on social targets, and delivered at team and catchment area as well as individual levels. Observational studies can yield helpful evidence on treatment outcomes in naturalistic settings, sometimes providing pointers to interventions worth researching through randomised trials or allowing questions to be addressed, such as about area-level service design, that are difficult to investigate through trials [27]: we thus aimed also to capture evidence from such designs. We also aimed to identify preliminary investigations of feasibility and reports on adaptations of interventions to new populations or new settings, as these have potential to inform further research and intervention development. Therefore, by considering this broader evidence base, we aim to take stock of what is known so far, highlight important gaps, and inform future research in this area.

## Methods

### Study Design

We conducted a systematic scoping review [28, 29] to map the evidence from studies using a range of quantitative designs on community-based treatments for CEN and to identify gaps in the literature.

### Search Strategy

The current review was part of a programme of work commissioned from the National Institute for Health Research Mental Health Policy Research Unit to inform policy on services for CEN. This programme of work included evidence reviews and studies of stakeholder views and experiences, and was supported by a working group that included people with relevant lived experience of using services and clinicians from a range of disciplines and service contexts.

The programme included four individual systematic reviews, for which we used a single overall search strategy, developed in collaboration with the working group of researchers, clinicians, and people with relevant lived experience. This review follows the PRISMA guidelines [30] and the protocol was prospectively registered on PROSPERO (CRD42019143165). The protocol for the wider programme of work was also registered on PROSPERO (CRD42019131834). The original protocol encompassed a meta-analysis of quantitative data, however, the extent and heterogeneity of important literature led to a decision to conduct such analyses on a more limited subset of data. This will be reported in a separate paper, and meta-syntheses of the qualitative evidence have also been conducted [10, 20, 31].

We conducted a comprehensive search of MEDLINE, Embase, HMIC, Social Policy and Practice, CINAHL and ASSIA, from database inception to December 2019. Search terms included terms relating to CEN, community/outpatient setting, and psychological or psychosocial treatments. An update search was conducted in November 2020. The search strategy was supplemented with a reference search of relevant systematic reviews following the original and updated search. Forward and backward citation searches using Web of Science were also performed for all included papers. No limits were placed on the language or country. Details of the search strategy are available in Appendix 1.

### Study selection

All titles and abstracts were independently screened, with 10% double checked by a second researcher. Studies not meeting inclusion criteria were excluded. Subsequently, full-text articles were screened according to the specific inclusion criteria for this review by two researchers. Unclear cases and disagreements were resolved through discussion with the wider research team.

### Selection criteria

Studies were included if they met the following criteria:

#### Participants

Adults (operationalised as 90% of the sample over 16 years old or mean sample age of 18 or over) in which a majority (>50%) had received a diagnosis of “personality disorder”. In order not to exclude studies in which authors wished to avoid use of this diagnostic term, or which focused on participants who had not received a formal diagnosis, we also ran searches using search terms intended to capture difficulties comparable to those experienced by people with a “personality disorder” diagnosis, including searches for samples presenting with repeated self-harm or suicide attempts, complex trauma or complex post-traumatic stress disorder (PTSD), and emotional dysregulation or instability. Clinical members of the team were consulted to achieve a consensus on the inclusion of such papers, although the large majority of the included papers did involve participants identified by a “personality disorder” diagnosis.

#### Interventions

Treatments with a primary focus on ‘personality disorder’ or associated needs, including psychotherapeutic treatments and service models, conducted in a community mental health care setting, or delivered to participants living in the community during treatment.

#### Controls

All comparators were considered (randomised and non-randomised), and we also included before and after study designs with no specific comparator group and studies in which the primary aim was uncontrolled preliminary testing of a new or newly adapted intervention.

#### Outcomes

Any measure of global clinical or symptom severity; psychiatric hospitalisation or emergency hospital presentations; self-harm or suicide-related outcomes; quality of life or general wellbeing; general, occupational, or social functioning (including interpersonal relations).

#### Study design

Quantitative studies, including randomised and non-randomised comparison studies and non-controlled studies with pre-post comparisons.

We excluded studies whose primary focus of treatment was not “personality disorder” diagnoses or comparable needs, or if the treatment was conducted in forensic, crisis care, or inpatient care settings. We also excluded theses and conference abstracts. Given the very broad nature of our searches, for feasibility we included only studies published in English. The full search and screening process is depicted in Figure 1.

**Figure 1.**
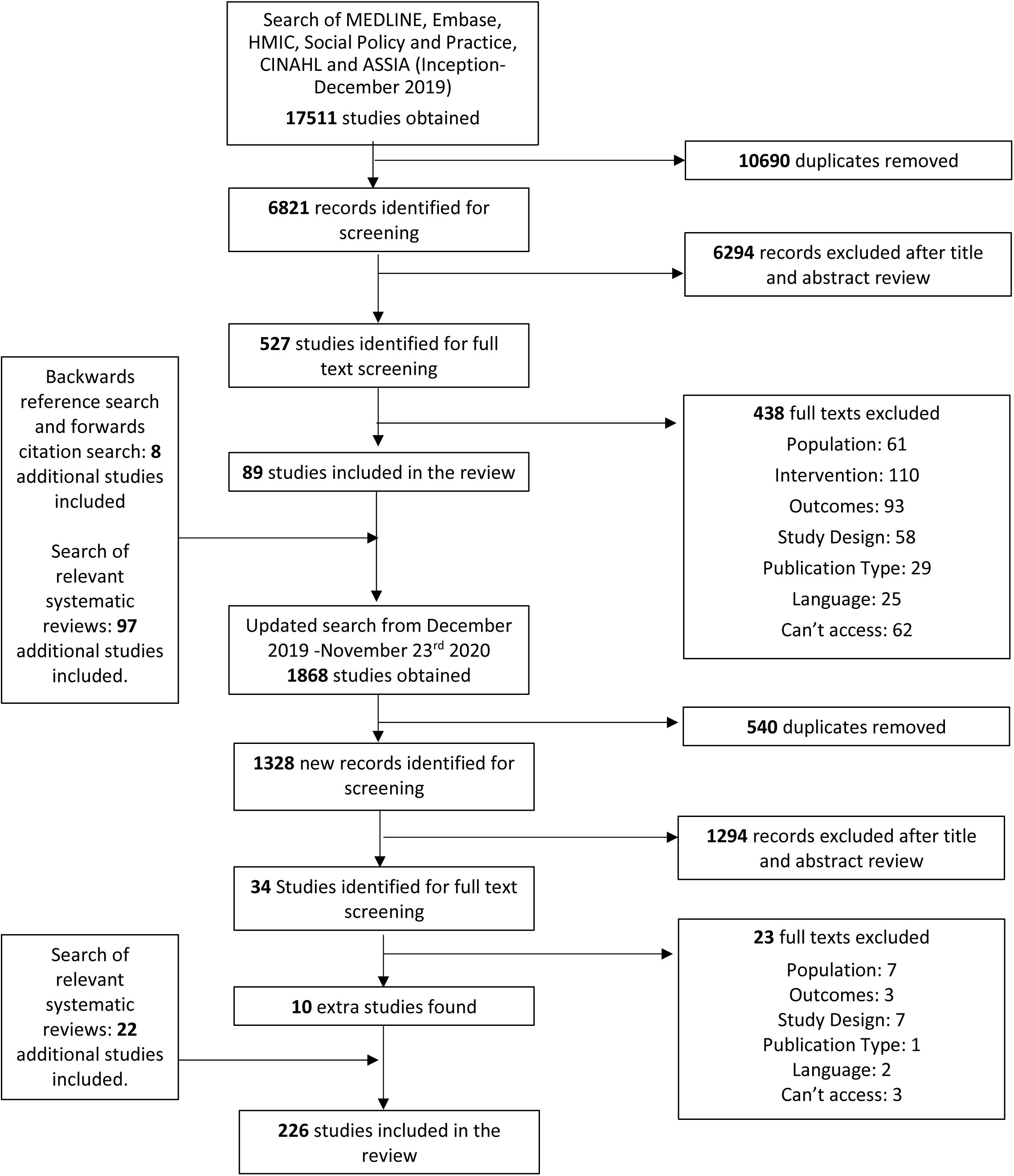
PRISMA Diagram

### Data extraction and synthesis

Data was extracted using a standardised extraction form and 10% was double checked for accuracy. Disagreements or errors were resolved by discussion with the team and corrected where required. Data extracted included study aims, study design, treatment and comparator details, sample characteristics and size, outcome measures, and study results. To present extracted data, papers were grouped by treatment modality, treatment/comparator category, and study design category. Treatment modality categories included: 1) DBT; 2) Cognitive and behavioural therapies; 3) MBT; 4) psychodynamic therapy; 5) schema therapy; 6) mixed modality psychotherapy; 7) other individual psychotherapy modalities; 8) Social or functional orientated therapy; 9) tests of service models or service re-organisation; 10) self-management or care planning; 11) family, couple, or parenting therapies. Treatment/comparator categories included: 1) non-active/treatment as usual comparator; 2) specialist or active comparator; 3) test of a modified version of the intervention; 4) test of a therapy adapted to a particular population. Study designs were categorised as follows: 1) Randomised Controlled Trials (RCTs) (noting where the study is clearly described as a pilot); 2) observational studies, including non-randomised controlled studies, and studies making pre-post comparisons within the same cohort; 3) Intervention development studies.

In keeping with guidance for scoping reviews, we did not carry out quality appraisal, but have placed a greater emphasis on more robust designs in our reporting.

## Results

Searches of bibliographic databases returned a total of 17,511 papers of which 5,385 papers were duplicates. After screening 6,821 titles and abstracts, reviewers screened 527 full texts. 438 papers did not meet our inclusion criteria and were excluded, resulting in 88 studies included in the review. Ninety-six additional studies were identified by searching relevant systematic reviews and eight studies through reference and citation searches. The search was updated on 23/11/2020 obtaining 1,868 records. After screening 34 full texts, 10 additional studies were included in the scoping review. Overall, we identified 226 papers for inclusion (Figure 1), reporting data from 210 distinct trials.

### Intervention types

Tables 1-5 provide a summary of included studies, and a detailed description of individual papers is presented in appendices 2-5. There have been more studies of DBT (Table 1, Appendix 2) than any other therapy modality or community-based treatment in this group (n= 66). We found 49 papers reporting studies of Cognitive and behavioural therapies (Table 2, Appendix 3), six of schema therapy (Table 2, Appendix 3), 54 of psychodynamic therapy (Table 3, Appendix 4), 20 of MBT (Table 3, Appendix 4), ten of mixed modality psychotherapy (Table 4, Appendix 5), seven of other individual psychotherapy modalities (Table 4, Appendix 5), five of socially or functionally orientated therapy (Table 4, Appendix 5), six of self-management or care planning (Table 4, Appendix 5), and 13 tests of novel mental health service models (Table 4, Appendix 5).

**Table 1:**
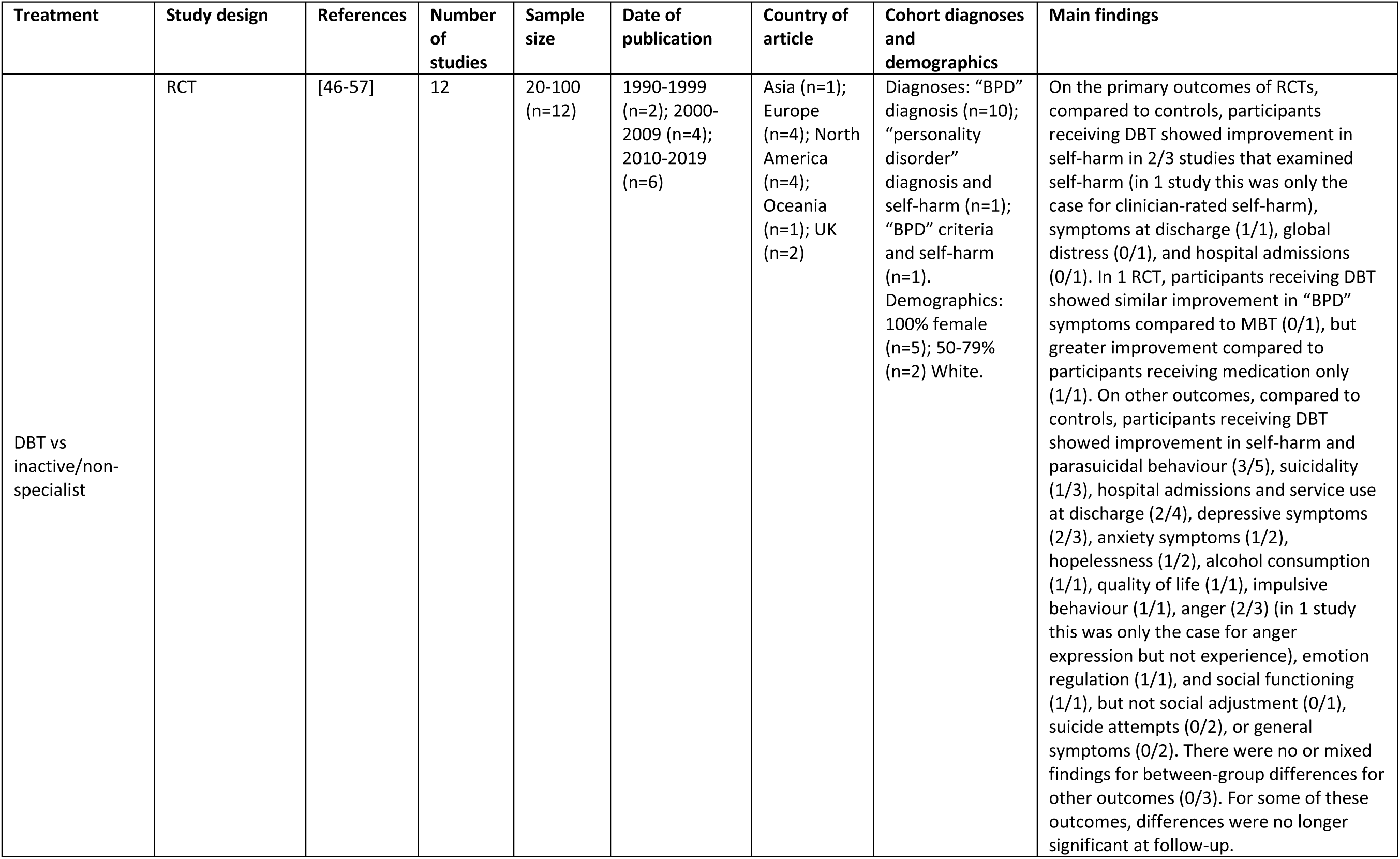
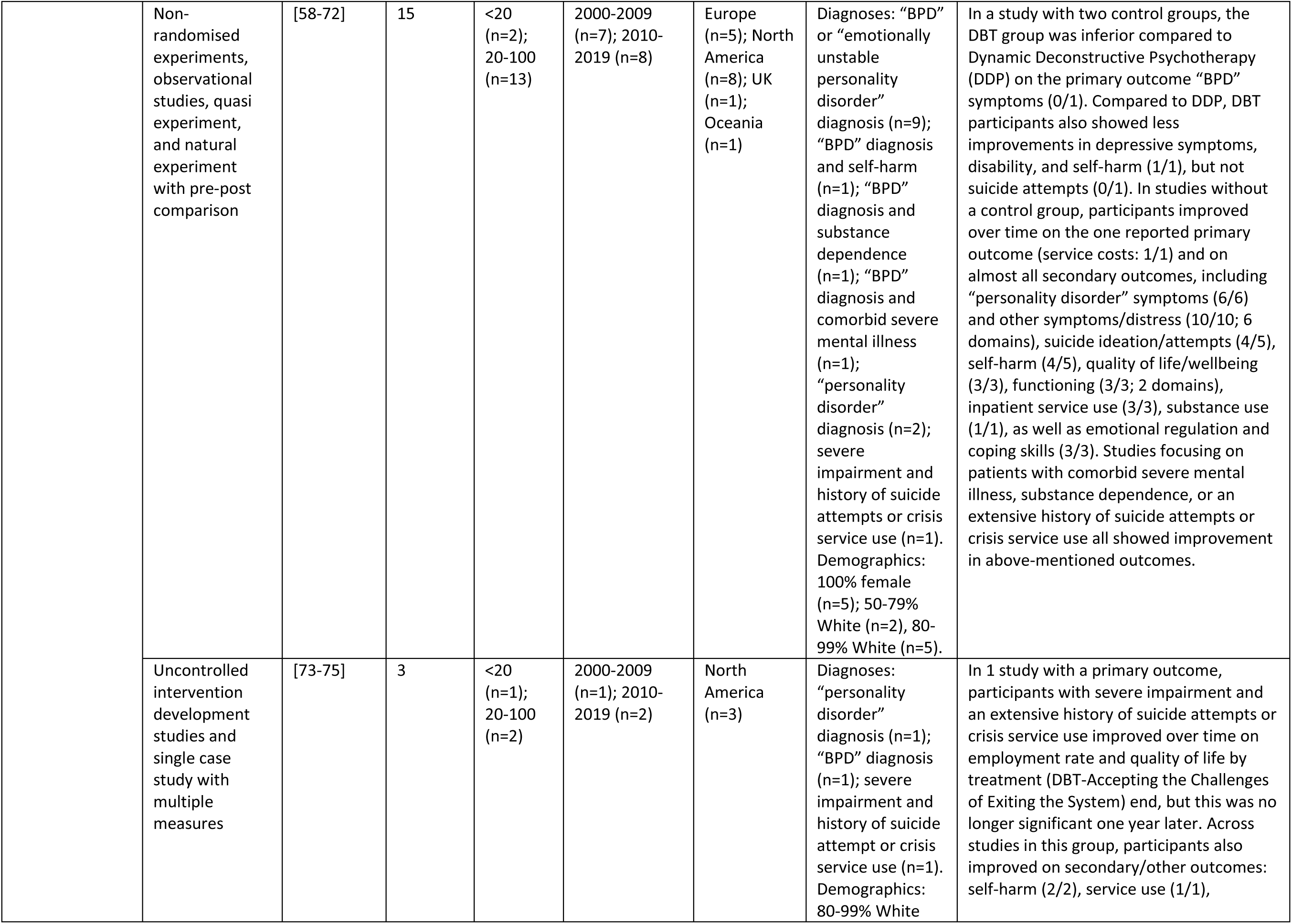
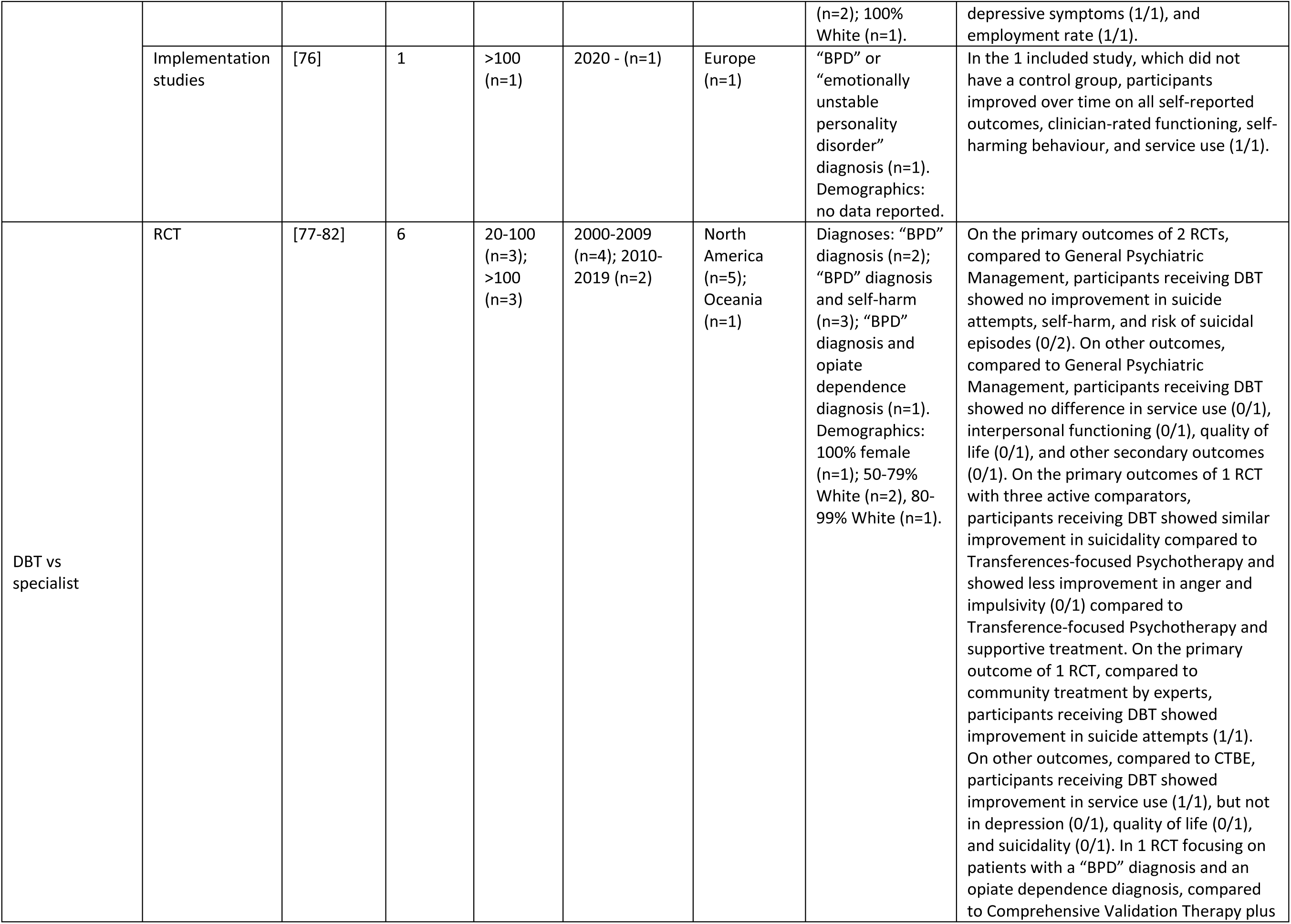
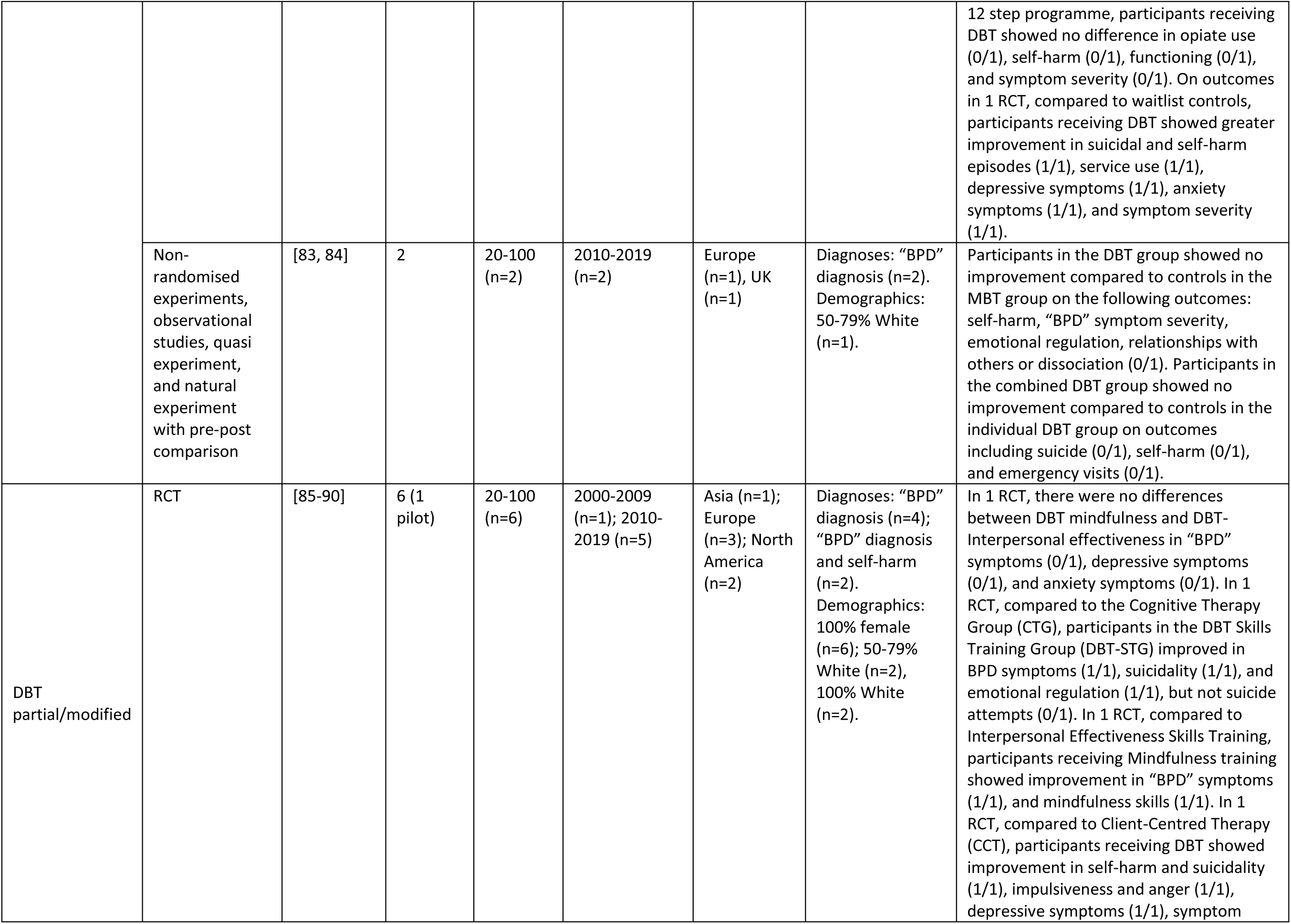
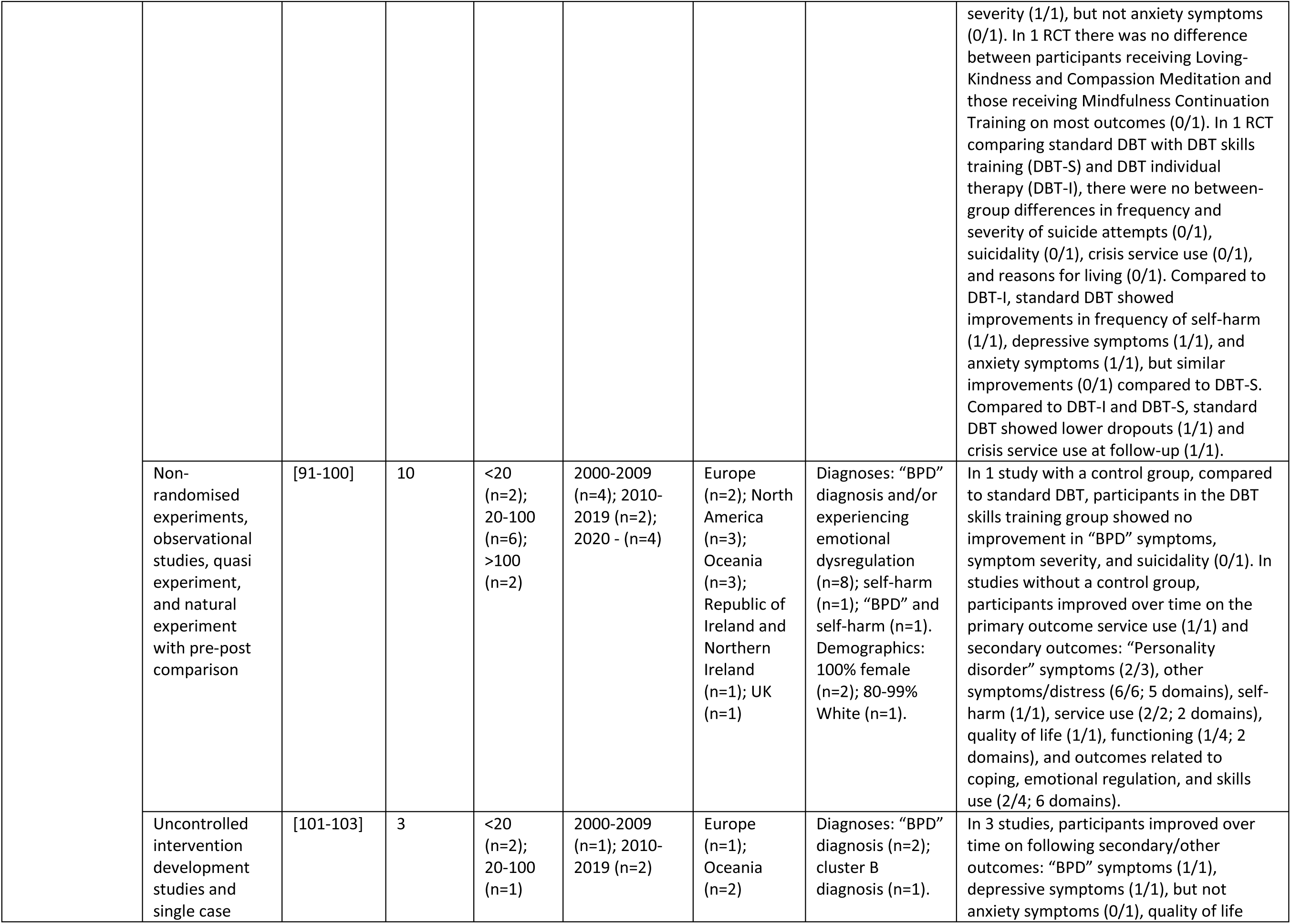
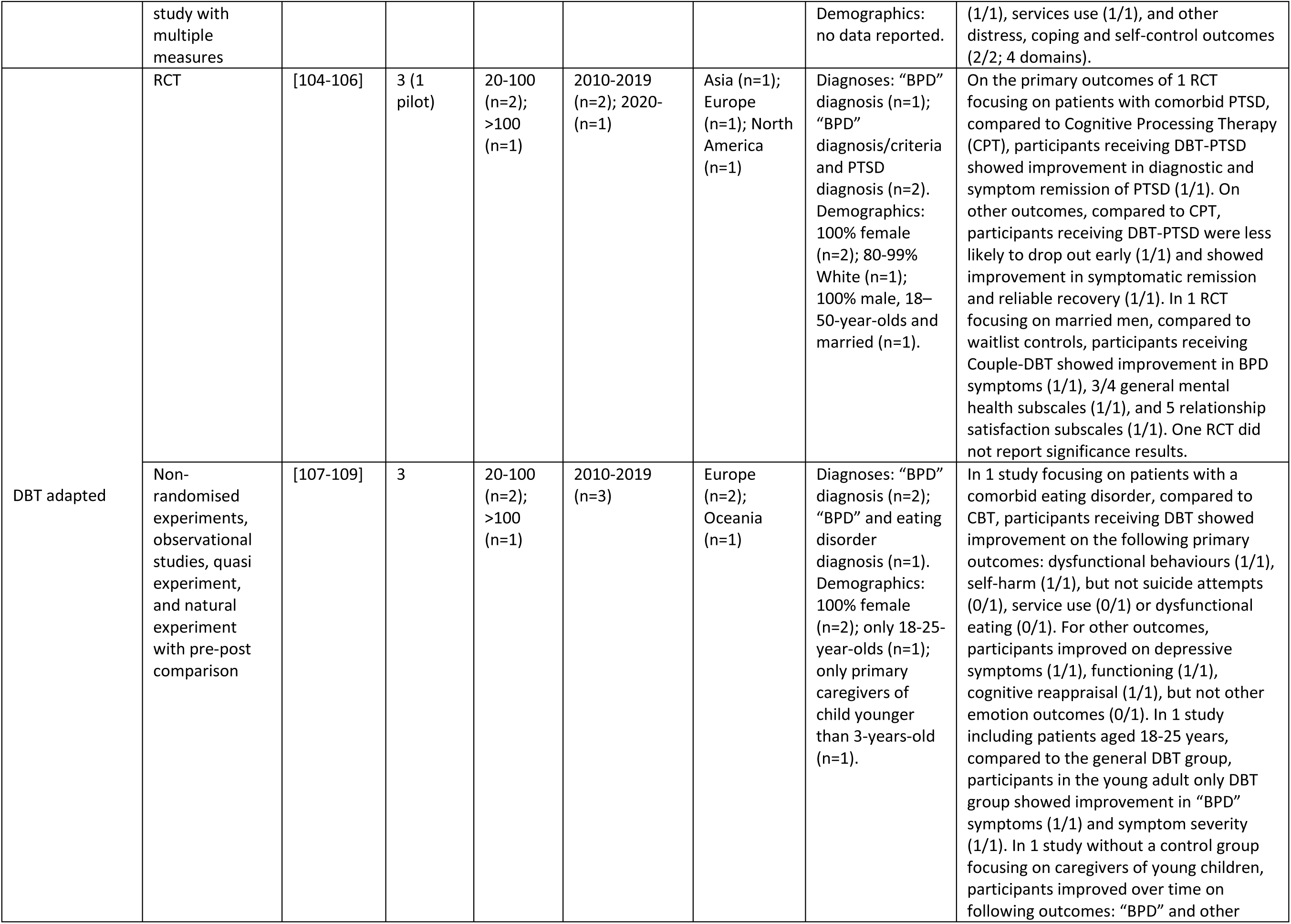
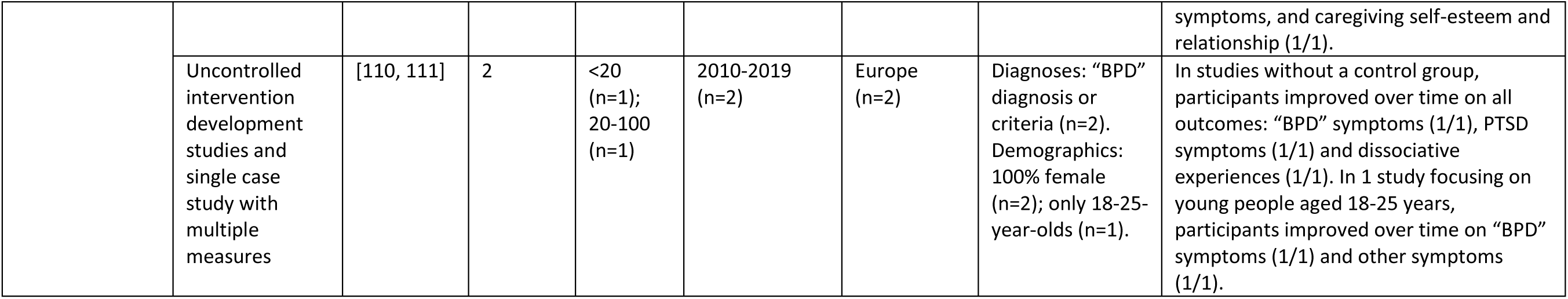
DBT

**Table 2:** Cognitive and behavioural and Schema therapies

| Treatment | Study design | References | Number of studies | Sample size | Date of publication | Country of article | Cohort diagnoses and demographics | Main findings |
| --- | --- | --- | --- | --- | --- | --- | --- | --- |
| Cognitive and behavioural vs inactive/non-specialist | RCT | [112-129] | 18 (4 pilot) | 20-100 (n=12); >100 (n=6) | 1990-1999 (n=2), 2000-2009 (n=7), 2010-2019 (n=9) | Europe (n=4); North America (n=6); Oceania (n=1); UK (n=7) | Diagnoses: Axis I and/or II diagnoses (n=1); avoidant “personality disorder” (n=1); “BPD” diagnosis or criteria (n=8); “personality disorder” diagnosis (n=3); “BPD” diagnosis/criteria and history of repeated self-harm (n=2); recent and previous self-harm (n=1); personality disturbance and recent and previous self-harm (n=1). Demographics: 100% female (n=4); 0-49% White (n=1), 80-99% White (n=5); 100% White (n=5). | On the primary outcomes of RCTs, compared to controls, participants receiving cognitive and behavioural therapies showed improvement in “personality disorder” symptoms (3/3), symptom severity (1/2), and social functioning (1/2), but not depressive (0/1) or (social) anxiety symptoms (0/1), service use (0/1), or frequency/number of participants with self-harming/suicidal behaviour (0/4). Compared to controls, a greater proportion of participants receiving cognitive and behavioural therapy recovered on symptoms (1/1). In other outcomes, compared to controls, participants receiving Cognitive and behavioural therapies showed improvement in symptom distress/severity (6/6), overall mental health (1/1), “personality disorder” symptoms (4/4), depressive symptoms (4/4) (with one study reporting mixed findings (0/1)), anxiety (2/6), stress (2/2), and dissociative (1/1) symptoms, hopelessness (1/1), quality of life (3/5), emotional regulation (3/4), self-harm (4/6), social functioning (1/4), global functioning (1/1), schemas (1/2), metacognition (1/1), and psychological flexibility (1/1). Cognitive and behavioural therapies were not superior on outcomes including proportion of participants meeting “BPD” criteria (0/1), service use (0/3), suicide attempts (0/1), suicidality (0/1), shyness (0/1), alexithymia (0/1), and costs (0/1) with one study reporting mixed findings (0/1). |
|  | Non-randomised experiments, observational studies, quasi experiment, and natural experiment with pre-post comparison | [130-137] | 8 | <20 (n=1); 20-100 (n=7) | 1990-1999 (n=1), 2000-2009 (n=2), 2010-2019 (n=5) | Europe (n=2); North America (n=3); UK (n=3) | Diagnoses: Avoidant “personality disorder” diagnosis (n=1); “BPD” diagnosis (n=3); “BPD” criteria/diagnosis and repeated self-harm or suicidality (n=2); “BPD” diagnosis/criteria, mood disorder, history of self-harm, and emotional and behavioural dysregulation (n=1); childhood sexual abuse (n=1). Demographics: 100% female (n=1); 50-79% White (n=1), 80-99% White (n=1). | In studies without a control group, participants improved over time on the one reported primary outcome (self-harm: 1/1) and secondary outcomes: “personality disorder”/”BPD” symptoms (2/3), other symptoms (4/4; 5 domains), self-harm and suicide ideation/attempts (3/3), hospitalisation (1/1), quality of life (1/1), emotional regulation/intensity (2/2), schemas (1/1), personality beliefs (1/2), combination of measures (1/1), and most clinical and social outcomes (1/1), but not cognitive filter (0/1). In one study focusing on patients with childhood sexual abuse, participants improved over time on emotional regulation (1/1), interpersonal problems (1/1), and trauma symptoms (1/1). |
|  | Uncontrolled intervention development studies and single case study with multiple measures | [138-148] | 11 | <20 (n=8); 20-100 (n=3) | 2000-2009 (n=2), 2010-2019 (n=9) | Asia (n=2), Europe (n=3); North America (n=1); Oceania (n=1); UK (n=4) | Diagnoses: "BPD" diagnosis (n=4); "BPD" diagnosis and comorbid emotional disorder (n=1); current or historic "BPD" diagnosis, "BPD" features, and current drug/alcohol disorder (n=1); "obsessive-compulsive personality disorder" diagnosis (n=1); chronic mood or adjustment disorder and comorbid "personality disorder" diagnosis/features (n=1); cluster-B or cluster-C "personality disorder" diagnosis or Axis II features (n=1); "personality disorder" diagnosis (n=2). Demographics: 80-99% White (n=1); 100% White (n=1); older age (n=1) | In studies without a control group, participants improved over time on the primary outcomes symptoms/distress (2/2; 2 domains) and quality of life (1/1), and also showed no dropouts (1/1). Participants improved on the following secondary outcomes: "personality disorder" symptoms (4/4) and other symptoms (4/4; 6 domains); functioning (2/2; 2 domains); personality integration/beliefs (2/2); emotional regulation, coping, and skills (3/3; 5 domains). Additionally, patients with a current substance misuse disorder showed a reduction in drug use. Elderly patients with a chronic mood or adjustment disorder showed improvement in symptom distress (1/1) and some but not all aspects of schema and coping variables (1/1). |
| Cognitive and behavioural vs specialist | RCT | [149-152] | 4 | 20-100 (n=4) | 2000-2009 (n=3), 2010-2019 (n=1) | Europe (n=3); North America (n=1) | Diagnoses: "BPD" features/diagnosis (n=2); cluster C or self-defeating "personality disorder" (n=2). Demographics: 100% White (n=1). | On the primary outcomes of 1 RCT, there was no difference between cognitive therapy and Rogerian Supportive Therapy in symptom improvement (0/1) as well as no between-group difference on secondary outcomes (0/1). In a RCT comparing Schema Focused Therapy (SFT) with cognitive therapy, significantly more participants receiving SFT recovered on the primary outcome ("BPD" symptoms: 1/1) as well as on secondary outcomes: symptom severity (1/1), "BPD" symptoms (1/1), and quality of life (1/1). In another RCT, there was no difference between patients receiving cognitive therapy and those receiving standalone outpatient treatment in the primary outcomes: symptom severity (0/1) and interpersonal problems (0/1). In a RCT comparing Dynamic psychotherapy with CT there were no between-group differences in outcomes: symptom severity (0/1), interpersonal problems (0/1), and “personality disorder” symptoms (0/1). |
|  | Non-randomised experiments, observational studies, quasi experiment, and natural experiment with pre-post comparison | [153-155] | 3 | 20-100 (n=1); >100 (n=2) | 2000-2009 (n=1), 2010-2019 (n=2) | Europe (n=3) | Diagnoses: “personality disorder” diagnosis (n=1); “personality disorder” NOS diagnosis (n=1); “BPD” diagnosis or other cluster-B “personality disorder” diagnosis with comorbid Axis I disorder (n=1). Demographics: no data report. | In studies with a control group, there was no difference between the TAU-CBT group and participants receiving ACT on the only primary outcome (personality functioning: 0/1) and most secondary outcomes (0/1). There were no differences between individuals receiving Double Setting Cognitive-Evaluation Therapy (DS-CET) and those receiving Individual Cognitive-Evolution therapy (I-CET) on any of the outcomes (0/1). In one study comparing six active groups, there was no between-group difference on the primary outcome (symptom severity: 0/1), and most groups improved on the secondary outcomes of social functioning and quality of life. |
|  | Uncontrolled intervention development studies and single case study with multiple measures | [156] | 1 | <20 (n=1) | 2010-2019 (n=1) | North America (n=1) | Diagnoses: NSSI disorder (n=1). Demographics: 50-79% White (n=1). | No significant results reported for outcomes in the 1 included study on patients with NSSI disorder. However, 8/10 participants reported meaningful reductions in self-harming behaviour. |
| Cognitive and behavioural modified | RCT | [157, 158] | 2 (1 pilot) | <20 (n=1); 20-100 (n=1) | 1990-1999 (n=1), 2010-2019 (n=1) | North America (n=1); UK (n=1) | Diagnoses: "BPD" diagnosis (n=1); previous suicide attempts, antidepressants taken as part of an overdose, and suicidal behaviour (n=1). Demographics: 80-99% White (n=1). | On the primary outcome of 1 RCT, findings for differences between the Cognitive Behavioural Problem Solving and TAU group on suicidality were mixed (0/1). On other outcomes, findings were mixed or showed no between-group differences (0/2). |
|  | Non-randomised experiments, observational studies, quasi experiment, and natural experiment with pre-post comparison | [159] | 1 | 20-100 (n=1) | 2000-2009 (n=1) | Europe (n=1) | Diagnoses: "personality disorder" diagnosis, excluding borderline, schizotypal, schizoid, antisocial "personality disorder", or "personality disorder" NOS (n=1). Demographics: no data reported. | The 1 study utilised a crossover design and showed significant improvements over the treatment period as a whole, but no between-group differences. |
| Cognitive and behavioural adapted | Uncontrolled intervention development studies and single case study with multiple measures | [160] | 1 | <20 (n=1) | 2010-2019 (n=1) | Oceania (n=1) | Diagnoses: "personality disorder" diagnosis (n=1). Demographics: no data reported. | No statistical analysis conducted in the 1 included study. However, 5/8 patients no longer met criteria for an avoidant "personality disorder" at end of follow-up. |
| Schema therapy vs inactive/non-specialist | RCT | [161] | 1 | >100 (n=1) | 2010-2019 (n=1) | Europe (n=1) | Diagnoses: Avoidant, dependent, obsessive-compulsive, paranoid, histrionic, or narcissistic "personality disorder" diagnosis (n=1). | On the primary outcome of the 1 RCT, compared to controls, a greater proportion of participants receiving schema therapy recovered (1/1). On other outcomes, compared to controls, participants receiving schema therapy showed improvement on 2/3 measures of functioning, but not quality of life (0/1). |
|  | Uncontrolled intervention development studies and single case study with | [162-165] | 4 | <20 (n=4) | 2000-2009 (n=1), 2010-2019 (n=3) | Europe (n=3); North America (n=1) | Diagnoses: "BPD" diagnosis (n=3); cluster C "personality disorder" or "personality disorder" not otherwise specified with cluster C traits (n=1). | In the 1 study that reported significance results, participants improved on "BPD" symptoms (1/1) and most other outcomes over time. |
|  | multiple measures |  |  |  |  |  | Demographics: 100% female (n=3); old age (n=1) |  |
| Schema therapy modified | RCT | [166] | 1 | 20-100 (n=1) | 2000-2009 (n=1) | Europe (n=1) | Diagnoses: "BPD" diagnosis (n=1). Demographics 80-99% (n=1). | On the primary outcome of the 1 RCT, there was no difference between participants receiving schema therapy with and those without phone support on recovery from "BPD" (0/1). There was also no significant difference on other outcomes (0/1). |

**Table 3.** MBT and psychodynamic therapies

| Treatment | Study design | References | Number of studies | Sample size | Date of publication | Country of article | Cohort diagnoses and demographics | Main findings |
| --- | --- | --- | --- | --- | --- | --- | --- | --- |
| MBT vs inactive/non-specialist | RCT | [56, 167-169] | 4 | 20-100 (n=4) | 1990-1999 (n=1), 2000-2009 (n=2), 2010-2019 (n=1) | Asia (n=1), UK (n=3) | Diagnoses: "BPD" diagnosis (n=4). Demographics: no data reported. | In the primary outcomes of RCTs, compared to controls, participants receiving MBT showed improvement in the proportion of patients making suicide attempts in 1/1 studies. In 1 RCT, participants receiving MBT showed similar improvement in "BPD" symptoms compared to DBT (0/1), but greater improvement compared to participants receiving medication only (1/1). In other outcomes, compared to controls, participants receiving MBT showed improvement in symptom severity (1/1), depressive and anxiety symptoms (1/1), other symptoms (1/1), self-harming behaviour (1/1), medication use (1/1), social functioning (2/2), number of patients engaging in self-harm or suicide attempts (1/1) and being admitted to the hospital (1/1). |
|  | Non-randomised experiments, observational studies, quasi experiment, and natural experiment with pre-post comparison | [170-175] | 6 | <20 (n=2), 20-100 (n=3), >100 (n=1) | 2010-2019 (n=6) | Europe (n=6) | Diagnoses: "BPD" diagnosis (n=4), Generic "personality disorder" diagnosis (n=1), Generic "personality disorder" diagnosis and poor functioning (n=1)<br>Demographics: 100% female (n=1). | In one study with a control group , no primary outcomes were reported. In other outcomes, participants improved in symptom distress, interpersonal functioning, global functioning, and occupational functioning. Participants did not improve compared to control in suicidal acts or self-harm, hospital admissions, and or medication use. In studies without a control group, participants showed improvements over time on the following primary outcomes: "personality disorder" symptoms (2/2), Interpersonal problems (1/1). In other outcomes, participants showed improvements over time in symptoms (3/3), global functioning (2/2), suicidality (1/1), service use (1/1), and unemployment (1). |
| MBT vs specialist | RCT | [176-179] | 4 | 20-100 (n=1), >100 (n=3) | 2000-2009 (n=1), 2010-2019 (n=3) | Europe (n=3), UK (n=1) | Diagnoses: "BPD" diagnosis (n=3), "BPD" and suicide attempt or life-threatening self-harm (n=1).<br>Demographics: 50-79% White (n=1). | In the primary outcomes of RCTs, compared to specialist TAU psychotherapy, participants receiving MBT did not show improvement of borderline symptoms (0/1). Compared to structured clinical management, participants receiving MBT showed improvement in suicidal behaviours (1/1) and number of hospitalisations (1/1). In other outcomes, compared to specialist TAU psychotherapy, participants receiving MBT did not show improvements in general symptom severity (0/1), depressive symptoms (0/1), interpersonal problems (0/1) or quality of life (0/1). Compared to supportive group therapy, participants receiving MBT showed improvement in global functioning (1/2) but did not show improvement in depressive symptoms (0/2), anxiety symptoms (0/2), interpersonal functioning (0/2) or social functioning (0/1). Compared to structured clinical management, participants receiving MBT showed improvements in symptoms (1/1) and social functioning (1/1), but did not show improvements in depressive symptoms (0/1) and global functioning (0/1). |
|  | Non-randomised experiments, observational studies, quasi experiment, and natural experiment with pre-post comparison | [83, 180-183] | 5 | 20-100 (n=4), >100 (n=1) | 2010-2019 (n=5) | Europe (n=1), UK (n=4) | Diagnoses: "BPD" diagnosis (n=2), "personality disorder" diagnosis (n=3). Demographics: 50-79% White (n=1), 80-99% White (n=3). | In studies with a control group, the primary outcome of bed use compared to an alternative psychoanalytic model did not significantly improve in the MBT group. For other outcomes, compared to DBT, participants receiving MBT did not have significantly reduced self-harm (0/1), symptom severity (0/1), emotional dysregulation (0/1), interpersonal problems (0/1) or dissociation (0/1). Compared to various other specialist treatments, participants receiving MBT had improved symptoms (1/1) and personality functioning (1/1) but did not have improved relational functioning (0/1). In studies without a control group, participants improved over time on bed use (1/1), global functioning (1/1), and symptom severity (1/1) but did not improve over time on other symptom measures (0/2), social adjustment (0/1), self-esteem (0/1), and quality of life (0/1). |
| MBT modified | RCT | [184] | 1 | >100 (n=1) | 2020- (n=1) | Europe (n=1) | Diagnoses: Generic "personality disorder" diagnosis: (n=1). Demographics: no data reported. | Compared to lower intensity outpatient MBT, higher intensity day hospital MBT showed no difference in the primary outcome of symptom severity. In other outcomes, there was no difference in personality functioning, interpersonal problems, quality of life and or suicide and self-harm. |
| Psychodynamic vs inactive/non-specialist | RCT | [122, 185-189] | 6 | 20-100 (n=4), >100 (n=2) | 1990-1999 (n=2), 2000-2009 (n=3), 2010-2019 (n=1) | Canada (n=2), Europe (n=3), USA (n=1) | Diagnoses: "BPD" diagnosis (n=1), Generic "personality disorder" diagnosis (n=2), Avoidant "personality disorder" (n=1), "personality disorder" diagnosis other than paranoid, schizoid, schizotypal, narcissistic, or borderline (n=1), long term psychiatric difficulties disrupting functioning (n=1). Demographics: no data reported. | In the primary outcomes of RCTs, compared to controls, participants receiving psychodynamic therapy showed improvement in symptom severity (2/2), social functioning (1/2), and interpersonal functioning (1/1) but not dysfunctional borderline beliefs (0/1), anxiety symptoms (0/1) or the number of participants meeting diagnostic criteria for a "personality disorder" diagnosis (0/1). In other outcomes, compared to controls, participants receiving psychodynamic therapy showed improvements in symptom severity (3/4), depressive symptoms (2/2), suicide intentionality (1/1), self-esteem (2/2), life satisfaction (1/1), social functioning (3/3), interpersonal functioning (1/1), global functioning (1/2), and occupational functioning (1/1), but did not show improvements in the number of patients meeting diagnostic criteria for a "personality disorder" diagnosis (0/1), emotional reliance (0/1) or anxiety symptoms (0/1). |
|  | Non-randomised experiments, observational studies, quasi experiment, and natural experiment with pre-post comparison | [61, 190-214] | 26 | <20 (n=1), 20-100 (n=18), >100 (n=7) | 1990-1999 (n=6), 2000-2009 (n=12), 2010-2019 (n=7), 2020- (n=1) | Australia (n=7), Canada (n=4), Europe (n=10), UK (n=3), USA (n=2) | Diagnoses: Generic "personality disorder" (n=8), "BPD" diagnosis (n=8), "personality disorder" diagnosis or significant traits of personality dysfunction (n=2), treatment resistant depression with comorbid personality disorder and childhood trauma (n=1), "personality disorder" diagnosis and poor interpersonal functioning (n=2), problematic interpersonal functioning (n=1), "personality disorder" diagnosis and comorbid axis I mental health problem (n=3), | In studies with a control group, participants showed improvements compared to controls on the following primary measures: reflective functioning (2/2), "personality disorder" symptoms (1/1), social functioning (1/1), and depressive symptoms (1/1). In other outcomes, participants improved compared to the control on "personality disorder" symptoms (6/6), global functioning (4/4), social functioning (5/5), depressive symptoms (2/2), suicidal ideation or self-harm (2/3), interpersonal functioning (2/2), anxiety symptoms (1/1), and number of emergency contacts (1/1). In studies without a control group, participants improved over time in primary outcomes in interpersonal functioning (3/3) and symptom severity (1/1). In secondary outcomes, participants improved over time in symptom severity (13/13), other symptom measures (depression (6/6) the number of participants meeting diagnosis (4/4), anxiety (4/4), suicide and self-harm (5/5), functioning measures (11/12), service use (3/3), drug use (2/2), violence (1/1), life satisfaction (1/1), and self-esteem (1/1). Above-mentioned findings include studies that focused on patients with treatment |
|  |  |  |  |  |  |  | Avoidant or obsessive-compulsive "personality disorder" (n=1). Demographics: 100% female (n=1); 80-99% White (n=3), 100% White (n=1). | resistant depression and comorbid personality disorder and childhood trauma (n=1), "personality disorder" diagnosis and poor interpersonal functioning (n=2), problematic interpersonal functioning (n=1), and "personality disorder" diagnosis and comorbid axis I mental health problem (n=3). |
|  | Uncontrolled intervention development studies and single case study with multiple measures | [215] | 1 | 20-100 (n=1) | 2000-2009 (n=1) | USA (n=1) | Diagnoses: "BPD" symptoms and suicidal or self-injurious behaviour (n=1). Demographics: 100% female (n=1); >50% white (n=1). | One uncontrolled feasibility trial found that patients given psychodynamic therapy improved over time in functioning, parasuicide and service utilisation (1/1). |
| Psychodynamic vs specialist | RCT | [80, 151, 152, 216-220] | 8 | 20-100 (n=8) | 1990-1999 (n=1), 2000-2009 (n=2), 2010-2019 (n=5) | Europe (n=5), Europe and North America (n=1), North America (n=2) | Diagnoses: "BPD" diagnosis (n=5), "personality disorder" diagnosis (n=1), Cluster C "personality disorder" diagnosis (n=2). Demographics: 50-79% White (n=1), 80-99% White (n=2), 100% white (n=1). | In primary outcomes of RCTs, compared to cognitive therapy, participants receiving psychodynamic therapy did not have significantly improved "personality disorder" symptoms (0/1). Compared to General Psychiatric Management, participants receiving psychodynamic therapy made significantly more overall progress in therapy (1/3) overall. Though no direct contrasts were made, in an RCT of DBT, supportive treatment and psychodynamic therapy, participants receiving psychodynamic therapy improved significantly in suicidality, aggression and impulsivity (1/1). In other outcomes, compared to cognitive therapy, participants receiving psychodynamic therapy did not improve interpersonal functioning (0/2), symptoms (0/1), personality functioning (0/1) or the number of patients with a "personality disorder" diagnosis (0/1). Compared to General Psychiatric Management, participants receiving psychodynamic therapy had improved<br>symptom distress (1/1) but did not have improved interpersonal functioning (0/2), symptom severity (0/2), social functioning (0/1), number of crisis consultations (0/1) or number of days spent in inpatient treatment (0/1). Compared to Short Term Dynamic Therapy, participants receiving Brief Supportive Psychotherapy showed no improvement in symptoms (0/1) or interpersonal functioning (0/1). Though again no direct contrasts were made, in the RCT of DBT, supportive treatment and psychodynamic therapy, participants receiving psychodynamic therapy improved significantly in depression (1/1), anxiety (1/1), global functioning (1/1) and social functioning (1/1). |
|  | Non-randomised experiments, observational studies, quasi experiment, and natural experiment with pre-post comparison | [221-224] | 4 | 20-100 (n=3), >100 (n=2) | 1990-1999 (n=1), 2010-2019 (n=3) | Europe (n=3), USA (n=1) | Diagnoses: Generic “personality disorder” diagnosis (n=1), generic “personality disorder” diagnosis with comparison between comorbid substance misuse (n=1), “BPD” diagnosis (n=2). Demographics: no data reported. | In studies with a control group, compared to stabilising treatments, participants given stabilising treatments had significantly higher improvements in symptom severity (1/1) and interpersonal functioning (1/1). Compared to DBT, participants given DDP had significantly greater improvement in the primary outcome of symptom severity (1/1), and other outcomes of self-harm (1/1), depression (1/1), and social and occupational impairment (1/1). Compared to day treatment without follow-up group psychotherapy, participants who were provided with follow-up group psychotherapy showed significant improvements in health sickness (1/1) and symptom severity (1/1) but did not show significantly different improvements in rehospitalisation (0/1) or suicide attempts (0/1). In one study with a pre-post comparison of patients with and without comorbid substance misuse in General Psychiatric Management, borderline symptoms improved significantly over time for both groups. One study found improved<br>global functioning in group therapy compared to individual therapy. |
| Psychodynamic treatment setting comparisons | Non-randomised experiments, observational studies, quasi experiment, and natural experiment with pre-post comparison | [154, 225-229] | 6 | >100 (n=6) | 2000-2009 (n=2), 2010-2019 (n=4) | Europe (n=3), UK (n=2), Europe and UK (n=1) | Diagnoses: Generic "personality disorder" (n=4), "personality disorder" NOS (n=1), severe "personality disorder" (n=1). Demographics: no data reported. | Six studies compared psychodynamic treatment in varying contexts. Primary outcomes were rarely reported. In studies comparing community-based services or step-down services to residential services, community or step-down services resulted in significantly improved symptom severity (1/1), psychiatric distress (1/1), self-harm and suicide (2/2), social adaption (1/1), and global functioning compared to residential services. In studies comparing day hospital, outpatient and inpatient services, there were no significant differences in outcomes between settings in symptom severity (0/4), psychosocial function (0/4), quality of life (0/3) or interpersonal functioning (0/2). |
| Psychodynamic adapted | RCT | [230, 231] | 2 | 20-100 (n=2) | 2000-2009 (n=1), 2010-2019 (n=1) | USA (n=2) | Diagnoses: "BPD" and alcohol use or substance dependence (n=2). Demographics: no data reported. | In the primary outcomes of RCTs comparing DDP combined with alcohol rehabilitation compared to TAU with alcohol rehabilitation for patients with co-occurring substance use disorders, DDP patients showed significantly higher clinically meaningful improvement (1/1), alcohol misuse (1/1), and use of institutional care (1/1). In other outcomes, participants receiving DDP showed significant improvements in symptom severity (2/2), depression (2/2), parasuicide (1/1), recreational drug use (1/1), and perceived social support (1/2) but did not show improvement compared to TAU in dissociation (0/1), heavy drinking days (0/1), and days employed (0/1). |
|  | Non-randomised experiments, observational studies, quasi experiment, and natural experiment with pre-post comparison | [232] | 1 | 20-100 (n=1) | 2011-2019 (n=1) | Europe (n=1) | Diagnoses: "BPD" diagnosis (n=1) Demographics: relatively low socio-economic status (n=1). | A brief psychoeducational program based on General Psychiatric Management was more effective than generic outpatient treatment in improving symptom severity (1/1), except for the impulsivity subscale. |

**Table 4:**
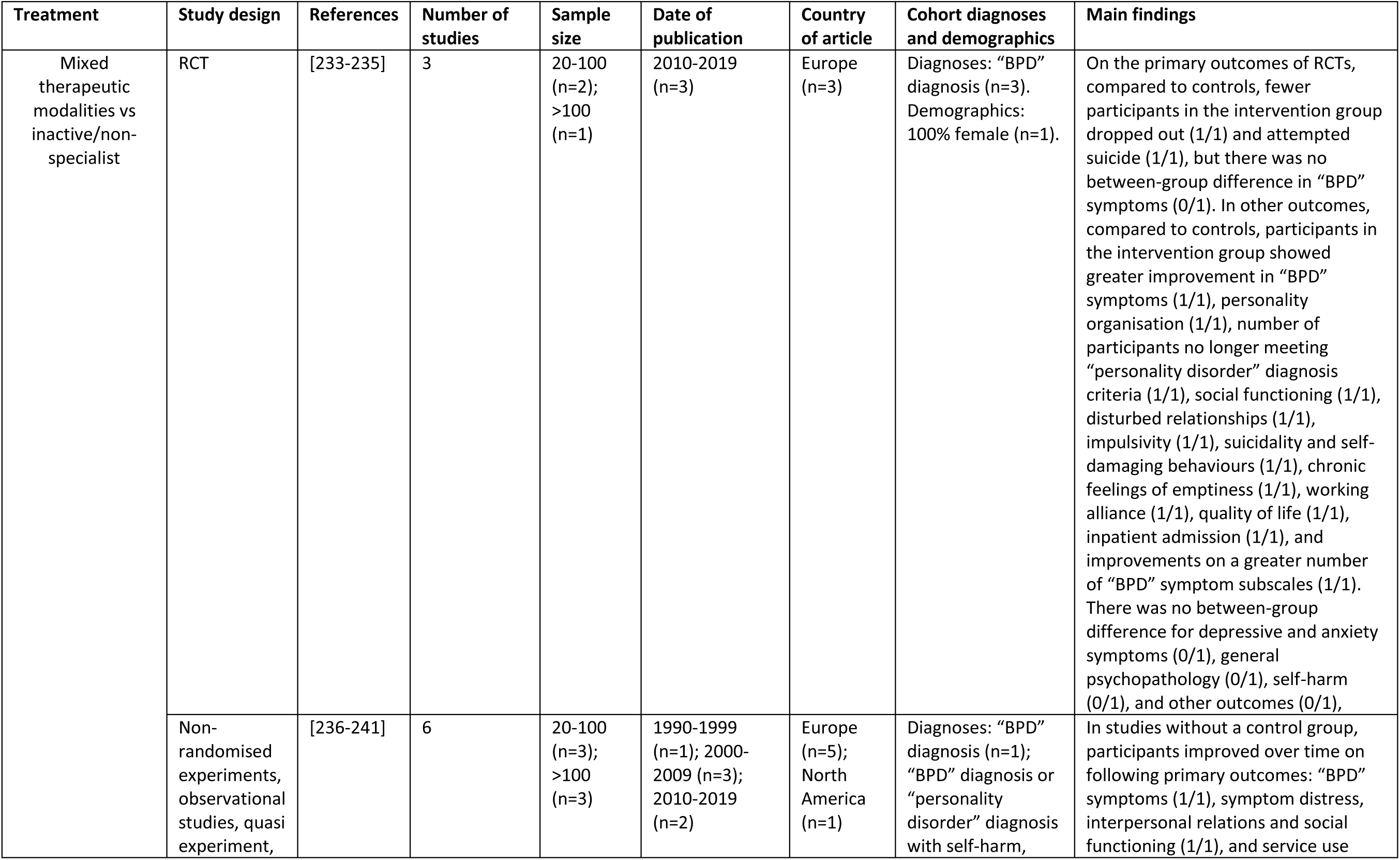
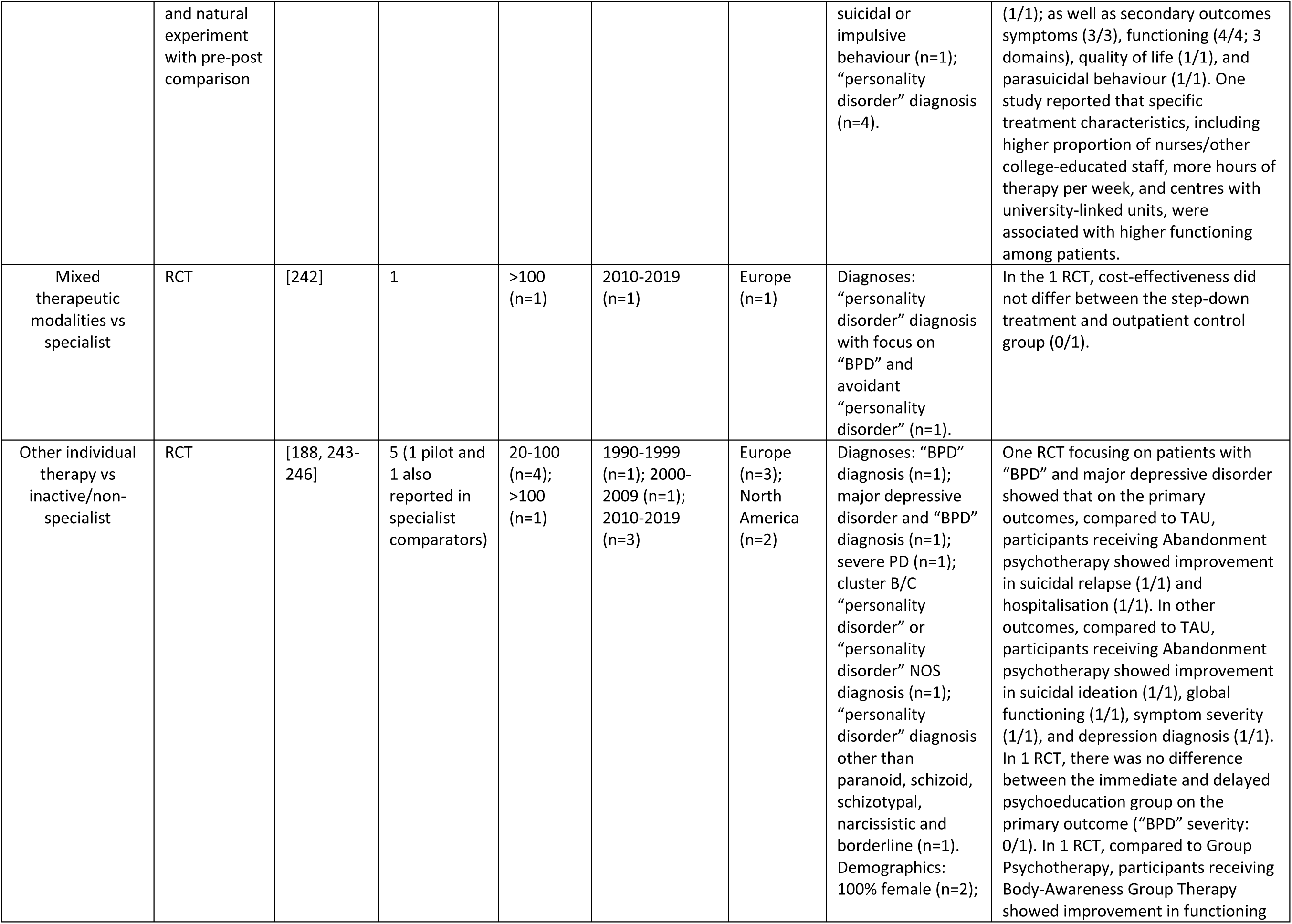
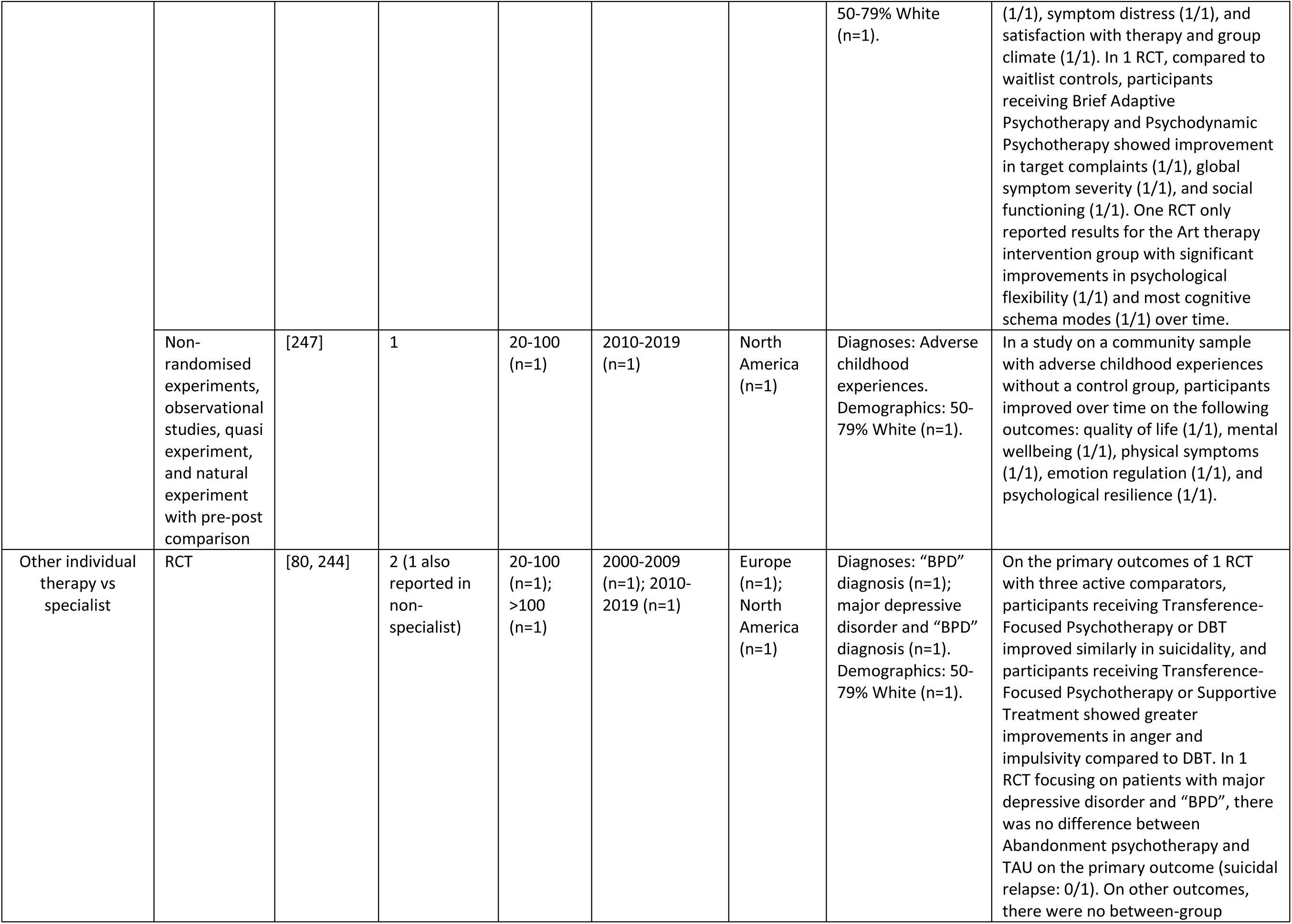
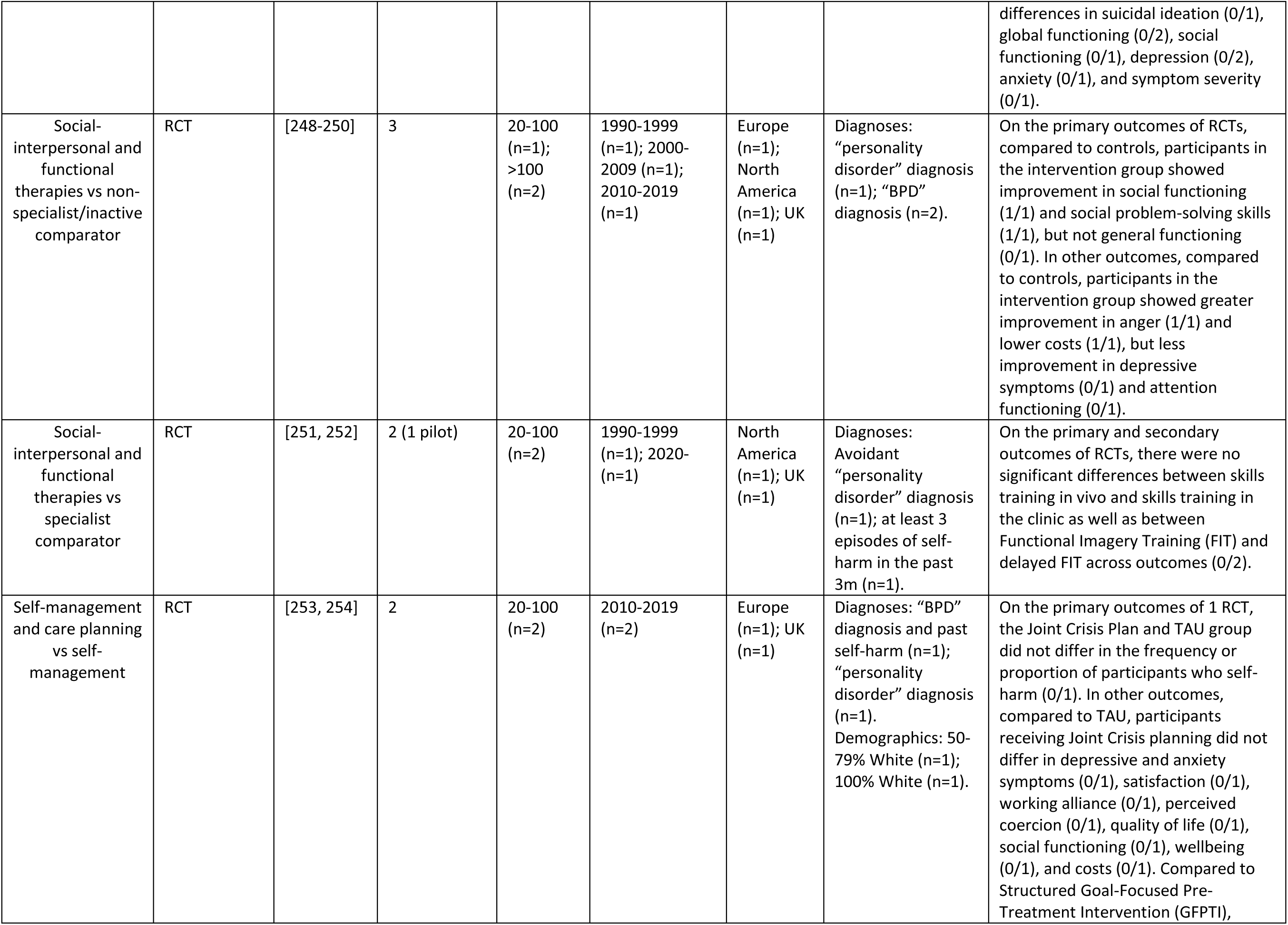
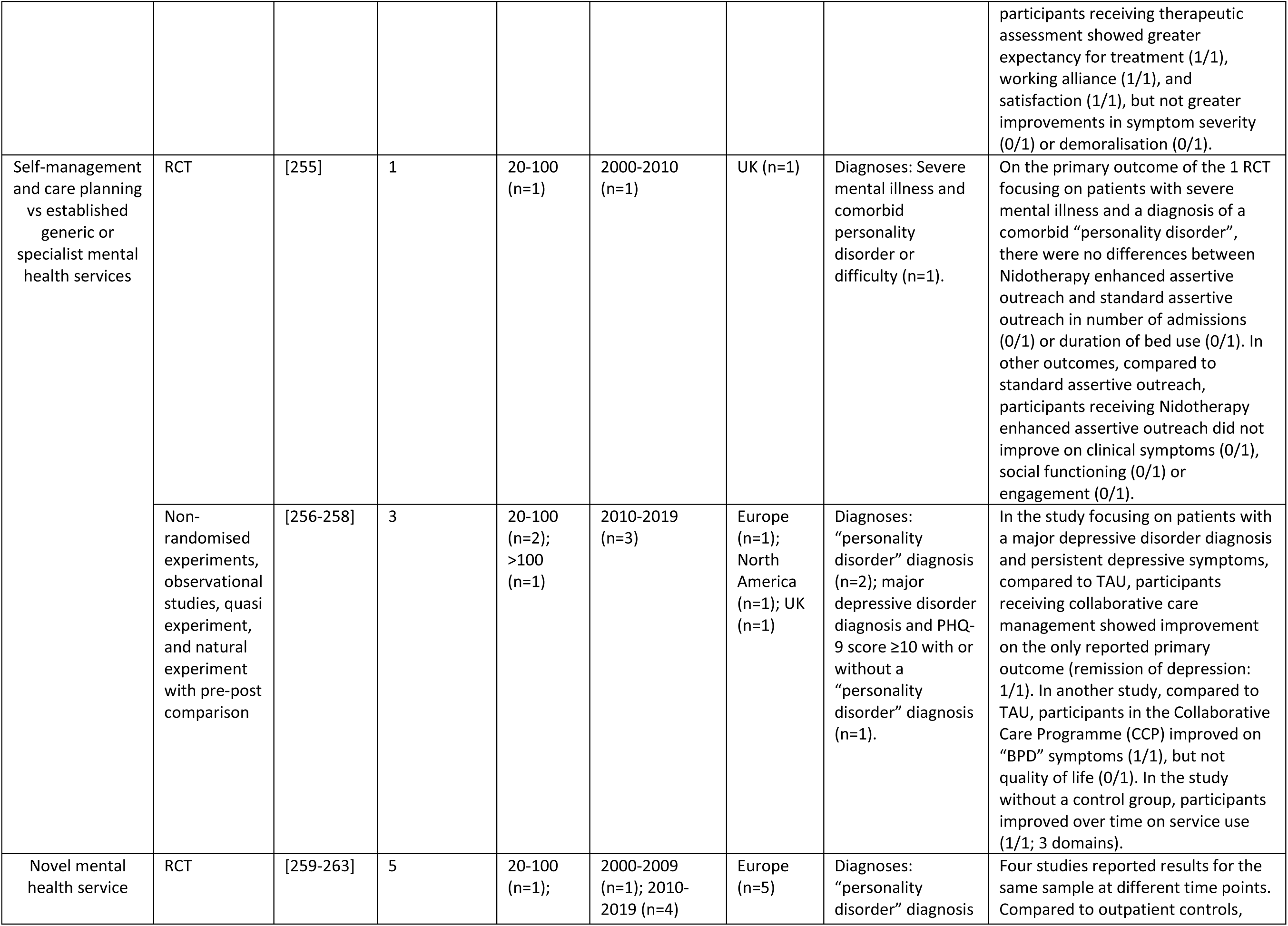
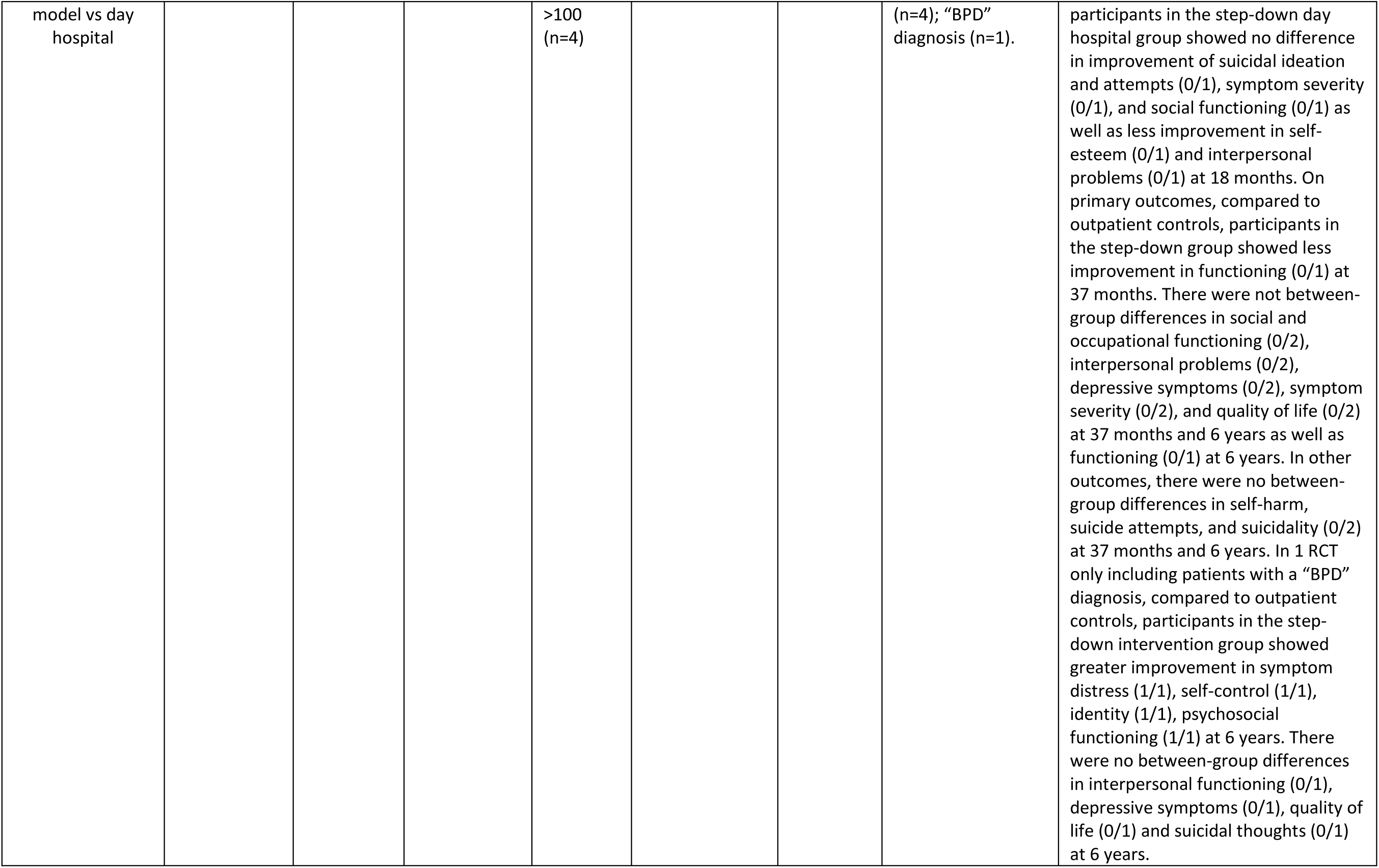
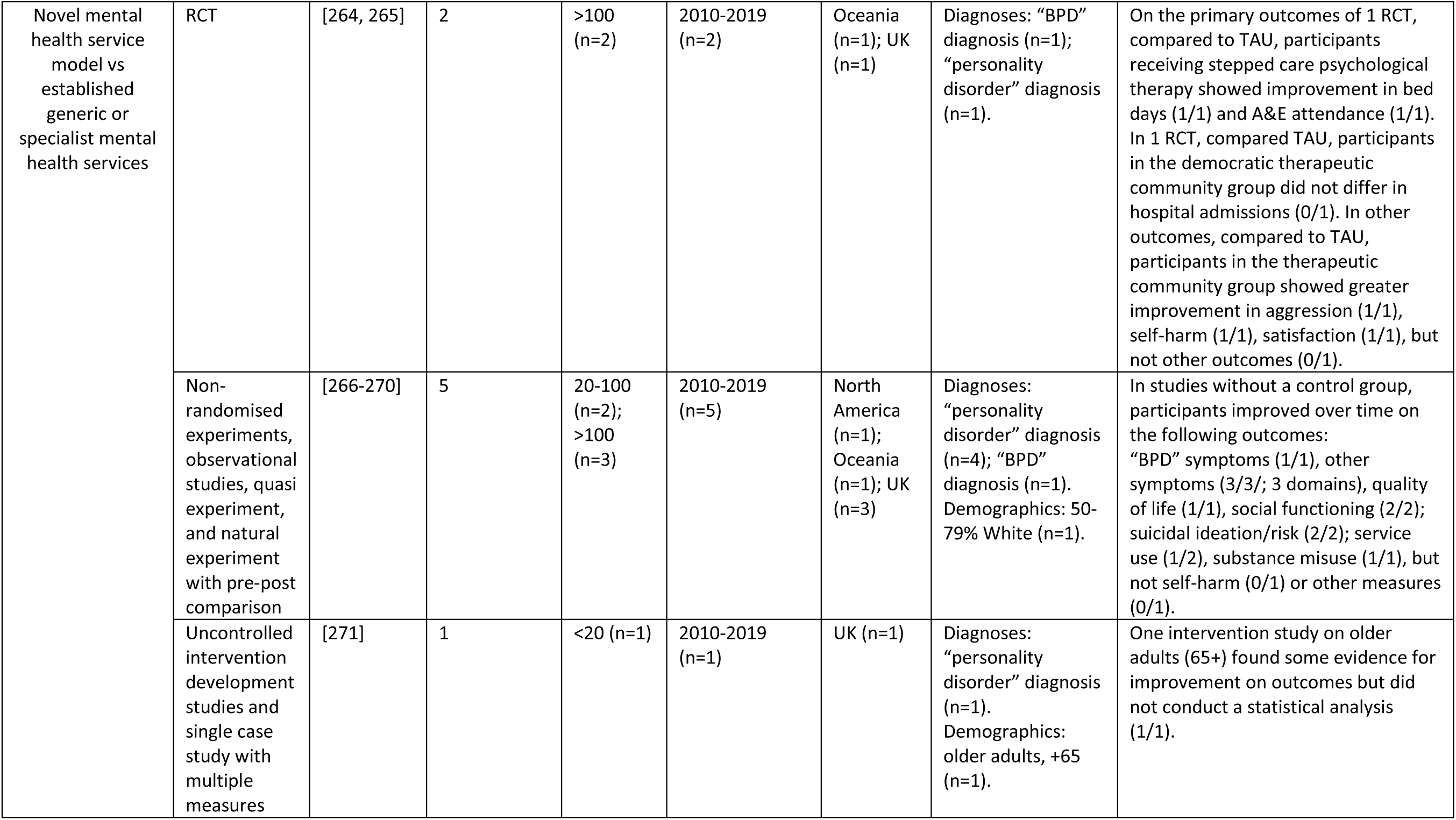
Other studies

Included papers were published between 1989 and 2020. As shown in Figure 2, there has been a progressive increase in papers over this time, with both the number of RCTs and other study designs increasing from a very small number per year in the 1990s, to 10-20 per year from 2010 onwards. As shown by Figure 3, studies testing psychodynamic therapy were the most frequent until 2005, with studies of Cognitive and behavioural therapies and DBT becoming the most prevalent in the last 15 years. There has also been an increase in the number of studies evaluating mixed therapeutic approaches over time. However, the number of studies exploring service models has remained very low (n=13; 2010 to 2019) (*see* Table 4).

**Figure 2.**
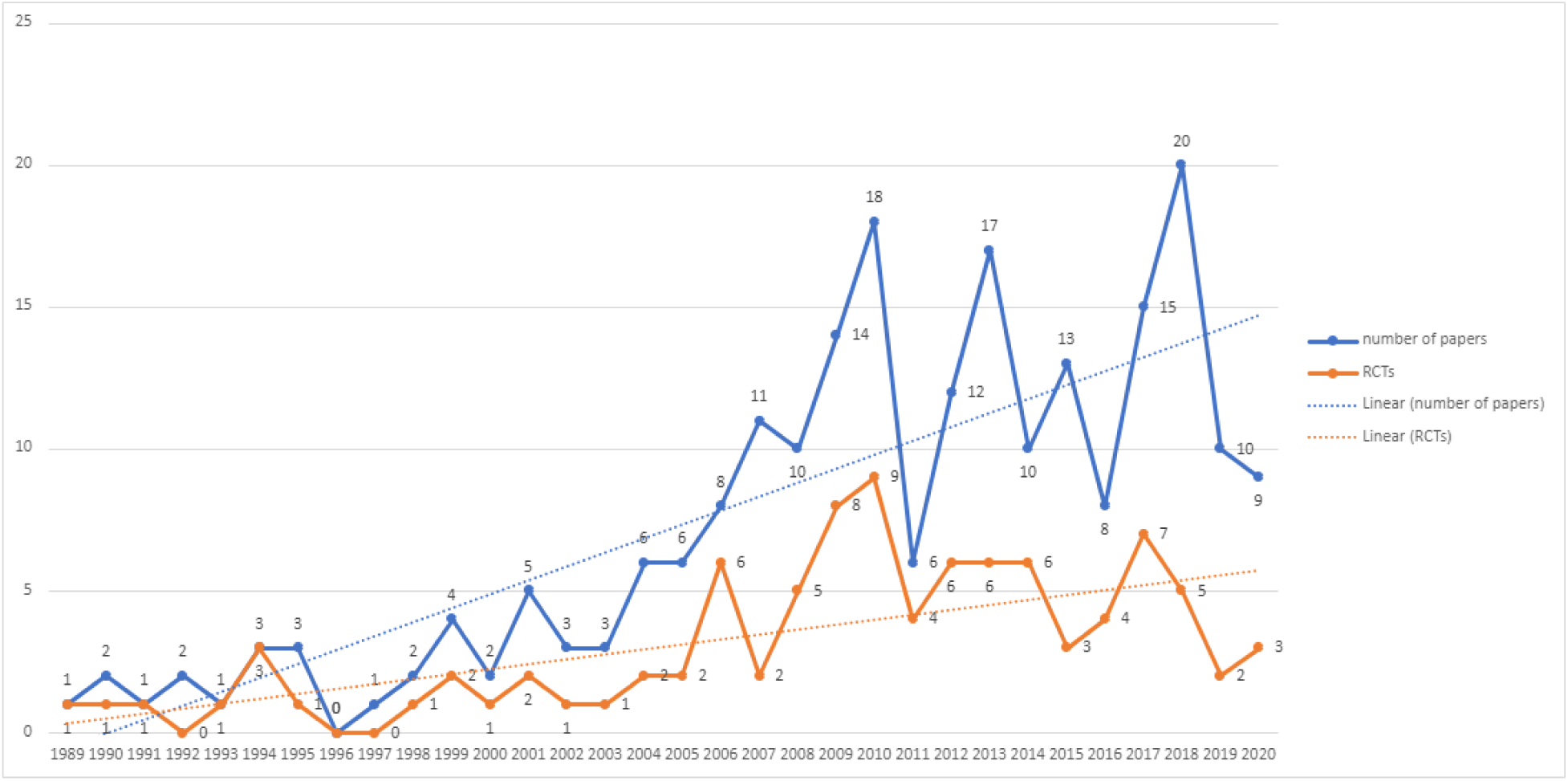
Number of Papers by Year

**Figure 3.**
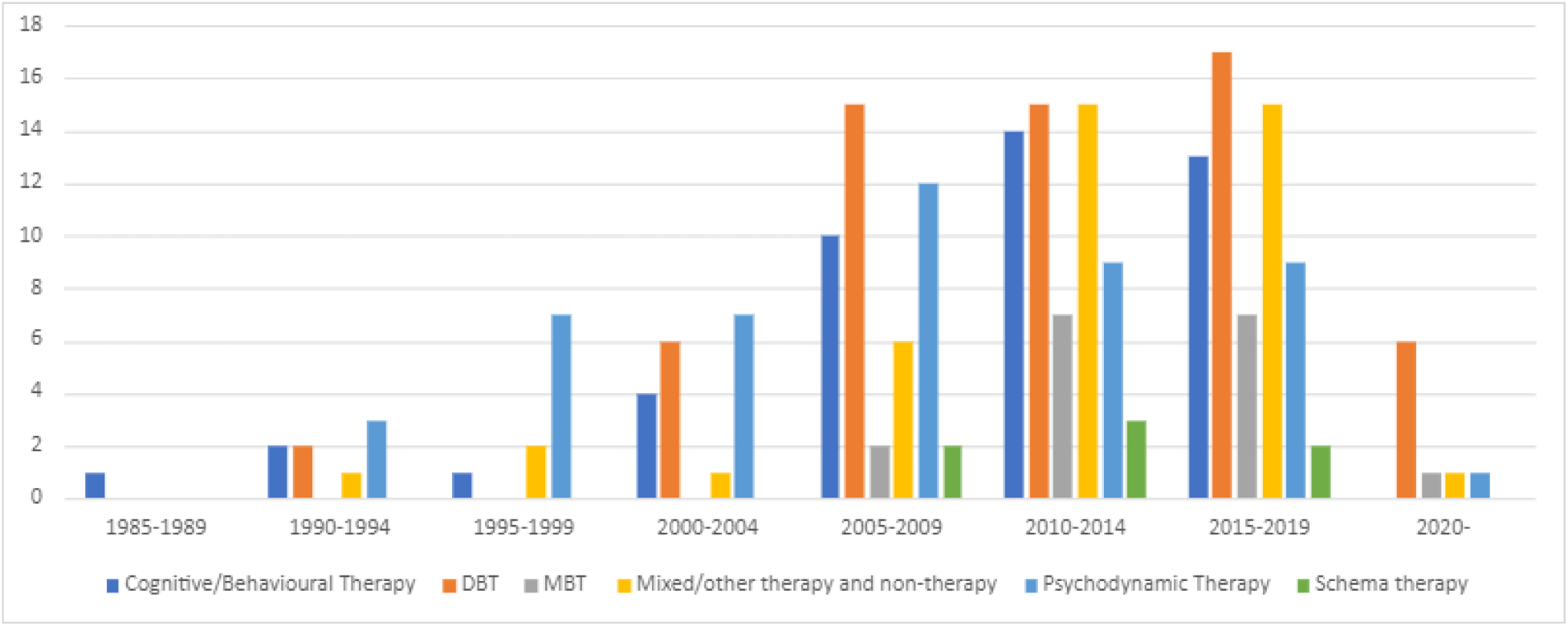
Number of Papers by Treatment Type and by Year

### Locations of interventions

Studies were conducted in a range of countries across Asia (n=5), Europe (European countries other than the UK) (n=95), North America (n=59), Oceania (n=21), and the UK (n=44). Two studies were conducted in two different continents. DBT studies made up around half of all studies conducted in North America (n=26) and Oceania (n=9), but a much smaller proportion in Europe (n=21), the UK (n=7), and Asia (n=3). Cognitive and behavioural therapies and schema studies made up around a third or more of studies in Asia (n=2) and the UK (n=15), but a lower proportion in Europe (n=21), North America (n=13), and Oceania (n=3). One Cognitive and behavioural therapy study was conducted in both Europe and North America. Psychodynamic and MBT therapies also made up a third or more of studies in the UK (n=13) as well as in Oceania (n=7) and Europe (n=36), but a lower proportion elsewhere (Asia n=1; North America n=15). Two psychodynamic studies were conducted in multiple continents. Studies exploring other types of treatment were mainly conducted in Europe (n=20), followed by the UK (n=10), North America (n=9), and Oceania (n=2). Study sample sizes varied from five to 9,614, and as shown in Figure 4, these have generally increased over the last 30 years. Overall, around half to two thirds of studies of each therapeutic modality had samples between 20 and 100. Cognitive and behavioural and schema therapy studies were generally smaller (samples <20=16/55; >100=9/55), and psychodynamic and MBT therapies were larger (samples <20=3/74; >100=22/74). As shown in Figure 4 sample sizes of RCTs have also risen during this period. The mean sample size rose from 55.3 (SD=35.7) between 1990 and 1999 to 97.4 (SD=98.1) between 2010 and 2019.

**Figure 4.**
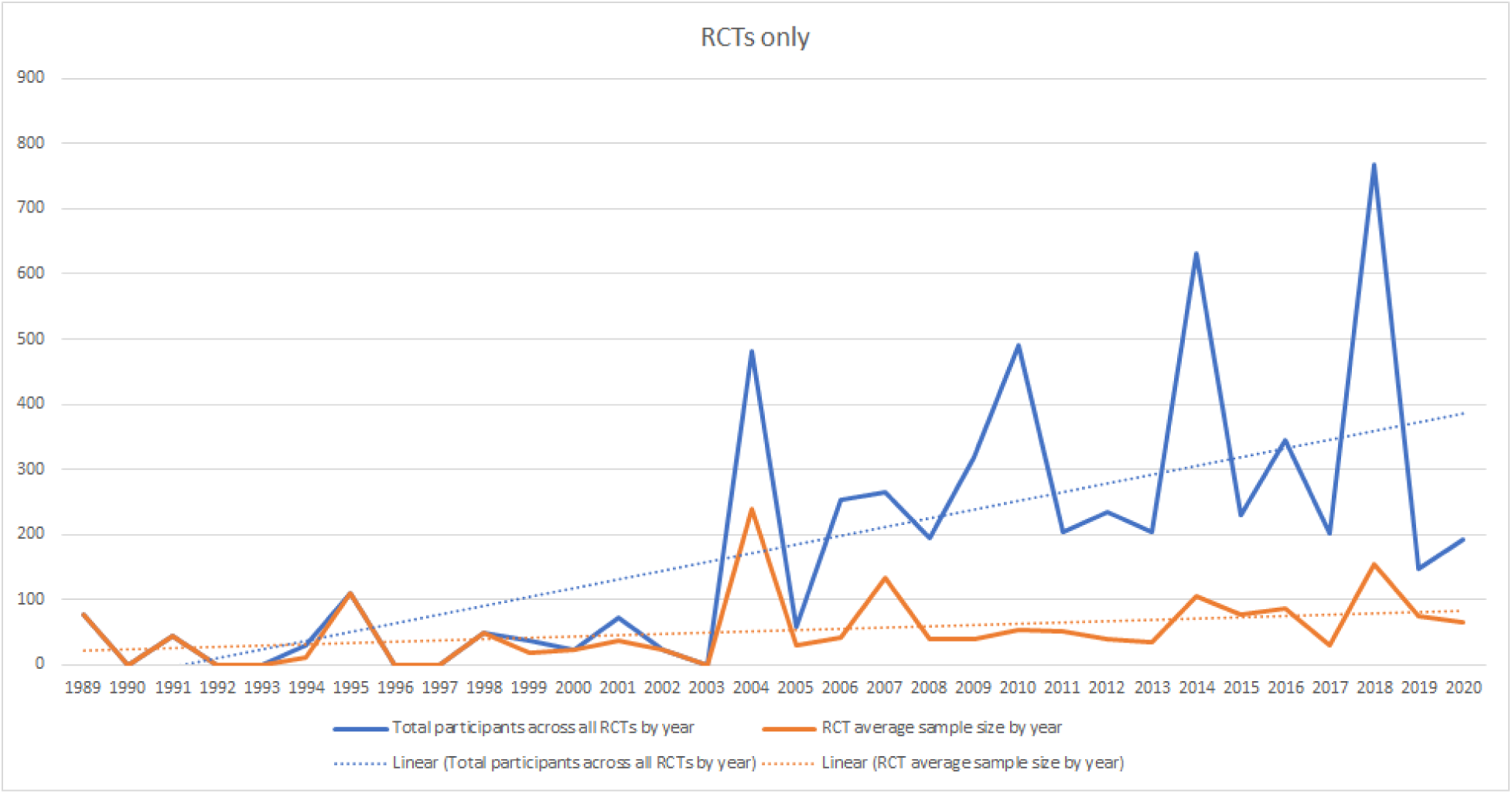
Sample Sizes of Randomised-Controlled Trials by Year

### Outcomes

Overall, “borderline personality disorder” (BPD) was the most studied condition, with 128 studies (57%) including samples that are partially or wholly given a diagnosis of “BPD”, followed by studies including participants with a mixture of “personality disorder” diagnoses (n=79, 35%). Fourteen (6%) studies did not have “personality disorder” as an inclusion criterion, but used inclusion criteria that in the judgement of the team, including clinicians, appeared to encompass similar difficulties, for example focusing on repeated self-harm or suicide attempts, complex trauma or PTSD, and emotional dysregulation or instability. These studies were included in an attempt to capture studies relevant to people with CEN in which investigators had decided not to use the “personality disorder” label as a primary way of identifying participants. “BPD” was the most studied diagnosis across treatment types, except for psychodynamic therapies and other therapies, where the largest category was studies in which participants had a mixture of “personality disorder” diagnoses. Most samples of studies that reported the sex or gender and/or ethnicity of participants were mostly female and white with thirty-nine studies including only women and thirteen studies only white participants. One study included a 100% male sample. The remaining studies included mixed samples or did not report sex or gender and/or ethnicity. 96 out of 126 studies had specified primary outcomes, including 21/65 studies on DBT, 10/20 studies on MBT, 23/54 studies on psychodynamic therapy, 24/49 studies on Cognitive and behavioural therapy, 5/6 studies on schema therapy, and 20/41 studies on other treatment. The most studied outcomes were improvement in overall symptom severity (approximately N=106), personality symptoms/functioning/diagnosis (approx. N=113), as well as other symptoms, such as anxiety, depressive, or PTSD symptoms (approx. N=115). Other commonly examined outcomes were social functioning and interpersonal symptoms and problems (approx. N=88), self-harm, suicide attempts and suicidality (approx. N = 87), service use, such as crisis service use and length and number of hospitalisations (approx. N=66), as well as quality of life (approx. N=44) and general functioning (approx. N=48). Approximately 145 studies also examined a range of other outcomes.

#### DBT interventions: findings

Table 1 and Appendix 2 summarise studies investigating the effectiveness of DBT (n=66), of which the largest group was RCTs (n=27), followed by uncontrolled studies making only pre-post comparisons (n=24), non-randomised studies with contemporaneous comparators (including quasi and natural experiments) (n=6), uncontrolled intervention development studies (n=8), and one implementation study.

Thirteen studies involved comparisons with an inactive or non-specialist treatment control, such as treatment as usual (TAU) or waitlist. Results of these studies with inactive/non-specialist comparators showed that in RCTs (n=12) and one non-randomised trial with a contemporaneous comparator (Dynamic Deconstructive Psychotherapy), DBT was superior only on some outcomes compared to controls. 6/12 of said RCTs identified primary outcomes, including self-harm, symptoms such as “BPD” symptoms, global distress, and hospital admission. DBT was superior compared to comparators on some of these primary outcomes (*see* Table 1). In one non-randomised study with contemporaneous comparators, DBT showed significantly greater improvement on the primary outcome “BPD” symptoms compared to treatment as usual (TAU), but less improvement than with Dynamic Deconstructive Psychotherapy (DDP). Sample sizes ranged from 20 to 100 for studies with primary outcomes that compared DBT with inactive/non-specialist comparators.

For studies comparing DBT with other forms of specialist psychotherapy, including General Psychiatric Management, Community Treatment by Experts, Comprehensive Validation Therapy plus 12 step programme, and clinical case management (n=8), DBT was not superior to comparators on the majority of outcomes in RCTs (n=6) and non-randomised studies with contemporaneous comparators (n=2). For studies with specified primary outcomes, DBT showed similar or less improvement in self-harm and suicidality compared to controls in 3/4 RCTs, but was superior to Community Treatment by Experts on suicide attempts in the fourth RCT. Three of these RCTs had sample sizes greater than 100.

Nineteen studies investigated partial or modified DBT therapies. In these studies, DBT was superior to comparators on some outcomes in RCTs (n=6), including three RCTs with sample sizes greater than 100 and one pilot RCT, but inferior to controls on outcomes in one non-randomised trial with a contemporaneous comparator. No study that investigated partial or modified DBT therapies had both a specified primary outcome and a control group (n=19).

Across studies reporting pre-post-treatment comparisons but no comparison with a control group (n=31), participants improved over time on all or almost all outcomes.

#### DBT interventions: Adaptions to specific populations

Seven of the above studies focused on delivery of DBT to samples defined other than solely by a “personality disorder” diagnosis, including investigations of outcomes of DBT for individuals with a “BPD” diagnosis and comorbid severe mental illness (n=1) or substance use disorder (n=2). Four DBT studies used criteria other than “personality disorder” diagnosis, including emotional dysregulation (n=1), parasuicidal behaviours in the past six months (n=1), and severe difficulty in functioning together with frequent suicide attempts (n=1) or crisis service use (n=1). These studies included one RCT, one intervention development study and five studies involving only pre-post comparisons.

Additionally, a total of eight studies examined the effectiveness of DBT adapted for specific clinical or demographic populations. Adapted DBT treatments included DBT-PTSD (n=2) and DBT with prolonged exposure for people with comorbid PTSD (n=1), DBT for people with comorbid eating disorders (n=1), DBT for young adults aged 18 to 25 years (n=2), mother-infant DBT for female primary caregivers of children younger than 3-years-old (n=1), and couples DBT for 18-50-year-old married men (n=1). The studies included RCTs (n=3) including one pilot RCT), intervention development studies (n=2), and non-randomised studies with contemporaneous comparators (n=2) or only making pre-post (n=1) comparisons. In one RCT including 193 participants, DBT-PTSD was superior to Cognitive Processing Therapy (CPT) for participants with complex PTSD and a history of childhood abuse, on the primary outcome, PTSD diagnosis. Additionally, DBT was superior to CBT on some primary outcomes in one non-randomised controlled study including 118 participants. DBT was superior to controls on all non-primary outcomes across RCTs (n=3) and most non-primary outcomes across non-randomised controlled studies (n=2): sample sizes for these studies without specified primary outcomes varied from 11 to 37.

#### Cognitive and behavioural and Schema interventions: Findings

Table 2 and Appendix 3 present study characteristics and findings of Cognitive and behavioural and schema therapies (n=55). There were 26 RCTs, 17 uncontrolled intervention development studies, three non-randomised studies with contemporaneous controls and nine uncontrolled studies making only pre-post-treatment comparisons.

Nineteen studies had inactive/non-specialist comparators. In RCTs (n=19 including 4 pilot studies), compared to inactive/non-specialist controls, participants receiving Cognitive and behavioural or schema therapy showed improvement on some outcomes. 12/19 RCTs had specified primary outcomes, with sample sizes ranging from 34 to 480. Cognitive and behavioural or schema therapy was superior compared to controls on primary outcomes in some studies, including for “personality disorder” symptoms (n=3), “recovery” (n=1) and symptom severity and social functioning in 1/2 RCTs. Cognitive and behavioural or schema therapy was not superior for primary outcomes in other studies, including for depressive or (social) anxiety symptoms, and service use and/or self-harm (*see* Table 3).

In studies making only pre-post comparisons with no control groups (n=23), participants showed improvement on almost all outcomes over time.

In studies with specialist treatment comparators, including Rogerian Supportive Therapy, Transference-Focused Therapy, Dynamic psychotherapy, group-based CBT, individual Cognitive-Evolution Therapy, Mindful Emotion Awareness and Cognitive Reappraisal, and different treatment settings (n=7), Cognitive and behavioural therapy was inferior to or showed similar improvements to control treatments for all outcomes in RCTs (n=4) and non-randomised studies with contemporaneous comparators (n=3). This included the results of three RCTs and two non-randomised studies with contemporaneous controls with specified primary outcomes (“BPD” symptoms, symptom severity, personality functioning and interpersonal problems).Sample sizes of studies with primary outcomes ranged from 46 to 205.

Lastly, three RCTs, including one pilot RCT, examined modifications of Cognitive and behavioural or schema therapies. These interventions were not superior to comparators on any outcomes. One RCT found no difference between schema therapy with and without phone support on “BPD” recovery as a primary outcome. This was also the case for a Manual-Assisted Cognitive Therapy (MACT) with and without a therapeutic assessment intervention. One RCT investigating a cognitively-based problem-solving treatment delivered at home obtained mixed findings on the primary outcome recovery from “BPD”. The 2/3 RCTs with identified primary outcomes included 20 and 62 participants.

#### Cognitive and behavioural and schema treatments: Adaptions to specific populations

Of the above studies ten examined the effectiveness of Cognitive and behavioural treatments for clinical populations with “personality disorder” diagnoses and comorbid mental health problems, or individuals without a formal “personality disorder” diagnosis but CEN. These studies looked at individuals with experiences of childhood sexual abuse (n=1), “BPD” symptoms and comorbid substance use disorder (n=1), (longstanding or treatment-resistant) mood disorder or chronic adjustment disorder and a comorbid “personality disorder” or “BPD” diagnosis (n=3), non-suicidal self-injury (NSSI) disorder (n=1), previous and predicted suicide attempts (n=1), repeated self-harm (n=1), and a “BPD” or cluster B “personality disorder” diagnosis or transversal or longitudinal comorbidity (n=1). One study included participants with various diagnoses, including “personality disorder” diagnoses. Studies included RCTs (n=3), non-randomised trials with contemporaneous comparators (n=1) or pre-post comparisons (n=1), observational studies with contemporaneous comparators (n=1), intervention development or uncontrolled feasibility studies (n=2), single case studies (n=2).

#### Psychodynamic and MBT interventions: Findings

Table 3 and Appendix 4 summarise studies investigating the effectiveness of MBT (n=20) and psychodynamic interventions (n=54). There were 25 RCTs, and 48 non-randomised studies, which included non-randomised studies with contemporaneous controls (n=17) and studies without control groups making only pre-post comparisons (n=31). One uncontrolled study focused on intervention development. Four RCTs compared MBT with an inactive/non-specialist treatment control (as did a non-randomised study comparing with a historical cohort). In the RCTs sample sizes ranged from 38-51 and MBT was superior to the control on most outcomes. Two RCTs specified primary outcomes, and MBT proved superior in reducing both “BPD” symptoms (n=1) and suicide attempts (n=1) compared to controls, though MBT did not reduced “BPD” symptoms more than DBT.

For studies comparing MBT with other forms of specialist treatment, including specialist TAU, supportive group therapy, structured clinical management, and DBT, (n=9), MBT showed no significant difference in most outcomes in three of four RCTs with sample sizes ranging from 95-111, with the fourth reporting greater improvements in the primary outcome of parasuicidal behaviours compared with structured clinical management. In non-randomised studies, results were mixed with few additional benefits of MBT reported. In primary outcomes, one study reported similar reductions in bed days to the specialist treatment comparator.

One RCT comparing MBT settings (sample size 114) found no differences on primary or secondary outcomes between MBT at a day hospital compared to an intensive out-patient MBT.

In seven MBT studies, pre-post-treatment comparisons were made without a control group: improvements were reported on most or all outcomes.

Thirteen studies on psychodynamic treatments had inactive/non-specialist comparators including six RCTs and seven non-randomised studies. Participants receiving psychodynamic therapy showed greater improvements compared to inactive/non-specialist comparators in the majority of outcomes in RCTs (which included sample sizes of 50-100 and <50 in two studies) and close to all outcomes in non-randomised studies with control groups. Improvement in the primary outcome was reported in 2/3 RCTs and all non-randomised studies who specified one compared to the comparator (*see* Table 3).

In studies with specialist comparators (n=11), including manual-based Psychiatric-Psychodynamic sessions, General Psychiatric Management, cognitive therapy, and Transference-Focused Therapy plus supportive treatment, the intervention group was superior compared to the control group only on some outcomes in RCTs (n=8), but most outcomes in non-randomised studies (n=3). Of those specifying primary outcomes, only 2/5 of RCTs (both with sample sizes <50) reported greater improvements in suicidality (n=1) or progress in therapy (n=1) than for other specialist treatments, and 1/2 non-randomised studies reported greater improvement in “personality disorder” symptoms for DDP compared to controls and DBT.

Participants improved on almost all or all outcomes over time in studies with pre-post designs (n=11).

Six non-randomised studies compared the outcomes of psychodynamic therapy delivered in different settings. There was no difference in outcomes in studies comparing day hospital, outpatient, and inpatient services, however, community and step-down services were superior to residential services on all outcomes, including the primary outcome of symptom severity in all three studies with an identified primary outcome. Lastly, participants receiving adapted psychodynamic treatments showed greater improvement compared to controls on most outcomes, including the primary outcomes of “BPD” severity or parasuicide behaviour in RCTs (n=2) and one non-randomised study with a contemporaneous control.

#### Psychodynamic and MBT interventions: Adaptions to specific populations

Of the above studies, six looked at adaptations for specific populations including for individuals with poor personal, social and/or interpersonal functioning with or without “personality disorder” diagnosis (n=4), “personality disorder” diagnosis with or without comorbid alcohol use disorder (n=1), and treatment-resistant depression with comorbid “personality disorder” diagnoses and histories of early childhood trauma (n=1).These studies included one RCT and five studies with pre-post comparisons.

Additionally, a total of three studies examined the effectiveness of psychodynamic treatments that were adapted to specific clinical or demographic populations, including two RCTs investigating interventions adapted for people with a “BPD” diagnosis and active alcohol use or dependence and one non-randomised trial with a contemporaneous comparator adapted to people with a “BPD” diagnosis in an underserved community-based outpatient setting. Compared to controls, the intervention group was superior on the majority of outcomes, including all primary outcomes (“BPD” symptom severity, parasuicidal behaviour, alcohol misuse and institutional care) identified in the two RCTs, which both included 30 participants.

#### Other interventions: Findings

Table 4 and Appendix 5 present studies on any treatment type other than the psychotherapies listed above (n=41). These included studies of mixed therapeutic modalities (n=10), other individual therapies (n=7), social-interpersonal and functional therapies (n=5), self-management and care planning interventions (n=6), as well as studies investigating outcomes of different approaches to service design and delivery (n=13). Most studies were RCTs (n=25), while three studies made comparisons with contemporaneous control groups, and 13 only pre/post comparisons.

In RCTs with inactive/non-specialist comparators examining mixed therapeutic modalities, the intervention group was superior to controls on most outcomes (n=3), including the primary outcomes drop out and suicide attempts, but not “BPD” symptoms of the two RCTs with identified primary outcomes.

Compared to controls, participants receiving individual therapies other than the psychotherapies listed above (including Art therapy, Abandonment psychotherapy, Body Awareness Group therapy, short-term psychotherapy, and psychoeducation) showed greater improvements in close to all outcomes in RCTs with inactive/non-specialist comparators (n=5 including one pilot RCT). However, in the two RCTs with specified primary outcomes Abandonment psychotherapy was superior to control for suicidal relapse and hospitalisation (n=1), but psychoeducation was not superior to control for “BPD” severity (n=1). Other individual therapies were not superior to controls in two RCTs with specialist treatment comparators, including Abandonment psychotherapy delivered by nurses instead of trained psychotherapists, Transference-Focused Therapy and DBT, on all outcomes including primary outcomes. Sample sizes of RCTs with primary outcomes ranged from 50 to 170.

Similar results were found for social and interpersonal interventions, with the intervention group being superior compared to controls on up to half of the outcomes in RCTs with inactive/non-specialist comparators (n=3). Additionally, the intervention group was superior on primary outcomes in only 1/2 RCTs with identified primary outcomes: Psychoeducation plus problem-solving therapy showed greater improvement in social functioning and social problem-solving skills compared to waitlist, however, the cognitive rehabilitation and psychoeducation groups improved similarly in general functioning. RCTs with primary outcomes included 70 and 176 participants. There were no between-group differences found in RCTs with specialist comparators, including delayed Functional Imagery Training and Social Skills Training in the clinic/hospital only (n=2 including one pilot RCT).

RCTs on self-management and care planning compared to self-management (n=2) or established generic or specialist mental health services (n=1) found no between-group differences on outcomes. This included the primary outcomes of two RCTs: The Joint Crisis Plan group and TAU group had similar rates of self-harm in one RCT including 88 participants. The second RCT with a sample size of 52 found no difference in service admissions in Nidotherapy enhanced assertive outreach compared to standard assertive outreach.

Regarding service design models, one RCT comprising four papers comparing step-down treatment with outpatient treatment showed no between-group differences on outcomes, including a range of primary outcomes (*see* Table 4). In the other RCT, the step-down intervention group was superior compared to the outpatient group on half of the outcomes. Lastly, two RCTs with samples >100 examining novel mental health service models compared to established generic or specialist mental health services found the intervention group to be superior on most outcomes compared to the control group, but on primary outcomes related to service use only in 1/2 RCTs.

Across treatment types, participants improved on most or all outcomes over time in studies with pre-post comparisons (*see* Table 4).

#### Other treatments: Adaptions to specific populations

Six of the above studies on other treatments examined specific populations, including three RCTs, one non-randomised study with a contemporaneous control, and one uncontrolled study making only pre-post comparisons. One RCT compared the effectiveness of Abandonment psychotherapy and intensive TAU for individuals with major depression and a comorbid “BPD” diagnosis. Another RCT investigated a joint crisis plan and TAU for young people without a “personality disorder” diagnosis but at least two episodes of self-harm in the previous three months. A third RCT compared the effectiveness of Nidotherapy and TAU for individuals with severe mental illness and a comorbid “personality disorder” diagnosis. One non-randomised study examined collaborative care management and TAU for individuals with major depression with or without a comorbid “personality disorder” diagnosis. Lastly, an uncontrolled study investigated emotion regulation skills training for a community-based sample of individuals with adverse childhood experiences over time.

## Discussion

Our scoping review collated quantitative evidence regarding community-based psychological, psychosocial, and service level interventions designed for people with CEN. Most studies focused on people given a “personality disorder” diagnosis, with a small number relating to people who appeared to have comparable difficulties (6%). Some observations may be made from this literature, but large gaps are prominent.

### What *does* the literature tell us?

We identified 226 papers reporting on 210 distinct studies carried out in a range of countries, the majority in Europe or North America. The largest group of studies evaluated the effectiveness of DBT, followed by psychodynamic therapy, Cognitive and behavioural therapy, MBT, and schema therapy. Research on psychological treatments dominated, with only a small handful of studies using any method to investigate interventions with primarily social targets, self-management, care planning or models of service delivery.

The total quantity of studies, given the breadth of the search and inclusion of uncontrolled studies and studies with very small samples, is small. Little literature was published in the 20^th^ century, with most included studies published after 2005, since when annual publication rates have slowly risen. This may reflect to reflect a shift internationally away from the view of “personality disorder” as untreatable and justifying exclusion from mental health services that prevailed in the 20^th^ century [32]. In the early 2000s, factors including the publication of trials that held out prospects for successful treatment, service user activism, and key policy documents such as the UK’s “Personality Disorder: No longer a diagnosis of exclusion” may have contributed to greater confidence that research in this area is potentially fruitful [6, 33, 34]. However, stigma, therapeutic pessimism and difficulty accessing any kind of helpful care are still widely reported [10-12, 20, 35]. The results of our searches suggest that investment in large well-designed studies that test clear primary hypotheses has remained very limited around the world, which may reflect a continuing lack of optimism, and the impacts of the particularly severe stigma that appears associated with CEN.

The evidence base that has been established thus far relates mainly to specialist psychotherapies, delivered especially to people with a “BPD” diagnosis. Many studies are small and/or non-randomised, but studies with any methodology have tended to suggest benefits for specialist psychotherapies of a range of types compared with inactive/non-specialist controls, both in studies focused on people with a “BPD” diagnosis and with broader groups. However, results do not tend to suggest one kind of specialist treatment is clearly superior to another – this coheres with the results of more narrowly focused systematic reviews that do not identify a clear gold standard but suggest a variety of psychological treatments are helpful for those who engage with them [23, 26]: a focus on what works well for whom, and why, would be helpful in further work.

Contrary to the pessimistic outlook often reported regarding potential for improvement among people with a “personality disorder” diagnosis, a large majority of studies involving before and after comparisons find significant reductions in symptoms and self-harm as well as improvements in other outcomes. This seems to be the case across treatment types as well as diagnoses, often to the extent that a substantial minority of participants were assessed as no longer meeting criteria for a “personality disorder”. Study methods often made it hard to assess how far this was a result of treatments received, including those being investigated, and how far of the natural improvement in symptoms and difficulties (people may also tend to be recruited to studies at times when difficulties are especially severe). Findings from these studies suggest the value of uncontrolled studies and of before and after treatment comparisons is very limited except where the main purpose is to test the feasibility and acceptability of delivering an intervention: it appears likely that improvement will be found whatever interventions are offered.

Regarding specific populations such as those who are younger or older or who have some of the conditions that are frequently comorbid with CEN, such as substance misuse or psychosis, we found substantial numbers of interesting small studies, mainly aimed at intervention development, or establishing that treatments are feasible and acceptable in specific populations. These provide potential building blocks for further design and testing of interventions in important populations where substantial trials have yet to be reported.

### What does the literature *not* tell us?

Gaps in the evidence needed to underpin high quality service delivery for people with CEN are large. Service users and clinicians report that mental health care systems appear ill-equipped to deliver accessible care of high quality [10, 15, 20], yet there are hardly any published investigations of the best approaches to designing teams and systems. Care planning, crisis planning, and self-management are to a large extent not investigated as applied to people with CEN. We identified very few studies of interventions with social targets, including employment and social relationships, even though people with CEN identify these as a priority [11, 36]. We found very little evidence of co-production or service user leadership in either research or intervention design, despite the benefits of these in producing research that aligns to service user needs and priorities [37]. We also found very little quantitative research on either trauma-informed care for this group, or interventions for people with comorbid PTSD, despite calls to place trauma at the centre of thinking about CEN [11, 35, 38].

Only a few studies evaluated treatments adapted to specific populations of interest, such as younger or older age groups, parents or patients with comorbid severe mental illnesses, substance misuse, or childhood trauma. As above, a number of small-scale initial studies appeared promising, but were limited by small sample sizes and/or observational or feasibility/intervention development study designs. Lack of more substantial evaluations of well-designed interventions for these groups who have tended to be still more under-served than others with CEN appears an important gap.

Most studies were conducted with participants with a “BPD” diagnosis, so that there is little evidence on effective interventions for people with other diagnoses, or who may have comparable difficulties but not have received a diagnosis. Samples are largely White and female samples with close to no papers focusing on diverse gender and sexual identities (despite some evidence of LGBTQ+ groups being more likely to receive a “personality disorder” diagnosis [39, 40]), or other ethnicities. Studies generally measured effectiveness of interventions by examining improvement in whether diagnostic criteria continued to be met for “personality disorder”, symptom outcomes, self-harm, and service use. However, outcomes prioritised by service users such as personal achievements, employment, and social connections were reported much less [36], and the possibility of iatrogenic harm was also rarely examined. Interventions addressing social needs are especially important in the light of findings of longitudinal studies showing that while symptoms and suicidal behaviour tend to improve with time, this is less the case for psychosocial functioning including rate of employment [41, 42]. Implementation studies examining how to embed successful interventions in real-world settings were also largely absent.

### Limitations

Despite the breath of our approach, the findings of the present review must be considered in light of several limitations. In order to provide an overview of evidence acquired so far and identify gaps, we have created broad, often heterogenous, categories of study designs. This approach is inevitably superficial and limits how far meaningful comparisons can be made across study types, treatments, and subpopulations. In keeping with scoping review methodology recommendations, we did not formally assess the quality of the studies, although we have commented on some obvious limitations, for example relating to small trial populations or uncontrolled study designs.

Additionally, while inclusion criteria were kept broad, and a variety of search terms applied to try to include studies with participants with any diagnosis of “personality disorder” as well as those with comparable difficulties, capturing the latter reliably is likely to have been particularly difficult, and only a small number of studies not based on such criteria were included. We also have not included many studies that are transdiagnostic or include mixed populations of mental health service users Lastly, in order to make this very broad search feasible, we included only studies published in English and this may well have excluded some relevant evidence, although the number of studies excluded on grounds of language was small.

### Conclusions

Our overall conclusion from this scoping review is that people with CEN, despite being numerous among community mental health service users [43] have thus far been poorly served by clinical research. Mental health research is in general under-funded compared with other areas of health [44]. Our findings suggest that this is especially striking in the field of CEN, in which little was published prior to 2005 and the tally has increased only gradually subsequently, now only just exceeding two hundred quantitative studies including 96 RCTs of community and outpatient interventions, even including studies of any scale using any method.

Much therefore needs to be done to develop a robust evidence base in this area, especially beyond a narrow focus on specialist psychotherapies for people with a “BPD” diagnosis, where a substantial number of trials have resulted in a finding that several specialist therapies appear better than treatment as usual, but not in a clear finding that any treatment is clearly superior. Future research should address outcomes valued by patients rather than being limited to a focus on self-harm and symptoms: relevance to service users is much more likely to be achieved by the adoption of co-production in design of both interventions and research studies. The recent service user-led StopSIM campaign against the Serenity Integrated Monitoring intervention [45], which involved police in response to some people with frequent contact with emergency services, exemplifies the potential for iatrogenic harm from interventions that are unevaluated, or where the potential for harm has not been assessed. Research on important populations such as older and younger people and people with major comorbidities, and on interventions focusing on people with CEN as parents, partners or relatives needs to progress beyond the feasibility studies conducted so far. Larger and more diverse samples are needed to be confident of relevance across service user populations.

Models of service delivery have been largely neglected in research so far despite recurrent complaints from service users and clinicians that current systems are fragmented and inaccessible. Realist evaluations may shed a light on what mechanisms underly the effectiveness of different interventions as well as what type of intervention works for which patient group and in what context. Relevant contexts may be individual, such as personal life and stage of life, as well as systemic. Additionally, services need to deliver holistic and person-centred care that addresses service users’ interconnected needs and intersecting experiences over several years: large-scale observational designs may be helpful in understanding outcomes over longer periods [10, 11]. Lastly, patients and carers with relevant experiences need to be invited to co-produce the development and evaluation of treatments to not only ask the right questions but also examine these in a meaningful way.

#### Box1 Lived experience commentary written by Sarah Labovitch and Jennie Parker

In light of the Community Mental Health Framework (CMHFA), this review is well timed to revise thinking around what *should* be available to people who may meet the diagnostic criteria for “personality disorder”/CEN. It may also prompt researchers and service-providers to consider what is important to us - it was disappointing to see that only 44/226 studies reported on quality of life, whilst most primary outcomes focused on diagnostic-related criteria.

Time to follow-up in many studies discussed is limited. Side-effects of funding constraints typically lead to quantitative research and RCTs being prioritised. We agree with the question of what underlies reported improvements, and would say this is not just in relation to observational studies. It would be interesting to delve further into this.

Despite advancements in recent years, community service-provision for “personality disorder”/CEN is nevertheless lagging behind other areas of mental health. Treatment in the community must be patient-centred: adapted to factors such as age, culture, comorbidity, substance misuse and trauma. Some health professionals still display discriminatory attitudes towards CEN, or simply don’t know how to help. Finding a clinician with the right skills and compassion is depressingly arduous. Further, exclusion criteria and high thresholds can make “specialist” services inaccessible. Meanwhile, the notion of individuals actually having a choice in therapist is vanishingly slim, adding to the risk of iatrogenic harm and a “cliff-edge” of care. Services need to commit to consistent long-term contact, as well as tailoring treatment to individual needs.

As with others, we have experienced stigma, rejection, and repeated/inappropriate referrals This paper leaves us with a conundrum, both in relation to the integrated approach proposed by the CMHFA and access to good and timely support. Whilst this is a scoping review of quantitative research, our recommendation is for further investigation into the active ingredients of therapy: what makes good outcomes for some but not others, the importance of the relationship, and whether we have a choice of therapist (considerate of age, culture, gender, etc) or of intervention. We also noted the limited research on peer support, compared to our experience of its value. With such a diverse population and diverse range of therapies (and variance within specific models), clearer guidance would be helpful so that we can all make fully-informed choices.

## Supporting information

PRISMA checklist

## Data Availability

All data produced in the present work are contained in the manuscript (see appendices).

