## Appendices for "Current state of the evidence on community treatments for people with complex emotional needs: a scoping review"

### Table of contents

Appendix 1 – search strategy ........... p. 2

Appendix 2 – Table of studies testing Dialectical Behavioral Therapy treatments …. p. 7

Appendix 3 – Table of studies testing Cognitive and Behavioral Therapy and Schema Therapy treatments … p. 37

Appendix 4 – Table of studies testing MBT and Psychodynamic Therapy treatments …. p. 63

Appendix 5 – Table of studies testing other treatments …. p. 105

### Appendix 1 – Search Strategy overview and MEDLINE example

| | |
|---|---|
| Ovid MEDLINE(R) and Epub Ahead of Print, In-Process & Other Non-Indexed Citations and Daily | 3984 |
| Embase | 6002 |
| Social Policy and Practice (SPP) | 402 |
| PsycINFO | 5285 |
| HMIC | 218 |
| CINAHL | 1620 |
| total | 17511 |
| - duplicates | 10690 |
| | 6821 |
| UPDATE SEARCH | |
| Ovid MEDLINE(R) and Epub Ahead of Print, In-Process & Other Non-Indexed Citations and Daily | December 10th 2019-November 23 2020: 409 |
| Embase | December 11th 2019-November 23 2020: 918 |
| Social Policy and Practice (SPP) | December 1 2019 – November 23 2020: 82 |
| PsycINFO | December 11th 2019-November 23 2020: 430 |
| HMIC | January 1st 2019-November 23 2020: 4 |
| CINAHL | December 1st 2019-November 23 2020: 25 |
| total | 1868 |
| - duplicates | 540 |
| | 1328 |

Database: Ovid MEDLINE(R) and Epub Ahead of Print, In-Process & Other Non-Indexed Citations and Daily
Host: Ovid
Data Parameters: 1946 to December 10, 2019

Date of search: Wednesday 11th December 2019
Search strategy:

Search Strategy:

| # | Searches | Results |
|---|---|---|
| 1 | exp *Personality Disorders/ | 28263 |
| 2 | ((personality or character*) adj3 disorder$).ti,ab,kw. | 65164 |
| 3 | "axis II".ti,ab,kw. | 1959 |
| 4 | ("Complex trauma" or CPTSD or "complex post-traumatic stress disorder").ti,ab,kw. | 535 |
| 5 | (Complex adj (needs or mental)).ti,ab,kw. | 1929 |
| 6 | *Self-Injurious Behavior/ | 5465 |
| 7 | (Self-harm or self-injury).ti,ab,kw. | 7484 |
| 8 | (emotion* adj2 (regulation or dysregulation or unstable or instability)).ti,ab,kw. | 10588 |
| 9 | mood instability.ti,ab,kw. | 254 |
| 10 | 1 or 2 or 3 or 4 or 5 or 6 or 7 or 8 or 9 | 103674 |
| 11 | Community Health Services/ | 31050 |
| 12 | Community Mental Health Services/ | 18317 |
| 13 | ((commun$ adj5 (mental health or model$1 or pathway$1 or program$ or evaluat$ or intervention$ or implement$)) or camhs or cmht$1).ti,ab,kw. | 79584 |
| 14 | (community adj5 (agenc$ or care or center$ or centre$ or clinic$ or consultant$ or doctor$ or employee$ or expert$ or facilitator$ or healthcare or instructor$ or leader$ or manager$ or mentor$ or nurs$ or personnel$ or pharmacy or pharmacist$ or psychiatrist$ or psychologist$ or psychotherapist$ or specialist$ or skill$ or staff$ or team$ or therapist$ or tutor$ or visit$ or worker$ or group$ or independent or (peer$ adj3 support$) or survivor or outpatient$ or "out patient$")).ti,ab,kw. | 96749 |
| 15 | (commun$ adj5 (service or hub$ or based or deliver$ or interact$ or led or maintenance or mediat$ or operated or provides or provider$ or run or setting$ or support or rehab$ or therap$ or service$ or treatment or management or assessment or assistance or care or day or week)).ti,ab,kw. | 205151 |

| | | |
|---|---|---|
| 16 | (Independent sector or ((non institutional$ or noninstitution$) adj2 (sector$ or setting$))).ti,ab,kw. | 367 |
| 17 | (network or outreach or ((specialist or day or whole) adj3 service)).ti,ab,kw. | 351098 |
| 18 | ((treatment* or (Dialectical behavior therapy or Dialectical behaviour therapy or DBT) or Psychotherapy* or specialist or psychiatry* or therapeutic or day or outreach or therap*) adj3 (Outpatient* or community or Service* or Center* or Centre* or Clinic*1 or Team* or program* or provider* or practice or setting* or care or community or unit* or hospital*)).ti,ab,kw. | 244595 |
| 19 | 11 or 12 or 13 or 14 or 15 or 16 or 17 or 18 | 851066 |
| 20 | Interview*.af. | 373632 |
| 21 | Experience*.af. | 1028333 |
| 22 | qualitative.tw. | 212806 |
| 23 | Qualitative Research/ | 50164 |
| 24 | 20 or 21 or 22 or 23 | 1444445 |
| 25 | randomized controlled trial.pt. | 495635 |
| 26 | controlled clinical trial.pt. | 93449 |
| 27 | (randomized or randomised).ab. | 553632 |
| 28 | placebo.ab. | 203251 |
| 29 | clinical trials as topic.sh. | 189357 |
| 30 | randomly.ab. | 322897 |
| 31 | trial.ti. | 209094 |
| 32 | 25 or 26 or 27 or 28 or 29 or 30 or 31 | 1289520 |
| 33 | Epidemiologic studies/ | 8156 |
| 34 | exp case control studies/ | 1037554 |
| 35 | Case control.tw. | 120110 |
| 36 | (cohort adj (study or studies)).tw. | 190004 |
| 37 | Cohort analy$.tw. | 7484 |

| | | |
|---|---|---|
| 38 | (Follow up adj (study or studies)).tw. | 47969 |
| 39 | (observational adj (study or studies)).tw. | 98982 |
| 40 | Longitudinal.tw. | 232846 |
| 41 | Retrospective.tw. | 497909 |
| 42 | Cross sectional.tw. | 329612 |
| 43 | Cross-sectional studies/ | 311409 |
| 44 | 33 or 34 or 35 or 36 or 37 or 38 or 39 or 40 or 41 or 42 or 43 | 2028189 |
| 45 | 32 or 44 | 3197720 |
| 46 | "Surveys and Questionnaires"/ | 443562 |
| 47 | survey$.tw. | 612248 |
| 48 | exp clinical pathway/ | 6469 |
| 49 | exp clinical protocol/ | 163100 |
| 50 | exp consensus/ | 11712 |
| 51 | exp consensus development conference/ | 11685 |
| 52 | exp consensus development conferences as topic/ | 2772 |
| 53 | critical pathways/ | 6469 |
| 54 | exp guideline/ | 33000 |
| 55 | guidelines as topic/ | 38818 |
| 56 | exp practice guideline/ | 26125 |
| 57 | health planning guidelines/ | 4067 |
| 58 | (guideline or practice guideline or consensus development conference or consensus development conference, NIH).pt. | 42151 |
| 59 | (position statement* or policy statement* or practice parameter* or best practice*).ti,ab,kf,kw. | 31048 |
| 60 | (standards or guideline or guidelines).ti,kf,kw. | 105440 |

| | | |
|---|---|---|
| 61 | ((practice or treatment* or clinical) adj guideline*).ab. | 37832 |
| 62 | (CPG or CPGs).ti. | 5569 |
| 63 | consensus*.ti,kf,kw. | 24689 |
| 64 | consensus*.ab. /freq=2 | 23911 |
| 65 | ((critical or clinical or practice) adj2 (path or paths or pathway or pathways or protocol*)).ti,ab,kf,kw. | 19229 |
| 66 | recommendat*.ti,kf,kw. | 39030 |
| 67 | (care adj2 (standard or path or paths or pathway or pathways or map or maps or plan or plans)).ti,ab,kf,kw. | 55156 |
| 68 | (algorithm* adj2 (screening or examination or test or tested or testing or assessment* or diagnosis or diagnoses or diagnosed or diagnosing)).ti,ab,kf,kw. | 7192 |
| 69 | (algorithm* adj2 (pharmacotherap* or chemotherap* or chemotreatment* or therap* or treatment* or intervention*)).ti,ab,kf,kw. | 9314 |
| 70 | 46 or 47 or 48 or 49 or 50 or 51 or 52 or 53 or 54 or 55 or 56 or 57 or 58 or 59 or 60 or 61 or 62 or 63 or 64 or 65 or 66 or 67 or 68 or 69 | 1422311 |
| 71 | (systematic adj3 review$).ti,ab,kw. | 164344 |
| 72 | 24 or 45 or 70 or 71 | 5190597 |
| 73 | 10 and 19 and 72 | 3984 |

#### Appendix 2 - Dialectical Behavioural Therapy (DBT)


1. DBT treatments vs. non-active comparators
    a. Randomised Controlled Trials .......................................................... p. 7
    b. Non-randomised experiments and observational studies ........... p. 12
    c. Uncontrolled intervention development studies ......................... p. 18
    d. Implementation studies .................................................................. p. 19
2. DBT treatments vs. specialist comparators
    a. Randomised Controlled Trials .......................................................... p. 19
    b. Non-randomised experiments and observational studies ........... p. 22
3. Tests of partial/modified DBT treatments
    a. Randomised Controlled Trials .......................................................... p. 23
    b. Non-randomised experiments and observational studies ........... p. 26
    c. Uncontrolled intervention development studies ......................... p. 31
4. Tests of DBT treatments adapted to specific cohorts
    a. Randomised Controlled Trials .......................................................... p. 32
    b. Non-randomised experiments and observational studies ........... p. 33
    c. Uncontrolled intervention development studies ......................... p. 36


| Study design and comparator | Paper | Aim | Treatment details | Sample details | Outcomes | Main findings |
|---|---|---|---|---|---|---|
| **1. DBT vs Non-active comparators**<br>a. Randomised Controlled Trials | | | | | | |
| Randomised Controlled Trial. Non-specialist/inactive comparator. | Khabir et al. 2018 Iran | To investigate and compare clinical outcomes of DBT and MBT for people with BPD in an Iranian setting. | Treatment:<br>DBT – DBT based group therapy.<br>MBT – MBT based group therapy.<br><br>Duration/Intensity: Programme length unclear; twice weekly sessions (120 minutes).<br><br>Comparator: Medication only.<br><br>Service setting: Standalone outpatient intervention. | Sample Size: 51 (treatment completers N=36).<br><br>Demographics: 25/36 female; mean age 22.61 (only 18-27); no ethnicity data provided.<br><br>Diagnoses: BPD diagnosis. | Primary outcome: BPD symptoms (BPDSI-IV). Secondary outcomes: Anxiety symptoms (BAI), depression symptoms (BDI-II). | Primary outcome: Both treatments were more effective than the control treatment, involving medication only (p=.0001), in reducing BPD symptoms, but no difference was found between MBT and DBT (p=.4). Similar patterns were seen at follow-up two months after the end of treatment and for secondary outcomes. |

| | | | | | | |
|---|---|---|---|---|---|---|
| Randomised Controlled Trial. Non-specialist/inactive comparator. | McMain et al. 2017 Canada | To evaluate the clinical effectiveness of brief DBT skills training as an adjunctive intervention in people with BPD. | Treatment: Adapted brief group DBT - training uses a psycho-educational focus to enhance capabilities based on Linehan's skills manual.<br><br>Duration/Intensity: 20-week programme; weekly groups (120 minutes).<br><br>Comparator: Active waitlist - ancillary treatments were unrestricted for both groups.<br><br>Service setting: Standalone outpatient intervention. | Sample Size: 84.<br><br>Demographics: DBT group 83.3% female, waitlist group 73.8% female; DBT mean age 27.3 (SD=7.5), waitlist mean age 32.1, (SD=9.1); no ethnicity data provided.<br><br>Diagnoses: DSM-IV BPD diagnosis (SCID-II). | Primary outcome: Self-report frequency of suicidal or non-suicidal self-injurious (NSSI) episodes (LSASI, DSHI). Secondary outcomes: Service use (THI-2); BPD symptoms (BSL-23); anger (STAXI); symptom severity (SCL-90-R); impulsiveness (BIS-11); depressive symptoms (BDI-II); social functioning (SAS-SR); coping (DERS). | Primary outcome: The DBT group showed statistically greater reductions in the frequency of suicidal and self-harm episodes as measured by the clinician reported LSASI at 32 weeks (Chi squared = 6.71, p<.04), but the difference on the self-reported DSHI was in the same direction but did not reach statistical significance (Chi squared = 5.32, p=.08). Secondary outcomes: The DBT group had significantly fewer admissions up to 20 weeks, but this was not found at 32 weeks. Results on other secondary outcome measures were mixed, with DBT participants showing greater improvements than controls on measures of anger, distress tolerance and emotion regulation at 32 weeks, but not on other outcomes. |
| Randomised Controlled Trial. Non-specialist/inactive comparator. | Kramer et al. 2016 Switzerland | To investigate the effect of a 20-session group DBT skills module on symptoms and anger added to treatment as usual for people diagnosed with BPD. | Treatment: DBT-informed skills group training, with focus on emotion regulation (specifically problematic anger), plus TAU.<br><br>Duration/Intensity: 20-week programme; weekly.<br><br>Comparator: TAU involving generic mental health care of various types.<br><br>Service setting: Outpatient intervention added to various types of generic mental health care. | Sample Size: 41.<br><br>Demographics: 36/41 female; mean age 34.4; ethnicity data not provided.<br><br>Diagnoses: DSM-IV BPD diagnosis. | Primary outcome: Psychosocial functioning (OQ-45). Secondary outcome: Anger (CAMS) | Primary outcome: In the intention to treat analysis, repeated measures showed a reduction in symptoms across groups. MANCOVA showed a significant omnibus effect favouring overall symptom reduction in the DBT trained group at discharge (F(3, 34) = 2.92; p=.04). There was no significant difference at 3-month FU. Anger was also examined as a possible mediating variable in DBT treatment. |
| Randomised Controlled Trial. Non-specialist/inactive comparator. | Feigenbaum et al. 2012 UK | To evaluate the effectiveness of DBT delivered by staff with a level of training readily achievable in National Health Service care settings for individuals with a Cluster B personality disorder. | Treatment: Dialectical behavioural therapy (DBT)<br><br>Duration/Intensity: 12-month programme plus 3–6-week pre-treatment phase; weekly individual sessions (60 minutes) + weekly group skills training (150 minutes) + out of hours phone consultation.<br><br>Comparator: TAU<br><br>Service setting: Experimental group treated in specialist PD service; control group in generic community mental health services. | Sample Size: 42.<br><br>Demographics: 30/42 female; mean age 35.4 DBT group, 34.6 TAU group; no ethnicity data provided.<br><br>Diagnoses: Cluster B PD diagnosis (anti-social, borderline, histrionic, narcissistic). BPD (93%). | Primary outcome: Global distress (CORE-OM). Secondary outcomes: Self-harm and suicide attempts (SASII); length of psychiatric hospital admissions (THI); aggression, irritability, and suicidality (OAS); anger (STAXI); PTSD symptoms (modified PTSD symptom scale); dissociative symptoms (DES) | Primary outcome: A non-significant time × group interaction suggested that individuals from both groups had comparable declining slopes on CORE-OM, with no significant difference. There was some evidence of a greater decline in self-rated risk for the DBT group, but no significant evidence of differences on other secondary outcomes, including symptoms, suicidality or risk. A high drop-out rate from DBT was found (11 out of 26 still in treatment after a year). |

| | | | | | | |
|---|---|---|---|---|---|---|
| Randomised Controlled Trial. Non-specialist/inactive comparator. | Priebe et al. 2012 UK | To assess the effectiveness and cost-effectiveness of 12 months of DBT as compared to TAU in reducing self-harm in patients with a personality disorder. | Treatment: DBT - based on the principles of cognitive behavioural therapy with the inclusion of mindfulness, validation and supportive therapy techniques, and holds as its core the key dialectic of the acceptance of the individuals as they are with the acknowledgement of the need for change.<br><br>Duration/Intensity: 12-month programme; weekly individual therapy (60 minutes) + weekly skills training group (120 minutes) + out of hours telephone coaching as needed.<br><br>Comparator: TAU consisting of a variety of different forms of treatment other than DBT, delivered by any service.<br><br>Service setting: Standalone outpatient intervention for treatment group | Sample Size: 80.<br><br>Demographics: 87.5% female; mean age 32.2; ethnicities White 57.5%, Black 15%, Asian 21.3%, Mixed/Other 6.3%.<br><br>Diagnoses: 1) Five days or more with self-harm; and 2) PD diagnosis. | Primary outcome: Self-harm (self-reported). Secondary outcomes: BPD symptoms (ZAN-BPD); symptom severity (BPRS; BSI); quality of life (MANSA). | Primary outcome: Statistically significant treatment by time interaction for self-harm, incidence rate ratio 0.91; p = 0.001. For every 2 months spent in DBT, the risk of self-harm decreased by 9% relative to TAU. There was no evidence of differences on any secondary outcomes. The economic analyses revealed a total cost of a mean of 5,685 GBP in DBT compared to a mean of 3,754 GBP in TAU, but the difference was not significant. 48% of patients completed DBT. They had a greater reduction in self-harm compared to dropouts. |
| Randomised Controlled Trial. Non-specialist/inactive comparator. | Carter et al. 2010 Australia | To compare the outcomes of DBT (in an Australian context) with a waitlist control also receiving treatment as usual. | Treatment: DBT<br><br>Duration/Intensity: 6-month programme.<br><br>Comparator: The control condition was a 6-month WL for DBT while receiving TAU (TAUWL).<br><br>Service setting: Standalone outpatient intervention | Sample Size: 73.<br><br>Demographics: 100% female; mean age 24.5 (SD=6.10); ethnicity data not provided.<br><br>Diagnoses: DSM-IV BPD diagnosis (SCID-II). | Primary outcomes: Number and length of hospital admissions for DSH and psychiatric hospital. Secondary outcomes: Disability, i.e., days out of role and days spent in bed (unclear); quality of life (WHOQOL-BREF). | Primary outcomes: No statistically significant differences found between DBT and waitlist/TAU group in indicators related to deliberate self-harm or hospital admission. Secondary outcomes: Disability and quality of life were significantly better for the DBT group. |

| | | | | | | |
|---|---|---|---|---|---|---|
| Randomised Controlled Trial. Non-specialist/inactive comparator. | Soler et al. 2009 Spain | To compare the efficacy of DBT-skills training and standard group therapy (SGT) for outpatients with BPD. | Treatment: DBT skills group training - The DBT format used was adapted from the standard version (Linehan, 1993a, 1993b), applying one of the four modes of intervention: skills training. DBT-ST included all the original skills. These skills can be divided into those that promote change, interpersonal effectiveness and emotional regulation skills, and those that promote acceptance, mindfulness and distress tolerance skills.<br><br>Duration/Intensity: Programme length unclear; 13 sessions (120 minutes).<br><br>Comparator: Standard group therapy with same number of hours as intervention therapy over 3 months. The SGT format was oriented to provide a relational experience, allowing people with BPD to share their characteristic difficulties. Prominent techniques used were interpretation (although this was not used systematically), highlighting, exploration, clarification and confrontation.<br><br>Service setting: Standalone outpatient intervention | Sample Size: 60.<br><br>Demographics: Intervention group 79.3% female, control group 86.7% female; mean age intervention 28.45 (range 19-41), mean age control 29.97 (range 21-39); no ethnicity data provided.<br><br>Diagnoses: DSM-IV BPD diagnosis (SCID-II; DIB-R). | No primary outcome specified. BPD symptom severity (CGI-BPD); depressive symptoms (HRSD-17); anxiety symptoms (HRSA); symptom severity (BPRS; SCL-90-R); hostility/irritability (Buss–Durkee Inventory); impulsiveness (BIS); self-injury, suicide attempts, and visits to psychiatric emergency services. | No primary outcome specified. DBT-ST was associated with lower dropout rates (34.5% compared to 63.4% with SGT). It was superior to SGT in improving several mood and emotion areas, such as: depression, anxiety, irritability, anger and affect instability. Other measures of mood and emotion showed no significant difference, and no difference is reported on self-injury, suicide attempts or emergency service use. |
| Randomised Controlled Trial. Non-specialist/inactive comparator. | Van den Bosch et al. 2005 The Netherlands (FU to Verheul et al. 2013) | To examine whether the treatment effects of a previous 12 months randomised controlled trail comparing DBT to TAU were sustained over a 6-months follow-up. | Treatment: DBT.<br><br>Duration/Intensity: 12-month programme; weekly individual CBT + weekly skills training group (120-150 minutes).<br><br>Comparator: TAU (outpatient treatment from the original referral source).<br><br>Service setting: Standalone outpatient intervention | Sample Size: 58.<br><br>Demographics: 100% female; age range 18-65; no ethnicity data provided.<br><br>Diagnoses: DSM-IV BPD diagnosis (SCID-II). | No primary outcome specified. BPD symptom severity (BPDSI); parasuicidal/self-mutilating behaviours (LPC). | No primary outcome specified. Across the treatment period BPD symptoms (BPDSI), including impulsive behaviour ($F(1,248) = 11.93$, $p<.01$), self-mutilating behaviour ($F(1,51) = 11.85$, $p<.01$), and alcohol consumption ($F(1,54) = 5.33$, $p=.02$) decreased to a greater extent in the DBT group compared to the control group. These treatment effects were sustained in the first six months following treatment discontinuation ($p>.05$). Fewer patients in the DBT compared to the control group attempted suicide (LPC) during the treatment and follow-up period, however, this difference was not significant. |

| | | | | | | |
|---|---|---|---|---|---|---|
| Randomised Controlled Trial. Non-specialist/inactive comparator. | Verheul et al. 2003 Netherlands | To compare the effectiveness of DBT with treatment as usual for patients with BPD and to examine the impact of baseline severity on effectiveness. | Treatment: DBT - individual therapy and group therapy.<br><br>Duration/Intensity: 12-month programme; weekly individual therapy + weekly group sessions (150 minutes).<br><br>Comparator: TAU -clinical management from the original referral source (addiction treatment centres n=11, psychiatric services n=20). No more than two session per month.<br><br>Service setting: Standalone outpatient | Sample Size: 58.<br><br>Demographics: 100% females; mean age 34.9 (SD=7.7); ethnicity data not provided.<br><br>Diagnoses: DSM–IV version BPD diagnosis (SCID–II). | No primary outcome specified. Recurrent parasuicidal and self-damaging impulsive behaviours (BPDSI); self-mutilating behaviours (LPC). | No primary outcome specified. The frequency and course of suicidal behaviours were not significantly different across treatment conditions: neither treatment condition nor time x condition (t(1,166)=0.22; p=.639) reached statistical significance. Self-mutilating behaviours appeared to diminish gradually in the DBT group over the treatment year, resulting in a significant effect for the time x group interaction, but not for treatment condition alone. There was a large difference in drop out from treatment, with 77% of the DBT group but only 37% of the control group retained in treatment at the end of a year. |
| Randomised Controlled Trial. Non-specialist/inactive comparator. | Koons et al. 2001 USA | To compare DBT to treatment as usual in a veterans' mental health centre. | Treatment: DBT - individual and group DBT + therapist consultation meetings. Shortened from one year to six months.<br><br>Duration/Intensity: 6-month programme; weekly individual therapy + weekly group therapy (90 minutes).<br><br>Comparator: TAU – medication and psychotherapy.<br><br>Service setting: Community mental health centre providing general mental health care for veterans | Sample size: 28<br><br>Demographics: 100% female (inclusion criterion); mean age 35 (range 21-46); ethnicities 75% Caucasian and 25% African American.<br><br>Diagnoses: BPD diagnosis (DSM-III). | No primary outcome specified: Parasuicidal behaviour (PHI); suicidal ideation (SSI); hopelessness (BHS), depressive symptoms (BDI, 25-item version of the HAM-D); anxiety (HARS); anger (Spielberger Anger Expression Scale); dissociation (DES); healthcare utilisation including inpatient admissions (THI). | No primary outcome specified: Compared with patients in TAU, those in DBT reported significantly greater decreases in suicidal ideation, hopelessness, depression, and anger expression after 6 months. Differences on other outcome measures, including parasuicidal acts, hospitalisations, and anxiety, did not reach statistical significance. |
| Randomised Controlled Trial. Non-specialist/inactive comparator. | Linehan et al. 1994 USA | To evaluate the efficacy of dialectical behaviour therapy for improving interpersonal outcomes compared to TAU. | Treatment: DBT - individual behavioural psychotherapy and psychoeducational group sessions concurrently.<br><br>Duration/Intensity: 12-month programme; weekly individual therapy + weekly psychoeducational skills training group.<br><br>Comparator: TAU comparison group was a naturalistic condition. Subjects assigned to treatment as usual received alternative therapy referrals and were allowed to participate in any type of treatment available in the community.<br><br>Service setting: Standalone outpatient intervention | Sample Size: 26.<br><br>Demographics: 100% female; mean age 26.7 (SD=7.8); ethnicity data not provided.<br><br>Diagnoses: DSM-III-R BPD diagnosis (SCID-II). | No primary outcome specified. Service use (THI); social functioning (SAS-R); global functioning (GAS); anger (STAXI). | No primary outcome specified. One-way ANCOVA (with pre-treatment as a covariate) indicated that people in DBT improved significantly more than people receiving TAU on anger, global functioning and social functioning, but not social adjustment. |

| | | | | | | |
|---|---|---|---|---|---|---|
| Randomised Controlled Trial. Non-specialist/inactive comparator. | Linehan et al. 1991 USA | To evaluate the effectiveness of dialectical behaviour therapy for the treatment of recurrently parasuicidal women who meet the criteria for borderline personality disorder. | Treatment: DBT<br><br>Duration/Intensity: 12-month programme; weekly group therapy (150 minutes) + weekly individual therapy (60 minutes).<br><br>Comparator: TAU - subjects were given alternative community therapy referrals, usually by the original referral source, from which they could choose.<br><br>Service setting: Standalone outpatient intervention | Sample Size: 44.<br><br>Demographics: No demographics provided.<br><br>Diagnoses: 1) At least 7/10 on the DIB-R and met DSM-III criteria for BPD; and 2) at least two incidents of parasuicide in the last 5 years, with one during the last 8 weeks. | No primary outcome specified. Parasuicidal behaviour (PHI); mental health, medical treatment, and psychiatric inpatient care (THI); suicidal ideation (self-report form); depressive symptoms (BDI); hopelessness (BHS); reasons for living (Reasons for Living Inventory). | No primary outcome specified. The likelihood of any parasuicide was lower for treatment subjects (subjects assigned to DBT = 63.6%, control subjects = 95.5%, z=2.26, p<.005). There were significantly fewer parasuicidal acts per person and hospital days for participants in DBT than control group members. There were no between-group differences on measures of depression, hopelessness, suicide ideation, or reasons for living although scores on all four measures decreased throughout the year. |
| 1. DBT vs Non-active comparators<br>b. Non-randomised experiments and observational studies | | | | | | |
| Natural experiment with pre-post comparison. Non-specialist/inactive comparator. | Robinson et al. 2018 Canada | To evaluate the effectiveness of interdisciplinary administration of DBT to persons with a BPD diagnosis when delivered by an interdisciplinary team. | Treatment: DBT - Intensive DBT Programme were adaptive skills training groups, weekly individual psychotherapy, and phone coaching sessions. The distinctive aspect was delivery by a multidisciplinary team.<br><br>Duration/Intensity: 12-month programme; weekly group sessions (120 minutes).<br><br>Comparator: N/A<br><br>Service setting: Standalone outpatient intervention | Sample Size: 97.<br><br>Demographics: 81.1% female; no additional demographics provided.<br><br>Diagnoses: 1) BPD diagnosis; and 2) acute suicidal and self-harm behaviours. | No primary outcome specified. Symptom severity (BSI); BPD symptom severity (ZAN-BPD; BSL-23); coping skills (DBT-WCCL); quality of life (QOLI); service use (original client questionnaire). | No specified primary outcomes. Significant improvements over the treatment period found following interdisciplinary treatment on most outcomes, including borderline symptoms, quality of life and coping skills. |
| Natural experiment with pre-post comparison. Non-specialist/inactive comparator. | Flynn et al. 2017 Ireland | 1) To determine if completion of a 12-month DBT programme is associated with improved outcomes in terms of borderline symptoms, anxiety, hopelessness, suicidal ideation, depression and quality of life. 2) To assess client progress across multiple timepoints throughout the treatment. | Treatment: DBT<br><br>Duration/Intensity: 12-month programme; weekly individual sessions + weekly group skills training sessions + as needed phone coaching.<br><br>Comparator: N/A<br><br>Service setting: DBT teams established within generic community mental health service setting | Sample Size: 71.<br><br>Demographics: 61/71 female; mean age 40 (SD=9.76); no ethnicity data provided.<br><br>Diagnoses: DSM-IV-TR BPD diagnosis or emotionally unstable personality disorder (ICD-10). | No primary outcome specified. BPD symptoms (BSL-23); anxiety symptoms (BAI); hopelessness (BHS); suicidal ideation (BSS); depressive symptoms (BDI); quality of life (WHOQOL-BREF). | No primary outcome. At the end of the 12-month programme, significant reductions in borderline symptoms, anxiety, hopelessness, suicidal ideation and depression were observed and an increase noted in overall quality of life. Gains were especially made during the first 6 months of the programme with a tendency for scores to slightly regress after the six-month mark which marks the start of the second delivery of the group skills cycles. |

| | | | | | | |
|---|---|---|---|---|---|---|
| Quasi-experiment with pre-post comparison. Non-specialist/inactive comparator. | Rizvi et al. 2017 USA | To investigate outcomes of a 6-month course of comprehensive dialectical behaviour therapy (DBT) provided in a US training clinic with doctoral students as therapists and assessors. | Treatment: DBT - in accordance with the DBT treatment manuals (Linehan, 1993, 2014).<br><br>Duration/Intensity: 6-month programme; weekly individual therapy (60 minutes) + weekly group skills training (120 minutes) + as needed coaching.<br><br>Comparator: N/A<br><br>Service setting: Outpatient standalone intervention (DBT training clinic) | Sample Size: 50.<br><br>Demographics: 80% female; age 18+; no ethnicity data provided.<br><br>Diagnoses: BPD diagnosis. | No primary outcome specified. Suicidal behaviours (SASII; SITBI); BPD symptoms (BSL-23); emotional regulation (DERS); symptom severity (BSI); depressive symptoms (BDI-II); social and occupational functioning (WSAS); DBT skills (DBT-WCCL). | No primary outcome. During the 6 months of treatment, four participants made at least one suicide attempt (range: 1–2) and 14 participants reported at least one episode of non-suicidal self-injury (NSSI). Comparing rates of suicide attempts and NSSI from the 6 months before starting treatment to the 6 months during treatment showed that there was a significant decrease in rates for both SA (x2(1) = 4.17, p=.041), and NSSI (X2(1) = 4.37, p=.037). Across all participants, HLM analyses indicated that there was a significant decrease in all measures of psychopathology (BSL-23, DERS, BSI-GSI, and BDI) over the course of treatment and a significant increase in skills use (DBT-WCCL) and functioning (WSAS). |
| Natural experiment with contemporaneous comparison. Non-specialist/inactive comparator. | Gregory et al. 2016 USA | To examine the effectiveness of DDP and DBT in real-world settings. | Treatment: DBT and Dynamic deconstructive psychotherapy (DDP).<br><br>Duration/Intensity:<br>DBT: weekly individual sessions (60 minutes) + weekly group sessions (120 minutes).<br>DDP: 12-month programme; weekly individual sessions.<br><br>Comparator: TAU<br><br>Service setting: Standalone outpatient intervention | Sample Size: 68.<br><br>Demographics: DDP group 85% female, 84% DBT group female, 69% TAU female; mean age 28.0 (SD= 11.7) DDP, 36.6 (SD=10.2) DBT, 29.3 (SD=11.5) TAU; ethnicities Caucasian 89% DDP, 84% DBT, 94% TAU.<br><br>Diagnoses: BPD diagnosis. | Primary outcome: BPD symptoms (BEST). Secondary outcomes: Axis I diagnosis (PDSQ); depressive symptoms (BDI); social and occupational impairment (SDS); suicidal ideation and parasuicidal behaviour (SBQ). | Primary outcome: Attrition from DBT was high and DDP obtained better mean BPD symptom (BEST) score after 12 months of treatment than DBT (d=0.53, p=.042). Both active treatments performed better than TAU control and were associated with significant improvements over time on BEST. Secondary outcomes: Greater improvements were reported for DDP than for DBT for depression, disability, and self-harm, but not suicide attempts. |
| Natural experiment with pre-post comparison. Non-specialist/inactive comparator. | Stiglmayr et al. 2014 Germany | To investigate the effectiveness of DBT with BPD patients under routine mental health care conditions in Germany. | Treatment: DBT<br><br>Duration/Intensity: Programme length unclear; weekly individual therapy (50 minutes) + weekly skills group training (120 minutes) + telephone contacts as needed + consultation meeting (60 minutes).<br><br>Comparator: N/A<br><br>Service setting: Standalone outpatient intervention | Sample Size: 78.<br><br>Demographics: 91.5% females; mean age 30.1 (SD=8.1); no ethnicity data provided.<br><br>Diagnoses: DSM-IV-TR BPD (SCID-II). | No primary outcome specified. Suicide attempts and non-suicidal self-injury (LPC); number and length of inpatient or partial inpatient stays; BPD symptom severity (BSL); BPD diagnosis (SCID-II); borderline-specific thinking patterns (QTF); symptom severity (BSI); depressive symptoms (BDI; HRSD); dissociative symptoms (DSS). | No primary outcome specified. Uncontrolled study in which measurements are between time points with no specified outcome measure. Over the first 12 months of treatment, significant improvements found during treatment in NSSI, frequency and duration of inpatient treatment, severity of borderline symptoms, depression, dissociation and overall symptom severity. No significant difference in number of suicide attempts. |

| | | | | | | |
|---|---|---|---|---|---|---|
| Natural experiment with pre-post comparison. Non-specialist/inactive comparator. | Koons et al. 2013 US | To evaluate improvements in a cohort receiving DBT in a naturalistic setting, demonstrating how this data can be used to obtain satisfactory reimbursement for DBT. | Treatment: DBT<br><br>Duration/Intensity: a year of 50 minutes of weekly therapy, 2 hours weekly of skills training, 2 hours weekly of therapist consultation, and intersession coaching via telephone (8 participants have not completed a year though).<br><br>Comparator: N/A<br><br>Service setting: Standalone outpatient intervention | Sample Size: 49.<br><br>Demographics: 45/49 female; mean age 36; ethnicities White 31/49, Hispanic 16/49, Other 2/49.<br><br>Diagnoses: Personality dysfunction. BPD (69.4%); PTSD (34.7%); MDD or bipolar (16.7%). | No primary outcome specified. Depressive symptoms (BDI-II); hopelessness (BHS); global distress (CORE-OM). | No primary outcome specified. Uncontrolled design in which only change over time was measured. There were significant improvements in functioning and risk, depression, and hopelessness. However, the data suggest that the initial improvements at 3 or 4 months are much larger than at subsequent follow-ups. |
| Quasi-experiment with pre-post comparison. Non-specialist/inactive comparator. | Gutteling et al. 2012 The Netherlands | To evaluate outcomes of a 12-month adapted outpatient group dialectical behaviour therapy (DBT) programme for patients with a BPD. | Treatment: Outpatient group delivery of DBT, including standard DBT Skills Training Group (DBT-STG), DBT Group Therapy (DBT-GT), psychomotor group therapy, telephone consultation, and therapist consultation team meetings.<br><br>Duration/Intensity: 12-month programme; weekly sessions of group skills training (150 minutes), group therapy (120 minutes) and psychomotor group therapy (90 minutes) + fortnightly behaviour analysis (30 minutes) + on demand telephone consultations.<br><br>Comparator: N/A - pre-post<br><br>Service setting: Standalone outpatient intervention | Sample Size: 34.<br><br>Demographics: 100% female; mean age 32.65 (SD=7.59); no ethnicity data provided.<br><br>Diagnoses: DSM-IV BPD (SCID-II). | No specified primary outcome. Depressive symptoms (BDI-II); symptom severity (SCL-90-R), anger (STAXI); anxiety symptoms (STAI). | No specified primary outcome. At end of treatment, reductions over time reported in symptoms of depression, state and trait anxiety and parasuicidal behaviour. |

| | | | | | | |
|---|---|---|---|---|---|---|
| Quasi-experiment with pre-post comparison. Non-specialist/inactive comparator. | Axelrod et al. 2011 USA | To assess for improvement in emotion regulation and to examine the relationship between improvements in the emotion regulation and substance use problems following DBT treatment delivered to people with both substance use and borderline personality disorder diagnoses. | Treatment: DBT<br><br>Duration/Intensity: 20-week programme; weekly individual therapy (60 minutes) + weekly group skills (90 minutes) + as needed telephone skills coaching.<br><br>Comparator: N/A- pre-post<br><br>Service setting: Community substance abuse facility | Sample Size: 27.<br><br>Demographics: 100% female; mean age 38 (range 27–51); ethnicities 92% Caucasian and 8% Hispanic.<br><br>Diagnoses: 1) DSM-IV BPD diagnosis; and 2) substance dependence. | No primary outcome specified. Depressive symptoms (BDI); emotion regulation (DER); substance use in past 30 days (self-report, clinician report, tests). | No identified primary outcome. Significant reductions were observed in depression and in substance use - together with improved emotional regulation - over the treatment period. An interaction was observed between frequency of substance use and emotion regulation. |
| Quasi-experiment with pre-post comparison. Non-specialist/inactive comparator. | Blennerhauss ett et al. 2009 Ireland | To investigate whether a DBT programme run in a generic community mental health setting could achieve successful outcomes in respect of self-harming behaviour, inpatient psychiatric hospitalisation and general functioning as has been reported in studies in specialist settings. | Treatment: DBT<br><br>Duration/Intensity: 26-week programme; weekly group sessions.<br><br>Comparator: N/A - pre-post<br><br>Service setting: Generic community mental health centre | Sample Size: 8.<br><br>Demographics: 8/8 female; mean age 29.4 years (range 18-44 years); no ethnicity data provided.<br><br>Diagnoses: DSM-lll-R BPD diagnosis (SCID-II). | No primary outcome specified. Global distress (CORE); symptom severity (SCL-90-R); self-harm, suicidal ideation, substance abuse, and skills use (DBT diary cards). | No primary outcome specified. Significant improvements reported following treatment after completion of the DBT programme, significant improvement (p<.005) was seen on all subscales of the CORE including risk, symptoms, or problems, functioning and subjective wellbeing. Examination of diary cards showed that of the four patients who reported active self-harm episodes in the week prior to initial assessment, three reported reduced self-harm episodes in the week of the final assessment. |

| | | | | | | |
|---|---|---|---|---|---|---|
| Quasi-experiment with pre-post comparison. Non-specialist/inactive comparator. | Comtois et al. 2007 USA | To examine the effectiveness of a comprehensive DBT programme for individuals with BPD receiving outpatient care in a community mental health centre. | Treatment: DBT<br><br>Duration/Intensity: Programme length unclear; orienting and commitment sessions 2-3 meetings (30-60 minutes) + weekly individual therapy (60 minutes) + twice weekly group skills training (90 minutes) + phone consultation as needed (10-20 minutes) + DBT oriented case management as needed (30 minutes) + 1-3 months medication management (30 minutes).<br><br>Comparator: N/A - pre-post<br><br>Service setting: Generic community mental health centre | Sample Size: 38.<br><br>Demographics: 96% female; mean age 34 (range 19-54); ethnicities 96% Caucasian.<br><br>Diagnoses: 1) Severe impairment; and 2) extensive history of suicide attempt or crisis service use. Primary diagnosis of depression or dysthymia (87%); schizoaffective disorder (4%); bipolar disorder (4%); and schizotypal disorder (4%). Comorbidities: BPD (96%); history of at least one suicide attempt (91%). | No primary outcome specified. Service use (THI). | No primary outcome specified. Most outcomes improved significantly over the year of treatment, including medically treated self-inflicted injuries, inpatient admissions, and use of psychiatric services. |
| Quasi-experiment with pre-post comparison. Non-specialist/inactive comparator- | Harley et al. 2007 USA | To describe the modified DBT skills training programme, involving the group element in DBT only, and to explore its potential benefit to patients when delivered with separate individual therapy. | Treatment: DBT skills group therapy delivered alongside separate individual therapy, which may be non-DBT. Four skills modules delivered (i.e., mindfulness, interpersonal effectiveness, emotion regulation, and distress tolerance.<br><br>Duration/Intensity: 7-month programme; weekly sessions (105 minutes).<br><br>Comparator: N/A - pre-post<br><br>Service setting: Standalone outpatient intervention | Sample Size: 45.<br><br>Demographics: 92% female; mean age 40; ethnicities 96% Caucasian.<br><br>Diagnoses: BPD diagnosis (SCID-II). | No specified primary outcome. BPD, depressive, and anxiety symptoms (PAI); suicide (PAI Suicide scale); psychological wellbeing (SOS-10); tendency to portray events negatively (PAI NIM). | No primary outcome specified. Uncontrolled study reporting changes over time. At end of treatment, participants demonstrated significant improvement in NIM (p=.002), anxiety (p=.014), depression (p=.001), BPD symptoms (p<.001), suicide (p=.002), and SOS-10 (p<.001). Dropout was lower if individual therapists (who were separate from the DBT and not necessarily employing this model) were within the same treatment centre as the delivery of DBT. |
| Natural experiment with pre post comparison. Non-specialist/inactive comparator. | Zinkler et al. 2007 United Kingdom | To describe implementation and outcomes of DBT in a naturalistic setting with therapists as care coordinators. | Treatment: DBT delivered by therapists who also act as care coordinators.<br><br>Duration/Intensity: 12-month programme; weekly individual session (60 minutes) + weekly group session (120 minutes) + telephone coaching as needed.<br><br>Comparator: N/A<br><br>Service setting: Specialist PD service | Sample Size: 86.<br><br>Demographics: 88% female; mean age 33.21; ethnicities Black 16%, White 71%, Asian 8%, Other 4%.<br><br>Diagnoses: DSM-IV PD diagnosis (SCID II). Avoidant (42%); dependent (28%); obsessive-compulsive (30%); paranoid (51%); schizotypal (20%); schizoid (4%); histrionic (6%); narcissistic (6%); and borderline (91%) PD. | No primary outcome specified. Self-harm and suicide attempts (self-report and diary cards); psychiatric inpatient stays; quality of life (MANSA); BPD symptom severity (ZAN-BPD); service use and costs. | No primary outcome specified or testing for statistical significance. Large reductions were seen in incidents of self-harm (from 5.3 per service users per month in the year before entering the service to 1.2 during treatment), and in hospital days per service user per month (from 1.66 pre-treatment to 0.11 once in treatment). Of 49 service users who began treatment, 31 (63%) dropped out before completing 12 months. |

| | | | | | | |
|---|---|---|---|---|---|---|
| Quasi-experimental with pre-post comparison (pilot study). Non-specialist/inactive comparator. | Brassington and Krawitz. 2006 New Zealand | To describe the outcome of 10 patients treated in a New Zealand pilot study of dialectical behaviour therapy (DBT) for people with borderline personality disorder (BPD), and to ascertain the clinical utility and feasibility of implementing DBT into a standard New Zealand public mental health service. | Treatment: DBT<br><br>Duration/Intensity: programme length unclear; weekly individual therapy (60-90 minutes) + weekly group skills training (120 minutes) + weekly telephone calls and therapist consultation (90 minutes).<br><br>Comparator: N/A - pre-post<br><br>Service setting: Generic public mental health service | Sample Size: 10.<br><br>Demographics: 100% female; mean age 34.3 (range 21- 53); no ethnicity data provided.<br><br>Diagnoses: BPD diagnosis (IPDE). | No primary outcome specified. Personality functioning (MCMI-III); symptom severity (SCL-90-R). | Uncontrolled pilot study with no specified primary outcome. Statistically significant improvements reported on 10 of the 24 MCMI-III subscales, including the borderline personality, depression, and anxiety subscales, and on the Global Severity Index of the SCL-90-R and on 10 of the 12 SCL-90-R scales. |
| Quasi-experiment with pre-post comparison. Non-specialist/inactive comparator. | Ben-Porath et al. 2004 USA | To investigate the feasibility of delivering DBT to case management clients with both severe mental illness and a borderline personality disorder diagnosis in a public community mental health centre. | Treatment: DBT<br><br>Duration/Intensity: 6-month programme; weekly DBT skills training (90 minutes) + weekly individual therapy + on-demand telephone consultation + psychiatric services and case management.<br><br>Comparator: N/A - pre-post<br><br>Service setting: Community mental health centre providing case management for people with severe mental illness | Sample Size: 26.<br><br>Demographics: 25/26 female; mean age 35.48 (SD=10.19); ethnicity 26/26 Caucasian.<br><br>Diagnoses: 1) BPD diagnosis; 2) comorbid DSM-IV severe mental illness, including bipolar disorder, major depression, schizophrenia, or schizoaffective disorder; and 3) engaging in "system-interfering behaviours". | No primary outcomes specified. Suicidal ideation/suicidal thoughts and self-harm behaviours (diary card); employment status; hopelessness (BHS); symptom severity (SCL-90-R). | No primary outcome specified. Statistically significant reductions were reported in suicidal thoughts and unemployment. On average, clients also rated themselves as improved, however no statistically significant improvements were observed in parasuicidal behaviours or symptoms. |
| Quasi-experiment with pre-post comparison (pilot study). Non-specialist/inactive comparator. | Perseius et al. 2004 Sweden | A preliminary estimation of treatment costs before and after DBT. | Treatment: DBT<br><br>Duration/Intensity: 18-month programme; weekly individual + weekly group therapy + 24-hour telephone intervention as needed.<br><br>Comparator: N/A<br><br>Service setting: Range of types of mental health care | Sample Size: 22.<br><br>Demographics: 100% female; median age 29 years old (range 21–45); no ethnicity data provided.<br><br>Diagnoses: DSM-IV BPD diagnosis (SCID-II). One patient did not fulfil all DSM-IV criteria for BPD but was included due to repeated self-harm and suicide attempts. | Primary outcome: Costs. Secondary outcomes: Suicide attempts, deliberate self-harm, and psychiatric inpatient days (structured interviews, medical files). | Primary outcome: The total mean cost per patient and year, decreased from 320,627 SEK 12 months before therapy start to 210,858 SEK the last 12 months of the therapy, which is equivalent to a 35% cost reduction. The authors suggest that this uncontrolled study may indicate a reduction in costs when DBT is provided. This was primarily related to a significant reduction in use of inpatient beds. |

1. DBT vs Non-active comparators

c. Uncontrolled intervention development studies

| | | | | | | |
|---|---|---|---|---|---|---|
| Intervention development and uncontrolled preliminary testing. Non-specialist/inactive comparator. | Lopez and Chessick 2013 USA | To explore in a pilot study whether clients participating in a DBT graduate can achieve target goals relevant to establishing a life worth living. | Treatment: DBT graduate group focused on identifying important life goals such as work or education and making action plans to achieve them.<br><br>Duration/Intensity: 9-month programme; weekly groups (90 minutes).<br><br>Comparator: N/A pre-post<br><br>Service setting: Standalone outpatient intervention for graduates of DBT programme | Sample Size: 11.<br><br>Demographics: 54.6% female; mean age 41 (SD=9.5); ethnicities 90% Caucasian.<br><br>Diagnoses: Patients who completed DBT. Axis II diagnosis (100%) with BPD (90%). Primary diagnosis of MDD (72%). | No primary outcome specified. Depressive symptoms (PHQ-9); quality of life; employment status. | No primary outcome specified. Depression scores decreased significantly, and employment rate increased by six months of group attendance. All achieved a target goal of some type by the end of therapy. By month six, all of the members began to decrease their regular attendance as they began to achieve the target goals, such as going to work, that required their time; and the group was reported as successfully completed at Month 9. |
| Intervention development/uncontrolled preliminary testing. Non-specialist/inactive comparator. | Comtois et al. 2010 USA | To examine the feasibility of DBT–Accepting the Challenges of Exiting the System (DBT-ACES), a follow-up to standard DBT (SDBT) focused on social recovery and employment. | Treatment: DBT - Accepting the Challenges of Exiting the System (DBT-ACES), a follow-up to standard DBT (SDBT) focused on employment and social recovery.<br><br>Duration/Intensity: One year programme; weekly individual therapy + weekly skills group.<br><br>Comparator: N/A - pre-post<br><br>Service setting: Outpatient mental health centre | Sample Size: 30.<br><br>Demographics: 80% female; mean age 37 (range 19-56); ethnicities 100% Caucasian.<br><br>Diagnoses: 1) Severe impairment; and 2) extensive history of suicide attempt or crisis service use. BPD (97%) and other mental disorders. | Primary outcomes: Competitive employment, school attendance, and subjective satisfaction (QOLI - abbreviated). Secondary outcomes: self-injury, service use (THI). | Primary outcomes: Logistic regression modelling indicated a significant improvement in participants' odds of being employed or in school between the end of SDBT and the end of DBT-ACES (OR=3.34, 95% CI 1.14, 9.8; p<.05). One year after DBT-ACES, gains were reduced somewhat, and there was no significant difference from the end of SDBT to one year after the end of DBT-ACES. Eighteen of 28 clients for whom this datum was available (64%) had left the public mental health system one year after the end of DBT-ACES and were receiving private, low-income, or no mental health services. The RRM showed significant improvement in Quality of Life from the end of SDBT to the end of DBT-ACES (B=.49, p=.03), which was mostly retained one year after DBT-ACES but no longer reached statistical significance (B=.47, p=.08). Secondary outcomes: There were also improvements over time in measures of self-injury and service use. |
| Intervention development and uncontrolled preliminary testing. Non-specialist/inactive comparator. | Stanley et al. 2007 USA | To investigate the effectiveness of a brief, targeted DBT intervention for individuals with BPD. | Treatment: DBT (6 month) - (DBT-B). DBT-B was delivered in the standard manner except for the shortened duration from one-year minimum to six months.<br><br>Duration/Intensity: 6-month programme.<br><br>Comparator: N/A<br><br>Service setting: Standalone outpatient | Sample Size: 20.<br><br>Demographics: 85% female; mean age 32.2 (SD=8.7); ethnicities Caucasian 85%.<br><br>Diagnoses: DSM-IV BPD diagnosis (SCID-II). | No primary outcome specified. Self-harm and suicidal outcomes (diary cards); depressive symptoms (HAM-D); hopelessness (BHI); subjective distress (self-report). | Uncontrolled study with comparisons between time points and no specified primary outcome. Significant improvements over the course of 6 months treatment on most outcome measures, including NSSI episodes. |

1. DBT vs Non-active comparators
    d. Implementation/Observational study

| | | | | | | |
|---|---|---|---|---|---|---|
| Implementation/Observational study. Non-specialist/inactive comparator. | Flynn et al. 2020 Ireland | 1) To investigate barriers and facilitators to implementing DBT in a public mental health system. 2) To evaluate the effectiveness of the DBT programmes established across multiple independent sites as part of the national coordinated implementation. | Treatment: DBT Duration/Intensity: 12-month programme; weekly individual sessions + weekly group skills training sessions + as needed phone coaching.<br><br>Comparator: N/A - pre-post<br><br>Service setting: DBT teams established within generic community mental health service setting | Sample Size: 196.<br><br>Demographics: 80.6% female; age range 25–44; no ethnicity data provided.<br><br>Diagnoses: DSM-IV-TR BPD or EUPD (emotionally unstable PD) diagnosis (ICD-10). | Primary outcome: Study implementation (CFIR). Secondary outcomes: BPD symptoms (BSL-23); hopelessness (BHS); depressive symptoms (BDI-II); suicidal ideation (QSI); anger (STAXI-2), DBT skills and coping (DBT-WCCL); service utilisation and resource use (client record). | Primary outcome: Barriers and facilitators were identified to DBT implementation, based on the CFIR. Secondary outcomes: Regarding outcomes of treatment, there were statistically significant changes from T1 to T3 on all self-report outcome measures. Improvements were maintained at follow-up. Therapist-Rated Assessment recorded an increase in patients' functioning. There was a significant decrease in the proportion of participants self-harming from T1 to T3. There was a decrease in the total number of emergency department visits from T1 to T3, and a further decrease at T4. |
| **2. DBT vs Specialist comparators**<br>a. Randomised Controlled Trials | | | | | | |
| RCT. Specialist/active comparator. | McMain et al. 2012 (FU to McMain al. 2009) Canada | To evaluate clinical outcomes 2 years post-treatment in groups who were randomly assigned to DBT or General Psychiatric Management for borderline personality disorders. | Treatment: DBT.<br><br>Duration/Intensity: 12-month programme; weekly individual sessions (60 minutes) + weekly skills group (120 minutes) + weekly phone coaching (120 minutes).<br><br>Comparator: General Psychiatric Management (psychodynamic psychotherapy, case management and pharmacotherapy).<br><br>Service setting: Standalone outpatient intervention | Sample Size: 180.<br><br>Demographics: 86.1% female; mean age 30.4 (SD=9.9); no ethnicity data provided.<br><br>Diagnoses: 1) BPD; and 2) at least two suicidal or non-suicidal self-injurious episodes. | Primary outcome: Suicidal and non-suicidal self-injurious behaviour (SSHI). Secondary outcomes: BPD diagnosis and symptoms (ZAN-BPD); symptom severity (SCL-90-R); remission of BPD (IPDE); anger (STAXI), depressive symptoms (BDI-II); interpersonal problems (IIP-64); quality of life (EQ-5D); service use (THI). | 48% of patients originally randomised completed all four follow-up assessments. Primary outcome: No difference was found between groups for number of suicidal episodes over three years after the study baseline (z=–0.21, p=.83), with both groups maintaining the reduced rates of suicide attempts (z=0.47, p=.64), non-suicidal self-injurious behaviours (z=-1.82, p=.07), and medical severity of behaviours (t=0.78, df=933, p=.44) observed during treatment. Secondary outcomes: 57% in the DBT and 68% in the GPM group achieved diagnostic remission at three-year follow-up (IPDE). There were no significant group effects for most other service user outcomes, such as hospitalisation and use of emergency care, as well as clinical outcomes, such as interpersonal functioning and quality of life, while treatment effects were maintained. |

| | | | | | | |
|---|---|---|---|---|---|---|
| RCT. Specialist/active comparator. | Pasieczny et al. 2011 Australia | To contribute to effectiveness research on the use of DBT for BPD in routine clinical services in a culture and setting outside that of the existing efficacy research. | Treatment: DBT - DBT as described in Cognitive Behavioural Therapy of Borderline Personality Disorder (Linehan, 1993a) and Training Manual for Treating Borderline Personality Disorder (Linehan, 1993b).<br><br>Duration/Intensity: 6-month programme; weekly individual therapy (60 minutes) + weekly group skills training (120 minutes) + phone coaching as needed + weekly consultation meeting (90 minutes).<br><br>Comparator: Waitlist control (TAU). The control group received TAU, clinical case management (Kanter, 1989). This consisted of engagement, ongoing assessment, planning, linking with community resources, consultation with carers, assistance expanding social networks, collaboration with medical staff, advocacy, individual counselling, living skills training, psycho education and crisis management.<br><br>Service setting: Generic community mental health services | Sample Size: 90.<br><br>Demographics: 84/90 female; mean age 33.58 (SD=10.10); no ethnicity data provided.<br><br>Diagnoses: BPD diagnosis. | No primary outcome specified. Depressive symptoms (BDI-II); suicide ideation (BSS); anxiety symptoms (STAI-Y); symptom severity (BSI; SCL-90-R GSI); suicide attempts and self-harm episodes; ED visits, psychiatric admissions, and hospital attendances. | No primary outcome specified. After six months of treatment the DBT group showed significantly greater reductions in suicidal/non-suicidal self-injury (Hotelling's T = 25.13, F(2, 78)=25.14, p<.001), emergency department visits, psychiatric admissions, and bed days. (Hotelling's T = 25.13, F(3, 77)=7.70, p<.001). DBT patients demonstrated significantly improved depression, anxiety and general symptom severity scores compared to TAU at six months. |
| RCT. Specialist/active comparator. | McMain et al. 2009 Canada | To evaluate the clinical efficacy of DBT compared with General Psychiatric Management for people diagnosed with borderline personality disorder. | Treatment: DBT.<br><br>Duration/Intensity: 12-month programme; weekly individual sessions (60 minutes) + weekly skills group (120 minutes) + weekly phone coaching (120 minutes).<br><br>Comparator: General Psychiatric Management (psychodynamic psychotherapy, case management and pharmacotherapy).<br><br>Service setting: Standalone outpatient intervention | Sample Size: 180.<br><br>Demographics: 86.1% female; mean age 30.4 (SD=9.9); no ethnicity data provided.<br><br>Diagnoses: 1) BPD; and 2) at least two suicidal or non-suicidal self-injurious episodes. | Primary outcomes: Frequency and severity of suicidal and non-suicidal self-injurious episodes (SASII). Secondary outcomes: BPD symptoms (ZAN-BPD); symptom severity (SCL-90-R); remission of BPD (IPDE); anger (STAXI); depressive symptoms (BDI-II); interpersonal functioning (IIP-64); quality of life (EQ-5D); service use (THI); treatment retention. | Primary outcome: There was no significant difference in frequency of suicidal episodes, non-suicidal self-injurious episodes, and medical risk of these behaviours between groups (t=–0.47, df=450, p=.64), with significant decreases over time in both groups. Secondary outcomes also showed no evidence of significant difference between groups, with both improving over time. |

| | | | | | | |
|---|---|---|---|---|---|---|
| RCT. Specialist/active comparator. | Clarkin et al. 2007 USA | To compare three-year long outpatient treatments for borderline personality disorder: dialectical behaviour therapy, transference-focused psychotherapy, and a dynamic supportive treatment. | Treatment: Transference focused therapy / DBT / Supportive treatment.<br><br>Duration/Intensity:<br>Transference focused therapy: 12 months programme; two individual weekly sessions.<br>DBT: 12 months programme; a weekly individual and group session and available telephone consultation.<br>Supportive treatment: 12 months programme; one weekly session with additional sessions as needed.<br><br>Comparator: Active comparator.<br><br>Service setting: Standalone outpatient interventions | Sample Size: 90.<br><br>Demographics: 92.2% female; mean age 30.9 (SD=7.85); ethnicities 67.8% Caucasian, 10% African American, 8.9% Hispanic, 5.6% Asian, 7.8% Other.<br><br>Diagnoses: DSM-IV BPD diagnosis (SCID-II). | Primary outcomes: Suicidality (MOAS); aggression (AIAQ); impulsivity (BIS-11). Secondary outcomes: depressive symptoms (BDI); global functioning (GAF); social functioning (SAS). | Regarding primary outcomes, both transference-focused psychotherapy and dialectical behaviour therapy were significantly associated with improvement in suicidality. Only transference-focused psychotherapy and supportive treatment were associated with improvement in anger. Transference- focused psychotherapy and supportive treatment were each associated with improvement in facets of impulsivity. Regarding secondary outcomes, all treatments were associated with improvements in depression, anxiety, global functioning, and social functioning. Only transference-focused psychotherapy was significantly predictive of change in irritability and verbal and direct assault. The authors suggest transference-focused psychotherapy may result in impacts on a wider range of outcomes than other treatment conditions. |
| RCT. Specialist/active comparator. | Linehan et al. 2006 USA | To compare outcomes from DBT with those from expert therapists using other models on suicidal behaviour and other outcomes in women with BPD. | Treatment: DBT - individual behavioural psychotherapy and psychoeducational group sessions concurrently.<br><br>Duration/Intensity: Programme length unclear; weekly individual psychotherapy (60 minutes) + weekly group skills training (150 minutes) + telephone consultation (as needed).<br><br>Comparator: Community Treatment by Experts (CTBE) - psychotherapy by experienced therapists using a variety of treatment models.<br><br>Service setting: Standalone outpatient intervention | Sample Size: 101.<br><br>Demographics: DBT group 68.7% female, CTBE group 64% female; mean age: DBT 29.0 (SD=7.3), CTBE mean age 29.6 (SD=7.8); ethnicities DBT Caucasian 86.5%, African American 3.8%, Asian American 1.9%, Native American 1.9%, other 5.8%, CTBE: Caucasian 87.8%, African American 4.1%, Asian American 2.0%, Native American 0%, Other 6.1%.<br><br>Diagnoses: 1) BPD diagnosis; and 2) two suicide attempts or self-injuries in the past 5 years, with at least one in the past weeks. | Primary outcome: Topography, suicide intent, medical severity of suicide attempts (SASII). Secondary outcomes: Suicide ideation (SBQ); reasons for living (Reasons for Living Inventory); experience of treatment and service use (THI); depressive symptoms (HRSD). | Primary outcome: There were no documented suicides in either condition during the 2-year study. The DBT group had half the rate of suicide attempts compared with the CTBE group (23.1% vs 46%, Chi squared =5.98, p=.01; hazard ratio, 2.66, p=.005). Outcomes were also significantly better for emergency department visits and hospital admissions in the DBT group. No significant differences were found in depression, quality of life or suicidal ideation between groups. Dropout was 3 times higher in CTBE compared to DBT. |

| | | | | | | |
|---|---|---|---|---|---|---|
| RCT. Specialist/active comparator. | Linehan et al. 2002 USA | To examine whether DBT is more effective than a Comprehensive Validation Therapy with 12 Step (CVT/12S) in treatment of women with both opioid dependence and borderline personality disorder. | Treatment: DBT - individual behavioural psychotherapy and psychoeducational group sessions concurrently, plus addiction treatment.<br><br>Duration/Intensity: 12-month programme; individual therapy weekly (40-90 minutes) + weekly group skills training (120 minutes) + weekly individual skills coaching (30 minutes) + other support group meetings as needed.<br><br>Comparator: Comprehensive validation therapy plus 12 step programme. CVT/12S focused on validating the client and their experience. Major contrast to DBT is therapists are non-directive and agenda determined by the client.<br><br>Service setting: Addiction treatment setting | Sample Size: 24.<br><br>Demographics: 100% female; mean age 36.19 (SD=7.3); ethnicities 66% Caucasian, 26% African American, 4% Hispanic.<br><br>Diagnoses: 1) DSM-IV BPD diagnosis (SCID-I; PDE); and 2) opiate dependence diagnosis (SCID-I). | No primary outcome specified. Drug use (TLFB); parasuicidal behaviours (PHI); social functioning (SHI; GAS); general functioning (GAF); symptom severity (BSI). | No primary outcome specified. Opiate use fell to relatively low levels in both groups by 16 months post-randomisation and both groups also showed significant reductions in psychopathology relative to baseline, but no significant differences on outcome measures were found between groups. |
| 2. DBT vs Specialist comparators<br>b. Non-randomised experiments and observational studies | | | | | | |

| | | | | | | |
|---|---|---|---|---|---|---|
| Natural experiment with contemporaneous control. Specialist/active comparator. | Barnicot et al. 2019 UK | To investigate whether clinical outcomes at 12 months in naturalistic personality disorder treatment settings differ between people receiving DBT and those receiving MBT. | Treatment: DBT and MBT.<br><br>Duration/Intensity:<br>DBT: 12-month programme; 4 weekly sessions.<br>MBT: 18-month programme; 2 weekly/fortnightly sessions + initial short-term psychoeducation.<br><br>Comparator: Mentalisation-based therapy - 18-month period, weekly or fortnightly individual therapy and weekly group therapy. They also provided a short-term group programme which involves weekly groups delivered over a 10-week period.<br><br>Service setting: Specialist PD services | Sample Size: 90.<br><br>Demographics: 72% female; mean age 31.0 (SD=13.0); ethnicities: White 64%, black and minority 36%.<br><br>Diagnoses: DSM-IV BPD diagnosis (SCID-II). | No primary outcome specified. Crisis service use: A&E and psychiatric hospital admissions; self-harm (SASII); BPD symptom severity (BEST); emotion regulation (DERS); dissociation (Dissociative Experience Scale); interpersonal problems (SIDES-SR). | No primary outcome specified. Patients receiving DBT were significantly less likely to complete at least 12 months of treatment than those receiving MBT (completion rate 42% v. 72%), but this was no longer significant after adjusting for baseline differences. At 12 months follow up, groups did not differ in adjusted or unadjusted comparisons of number of incidents of self-harm, BPD severity, emotional dysregulation, relationships with others or dissociation. In unadjusted models, participants receiving DBT reported a significantly steeper decline over time in incidents of self-harm and in emotional dysregulation than participants receiving MBT, remaining significant after adjusting for confounders. |
| Quasi-experiment with contemporaneous control. Specialist/active comparator. | Andión et al. 2012 Spain | To compare the effectiveness of individual vs. combined individual and group DBT. | Treatment: DBT with combined individual and group elements - a weekly skills training programme added to individual therapy, telephone consultation and consultation team meetings.<br><br>Duration/intensity: 12-month DBT programme; 48 hours of individual sessions + 96 hours of group skills training.<br><br>Service setting: Outpatient DBT programme | Sample size: 51.<br><br>Demographics: 96.2% female; mean age 25.63 (SD=6.46); no ethnicity data provided.<br><br>Diagnoses: DSM-IV BPD outpatients. | No primary outcome specified. Suicide attempts, self-harm, visits to emergency departments (measure unclear). | No primary outcome specified. No significant differences reported in outcomes between groups. Improvements on most outcome measures, including suicide and self-harm and emergency department visits, reported when comparing follow-up with pre-treatment, and sustained at 18 months. |

#### 3. Tests of partial/modified DBT treatments

##### a. Randomised Controlled Trials

| | | | | | | |
|---|---|---|---|---|---|---|
| RCT. Partial/modified. | Carmona I Farres et al. 2019 Spain | To examine the impact of mindfulness training on default mode network brain activation and deactivation during an executive task in a sample of individuals with BPD, and on BPD symptoms and other clinical variables. | Treatment: DBT-mindfulness module.<br><br>Duration/Intensity: 10-week programme; daily sessions (10-15 minutes).<br><br>Comparator: DBT-Interpersonal effectiveness. The aim of the DBT-IE is to teach participants how to effectively interact with others in interpersonal situations.<br><br>Service setting: Standalone outpatient setting | Sample Size: 65.<br><br>Demographics: DBT-M group 93.9% females, DBT-IE group 84.4%; DBT-M mean age 31.03 (SD=6.76), DBT-IE 33.75 (SD=8.78); no ethnicity data provided.<br><br>Diagnoses: DSM-IV BPD diagnosis (SCID-II). | No primary outcome specified. BPD symptom severity (BSL-23); depressive symptoms (BDI); anxiety symptoms (STAI); mindfulness (FFMQ). | No primary outcome specified. Both groups showed significantly decreased general borderline depressive and anxiety symptomatology. Activation of the left anterior insula of the brain on a relevant task increased in both groups following the intervention. |
| RCT. Partial/modified. | Lin et al. 2019 Taiwan | To evaluate the effectiveness of the condensed DBTSTG in comparison to a Cognitive Therapy Group (CTG) in reducing depression and suicide attempts in a sample of Taiwan college students with BPD. | Treatment: The Dialectical Behaviour Therapy Skills Training Group (DBTSTG) programme is a manualised group intervention, adapted from the Skills Training Manual for Treating Borderline Personality Disorder. Session topics included: mindfulness skills, distress tolerance skills reducing vulnerability to negative emotions, amongst many others.<br><br>Duration/Intensity: 8-week programme; weekly sessions (120 minutes).<br><br>Comparator: Active comparator - DBT skills training group - manualized group intervention. Series of topics including relationship between thoughts and feelings, identifying automatic thoughts, challenging thoughts and other topics.<br><br>Service setting: Student population at university mental health centre | Sample Size: 82.<br><br>Demographics: DBTSTG group 90.5% female, CTG group 85% female; mean age DBTSTG 20.38 (SD=.68), CTG mean age 20.46 (SD=.76); no ethnicity data provided.<br><br>Diagnoses: 1) BPD diagnosis; and 2) at least one suicide attempt in the past 6 months. | No primary outcome specified. Suicide attempt (interview); BPD symptoms (BPDFS; SCID-II); depressive symptoms (KDI); suicidal ideations (ASIQ-R); cognitive errors (CEQ-S); emotion regulation (DERS). | No primary outcome specified. No suicide reattempts were recorded in either group over the 6-month follow-up period. Reductions in depression were also not significantly different between groups, but at 6 months there were significant group x time effects favouring the DBSTG group for borderline features, suicidal ideation, and emotional regulation, whereas the CT group were significantly more able to detect cognitive errors. |

| | | | | | | |
|---|---|---|---|---|---|---|
| RCT. Partial/modified. | Elices et al. 2016 Spain | To evaluate the effects of a stand-alone mindfulness intervention on borderline symptoms and mindfulness-related capacities in patients with BPD. | Treatment: Mindfulness training, aiming to preserve the essence of mindfulness skills taught in DBT.<br><br>Duration/Intensity: 10-week programme; weekly group sessions (150 minutes).<br><br>Comparator: Active comparator - interpersonal effectiveness skills training.<br><br>Service setting: Standalone outpatient interventions | Sample Size: 64.<br><br>Demographics: 86 % female; mean age 30; ethnicities 100% Caucasian.<br><br>Diagnoses: BPD diagnosis (SCID-II, DIB-R). Comorbid disorders: Axis I (100%); Axis II cluster C diagnosis (31%), cluster A (30%), cluster B (26%). | No primary outcome specified. PD status (SCID-II; DIB-R); axis I comorbidities (PDSQ); BPD symptom severity (BSL-23); mindfulness (FFMQ). | No primary outcome specified: The mindfulness group showed greater improvement over time than the IE group in borderline symptoms and in some mindfulness skills. |
| 3-arm RCT. Partial/modified. | Linehan et al. 2015 USA | To evaluate the importance of the skills training component of DBT by comparing skills training plus case management (DBT-S), DBT individual therapy plus activities group (DBT-I), and standard DBT which includes skills training and individual therapy. | Treatment: DBT<br><br>Duration/Intensity:<br>DBT: Programme length unclear; weekly individual therapy (60 minutes) + weekly group skills training (150 minutes).<br>DBT skills training (DBT-S): Programme length unclear; individual sessions monthly plus additional as needed up to weekly sessions + weekly group skills training (150 minutes).<br>DBT individual therapy (DBT-I): weekly individual therapy (60 minutes) + weekly activity-based support group (150 minutes).<br><br>Comparator:<br>DBT-S - designed to evaluate the effect of DBT skills training by providing DBT group skills training while removing the DBT individual therapy component.<br>DBT-I - designed to eliminate all DBT skills training from the treatment by re-moving group skills training and prohibiting individual therapists from teaching DBT skill.<br><br>Service setting: Standalone outpatient intervention | Sample Size: 99.<br><br>Demographics: 100% female; mean age 30.3 (SD=8.9); ethnicities 71% White.<br><br>Diagnoses: 1) DSM-IV BPD diagnosis (IPDE; SCID-II); and 2) at least 2 suicide attempts and/or NSSI episodes in the past 5 years, at least 1 suicide attempt or NSSI act in the 8-week period before entering the study, and at least 1 suicide attempt in the past year. | No primary outcome specified. Frequency, intent, and severity of suicide attempts and NSSI acts (SASII); suicidal ideation (SBQ); reasons for living (Reasons for Living Inventory); use of crisis services and psychotropic medications (THI); depressive symptoms (HRSD); anxiety symptoms (HRSA). | No primary outcome specified. All treatment conditions resulted in similar improvements in the frequency and severity of suicide attempts, suicide ideation, use of crisis services, and reasons for living. Compared with the DBT-I group, interventions that included skills training resulted in greater improvements in the frequency of NSSI acts and in depression and anxiety. Compared with the DBT-I group, the standard DBT group had significantly lower dropout rates from treatment, and patients were less likely to use crisis services in follow-up. |

| | | | | | | |
|---|---|---|---|---|---|---|
| RCT. Partial/modified. | Turner et al. 2000 USA | To assess the effectiveness of a DBT-oriented therapy model compared to an alternative psychosocial treatment, i.e., client-centred therapy (CCT) treatment protocol. | Treatment: DBT - oriented treatment (added psychodynamic techniques, but without separate DBT skills training groups).<br><br>Duration/Intensity: Programme length unclear; 49-84 individual sessions + 6 group sessions.<br><br>Comparator: Active comparator (client-centred therapy control condition; CCT) which focuses on the empathic understanding of the patient's sense of aloneness and providing a supportive atmosphere for individuation.<br><br>Service setting: Standalone outpatient intervention | Sample Size: 24.<br><br>Demographics: 79.2% female; mean age 22 (range 18-27); ethnicities 79.2% Caucasian, 16.7% African American, 4.2% Asian American.<br><br>Diagnoses: DSM-III BPD diagnosis. Comorbid axis I disorder (95.3%). | No primary outcome specified. Depressive symptoms (HRSD; BDI); psychiatric symptoms (BPRS); anxiety symptoms (BAI); emotion regulation (Target Behaviour Ratings); suicidal ideation (BSI); suicide urges and attempts (daily patient logs); psychiatric hospitalisation (assessor rated). | No specified primary outcome. The DBT group showed greater improvement than the CCT group on most measures: self-harm behaviour and suicidality; impulsiveness, anger and depression, and overall symptom severity. Anxiety outcomes were not significantly different. |
| RCT (pilot). Partial/modified. | Feliu-Soler et al. 2017 Spain | To investigate the effects of a short training programme in loving-kindness and compassion meditation (LKM/CM) in patients with borderline personality disorder. | Treatment: Loving-Kindness and Compassion Meditation (LKM/CM).<br><br>Duration/Intensity: 13-week programme; 10-week mindfulness training followed by 3 weeks of weekly sessions.<br><br>Comparator: Active comparator: mindfulness continuation training.<br><br>Service setting: Standalone outpatient intervention | Sample Size: 32.<br><br>Demographics: 30/32 female; age rage 18-45; ethnicities 100% Caucasian.<br><br>Diagnoses: DSM-IV-TR BPD (DIB-R). | No primary outcome specified. BPD symptoms (DIB-R; BSL-23); self-compassion (SCS); critical and reassuring self-evaluative responses (FRCRS); mindfulness (PHLMS). | No primary outcome specified. Similar improvements were found on most outcome measures in both groups, with no clear statistically significant differences. |
| **3. Tests of partial/modified DBT treatments**<br>b. Non-randomised experiments and observational studies | | | | | | |
| Quasi-experiment with pre-post comparison. Partial/modified. | Kells et al. 2020 Ireland | To investigate the effectiveness of a 24-week DBT-ST intervention for people attending community services for BPD or emotion dysregulation who are not currently actively self-harming. | Treatment: DBT skills training (DBT-ST).<br><br>Duration/Intensity: 24-week programme; weekly sessions (150 minutes).<br><br>Comparator: N/A<br><br>Service setting: Generic community mental health services | Sample Size: 100.<br><br>Demographics: 71% female; 32% aged 25-34; ethnicity data not provided.<br><br>Diagnoses: 1) DSM-IV-TR BPD diagnosis, borderline personality traits, or emotion dysregulation; and 2) history of difficulties regulating emotions. Patients who actively self-harmed were excluded. BPD diagnosis (41%) and BPD traits (59%). | No primary outcome specified. Emotional regulation (DERS); mindfulness (FFMQ); coping skills (DBT-WCCL). | Uncontrolled design in which only change over time was measured. No primary outcome specified. There were significant improvements for dysfunctional coping, emotional regulation and DBT skill use and mindfulness, including all sub-scales, at the end of treatment. However, the drop-out rate was high (49% at post-intervention). |

| | | | | | | |
|---|---|---|---|---|---|---|
| Natural experiment with pre-post comparison. Partial/modified. | Lakeman et al. 2020 Australia | To describe implementation and evaluate outcomes from a high-fidelity Dialectical Behaviour Therapy (DBT) programme for young people with BPD or emerging BPD (15–25 years). | Treatment: DBT skills group followed the DBT Skills Manual (Linehan, 2015b). This not an accredited DBT programme and was adapted to focus on youth. 1) "pre-commitment" phase of therapy. This period focused on building an alliance with the client. 2) The skills-group consisted of four modules: Emotional Regulation, Distress Tolerance, Interpersonal Effectiveness and "Walking the Middle Path". All therapists were available to provide telephone coaching between face-to-face sessions. The skills-group size was limited to eight participants and was facilitated by two therapists. During the twenty-week cycle of skills-group, clients continued to meet with their therapist weekly and access telephone coaching as needed (this was rarely used more than once a week by any participant). At the end of the group programme clients were invited to continue for a further cycle if they wished.<br><br>Duration/Intensity: 20-week programme following a "pre-commitment" individual therapy phase; main programme: weekly group therapy (180 minutes).<br><br>Comparator: N/A - pre post study<br><br>Service setting: Standalone outpatient intervention (delivered through a partnership between public mental health and third sector organisations) | Sample Size: 22.<br><br>Demographics: 81% female; mean age 20 (SD=2.5); ethnicity data not provided.<br><br>Diagnoses: BPD diagnosis (by medical practitioner). | No primary outcome specified. BPD symptoms (BSL-23; BSL-Supp); overall wellbeing (visual rating); number of ED presentations and psychiatric hospital days (hospital records). | Uncontrolled study where reported outcomes relate to change over time. No primary outcome specified. Participants who remained in the programme for at least twelve weeks had significant reductions in borderline symptoms (BSL-23 scores), with several reporting no symptoms after completing the programme. Rates of hospital and emergency department use in the year also fell significantly. A further reported finding is that it is feasible to deliver a high fidelity DBT programme to youth in a public mental health context in Australia. |
| Quasi-experiment with contemporaneous comparison. Partial/modified. | Lyng et al. 2020 Republic of Ireland and Northern Ireland | To investigate outcomes for adults with BPD on waiting lists for full DBT of offering them standalone DBT group skills training. | Treatment: Standalone DBT skills training group - Skills training group from DBT without any of the additional therapy provided by standard DBT, offered to people on the waiting list for standard DBT.<br><br>Duration/Intensity: 24-week programme; weekly skills training group (150 minutes) + weekly therapist consultation.<br><br>Comparator: Standard DBT<br><br>Service setting: Standalone outpatient interventions recruiting from generic community mental health services | Sample Size: 88.<br><br>Demographics: treatment group 82% female, comparator group 83% female; mean age 33.5 (SD=10.46), comparator mean age 33.2 (SD=8.31); no ethnicity data provided.<br><br>Diagnoses: BPD diagnosis. | No primary outcome specified. BPD symptoms (BSL23); symptom severity (SCL-90-R GSI); suicidal ideation (SSI; BSS); hopelessness (BHS); emotion regulation (DERS). | No specified primary outcome. Dropout rates were higher for the standalone DBT skills training condition (38% vs. 17%). No statisticalclinically significant differences were found among completers between conditions for borderline symptoms, general psychopathology and suicide ideation after six months treatment. Higher risk individuals were excluded from the standalone skills condition. |

| | | | | | | |
|---|---|---|---|---|---|---|
| Natural experiment with pre-post comparison. Partial/modified. | Aafjes-van Doorn et al. 2020 US | To investigate the process of adding a DBT skills group to psychoanalytic psychotherapy, and to evaluate changes in psychiatric symptoms, quality of life, and mindfulness. | Treatment: DBT skills training group added to psychoanalytic psychotherapy.<br><br>Duration/Intensity: 20 sessions of weekly 2-hour DBT skills training<br><br>Comparator: N/A<br><br>Service setting: Standalone outpatient intervention | Sample Size: 8.<br><br>Demographics: 87.5% female; mean age 33; no ethnicity data provided.<br><br>Diagnoses: Experiencing emotional dysregulation. | No primary outcome specified. BPD symptoms (BSL-23; BSL-Supplement); anxiety symptoms (BAI); depressive symptoms (BDI-II); interpersonal problems (IIP-32); quality of life (Q-LES-Q); mindfulness (MAAS); global functioning (GAF). | Uncontrolled design in which only change over time was measured. No primary outcome specified. In a very small sample of 8, depressive and anxiety symptoms and quality of life improved significantly over the course of treatment, but not interpersonal functioning, PD symptoms or mindfulness. |
| Quasi-experiment with pre-post comparison (pilot study). Partial/modified. | Sandage et al. 2015 USA | To conduct a pilot study of inclusion of a novel manualized group forgiveness module within dialectical behaviour therapy (DBT). | Treatment: A novel DBT single module - forgiveness - forgiveness skills psychoeducational module adapted from the REACH forgiveness intervention (Worthington, 2006) and integrated with DBT language and skills in outpatient DBT therapy.<br><br>Duration/Intensity: Programme length unclear; 4 group sessions (120 minutes).<br><br>Comparator: N/A<br><br>Service setting: Standalone outpatient intervention | Sample Size: 40.<br><br>Demographics: 88.1% female; mean age 40.02 (SD=12.86); ethnicities 90.5% European American, 2.4% African American, 2.4% Arabic American, 2.4% multiracial, 2.4% did not identify.<br><br>Diagnoses: BPD diagnosis (MSI-BPD). | No primary outcome specified. Motivations toward a specific offender (TRIM); (emotional) forgiveness for a specific offence (DFS; EFS); proneness to forgive interpersonal transgressions (TFS); attachment (ECR-S); mental health symptoms (PSC). | Uncontrolled study in which comparisons are between timepoints. No primary outcome specified. Participants showed increases in all measures of forgiveness and decreases in attachment security and psychiatric symptoms during the forgiveness module and maintained to the 6-week follow-up. These were all statistically significant, except for anxious attachment. |
| Natural experiment with pre-post comparison. Partial/modified. | Williams et al. 2010 Australia | To investigate whether a DBT skills-training group programme is beneficial in decreasing BPD-related symptoms and functioning, and in decreasing service utilisation. | Treatment: DBT skills group (alongside individual DBT or other continuing individual therapy). DBT skills group focused on Emotional Regulation, Interpersonal Effectiveness, Core Mindfulness, and Distress Tolerance).<br><br>Duration/Intensity: 20-week programme; weekly sessions (120 minutes).<br><br>Comparator: N/A<br><br>Service setting: Standalone outpatient therapy | Sample Size: 140.<br><br>Demographics: group 55/68 female; mean age 35.59 (SD=10.02); no ethnicity data provided.<br><br>Diagnoses: DMS-IV-TR BPD diagnosis. | No primary outcome specified. Psychological distress and impairment (K10+); symptoms and functioning (BASIS-32); depressive symptoms (BDI-II); BPD symptoms (BSI); DSM-IV criteria for BPD (MSI-BPD); service use (Community Based Information System CBIS). | Uncontrolled study in which comparisons are between timepoints with no specified primary outcome. Individual DBT was related to a higher completion rate than individual. Improvements were seen over during psychotherapy on all outcome scales and on some service use measures. |

| | | | | | | |
|---|---|---|---|---|---|---|
| Natural experiment with pre post comparison. Partial/modified. | Yen et al. 2009 USA | To assess whether women with a BPD diagnosis improved over 3 months following a 5-days partial hospitalisation DBT programme. | Treatment: Study evaluates progress over 3 months after discharge from a DBT-based 5-day day hospital programme.<br><br>Duration/Intensity: 5-day DBT-based programme, 9am-3:30pm.<br><br>Comparator: N/A<br><br>Service setting: Specialist PD Day service (followed by discharge to various forms of TAU) | Sample Size: 47.<br><br>Demographics: 100% female.<br><br>Diagnoses: DSM-IV BPD diagnosis (SCID-II) | Six primary outcomes specified: Depressive symptoms (BDI); symptom severity (BSI); anger (STAXI); hopelessness (BHS); self-injury (Self-Injury Questionnaire, adapted from PHI). Secondary outcomes: Dissociation (DES). | Uncontrolled design in which only change over time was measured. Primary outcomes: At 3-month follow-up, patients showed significant improvement since discharge from the partial hospitalisation programme on all continuous outcomes (depressive symptoms, anger expression, and symptom severity (p<.05), as well as hopelessness and dissociation (p<.01); and in self-injury (p<.0001). However, scores on several measures remained in the clinical range. |
| Natural experiment with pre-post comparison. Partial/modified. | Prendergast and McCausland 2007 Australia | To examine the effect of an abbreviated and limited DBT programme on female clients who meet the criteria of BPD within a community mental health team setting. | Treatment: DBT (shortened and partial model - most had access only to either individual or group component).<br><br>Duration/Intensity:<br>Individual DBT: 6-month programme; 24 weekly individual sessions (60-90 minutes).<br>Group DBT: 6-month programme; 24 weekly group therapy sessions (150 minutes).<br><br>Comparator: N/A<br><br>Service setting: Generic community mental health services | Sample Size: 11.<br><br>Demographics: 100% female; mean age 36.35 (SD=7.42); no ethnicity data provided.<br><br>Diagnoses: DSM-IV BPD diagnosis. | No primary outcome specified. Depressive symptoms (BDI); anger (STAXI-2); coping skills (CSA); global functioning (GAF); frequency, medical severity, and intent of parasuicidal and suicidal behaviours (semi-structured interview schedule); number and length of hospitalisations (client files); number and duration of telephone and face-to-face contact (case management system). | Uncontrolled design in which only change over time was measured. No primary outcome specified. Depression (measured with BDI) significantly improved following the DBT programme, and number and length of hospital admissions and amount of face-to-face contact. Significant improvements were not seen on most other measures (but the sample size was only 11). Eleven out of an initial sample of 16 completed the programme. |

| | | | | | | |
|---|---|---|---|---|---|---|
| Quasi-experiment with pre-post comparison. Partial/modified. | Sambrook et al. 2007 UK | To evaluate the impact of Emotional Coping Skills (ECS) groups on clients exhibiting parasuicidal behaviours. | Treatment: Emotional coping skills (DBT based; ACT focus) - Groups were facilitated by two clinical psychologists trained in DBT. The groups balanced change and acceptance. Sessions were divided into reflection on use of current skills (pre-break) and teaching of new skills (post-break). Sessions comprised: Introductions and surviving crises (2 weeks); Introduction to mindfulness (2 weeks); Understanding emotions (2 weeks); Regulating emotions (2 weeks); Tolerating distress (3 weeks); Building skills into everyday life (1 week); Problem solving (1 week); Assertiveness (4 weeks); Preventing relapse (1 week).<br><br>Duration/Intensity: 18-week programme; weekly sessions (120 minutes).<br><br>Comparator: N/A<br><br>Service setting: Standalone outpatient intervention | Sample Size: 26.<br><br>Demographics: 92% female; no additional demographics provided.<br><br>Diagnoses: Parasuicidal behaviours (e.g., cutting, burning, frequent overdosing) in the last 6 months. | Primary outcome: Days spent in hospital or number of outpatient appointments (unclear measure). Secondary outcomes: Global distress (CORE); social and occupational functioning (WSAS). | Uncontrolled study in which comparisons are between timepoints. Primary outcome: Total bed days decreased by 30% across the sample from the 18-months prior to treatment to the 18-months after entry to treatment (no statistical test). For those with no in-patient dates, outpatient appointments reduced by 61%. Significant improvements were also reported in CORE and WSAS scores. |
| Natural experiment with pre-post comparison. Partial/modified. | McQuillan et al. 2005 Switzerland | To examine the effectiveness of an intensive 3-week version of dialectical behaviour therapy for patients in an outpatient setting who met criteria for borderline personality disorder and who were in crisis. | Treatment: DBT (3 week adapted intensive programme) - All patients have an individual therapist who works with them to define behavioural targets that will be the focus of treatment. Suicidal behaviours are treated as a priority, followed by behaviours that interfere with therapy, and then by behaviours that interfere with quality of life. Most group work consists of behavioural skills training.<br><br>Duration/Intensity: 3-week programme; 4-5 times weekly group therapy (13 hours total, 120–240-minute sessions) + individual sessions.<br><br>Comparator: N/A - pre post study<br><br>Service setting: Standalone outpatient intervention | Sample Size: 127.<br><br>Demographics: 81% female; mean age 30.7 (SD=8.1); no ethnicity data provided.<br><br>Diagnoses: 1) BPD diagnosis (IPDE); and 2) recent suicidal or parasuicidal behaviour. Comorbidities: Paranoid (53%); schizoid (33%); schizotypal (51%); histrionic (43%); antisocial (36%); narcissistic (32%); borderline (92%); obsessive-compulsive (57%); dependent (74%); and avoidant (82%) PD. | No primary outcome specified. PD diagnosis (IPDE); depressive symptoms (BDI); hopelessness (BHS); social functioning (SASS). | Uncontrolled study in which comparisons are over time and there is no specified primary outcome. Statistically significant improvements were seen in depression and hopelessness over the treatment period, but not in social adaptation. |
| **3. Tests of partial/modified DBT treatments**<br>c. Uncontrolled intervention development studies | | | | | | |

| | | | | | | |
|---|---|---|---|---|---|---|
| Intervention development/uncontrolled preliminary testing. Partial/modified. | Conrad et al. 2017 Australia | To evaluate the effectiveness of a pilot 10-week group programme based on DBT skills training intervention in reducing psychological symptoms and distress, and to examine the impact of the intervention on mental health service utilization. | Treatment: Short DBT-based group therapy.<br><br>Duration/Intensity: 10-week programme, weekly sessions (60 minutes).<br><br>Comparator: N/A - pre post study<br><br>Service setting: Community mental health centre | Sample Size: 38.<br><br>Demographics: 84% female, mean age 35.13 (range 20–63).<br><br>Diagnoses: Borderline PD (68.4%) and mood or bipolar disorder (31.6%). | No primary outcome specified. Hopelessness (BHS); impulsiveness (BIS); tendency to suppress unwanted thoughts (White Bear Suppression Inventory); quality of life (EQ-5D); service contacts and admissions (service level data). | Uncontrolled pilot study with no specified primary outcome. Over time, the group receiving treatment showed a significant drop in the average number of service contacts in the post-treatment period, along with improvements in clinical measures including hopelessness and cognitive instability, and in self-control and quality of life. |
| Intervention development and uncontrolled preliminary testing. Partial/modified. | Meaney-Tavers and Hasking 2013 Australia | To carry out a preliminary investigation of the effectiveness of a pilot programme, aimed at treating college students with borderline personality disorder (BPD) using short-term, modified group dialectical behavior therapy. | Treatment: DBT based programme (CARE Programme) - The CARE programme was based upon DBT and involved short term delivery of a group-based intervention based on DBT skills training, provided for students.<br><br>Duration/Intensity: 8-week programme; weekly group sessions (120 minutes).<br><br>Comparator: N/A - pre post study<br><br>Service setting: University mental health centre (student population) | Sample Size: 17.<br><br>Demographics: 76.5% female; mean age 22.5 (SD=3.84); no ethnicity data provided.<br><br>Diagnoses: BPD diagnosis | No primary outcome specified. Depressive symptoms (BDI-II); anxiety symptoms (BAI); coping strategies (CSA); BPD symptoms (DSM-IV-TR). | Uncontrolled study in which comparisons are over time and there is no specified primary outcome. Significant improvements reported in symptoms of depression, BPD traits and adaptive coping skills, but not in anxiety. |
| Intervention development and uncontrolled feasibility testing. Partial/modified. | Pozzi et al. 2008 Italy | To explore the effects of a pilot programme that aimed to integrate the group element in DBT with individual psychotherapy and general mental health care for people with Cluster B Personality Disorder. | Treatment: Group DBT added to general mental health care & individual psychotherapy.<br><br>Duration/Intensity: 2 years weekly individual psychotherapy & 6-months DBT group fortnightly<br><br>Comparator: N/A<br><br>Service setting: Specialist PD service | Sample Size: 12.<br><br>Demographics: 9/12 female; mean age 42 (SD= 5.5); no ethnicity data provided.<br><br>Diagnoses: DSM-IV PD cluster B diagnosis. Cluster C (33.3%); cluster A (8.3%); borderline (33.3%); sub-threshold for BPD (8.3%); paranoid (8.3%); and narcissistic (8.3%) PD | No primary outcome specified. Axis II diagnoses (SCID-II); symptom severity (SCL-90: BPRS); level of disability (DISS); global functioning (GAF); aggression (AQ); impulsiveness (BIS-11). | Uncontrolled design in which only change over time was measured. No primary outcome specified. After 24 months, improvements were reported on most sub-scales, but with no significance testing in this sample of only 6 participants. |

| 4. Tests of DBT treatments adapted to specific cohorts<br>a. Randomised Controlled Trials | | | | | | |
|---|---|---|---|---|---|---|
| RCT.<br>Adapted for particular groups/contexts. | Bohus et al. 2020<br>Germany | To test whether DBT-PTSD is more effective than CPT in outpatients with complex PTSD and a history of childhood abuse. | Treatment: DBT-PTSD - an adapted phase-based treatment programme for people with PTSD and a history of child abuse.<br><br>Duration/Intensity: One year programme; up to 45 individual sessions. Followed by 3 additional sessions during following 3 months.<br><br>Comparator: CPT, up to 45 individual sessions within 1 year and 3 additional sessions during the following 3 months.<br><br>Service setting: Standalone outpatient intervention | Sample Size: 193.<br><br>Demographics: 100% female; mean age 36.3 (SD=11.1); no ethnicity data provided.<br><br>Diagnoses: 1) DSM-V diagnosis of PTSD following sexual or physical abuse before age 18 years; and 2) 3 or more BPD criteria, including criterion affective instability. | Primary outcome: PTSD diagnosis (CAPS-5). Secondary outcomes: functioning (GAS); PTSD symptoms (PCL-5); BPD symptoms (BSL-23); depressive symptoms (BDI); dissociative symptoms (DSS). | Primary outcome: Outcome was significantly better for the DBT-PTSD group (group difference: 4.82 [95% CI 0.67, 8.96]; p=.02; d=0.33). Compared with the CPT group, participants in the DBT-PTSD group were also less likely to drop out early, and had higher rates of symptomatic remission, reliable improvement, and reliable recovery. |
| RCT.<br>Adapted for particular groups/contexts. | Kamalabadi et al. 2012<br>Iran | To examine the effect of couple dialectical behaviour therapy (CDBT) on symptoms and quality of marital relationships and mental health of couples in which the male partner is diagnosed with borderline personality disorder. | Treatment: Couple DBT - sessions included: accepting himself and his partner, training to stop making thing worse, being together in close relationship, reacting their relationship, accreting expression, validating responses, recovery from invalidation, managing problem and negotiating solutions and transforming conflict into closeness).<br><br>Duration/Intensity: 14-week programme; weekly sessions.<br><br>Comparator: Waitlist<br><br>Service setting: Standalone outpatient intervention | Sample Size: 30.<br><br>Demographics: 100% male and married; age range 18-50; no ethnicity data provided.<br><br>Diagnoses: BPD diagnosis. | No primary outcome specified. Symptom severity (BPDSI-IV); general mental health (GHQ); relationship satisfaction (PRQC). | No primary outcome specified. The treatment group had significantly lower scores than the control group one month after the end of sessions on measures of BPD symptom and 3 out of 4 subscales of general mental health, and higher scores of 5 subscales of PRQC (satisfaction, commitment, intimacy, passion, and love, but not on trust). |

| | | | | | | |
|---|---|---|---|---|---|---|
| RCT (pilot). Adapted for particular groups/contexts. | Harned et al. 2014 USA | To evaluate the efficacy of integrating PTSD treatment into DBT for women with BPD, PTSD, and intentional self-injury. | Treatment: DBT with DBT PE (prolonged exposure) - in vivo exposure and imaginal exposure followed by processing of the exposure experience. DBT strategies and procedures were incorporated into PE to monitor potential negative reactions to exposure, target problems that may occur during or as a result of exposure and utilise therapist strategies that address the particular characteristics of severe BPD patients. Also included PTSD treatment procedures. The treatment was implemented where patients received either one combined individual therapy session or two individual therapy sessions, as well as group DBT skills training and as needed phone consultation.<br><br>Duration/Intensity: 12-month programme; weekly individual therapy (120 minutes) or twice weekly individual therapy (90 minutes and 60 minutes).<br><br>Comparator: Active comparator (DBT)<br><br>Service setting: Standalone outpatient intervention | Sample Size: 26.<br><br>Demographics: 100% female; mean age 32.6 (SD=12); ethnicities Caucasian 80.8%, Biracial 15.4%, Asian 3.8%.<br><br>Diagnoses: 1) BPD diagnosis; 2) PTSD diagnosis; and 3) can remember at least some part of the index trauma, and recent and recurrent intentional self-injury. | Primary outcomes: PTSD diagnosis (PSS-I); intentional self-injury (SASII). Secondary outcomes: pathological dissociation (DES-T); trauma-related guilt cognitions (Trauma-Related Guilt Inventory; TRGI); shame (ESS); general psychological wellbeing (GSI); depressive symptoms (HRSD); anxiety symptoms (HARS). | Pilot study not powered to detect change at statistically significant level. Primary outcome: Intentional self-injury (suicide attempts) reduction was larger in DBT + DBT PE group (g = 0.6) than DBT (g = 0.4) at end of treatment. At follow-up, 91.7% of patients in DBT + DBT PE (g = 0.5) and 100% of patients in DBT (g = 0.1) were abstinent from suicide behaviour. PTSD severity reductions were greater in DBT + DBT PE group (g = 1.8) than DBT (g = 1.3) at end of treatment, in favour of DBT + DBT PE. At follow-up, the effect sizes were 1.4 for DBT + DBT PE and 0.9 for DBT groups. Dissociation, shame, anxiety, depression, and global severity all decreased at end of treatment and maintained at follow-up in both groups, but not trauma-related guilt cognitions. This was maintained at follow-up. A higher level of satisfaction was reported with combined DBT + PE. |
| **4. Tests of DBT treatments adapted to specific cohorts**<br>b. Non-randomised experiments and observational studies | | | | | | |
| Quasi-experiment with contemporaneous comparison. Adapted for particular groups/contexts. | Lyng et al. 2019 Republic of Ireland | To compare the benefits of DBT delivered in an age specific 18 to 25 group with those of an all-age group for young adults with a BPD diagnosis. | Treatment: Young adult only - DBT. Standard DBT - individual, group and telephone consultations. Only offered to young adults between 18 - to -25 years old.<br><br>Duration/Intensity: 12-month programme; weekly individual therapy (60 minutes) + weekly skills training (150 minutes) + weekly therapist consultation.<br><br>Comparator: General adult DBT. Same as intervention except for it was offered to all adults 18+.<br><br>Service setting: Generic community mental health services | Sample Size: 37.<br><br>Demographics: Treatment group 83% female, comparator group 69% female; only 18-25, mean age 20.5 (SD = 1.91), comparator mean age 21.5 (SD=2.15); no ethnicity data provided.<br><br>Diagnoses: BPD diagnosis. | No primary outcome specified. BPD symptoms (BSL23); symptom severity (SCL-90-R GSI); suicidal ideation (SSI); hopelessness (BHS); service discharge from community services. | No specified primary outcome. No difference in dropout. Greater improvements at a statistically significant level for completers of young adult DBT borderline symptoms and overall symptom severity. Significantly more in the young adult group had been discharged from community services by 24 months after the end of treatment. Differences on other measures not significant. It was suggested that greater social cohesion may be an advantage of youth specific DBT groups. |

| | | | | | | |
|---|---|---|---|---|---|---|
| Natural experiment with contemporaneous comparison. Adapted for particular groups/contexts. | Navarro-Haro et al. 2018 Spain | To Compare Standard Dialectical Behaviour Therapy with a Treatment as Usual Cognitive Behaviour Therapy (TAU CBT) for the treatment of borderline personality disorder when comorbid with an eating disorder. | Treatment: DBT - Standard DBT (Linehan 1993) includes four modes of intervention: individual psychotherapy, skills training, phone calls, and a consultation team. Individual therapy follows the principles and target hierarchy of standard DBT. Skill training consists of weekly group sessions. The aim of this group is to increase behavioural skills related to acceptance and awareness (mindfulness, distress tolerance) and skills related to behavioural change (emotion regulation and interpersonal effectiveness). This group lasted 24 sessions, and contents were taken from Linehan's manual and its version translated into Spanish). The phone call mode is mainly devoted to generalizing skills to daily life and learning how to ask for help.<br><br>Duration/Intensity: 6-month programme; weekly individual therapy (60 minutes) + weekly group skills training (120 minutes).<br><br>Comparator: TAU Cognitive Behavioural Therapy: A cognitive behavioural programme focused mainly on addressing ED psychopathology (awareness of the disorder and education, self-monitoring, establishing regular eating, reducing maladaptive eating behaviours, and changing misinterpretations about body image). In individual therapy, the programme also targets other symptoms that are more related to the personality psychopathology (self-harm, substance use, etc.) using CBT strategies. The TAU CBT was adapted to a group format by the clinical team. The TAU CBT group was adapted by dividing the treatment into three phases: Phase 1 (psychoeducation on adaptive eating and consequences of dysfunctional eating behaviours, and motivation toward the treatment); Phase 2 (cognitive restructuring and normalization of weight, as well as decreasing eating behaviours); Phase 3 (consolidation of the achievements obtained in the two previous phases, generating an internal attribution of the treatment result and relapse prevention). | Sample Size: 118.<br><br>Demographics: 100% female; mean age 27.37; no ethnicity data provided.<br><br>Diagnoses: 1) DSM-IV BPD diagnosis (SCID-II); and 2) DSM-IV eating disorder diagnosis (SCID-I). | Primary outcomes: Suicide attempts and non-suicidal self-injuries; hospitalisations; dysfunctional impulsive behaviours and maladaptive eating behaviours. Secondary outcomes: Diagnoses (DSM-IV-TR); global functioning (GAF); depressive symptoms (BDI-II); emotion regulation (ERQ); affect (PANAS). | There were multiple primary outcomes with differing findings: Outcomes were significantly better for DBT than TAU CBT for frequency of dysfunctional behaviours and non-suicidal self-injuries, but not for frequency of suicide attempts, hospitalisation or dysfunctional eating. Secondary outcomes showed a mixture of significant and non-significant findings: DBT showed greater improvement for depressive symptoms, cognitive reappraisal and global functioning, but there was no significant difference for negative and positive affect, and expressive suppression. |

| | | | | | | |
|---|---|---|---|---|---|---|
| | | | Service setting: Standalone outpatient or day hospital (both interventions delivered in both settings) | | | |
| Natural experiment with pre- post comparison. Adapted for particular groups/contexts. | Williams et al. 2018 Australia | To develop and evaluate a specialised group treatment for mothers with BPD, intended to improve their symptom management and to provide therapeutic guidance with regard to their relationship with infants. | Treatment: mother-infant DBT (MI-DBT) - specialized DBT groups focusing on parenting and the mother-infant relationship. Dyadic reunions were a further therapeutic focus each week. Additional material incorporated from mindfulness/acceptance commitment therapy, distress tolerance/Circle of Security, emotion regulation, interpersonal effectiveness.<br><br>Duration/Intensity: 24-week programme; weekly sessions (150 min).<br><br>Comparator: N/A - pre-post<br><br>Service setting: Standalone outpatient intervention | Sample Size: 29.<br><br>Demographics: 100% female and primary caregiver of at least 1 child younger than 3-years-old; mean age 31.97 (SD=5.88); no ethnicity data provided.<br><br>Diagnoses: Full or partial criteria for BPD diagnosis. Full BPD criteria (75%); and partial BPD criteria (25%). | No primary outcome specified. BPD symptom severity (MSI-BPD; BSL-23); postnatal depressive symptoms (EPDS); anxiety symptoms (BAI); parental self-esteem (PSOC); parental mentalisation (PRFQ); quality of caregiver-infant interaction (CARE Index). | Uncontrolled design in which only change over time measured. No primary outcomes specified. After participation in the programme, patients showed statistically significant improvement on all measures, including BPD, depressive and anxiety symptoms, parental self-esteem and quality of caregiving relationship. 21/29 completed the course of treatment. |
| 4. Tests of DBT treatments adapted to specific cohorts<br>c. Uncontrolled intervention development studies | | | | | | |

| | | | | | | |
|---|---|---|---|---|---|---|
| Intervention development and uncontrolled preliminary testing. Adapted for particular groups/contexts. | Steil et al. 2018 Germany | To investigate the feasibility, acceptance and safety of DBT-PTSD in an outpatient treatment setting. | Treatment: DBT - PTSD - A modular treatment programme. It is based on the principles and methods of Dialectical Behaviour Therapy (DBT; Linehan, 1993) and integrates trauma-focused cognitive and exposure-based interventions. The DBT-PTSD programme follows the DBT hierarchy of treatment targets, which prioritizes life-threatening behaviours, such as suicide attempts, and treatment-interfering behaviours, such as dissociation, over addressing problems reducing quality of life, such as sexual problems.<br><br>Duration/Intensity: 24-week programme; weekly sessions of flexible duration (50-120 minutes).<br><br>Comparator: N/A<br><br>Service setting: Specialist PTSD service | Sample Size: 21.<br><br>Demographics: 100% female; mean age 34.05; no ethnicity data provided.<br><br>Diagnoses: At least four BPD criteria (IPDE) | Primary outcomes: PTSD symptoms (CAPS; DTS); personality status (IPDE); BPD symptom severity (BSL-23). Secondary outcomes: Depressive symptoms (BDI-II); dissociative symptoms (FDS); history of self-harm behaviours and suicide attempts (SBD-I). | Uncontrolled study with comparisons between time points. Primary outcome: Significant reduction for treatment completers in PTSD symptoms (($t(15.12)=-5.44$; $p<.001$, Cohen's $d=1.30$). Significant reductions also observed in borderline symptomatology and dissociative experiences. 17 out of 21 completed the intervention. Self-harm rates were also reported to have fallen. |
| Intervention development and uncontrolled preliminary testing. Adapted for particular groups/contexts. | Lyng et al. 2015 Republic of Ireland | To explore impacts on outcomes and drop-out rate for a DBT programme delivered in an age specific 18 to 25 group. | Treatment: DBT - The five 'modes' of the DBT were included in the programme for younger adults, that is, individual psychotherapy, skills training, telephone consultation, structuring the environment, and therapist consultation group.<br><br>Duration/Intensity: 22-week programme.<br><br>Comparator: N/A - pre-post<br><br>Service setting: Generic community mental health services | Sample Size: 11.<br><br>Demographics: 100% female; age range 18-25; no ethnicity data provided.<br><br>Diagnoses: BPD diagnosis. | No primary outcome specified. BPD symptoms (BSL-23); symptom severity, depressive and anxiety symptoms (SCL-90); DBT coping skills (DBT-WCCL). | Uncontrolled study with comparisons only over time and no specified primary outcome. Statistically significant reductions were found in BPD symptoms and several mental health symptoms through the treatment period alongside an increase in DBT skills use. Dropout was 31% at 22 weeks of treatment. |

#### Appendix 3 – Cognitive and Behavioural Therapy and Schema Therapy


1. Cognitive and Behavioural Therapy treatments vs. non-active comparators
   a. Randomised Controlled Trials ........................................................ p. 38
   b. Non-randomised experiments and observational studies ........... p. 46
   c. Uncontrolled intervention development studies ......................... p. 50
2. Cognitive and Behavioural Therapy treatments vs. specialist comparators
   a. Randomised Controlled Trials ....................................................... p. 55
   b. Non-randomised experiments and observational studies ........... p. 56
   c. Uncontrolled intervention development studies.......................... p. 58
3. Tests of partial/modified Cognitive and Behavioural Therapy treatments
   a. Randomised Controlled Trials ....................................................... p. 59
   b. Non-randomised experiments and observational studies ........... p. 60
4. Tests of Cognitive and Behavioural Therapy treatments adapted to specific cohorts
   a. Uncontrolled intervention development studies ......................... p. 60
5. Schema therapy vs non-active comparators
   a. Randomised Controlled Trials ........................................................ p. 61
   b. Uncontrolled intervention development studies ......................... p. 61
6. Tests of partial/modified Schema therapy
   a. Randomised Controlled Trials ........................................................ p. 63


#### Cognitive and Behavioural Therapy

| Study design and comparator | Paper | Aim | Treatment details | Sample details | Outcomes | Main findings |
|---|---|---|---|---|---|---|
| 1. Cognitive and Behavioural treatments vs. Non-active comparators<br>a. Randomised Controlled Trials | | | | | | |

| | | | | | | |
|---|---|---|---|---|---|---|
| Randomised Controlled Trial. Non-specialist/inactive comparator. | McMurran et al. 2017 UK | The aim of this study was to compare the clinical and cost-effectiveness of PEPS therapy in addition to usual treatment with usual treatment alone in improving social functioning among people with a personality diagnosis. | Treatment: Psychoeducation and problem-solving therapy - Is a complex cognitive behavioural intervention with two distinct components.<br><br>Duration/Intensity: 12-week programme; weekly group sessions (120 minutes) + optional fortnightly support sessions.<br><br>Comparator: Treatment as usual.<br><br>Service setting: Generic community mental health teams | Sample Size: 308.<br><br>Demographics: PEPS group 82% female, TAU group 76% female; PEPS mean age 33.5 (SD=10.46), TAU mean age 37.8 (SD=11); Ethnicities PEPS 84% Caucasian, 4% Mixed, 3% Black-carribean,1% Black-other,8% Other. TAU 83% Caucasian,6% Mixed,1% black-other, 1% asian-indian,1% Asian-other, 4% Other.<br><br>Diagnoses: One or more PD diagnosis (IPDE). Paranoid (TAU: 11%; PEPS: 8%); schizoid (TAU: 1%; PEPS: 3%); schizotypal (0%); antisocial (TAU: 20%; PEPS: 15%); BPD (TAU: 59%; PEPS: 60%); histrionic (TAU: 4%; PEPS: 1%); narcissistic (TAU: 2%; PEPS: 1%); avoidant (TAU: 37%; PEPS: 37%); dependent (TAU: 5%; PEPS: 3%); obsessive-compulsive (TAU: 13%; PEPS: 9%); PD NOS (TAU: 7%; PEPS: 9%) PD. Simple PD (TAU: 51%; PEPS: 40%) and complex PD (TAU: 49%; PEPS: 60%). | Primary outcome: Social functioning (SFQ). Secondary outcomes: Cost-effectiveness and service use; anxiety and depression (HADS); quality of life (EQ-5D). | Primary care: PEPS therapy was no more effective than usual treatment for improving social functioning (adjusted difference in mean Social Functioning Questionnaire scores = –0.73; 95% CI [–1.83, 0.38]; p=.19). PEPS therapy is not an effective treatment for improving social functioning of adults with personality disorder living in the community. The trial was discontinued early because of safety concerns (there was an excess of adverse events in the PEPS arm) and did not achieve intended power. After adjusting for differences in baseline costs, there was a non-significant difference in favour of PEPS (–£1,174, 95% CI [–£3,720, £1,371], p=.19). |
| RCT. Non-specialist/inactive comparator. | Clarke et al. 2014 UK | To investigate the effectiveness of a group-based ACT intervention for "treatment-resistant" participants with various diagnoses, who had already completed at least one psychosocial intervention. | Treatment: Group-based Acceptance and Commitment Therapy<br><br>Duration/Intensity: 16-week programme; weekly sessions (120 minutes) + homework tasks.<br><br>Comparator: Treatment as usual based on Cognitive Behaviour Therapy (TAU-CBT)<br><br>Service setting: Standalone outpatient intervention | Sample Size: 61. Demographics: 67.21% female; mean age 43.46 (SD=12.35); no ethnicity data provided. Diagnoses: Participants with various diagnoses. Clinical psychiatric symptoms (72%); severe levels of depression (76%); PD (51%): depressive PD (1/3); avoidant, obsessive-compulsive, paranoid and borderline personality disorders (each 20–30%) | Primary outcomes: Symptom severity (SCL-90-R); depressive symptoms (BDI-II); Secondary outcomes: personality status (SCID-II); quality of life (WHOQOL). | Primary outcomes: The medium effect size values obtained for GSI (d=.39) and BDI-II (d=.54) at post-therapy reflected mean between-group differences favouring ACT. At follow-up at 6 months, group differences again favouring ACT were reflected in a medium effect size for GSI (d=.51) and a large effect size for BDI-II (d=.90). In comparison with TAU-CBT participants, a significantly greater number of ACT participants made reliable and clinically significant improvements according to scores on the GSI and BDI-II at both post-therapy (respectively, $\chi^2$=4.471, p=.034; $\chi^2$=4.127, p=.042) and follow-up (respectively, $\chi^2$=7.412, p=.006; $\chi^2$=7.519, p=.006). |

| | | | | | | |
|---|---|---|---|---|---|---|
| RCT. Non-specialist/inactive comparator. | Clarke et al. 2013 UK | To investigate the effectiveness of CAT in improving personality disorder outcomes in community settings. | Treatment: Cognitive analytical therapy<br><br>Duration/Intensity: 24-week programme; weekly sessions.<br><br>Comparator: Treatment as usual<br><br>Service setting: Standalone outpatient intervention provided in addition to generic acute community care received by both groups | Sample Size: 99.<br><br>Demographics: 72% female; mean age 36.0 (SD=9.5); no ethnicity data provided.<br><br>Diagnoses: PD diagnosis. Excluded people who self-harmed at least monthly. Diagnosis of two or more disorders (88%); diagnoses across two clusters (53%); across all three clusters (28%); and BPD (68%). | Primary outcomes: PD symptoms (SCID-II); interpersonal problems (IIP). Secondary outcomes: Global distress (CORE); dissociative symptoms (DisQ); frequency of dissociative experiences (DES), symptom severity (SCL-90-R); frequency and duration of A&E attendances and inpatient admissions (healthcare record). | Primary outcome: 9/27 (33%) CAT participants no longer met symptomatic criteria following treatment for any personality disorder, whereas all 30 (100%) TAU participants met the criterion for at least one ($P<0.001$, Fisher's exact test). ANCOVA indicated a significant between-group difference in favour of CAT ($F(1,69) = 16.507$, $p<.001$) with a large ES ($d = 1.00$) for interpersonal problems measured by the ITT. The CAT group also had better outcomes in the CORE, the DisQ, and the PSQ. However significant differences were not found in healthcare use. |
| RCT. Non-specialist/inactive comparator | Gratz et al. 2013 USA | To examine the efficacy of emotion regulation group therapy in a randomized controlled trial (RCT) and the durability of treatment gains over a 9-month uncontrolled follow-up period. | Treatment: Emotion regulation group therapy<br><br>Duration/Intensity: 14-week programme; weekly group sessions (90 minutes).<br><br>Comparator: TAU including individual therapy (TAU waitlist)<br><br>Service setting: Standalone outpatient intervention | Sample Size: 61.<br><br>Demographics: 100% female; ERGT+TAU group mean age 33.3 (SD=11), TAU group 33.0 (SD=10.9); ethnicities ethnic minority 16.1% ERGT+TAU, 26.7% TAU.<br><br>Diagnoses: 1) Threshold or subthreshold diagnosis of BPD; and 2) history of repeated DSH. | No primary outcome specified. Self-destructive behaviours (DSHI; SHI); BPD symptom severity (ZAN-BPD; BEST); depressive symptoms (BDI-II); depression, anxiety, and stress (DASS); interpersonal problems (IIP-BPD); social and occupational impairment (SDS); quality of life (QOLI); emotion dysregulation (DERS); psychological flexibility (AAQ). | No specified primary outcome. Significant effects of ERGT reported on DSH and other self-destructive behaviours, emotion dysregulation, BPD symptoms, depression and stress symptoms, and quality of life. Analyses of all participants who began ERGT (across treatment and waitlist conditions) revealed significant improvement from pre- to post-treatment on all outcomes, additional significant improvements from post-treatment to 9-month follow-up for DSH, emotion dysregulation/avoidance, BPD symptoms and quality of life, and no significant changes from post-treatment to 9-month follow-up on the other measures. |
| RCT. Non-specialist/inactive comparator | Bos et al. 2011 The Netherlands | To investigate whether STEPPS group psychotherapy is effective in a naturalistic sample given a BPD diagnosis in routine settings, and to test whether diagnosis and severity are associated with outcome. | Treatment: STEPPS group therapy (CBT principles) added to individual therapy of varying type and frequency<br><br>Duration/Intensity: 18-week programme; weekly sessions + fortnightly structured individual therapy followed by single follow up session 3-6 months later.<br><br>Comparator: Individual therapy of varying type and frequency<br><br>Service setting: Standalone outpatient therapy. | Sample Size: 168.<br><br>Demographics: STEPPS group 88.1% female, TAU group 85.7% female; mean age STEPPS 33.5 (SD=8.2), mean age TAU: 31.7 (SD=89.7); ethnicity 100% White.<br><br>Diagnoses: DSM-IV BPD diagnosis (SCID-II). | No primary outcome specified. Symptom severity (SCL-90); BPD symptom severity (BPD-40); quality of life (WHOQOL). | No specified primary outcome. STEPPS with individual therapy performed significantly better than individual therapy alone on general (SCL-90) and BPD-specific (BPD-40) psychopathology and on quality of life post-treatment and at follow-up. The superiority of the STEPPS condition was greater with greater initial symptom severity. |

| | | | | | | |
|---|---|---|---|---|---|---|
| RCT.<br>Non-specialist/inactive comparator | Bos et al. 2010 The Netherlands | To compare a version of the STEPPS group programme in which it is combined with basic structured individual therapy with TAU in a community setting. | Treatment: CBT based / STEPPS together with adjunctive structured individual therapy<br><br>Duration/Intensity: 18-week programme; weekly sessions + fortnightly structured individual therapy followed by single follow up session 3-6 months later.<br><br>Comparator: Treatment as usual - individual therapy from a psychotherapist, psychologist, or psychiatric nurse, offered every 1 to 4 weeks. In both conditions, the main treatment could be supplemented with (medication) contacts with a psychiatrist, social worker, or other health care professional.<br><br>Service setting: Standalone outpatient intervention | Sample Size: 79.<br><br>Demographics: STEPPS group 83.3% female, TAU group 89.2% female; STEPPS mean age 32.9 (SD=5.6), TAU mean age 31.8 (SD=9.2); no ethnicity data provided.<br><br>Diagnoses: DSM-IV BPD diagnosis (SCID-II). | Primary outcomes: Symptom severity (SCL-90); BPD symptom severity (BPD-40). Secondary outcomes: Impulsive and parasuicidal behaviour (BPDSI-IV); quality of life (WHOQOL). | Primary outcomes: Statistically significant end of treatment and follow-up differences between STEPPS (experimental group) and TAU for general and borderline symptoms. Secondary outcomes: This was also the case for psychological health and quality of life, but not for impulsive or parasuicidal behaviour. |
| RCT.<br>Non-specialist/inactive comparator | Davidson et al. 2010 UK | To follow up after 6 years participants in a randomised controlled trial comparing a CBT-based intervention with treatment as usual. | Treatment: Cognitive behaviour therapy in addition to treatment as usual (CBT plus TAU)<br><br>Duration/Intensity: 12-month programme; up to 30 sessions (60 minutes).<br><br>Comparator: Treatment as usual<br><br>Service setting: Generic community mental health teams | Sample Size: 106.<br><br>Demographics: 84% female; mean age 31.9 (SD=9.1); ethnicities 100% White.<br><br>Diagnoses: BPD diagnosis (SCID-II). | Primary outcomes: Suicidal acts (DHI); inpatient psychiatric hospitalisation (self-reported or hospital records); A&E department attendance (self-rated or hospital records). Secondary outcomes: Acts of self-mutilation (DHI); depressive symptoms (BDI-II); anxiety symptoms (STAI); symptom severity (BSI); interpersonal problems (IIP-32); social functioning (SFQ); maladaptive schemas (YSQ); quality of life (EQ-5D). | Primary outcome: The gains of CBT–PD over TAU in reduction of suicidal behaviour seen after 1-year follow-up were similar in magnitude to 1 year follow-up but did not reach statistical significance (mean difference adjusted for baseline characteristics = 1.26; 95% CI = -0.06, 2.58; p>.061). There were no significant differences in number of patients self-harming or attempting suicide between the treatment conditions. Secondary outcome: Over half the patients meeting criteria for borderline personality disorder at baseline no longer did so 6 years later, without any differences between groups. There were no differences in clinical outcome measures (BSI, BDI, STAI, IIP-32, SFQ, EQ-5D, YSQ) between the two groups during follow-up. There were no large between-group differences in service utilisation, but inpatient and A&E use was greater in the CPD group and total cost was lower, but not significantly so with adjustment for baseline. |

| | | | | | | |
|---|---|---|---|---|---|---|
| RCT. Non-specialist/inactive comparator | Farrell et al. 2009 USA | To test the effectiveness of adding an eight-month, thirty session schema-focused therapy (SFT) group to treatment-as-usual (TAU) individual psychotherapy for borderline personality disorder (BPD). | Treatment: Schema focused therapy<br><br>Duration/Intensity: 8-month programme; 30 weekly sessions (90 minutes).<br><br>Comparator: Treatment as usual (TAU)<br><br>Service setting: Standalone outpatient intervention | Sample Size: 32.<br><br>Demographics: 100% female; age rage 22–52; no ethnicity data provided.<br><br>Diagnoses: BPD diagnosis (DIB-R; BIS). | No primary outcome specified. BPD symptoms (BSI); symptom severity (SCL-90); BPD diagnosis (DIB-R); global functioning (GAFS). | No primary outcome specified. The treatment group had significantly lower scores at the end of thirty sessions of SFT-group psychotherapy on both measures of BPD symptoms (BSI and DIB-R) and on global severity of psychiatric symptoms (SCL-90); and had higher scores on global functioning (GAFS from individual psychotherapists). On all measures, this positive treatment effect was maintained or even increased at the six-month follow-up, when none of the treatment group and 83% of control group members still met criteria for a BPD diagnosis. |
| RCT. Non-specialist/inactive comparator | Blum et al. 2008 USA | To investigate the effects of adding adjunctive STEPPS to treatment as usual for people with a borderline personality diagnosis. | Treatment: CBT based / Systems Training for Emotional Predictability and Problem Solving (STEPPS)<br><br>Duration/Intensity: 20-week programme; weekly sessions (120 minutes).<br><br>Comparator: Treatment as usual of a variety of types, including individual psychotherapy, medication, and case management.<br><br>Service setting: Standalone outpatient intervention | Sample Size: 124.<br><br>Demographics: 83% female; mean age 31.5 (SD= 9.5); ethnicities Caucasian 94%, African American 2%, Other 3%.<br><br>Diagnoses: DSM-IV BPD diagnosis (SCID) | Primary outcome: BPD symptom severity (ZAN-BPD). Secondary outcomes: symptom severity (CGI; SCL-90-R); depressive symptoms (BDI); impulsiveness (BIS); social functioning (SAS); overall mental health (GAS). | Primary outcome: Significant difference in rate of change over treatment between STEPPS and TAU on Zanari Borderline Personality Disorder (total score) p<0.001 Effect size= 0.84 SE=0.25. Statistically significant differences also observed in CGI severity and improvement ratings, Global Assessment Scale, Beck Depression Inventory, Symptom Checklist-90-Revised Global Severity Index and Social Adjustment Scale total score, with most gains maintained at follow-up. No differences in suicide attempts, self-harm or hospitalisations. |
| RCT. Non-specialist/inactive comparator | Davidson et al. 2006 UK | To compare cognitive behaviour therapy in addition to treatment as usual with treatment as usual alone, for borderline personality disorder. | Treatment: Cognitive behaviour therapy in addition to treatment as usual (CBT plus TAU)<br><br>Duration/Intensity: 12-month programme; up to 30 sessions (60 minutes).<br><br>Comparator: Treatment as usual<br><br>Service setting: Generic community mental health teams | Sample Size: 106.<br><br>Demographics: 84% female; mean age 31.9 (range 18-57); ethnicities 100% White.<br><br>Diagnoses: BPD diagnosis (SCID-II). | Primary outcomes: Suicidal acts (DHI); inpatient psychiatric hospitalisation and A&E visits (self-reported; hospital records). Secondary outcomes: Acts of self-mutilation (DHI); psychiatric symptoms (BDI-II); anxiety (STAI); symptom severity (BSI); interpersonal problems (IIP-32); social functioning (SFQ); maladaptive schemas (YSQ); quality of life (EQ-5D). | Primary outcome: No significant difference was found at either 12 months (the end of the treatment period) or 24 months (the end of the follow-up period) in whether any suicidal act, inpatient hospitalisation or Accident and Emergency department attendance had taken place (CBT plus TAU vs. TAU alone: OR = 1.04 (95% CI 0.52, 2.00, p=.96) at 12 months (end of treatment) and OR = 0.86 (95% CI 0.45, 1.66, p=.66) at 24 months (end of follow-up). Secondary outcomes: There was a significant reduction over the two years in the adjusted mean number of suicidal acts in favour of CBT plus TAU over TAU, with a mean difference of −0.91 (95% CI −1.67, −0.15, p=.02). There were significant differences between CBT plus TAU compared with TAU alone in some secondary measures: State Anxiety, Young's Schema Questionnaire at 24 months and differences on the Brief Symptom Positive Symptom Distress Index at 12 months. Other secondary measures showed no significant difference. |

| | | | | | | |
|---|---|---|---|---|---|---|
| RCT. Non-specialist/inactive comparator | Emmelkamp et al. 2006 The Netherlands | To evaluate the comparative effectiveness of brief dynamic therapy and cognitive–behavioural therapy for patients with avoidant personality disorder as their primary problem. | Treatment: CBT or Brief Dynamic therapy (comparison between these two therapies and waitlist control)<br><br>Duration/Intensity: CBT: 6-month programme; 20 weekly individual sessions (45 minutes). Brief Dynamic therapy: 6-month programme; 20 weekly individual sessions (45 minutes).<br><br>Comparator: Waitlist<br><br>Service setting: Standalone outpatient intervention | Sample Size: 62.<br><br>Demographics: 32/62 female; mean age 34.3 (SD=8.9); ethnicity data not provided.<br><br>Diagnoses: Avoidant PD (SCID–II). | Primary outcomes. PD status (SCID–II); dysfunctional borderline beliefs (PDBQ); anxiety symptoms (LWASQ); social phobia (SPAI). | Primary outcomes: Post treatment, CBT was significantly superior to the control condition on primary outcome measures PDBQ avoidant sub-scale ($F(1,52)=7.39$, $p=.01$) and Avoidance Scale ($F(1,46)=5.39$, $p=.02$). No significant difference was found between BDT and control. CBT was significantly superior to BDT on all primary outcome measures: PDBQ avoidant sub-scale ($F(1,51)=5.92$, $p=.02$), LWASQ ($F(1,51)=5.69$, $p=.02$), SPAI social phobia sub-scale ($F(1,51)=2.98$, $p=.09$) and Avoidance Scale ($F(1,45)=5.25$, $p=.03$), and on the generalisation measure PDBQ obsessive–compulsive sub-scale ($F(1,51)=10.84$, $p=.002$). On none of the measures was BDT superior to CBT. Results were maintained at follow up. |
| RCT. Non-specialist/inactive comparator | Weinberg et al. 2006 USA | To investigate the efficacy of MACT for Deliberate Self Harm (DSH) in patients with BPD. | Treatment: MACT - a 6-session therapy that incorporates elements of DBT, CBT and bibliotherapy. Each session is structured around a chapter of a booklet, covering functional analysis of episodes of parasuicide (i.e., DSH and suicide attempts), emotion regulation strategies, problem-solving strategies, management of negative thinking, management of substance use, and relapse prevention strategies.<br><br>Duration/Intensity: 6-week programme; weekly sessions.<br><br>Comparator: Treatment as usual (TAU)<br><br>Service setting: Standalone outpatient intervention | Sample Size: 30.<br><br>Demographics: 100% female; MACT group mean age 30 (SD=8.61), control group mean age 26.33 (SD=7.67); ethnicities MACT 13 White, 2 Non-white, Control 15 White.<br><br>Diagnoses: 1) BPD criteria (DSM-V; DIB-R); and 2) history of repetitive DSH with at least one episode during the month before enrolment. | No primary outcome specified. BPD diagnosis (DIB-R); dates, method, severity and suicide intent of episodes of DSH (PHI); suicidal risk (SBQ); treatment use (TUI-FA). | No specified primary outcome. There was a significant difference between groups favouring MACT over control in DSH frequency and severity (self-reported), but not in time to repetition or suicidal ideation. |

| | | | | | | |
|---|---|---|---|---|---|---|
| RCT.<br>Non-specialist/inactive comparator | Tyrer et al. 2004 UK | to compare MACT to TAU in a multicentre trial for people with repeated deliberate self-harm episodes and to investigate whether personality status at baseline impacts treatment outcomes, particularly suicide repetition. | Treatment: Manual-assisted cognitive behaviour therapy (MACT) (a brief cognitively oriented and problem-focused therapy covering evaluation of the self-harm attempt, crisis skills, problem-solving, basic cognitive techniques to manage emotions and negative thinking and relapse prevention strategies).<br><br>Duration/Intensity: 6-month programme; up to 5 sessions in first 3 months followed by optional 2 booster sessions in following 3 months.<br><br>Comparator: TAU (standard treatment in the area or continuation of current therapy)<br><br>Service setting: Standalone outpatient intervention | Sample Size: 480.<br><br>Demographics: 68% female; mean age 32 (SD=11); ethnicities 90% White.<br><br>Diagnoses: Recent and previous episode of deliberate self-harm. PD diagnosis (42%). | Primary outcome: Deliberate self-harm (PHI; GP notes, A&E records). Secondary outcome: Costs (CSRI). | Primary outcome: The primary hypothesis - that fewer in the MACT group would repeat self-harm over the follow-up period than in the TAU group - was not confirmed (39% repeated self-harm in the MACT group vs. 46% in TAU, p=0.20). Secondary outcomes: Frequency of self-harm was significantly lower in the MACT group. Health economic analysis suggested that MAU was associated with higher costs of treatment in people with a borderline personality disorder diagnosis and lower costs in the remainder of the sample. |
| RCT.<br>Non-specialist/inactive comparator | Alden et al. 1989 Canada | To compare 3 behavioural interventions for avoidant personality disorder to treatment as usual as well as head-to-head. Secondly to examine whether effects are maintained post treatment. | Treatment: Behavioural treatment-based group therapy - These included 3 types of treatment 1) Graduated exposure (GE), in which subjects progressively learned interpersonal/social skills. 2) Interpersonal skills training (ST), which comprised GE plus additional interpersonal skills training to help establish better relationships and understand others more. 3) Intimacy focus, which comprised GE and ST skills but placed greater emphasis on the development of intimate relationships.<br><br>Duration/Intensity: 10-week programme; weekly group sessions (120 minutes).<br><br>Comparator: Waitlist<br><br>Service setting: Standalone outpatient intervention | Sample Size: 76.<br><br>Demographics: 34/76 female; mean age 27.5 years; ethnicities were 68 White, 8 Chinese.<br><br>Diagnoses: DSM-III avoidant PD diagnosis. | No primary outcome specified. Dispositional shyness (SORT; shyness questionnaire developed for the study); self-esteem (SRI); social functioning (Social targets measure); engagement with social activities (self-report). | Multiple comparisons made via MANCOVA between treatment groups and waiting list control, with no defined primary outcome. All treatment conditions reported to improve significantly more than control on social reticence, symptoms of anxiety and interpersonal functioning with no clear difference among treatment conditions. Gains were maintained at follow-up. |

| | | | | | | |
|---|---|---|---|---|---|---|
| RCT (pilot). Non-specialist/inactive comparator | Popolo et al. 2018 Italy | To investigate the acceptability and clinical effectiveness of MIT-G in a sample of young (18-25) patients diagnosed with mixed PDs (mostly of the over-regulated type) | Treatment: Metacognitive Interpersonal group therapy - Fixed structure sessions divided into blocks of 2 or 3 sessions for each motivation. Motivational structures were 1) social rank/competition, 2) group inclusion/affiliation, 3) attachment, 4) caregiving, 5) exploration, 6) sexuality, 7) cooperation.<br><br>Duration/Intensity: Programme length unclear; 16 sessions (120 minutes)<br><br>Comparator: Waitlist control (TAU). - weekly consultations with clinical psychologists.<br><br>Service setting: Standalone outpatient intervention | Sample Size: 20.<br><br>Demographics: 9/20 female; mean age intervention group 21.3 (SD=.68), mean age TAU group 21.8 (SD=2.04); no ethnicity data provided.<br><br>Diagnoses: Avoidant, dependent, obsessive-compulsive, narcissistic, paranoid, passive-aggressive, depressive, and PD NOS diagnosis (SCID-II). | No specified primary outcome measure. DSM-IV PD diagnosis (SCID-II); global distress (CORE-OM); synthetic metacognitive capacities (MAS-A); emotion regulation (DERS); alexithymia (TAS-20). | Pilot study not powered to detect significant differences; no primary outcome specified. At post-treatment assessment, the MIT-G patients had significantly lower scores on the CORE-OM (mean difference = 2.39, 95% CI 7.41, 0.79, t=2.6, df=18; p=.018; d=1.16). There was also a post-treatment group difference on self-related metacognition with MIT-G patients displaying significantly higher scores (mean difference = 2.40, 95% CI 1.16 to 3.64, t=4.10, df=18; p=.001, d=1.82). There were no significant post- treatment differences between groups on other measures. |
| RCT (pilot). Non-specialist/inactive comparator | Morton et al. 2012 Australia | To conduct a pilot study comparing outcomes from outpatient Acceptance and Commitment Therapy added to Treatment as Usual to Treatment as Usual only. | Treatment: Acceptance and Commitment therapy (ACT) plus TAU - The groups had a psychoeducational format.<br><br>Duration/Intensity: 12-week programme; weekly sessions (120 minutes).<br><br>Comparator: Treatment as usual (TAU)<br><br>Service setting: Standalone outpatient intervention with both experimental and control group continuing care from generic mental health services | Sample Size: 41.<br><br>Demographics: ACT+TAU group 90.5% female, TAU group 95% female; mean age ACT+TAU 35.6 (SD = 9.3), TAU 34 (SD=9), ethnicities ACT+TAU 88% White, 12% Non-white, TAU no ethnicity data provided.<br><br>Diagnoses: Four or more BPD criteria. | Primary outcome: BPD symptoms (BEST). Secondary outcomes: Depression, anxiety, and stress (DASS); hopelessness (BHS); psychological flexibility (AAQ); mindfulness skills (FFMQ); fear of emotions (ACS); emotion regulation (DERS). | Primary outcome: For BPD symptoms (BEST), there was a significant group x time interaction suggesting better outcomes for the ACT + TAU condition at the end of treatment (d = 0.81, 95% CI 1.13, 18.28, p=.028). Significantly better outcomes were also found for ACT + TAU for some other outcomes, including depression, emotion regulation, psychological flexibility, and hopelessness, but not anxiety. At 3-month follow-up fewer than half of participants responded, and the analysis was not repeated in full. |
| RCT (pilot). Non-specialist/inactive comparator | Gratz et al. 2006 USA | To evaluate the efficacy of a new, time-limited, emotion regulation group intervention for self-harm behaviour among women with BPD. | Treatment: Emotion regulation group therapy + TAU including individual therapy.<br><br>Duration/Intensity: 14-week programme; weekly group sessions (90 minutes).<br><br>Comparator: Treatment as usual including individual therapy (TAU waitlist)<br><br>Service setting: Standalone outpatient intervention | Sample Size: 22.<br><br>Demographics: 100% female; mean age 33.32 (SD=9.98); ethnicities 100% White.<br><br>Diagnoses: 1) Five or more criteria for BPD; and 2) score of 8 or higher on R-DIB. | No primary outcome specified. Self-harm (DSHI); emotion regulation (DERS); psychological flexibility (AAQ); BPD symptom severity (BEST); depression, anxiety, and stress (DASS). | No primary outcome specified. Results indicate significant differences between the ERGT and TAU groups on most measures, including symptoms, emotional regulation, and self-harm. |

| | | | | | | |
|---|---|---|---|---|---|---|
| RCT (pilot).<br>Non-specialist/inactive comparator | Evans et al. 1999 UK | To investigate the effectiveness of a new manual-based treatment varying from bibliotherapy (six self-help booklets) alone to six sessions of cognitive therapy linked to the booklets, which contained elements of dialectical behaviour therapy. | Treatment: Manual-assisted cognitive-behaviour therapy (MACT)<br><br>Duration/Intensity: Programme length unclear; 2-6 sessions.<br><br>Comparator: Treatment as usual (TAU)<br><br>Service setting: Generic community mental health services | Sample Size: 34.<br><br>Demographics: no gender data provided; age rage 16-50; no ethnicity data provided.<br><br>Diagnoses: 1) After episode of deliberate self-harm; 2) personality disturbance within the flamboyant personality cluster (antisocial (dissocial), histrionic or emotionally unstable (impulsive and borderline)); and 3) at least one other episode of deliberate self-harm in the previous 12 months. | Primary outcome: Parasuicide events (PHI). Secondary outcomes: service use (standardised measure). | This was identified as a pilot study from which no firm conclusions were expected. Primary outcome: The rate of self-harm episodes was lower in the MACT group but not significantly so (Mann Whitney Test p-0.11). Secondary outcomes: There was a significantly greater reduction in depressive symptoms with MACT on the depression section of the Hospital and Anxiety Depression Scale. Time to next self-harm episode, depression and anxiety symptoms and costs of care all showed trends in favour of MACT compared with TAU but these were not statistically significant. |
| 1. Cognitive and Behavioural treatments vs. Non-active comparators<br>b. Non-randomised experiments and observational studies | | | | | | |
| Observational study with pre post comparison.<br>Non-specialist/inactive comparator. | MacIntosh et al. 2018 Canada | To describe the implementation of the Skills Training in Affective and Interpersonal Regulation (STAIR), a manualized, evidence-based cognitive behavioural group treatment for childhood trauma at Cedar Centre, a community-based trauma treatment centre, and report on a preliminary evaluation of its effectiveness of the treatment. | Treatment: (STAIR) Group CBT - This model is based in attachment theory and interpersonal psychology, but the intervention draws from the cognitive behavioural tradition to assist individuals with childhood trauma.<br><br>Duration/Intensity: 10-week programme; weekly group sessions.<br><br>Comparator: N/A pre-post<br><br>Service setting: Standalone outpatient intervention in social services setting | Sample Size: 85.<br><br>Demographics: 77% female; mean age 43 (SD=11); no ethnicity data provided.<br><br>Diagnoses: Patients with childhood sexual abuse (CSA) | No primary outcome specified. Trauma history (LEC); emotion regulation (DERS); dissociative symptoms (DES - 28 items); PTSD symptom severity (ICD-11 PTSD); interpersonal problems (IIP). | Uncontrolled study with comparisons only over time and no specified primary outcome. Statistically significant improvements were found over time in emotional regulation, interpersonal problems and trauma symptoms. |

| | | | | | | |
|---|---|---|---|---|---|---|
| Quasi-experiment with pre-post comparison. Non-specialist/inactive comparator. | Sahlin et al. 2017 Sweden | To evaluate Emotion Regulation Group Therapy, a relatively brief group treatment approach to deliberate self-harm in BPD, as delivered by community clinicians at 14 psychiatric outpatient clinics. | Treatment: Emotion regulation group therapy - a 14-session, adjunctive, acceptance-based behavioural group treatment developed to treat deliberate self-harm (DSH) by targeting its underlying mechanism of emotion dysregulation. ERGT systematically teaches skills aimed at improving a number of dimensions of emotion regulation; emotional awareness, understanding and acceptance; the ability to control behaviours when experiencing negative emotions; the use of non-avoidant emotion regulation strategies to modulate the intensity and/or duration of emotional responses; and the willingness to experience negative emotions as part of pursuing meaningful activities in life.<br><br>Duration/Intensity: 14-week programme; weekly group sessions (120 minutes).<br><br>Comparator: N/A<br><br>Service setting: Standalone outpatient therapy | Sample Size: 95.<br><br>Demographics: 100% female; age ≥18 year; no ethnicity data provided.<br><br>Diagnoses: 1) ≥3 DSM-IV diagnostic criteria BPD (SCID-II); 2) ≥3 episodes of DSH in the past 6 months (DSHI). | Primary outcome: Frequency of deliberate self-harm (DSHI). Secondary outcomes: Emotion regulation (DERS); self-destructive behaviours (BSL); depression, anxiety, and stress symptoms (DASS); BPD-relevant interpersonal difficulties (IIP); Social and occupational impairment (SDS); treatment credibility and expectancy (Credibility/Expectancy Questionnaire). | Uncontrolled study in which measurements are of change over time. Primary outcome: There was a significant 52% reduction in DSH frequency from pre-treatment to post-treatment (d=0.52, 95% CI 0.30, 0.75) and a 76% reduction from pre-treatment to 6-month follow-up (d=0.99, 95% CI 0.70, 1.30). Results also revealed significant improvements in emotion dysregulation, self-destructive behaviours and depression and stress symptoms from pre-treatment to post-treatment. |
| Quasi-experiment with pre-post comparison. Non-specialist/inactive comparator. | Hill et al. 2016 UK | To assess outcomes of STEPPS delivered by a range of professionals in a UK community mental health care population. | Treatment: STEPPS (systems training for emotional predictability and problem solving) treatment programme: group treatment consisting of cognitive-behavioural principles and skills training with a systems component that includes family members and significant others. Delivered in this study by a range of professionals, not all extensively trained in psychotherapy<br><br>Duration/Intensity: 20-week programme; weekly sessions.<br><br>Comparator: N/A<br><br>Service setting: Generic community mental health teams | Sample Size: 45.<br><br>Demographics: 84.4% female; mean age 34.4 (SD=10.3); ethnicities White British 80%, Other 2.2%, did not state ethnicity 33.3%.<br><br>Diagnoses: BPD diagnosis. | No primary outcome specified. Symptom severity (ZAN-BPD); quality of life (QOL Scale); affinity for maladaptive schemas (Filter Questionnaire). | No primary outcomes specified: comparisons over time only in this uncontrolled observational study. At end of treatment, there were significant improvements in symptom severity (Zanarini-BPD), QoL (effect size = 0.73), p<.001) and maladaptive schemas on all domains of the Filter Questionnaire. |

| | | | | | | |
|---|---|---|---|---|---|---|
| Quasi-experiment with pre-post comparison. Non-specialist/inactive comparator. | Alesiani et al. 2014 Italy | 1) to confirm previously obtained results on STEPPS outcome in a larger sample and at a 12-month follow-up, and 2) to identify predictors of drop-out vs completion of STEPPS. | Treatment: CBT (STEPPS manualised group programme focused on emotion regulation)<br><br>Duration/Intensity: 20-week programme; twice weekly sessions (45 minutes) followed by 6-8 months additional open group.<br><br>Comparator: N/A<br><br>Service setting: Programme initiated during inpatient hospitalisation and continued as outpatient | Sample Size: 32.<br><br>Demographics: 26/32 (81%) female; mean age 44.41 (SD = 9.29; range 26–63); no ethnicity data provided.<br><br>Diagnoses: 1) DSM-IV-TR mood disorder (bipolar or unipolar) diagnosis; 2) DSM-IV-TR BPD diagnosis or severe PD with prominent borderline traits; 3) history of suicidal attempts or self-harm acts; and 4) emotional and behavioural dysregulation even in the euthymic period. | No primary outcome specified. Hospitalisations related to self-harm acts, suicidal attempts, perceived emotional intensity levels (measure unclear); cognitive schemas (Filters Questionnaire); personality symptom severity (BSI-11; AQ; NPI-40; HSNS). | No primary outcome specified. On pre-post comparison, programme completers reported to have a statistically significant reduction in hospitalisations and suicide attempts over 12 months, and in emotional intensity, but no clear changes in measures of personality traits or cognitive filter. Higher histrionic traits and traits on a score of self-transcendence (especially self-forgetting) predicted drop out. |
| Quasi-experiment with pre-post comparison (pilot study). Non-specialist/inactive comparator. | Harvey et al. 2010 UK | To investigate whether STEPPS is likely to prove to be a clinically effective intervention for a UK population. | Treatment: STEPPS (systems training for emotional predictability and problem solving) treatment programme: group treatment consisting of cognitive-behavioural principles and skills training with a systems component that includes family members and significant others.<br><br>Duration/Intensity: 21-week programme; 20 weekly group sessions (120 minutes).<br><br>Comparator: N/A - pre-post<br><br>Service setting: Standalone outpatient intervention - referrals from generic community mental health teams | Sample Size: 38.<br><br>Demographics: 32/38 female; mean age 37 (SD=8.1); ethnicity data not provided.<br><br>Diagnoses: BPD diagnosis. | No primary outcome specified. BPD symptoms (ZAN-BPD); global distress (CORE-OM); depressive symptoms (BDI); mood (PANAS-X); BPD severity (BEST). | No primary outcome specified in this preliminary study. At end of treatment, statistically significant improvements over time in all outcomes: depression, CORE, PANAS-X negative affect and positive affect, ZAN-BPD, and BEST. |
| Quasi-experiment with pre-post comparison. Non-specialist/inactive comparator. | Brown et al. 2004 USA | To examine whether cognitive therapy for BPD is associated with significant improvement on measures of psychopathology. | Treatment: Cognitive therapy<br><br>Duration/Intensity: 12-month programme; weekly sessions (50 minutes) followed by 12 additional sessions as needed.<br><br>Comparator: N/A- pre-post<br><br>Service setting: Standalone outpatient treatment | Sample Size: 32.<br><br>Demographics: 88% female; mean age 29 (range = 20-55); ethnicity 72% Caucasian, 19% African American, 9% Hispanic, Asian or other.<br><br>Diagnoses: 1) At least 4 BPD symptoms; 2) suicide ideation or self-harm behaviour in the past 2 months. | No primary outcome specified. BPD symptoms (SCID-II); suicide ideation (SSI); depressive symptoms (HRSD; BDI-II); hopelessness (BHS); self-harm behaviours (PHI); dysfunctional borderline beliefs (PBQ). | Uncontrolled trial with no specified primary outcome measures. Statistically significant decrease found on all outcome measures over time. Only 16% (4 of 24 assessed) met BPD diagnostic criteria at 18 months follow-up from baseline. |

| | | | | | | |
|---|---|---|---|---|---|---|
| Natural experiment with pre-post comparison.<br>Non-specialist/inactive comparator. | Ryle et al. 2000 UK | To explore the effectiveness of time-limited outpatient cognitive analytic psychotherapy for people with BPD. | Treatment: Cognitive Analytical Therapy - involves the early collaboration of patient and therapist in the identification and Cognitive analytic therapy of BPD characterization of the self-states and of switches between them. These understandings are recorded in writing and in diagrams which become the shared tools of therapy, providing the patient with a new basis for self-rejection and the therapist with a means of avoiding or correcting responses likely to reinforce negative interpersonal patterns and maintain fragmentation.<br><br>Duration/Intensity: Not recorded.<br><br>Comparator: N/A<br><br>Service setting: Standalone outpatient therapy | Sample Size: 27.<br><br>Demographics: 16/27 female; mean age 34.3 (SD=7.5); no ethnicity data provided.<br><br>Diagnoses: DSM-IV BPD diagnosis (PAS). | No primary outcome specified. BPD symptom severity (PAS); depressive symptoms (BDI); symptom severity (SCL-90); interpersonal problems (IIP); social functioning (SQ). | No primary outcome specified. Fourteen patients (52%) no longer met criteria for BPD according to PAS. Significant change was observed on most clinical and social outcome measures, but one-third of the sample had been lost to follow-up at 18 months. |
| Natural experiment including pre post and contemporaneous comparisons.<br>Non-specialist/inactive comparator. | Renneberg et al. 1990 USA | an initial evaluation of our behavioural group treatment programme for APD on intense fear of criticism, extreme fear of rejection, and a negative self-image. | Treatment: Intensive behavioural group treatment - 5-6 patients form a group led by 2-3 therapists. Day One: introduction and history-taking. Day Two: Desensitisation/Paradoxical Intention. Day Three: Behavioural Rehearsal. Day Four: Televised Feedback.<br><br>Duration/Intensity: 4-day programme; 32 hours total group treatment; 2 consecutive days followed by additional 2 consecutive days 1 week later.<br><br>Comparator: Wait-list control - After evaluation, 8 patients were assigned to a 3-months wait-list condition after which they were retested before all entered active treatment.<br><br>Service setting: Standalone outpatient intervention | Sample Size: 17.<br><br>Demographics: 47% female; mean age 34.3 (range 22-63); no ethnicity data provided.<br><br>Diagnoses: DSM-III-R avoidant PD diagnosis (SCID-II). | No primary outcome specified. Fear of rejection and criticism (FNE); negative self-image (PSS); interpersonal fear and functioning (SAD; GRAI); depressive symptoms (BDI); anxiety symptoms (STAI); social functioning (SAS-R). | No primary outcome specified. Most comparisons are regarding change between pre- and post-treatment, indicating significant improvements during treatment and stability over 1 year follow-up after treatment. No change was observed on any measure during the 3 months wait-list period for the control group, although subjects continued with individual treatment at the centre or elsewhere. |
| 1. Cognitive and Behavioural treatments vs. Non-active comparators<br>c. Uncontrolled intervention development studies | | | | | | |

| | | | | | | |
|---|---|---|---|---|---|---|
| Intervention development and uncontrolled preliminary testing. Non-specialist/inactive comparator. | Ng 2005 China (Hong Kong) | To make a preliminary assessment of the efficacy of cognitive therapy for outpatients with refractory depression and obsessive-compulsive personality disorder. | Treatment: Cognitive therapy - taught participants to identify and evaluate key negative automatic thoughts and applied schema re-structuring techniques to dispute core beliefs and to developed more adaptive beliefs and behaviours.<br><br>Duration/Intensity: 18-month programme; weekly sessions (60 minutes).<br><br>Comparator: N/A pre-post.<br><br>Service setting: Standalone outpatient intervention | Sample Size: 10.<br><br>Demographics: 80% female; mean 36.5 (range 28-45); no ethnicity data provided.<br><br>Diagnoses: DSM-IV obsessive compulsive PD diagnosis (SCID-II). Most common PD comorbidities: Cluster C diagnosis; dependent; narcissistic; and borderline PD. | No primary outcome specified. Depressive symptoms (BDI); hopelessness (BHS); anxiety symptoms (BAI); global functioning (GAF); personality disorder beliefs (PBQ); Number of DSM-IV criteria for OCPD (SCID-II). | Uncontrolled study with comparisons only over time and no specified primary outcome. Statistically significant improvements were reported over time on all main outcome measures. |
| Intervention development/uncontrolled feasibility study. Non-specialist/inactive comparator. | Nordahl et al. 2019 Norway | Explore the feasibility, tolerability and preliminary evidence of treatment associated effects of metacognitive therapy for patients with borderline personality disorder and a history of early trauma (a phase-II trial). | Treatment: Metacognitive therapy - The first phase in the protocol was to negotiate a contract and shape the patient's expectation about his/her and the therapist's role in the programme. In addition, there was some planning of the collaboration and availability of the therapist and early involvement of the community service. The second and the third phase focused on self-defeating beliefs and the self-regulatory executive functions of the patient.<br><br>Duration/Intensity: Average programme length 11.5 months; mean number of sessions 26.6.<br><br>Comparator: N/A – pre-post<br><br>Service setting: Standalone outpatient intervention | Sample Size: 12.<br><br>Demographics: 83% female; mean age 32.08 (range 19-51); no ethnicity data provided.<br><br>Diagnoses: Primary BPD diagnosis (SCID-II). | Primary outcomes: Drop-out and attendance rates for patients across treatment. Secondary outcomes: BPD symptoms (DSM-IV); depressive symptoms (BDI-II); anxiety symptoms (BAI); interpersonal problems (IIP-64); PTSD symptoms (PDS); metacognitive beliefs and cognitions (ERIS); quality of life (WHOQOL). | Primary outcome: 100% completion rate. There were no dropouts during the acute treatment phase. 11 patients completed the 1-year follow-up measures, but 11 of 12 filled in the measures at 2-year follow-up. One patient was lost to 2-year follow-up and did not fill in the questionnaires as we were unable to get in contact with her. There were no dropouts from pre- to post-treatment and there was a high retention rate where all attended between 70% and 90% of the sessions offered. Secondary Outcomes: Overall most patients were significantly less symptomatic and showed significant improvements on interpersonal problems and trauma symptoms after treatment and upheld the gains during the 1 to 2-year follow-up. |

| | | | | | | |
|---|---|---|---|---|---|---|
| Intervention development and uncontrolled preliminary testing. Non-specialist/inactive comparator. | Hall et al. 2018 Australia | To pilot an adjunctive ACT-based emotion regulation intervention in individuals with co-occurring BPD symptoms and substance use disorder in an outpatient addictions treatment setting. | Treatment: Emotional regulation intervention: group-based ACT intervention for BPD with a focus on emotion regulation, increasing emotion acceptance, reducing avoidance of both difficult emotions and thoughts, and building other emotion regulation skills. This was delivered as an adjunct to alcohol and other drug (AOD) counselling.<br><br>Duration/Intensity: Programme length unclear; 12 sessions.<br><br>Comparator: N/A pre-post<br><br>Service setting: Both groups receiving outpatient alcohol and other drug (AOD) treatment | Sample Size: 45.<br><br>Demographics: 64.4% female; mean age 35.8 (SD=10.4); ethnicities no ethnicity data provided.<br><br>Diagnoses: 1) Current drug and/or alcohol use disorder requiring treatment; 2) presence of three or more DSM-IV BPD symptoms; and 3) either a current or historic diagnosis of BPD. | Primary outcomes: Alcohol and drug use (ATOP); BPD symptoms (BEST); emotion dysregulation (DERS); psychological flexibility (AAQ-II). Secondary outcome: Treatment engagement (Treatment Engagement form). | Uncontrolled pilot study reporting change over time on multiple primary outcomes. At end of treatment, participants demonstrated significant reduction in number of occasions they had used drugs in the prior 28 days from baseline, BPD symptoms, emotion dysregulation, and acceptance, non-avoidance of thoughts and emotions, and psychological flexibility. Treatment engagement at end of treatment showed high participation, satisfaction, and rapport. |
| Pilot study using single case experimental design. Non-specialist/inactive comparator. | Mohammadi et al. 2018 Iran | The aim of the present study was to conduct a preliminary examination of the efficacy of the UP in treatment of Iranian patients with BPD and comorbid emotional disorders. | Treatment: CBT: Unified Protocol - Transdiagnostic Cognitive Behaviour Therapy for emotional disorders (UP): The UP is an emotion-oriented cognitive-behavioural intervention that has been designed recently to address a range of psychological disorders characterized by emotion dysregulation process as a shared vulnerability.<br><br>Duration/Intensity: 16–20-week programme (tailored to patient); weekly sessions.<br><br>Comparator: N/A pre-post<br><br>Service setting: Standalone outpatient intervention | Sample Size: 6.<br><br>Demographics: 83.3% female; mean age not provided; no ethnicity data provided.<br><br>Diagnoses: 1) BPD diagnosis (SCID-II); and 2) comorbid emotional disorder (SCID-I) | No primary outcome specified. PD symptoms (BPI); emotion regulation (DERS). | Uncontrolled study in which changes in outcome measures are reported separately for each of six participants, with some evidence of improvement over time for each. |

| | | | | | | |
|---|---|---|---|---|---|---|
| Single case series<br>Non-specialist/inactive comparator. | Sauer-Zavala et al. 2016 USA | to conduct a preliminary exploration of the efficacy of the Unified Protocol for Transdiagnostic Treatment of Emotional Disorders UP for treatment of BPD with comorbid depressive and/or anxiety disorders in a clinical replication series consisting of five cases. | Treatment: CBT-based treatment (Unified Protocol for Transdiagnostic Treatment of Emotional Disorders) - cognitive-behavioural intervention developed to address a range of psychological disorders characterized by shared underlying vulnerabilities. Specifically, the UP purports to address neuroticism by extinguishing distress in response to the experience of strong emotions, in turn leading to fewer negative emotions.<br><br>Duration/Intensity: Programme length unclear; 16-20 weekly sessions.<br>Comparator: N/A<br><br>Service setting: Standalone outpatient intervention | Sample Size: 5.<br><br>Demographics: 80% female; age range 19-38; ethnicities 4/5 Caucasian, 1/5 Hispanic.<br><br>Diagnoses: DSM-IV BPD diagnosis | No primary outcome specified. BPD symptom severity (ZAN-BPD); depression, anxiety, stress (DASS); emotion regulation (DERS). | Uncontrolled study in which comparisons are between timepoints in a sample of only 5. Some evidence presented of reductions in borderline symptoms and emotional regulatory capacity at a statistically significant level. |
| Intervention development/ uncontrolled feasibility or pilot study.<br>Non-specialist/inactive comparator. | Videler et al. 2014 Netherlands | To investigate in a proof-of-concept study whether SCBT-g appears is effective in older adults with personality disorder diagnosis or personality disorder features. | Treatment: Short-term group schema cognitive behaviour group therapy (SCBT-g) (in the first stage of the therapy (session 1–9), patients were educated about the schema model, specifically in relation to their own three most prominent EMS and modes. All patients had their own schema workbook in which cognitive techniques were applied to help them test and challenge the distorted views associated with their EMS. In the second stage (session 10–20), patients were tempted to respond to situations that triggered their EMS in a more adaptive manner, using their workbook exercises and role-playing).<br><br>Duration/Intensity: 18-week programme; weekly sessions (90 minutes) followed by 2 monthly follow up sessions (90 minutes).<br><br>Comparator: N/A<br><br>Service setting: Standalone outpatient intervention | Sample Size: 42.<br><br>Demographics: 71% female; mean age 68 (SD=4.6); no ethnicity data provided.<br><br>Diagnoses: Longstanding mood disorder or a chronic adjustment disorder with comorbid PDs or PD features, that had previously been treated by evidence based or best practice-based therapy without significant improvement. PDs (32%): PD NOS; dependent PD; paranoid PD. PD features (39%). Longstanding mood disorder without a comorbid PD or DSM-IV (21%) | Primary outcome: psychological distress (BSI). Secondary outcomes: maladaptive schemas (YSQ L-2); schema modes (SMI). | Uncontrolled study in which comparisons are between time points. Primary outcome: psychological distress decreased significantly from pre-treatment (M=63.58, SD=28.62) to end of- treatment (M=48, SD=28.31) (d=0.54, p<.05). There were also significant improvements in all schema- and coping-related variables. |

| | | | | | | |
|---|---|---|---|---|---|---|
| Single case series with multiple measures. Non-specialist/inactive comparator. | Kellett et al. 2013 UK | To examine change over time (with multiple time points) for patients receiving cognitive analytic therapy (CAT) for borderline personality disorder (BPD) , and to examine therapist competency in delivering this in routine settings. | Treatment: Cognitive Analytical Therapy<br><br>Duration/Intensity: 24 sessions + 4 follow up sessions over 6 months.<br><br>Comparator: N/A<br><br>Service setting: Standalone outpatient intervention | Sample Size: 17.<br><br>Demographics: 14/17 female; mean age female 28.27 (SD=8.7), mean age male 38 (SD=1.73); no ethnicity data provided.<br><br>Diagnoses: 1) DSM-IV BPD diagnosis; and 2) score of 28 or more on PSQ | No primary outcome specified. Identity disturbance (PSQ); dissociative symptoms (DES); general psychological distress (CORE-OM); BPD symptom severity (BSI). | Uncontrolled experimental design with multiple measurements for each participant. No primary outcome specified. Significant progressive reductions were observed over time in psychological distress, dissociation, risk and personality integration, with reductions in distress occurring early in the course of the CAT sessions and personality integration improving at a later stage. |
| Intervention development and uncontrolled preliminary testing. Non-specialist/inactive comparator. | Lucre et al. 2013 UK | To explore effects of Compassion-Focused Therapy on self-criticism and self-attacking thoughts, feelings, and behaviours, as well as the general symptoms of anxiety, stress, and depression among an outpatient group with personality disorder diagnoses. | Treatment: Compassion focused therapy - group therapy with three main components: formulation and psychoeducation, compassionate mind training, and planning for practice. CFT does not encourage clients to spend a lot of time engaging with or challenging self-criticism directly. Rather, the focus is on developing the compassionate attention, thinking, feeling, and behaviour that is linked to the development of soothing affiliative system.<br><br>Duration/Intensity: 16-week programme.<br><br>Comparator: N/A pre-post study<br><br>Service setting: Standalone outpatient | Sample Size: 8.<br><br>Demographics: 77.7% female; age range 18-54; ethnicities 100% Caucasian.<br><br>Diagnoses: ICD 10 PD diagnosis (IPDE). Diagnostic criteria: emotionally unstable, anxious (avoidant), anankastic, paranoid, and histrionic. | No primary outcome specified. Social comparison (social comparison scale); submissive behaviour (SBS); external shame (OAS); critical and reassuring self-evaluative responses (FSCRS); depression, anxiety and stress (DASS21); symptom severity (CORE). | No specified primary outcome - small uncontrolled study with measurements only over time. Significant improvements on most outcomes in the course of therapy. These were no longer statistically significant at one year follow-up. |

| | | | | | | |
|---|---|---|---|---|---|---|
| Intervention development/uncontrolled feasibility/pilot study. Non-specialist/inactive comparator. | Renner et al. 2013 The Netherlands | To test the effects of SCBT-g on global symptomatic distress in young adults with Cluster-B and Cluster-C personality disorders or with personality disorder features in a pilot study. | Treatment: Short term schema cognitive-behavioural group - SCBT-g protocol has a special emphasis on the cognitive and behavioural methods and techniques of schema therapy. The first phase consists of three sessions of psychoeducation; the second consists of seven sessions in which mainly cognitive techniques are used; the third phase lasts seven sessions and is primarily focused on identifying schema triggering events and prevention of schema triggering in the future.<br><br>Duration/Intensity: Programme length unclear; 18 weekly sessions + 2 booster sessions (90min).<br><br>Comparator: N/A<br><br>Service setting: Standalone outpatient intervention | Sample Size: 26.<br><br>Demographics: 17/26 female; mean age 22.5 (range 18-29); no ethnicity data provided.<br><br>Diagnoses: 1) Primary DSM-IV Cluster-B or Cluster-C PD diagnosis; or 2) subthreshold criteria of a DSM-IV axis-II disorder (SCID-II). Subthreshold PD (38.5%); avoidant (23.1%); borderline (19.2%); dependent (11.5%); narcissistic (3.8%); and obsessive compulsive (3.8%) PD. | Primary outcomes: Symptom severity (SCL-90); maladaptive schemas (SQ); schema modes (SMI). | Uncontrolled pilot study where measurements are changes over time. Primary outcome: Global symptomatic distress decreased substantially from pre-treatment to post-treatment (d=0.81, p<.001). Improvements were also seen in some, but not all aspects of maladaptive and adaptive schemas and coping strategies. |
| Intervention development/uncontrolled preliminary testing. Non-specialist/inactive comparator. | Clarke et al. 2012 UK | To pilot test the use of an ACT-based group intervention for a heterogeneous group of treatment-resistant clients. | Treatment: Group-based Acceptance and Commitment Therapy<br><br>Duration/Intensity: 16-week programme; weekly sessions (150 minutes).<br><br>Comparator: N/A - pre-post<br><br>Service setting: Standalone outpatient intervention | Sample Size: 10.<br>Demographics: 9/10 female; mean age 41 (SD=7.81); no ethnicity data provided.<br>Diagnoses: PD diagnosis | Primary outcomes: Symptom severity (SCL-90-R); quality of life (WHOQOL-BREF); depressive symptoms (BDI-II). Secondary outcomes: psychological flexibility (AAQ); mindfulness (MAAS); personality status (SCID-II). | Uncontrolled pilot study. Significant improvements over time for all three primary outcome measures: . F(1, 9)=4.92, p<.01 for GSI; F(1, 9)=3.63, p<.05 for QOL-BREF; F(1, 9)= 4.28, p<.05 for BDI-II. At T2, 50% of participants had either improved or recovered, which rose to 70% at T3, and fell back to 50% at T4. Changes in AAQ and MASS were marginally significant. Improvements were associated with ACT processes of change. |
| Single case series with multiple measures (intervention development and preliminary testing). Non-specialist/inactive comparator. | Wildgoose et al. 2001 United Kingdom | To evaluate the impact of cognitive analytic therapy on personality fragmentation and dissociation in a series of people with borderline personality disorder. | Treatment: Cognitive Analytical Therapy. The central aim of CAT with BPD patients is to provide a higher order understanding of the dissociative processes that maintain their fragmented sense of self and other<br><br>Duration/Intensity: 16-week programme; weekly sessions.<br><br>Comparator: N/A<br><br>Service setting: Standalone outpatient intervention | Sample Size: 5.<br><br>Demographics: 3/5 female; mean age 38; no ethnicity data provided.<br><br>Diagnoses: DSM-IV BPD diagnosis | No primary outcome specified. Axis II disorders (MCMM-II); personality integrity (PSQ); dissociative symptoms (DIS-Q); interpersonal problems (IIP). | A series of measures is individually reported for each of five participants. No primary outcome specified. Reductions in BPD severity were seen for all participants, such that two no longer met the criterion for caseness at the end of treatment and four at the end of 9 months follow-up. Changes on multiple measurements across the treatment period are also reported for most other participants. |

| 2. Cognitive and Behavioural treatments vs. Specialist comparators<br>a. Randomised Controlled Trials | | | | | | |
|---|---|---|---|---|---|---|
| RCT. Specialist/active comparator. | Kallestad et al. 2010 (same sample as Svartberg et al. 2004) Norway | To investigate long-term effectiveness of short-term dynamic psychotherapy (STDP) and cognitive therapy (CT) for reducing symptom severity in Cluster C personality disorders. To explore the role of insight in both STDP and CT for Cluster C personality disorders. | Treatment: Dynamic psychotherapy (short term) - McCullough's Short Term Dynamic Psychotherapy model, which is based on Malan's (1979) triangle of conflict / Cognitive Therapy (CT)<br><br>Duration/Intensity: 40-week programme; weekly sessions (50 minutes).<br><br>Comparator: Active comparator (Dynamic psychotherapy compared with Cognitive Therapy (CT))<br><br>Service setting: Standalone outpatient intervention | Sample Size: 49.<br><br>Demographics: no demographics provided.<br><br>Diagnoses: DSM-III cluster C PD diagnosis. | Primary outcomes: Symptom severity (SCL-90-R); interpersonal problems (IIP). | Primary outcomes: No statistically significant differences between the two treatment groups for symptom severity or interpersonal problems. No further details given. However, levels of insight increased significantly for those who received STDP but not CT at follow-up. |
| RCT. Specialist/active comparator. | Cottraux et al. 2009 France | To compare cognitive therapy (CT) with Rogerian supportive therapy (RST) in borderline personality disorder. | Treatment: Cognitive therapy<br><br>Duration/Intensity: 12 months programme; 6 months of 24 weekly sessions (60 minutes) followed by 6 months of 12 fortnightly sessions (60 minutes).<br><br>Comparator: Active comparator: Rogerian supportive therapy<br><br>Service setting: Standalone outpatient intervention | Sample Size: 65.<br><br>Demographics: 50/65 female; CT group mean age 34.3, RST group 32.6; no ethnicity data reported.<br><br>Diagnoses: At least 5 of 9 DSM-IV BPD criteria. | Primary outcome: Symptom severity (CGI). Secondary outcomes: Depressive symptoms (BDI); suicidal risk (BHS); anxiety symptoms (BAI); maladaptive schemas (YSQ-L-II); impulsivity, venturesomeness, empathy (IVE); self-harming behaviours (SHBCL); social functioning (SAS). | Primary outcome: At week 24, 13 of 26 patients (50%) in CT versus 7 of 25 (28%) in RST had improved (p = 0.15). At week 52, 10 of 20 patients (50%) in CT versus 12 of 18 (66.7%) in RST had improved (p=.34). At week 104, 8 of 10 patients (80%) in CT versus 6 of 11 (54%) in RST had improved (p=.36). Thus, no clear overall evidence of a statistically significant difference between therapies. Significant differences not found on outcome measures. |
| RCT. Specialist/active comparator. | Giesen-Bloo et al. 2006 the Netherlands | To compare the effectiveness of schema-focused therapy (SFT) and psychodynamically based transference-focused psychotherapy (TFP) in patients with borderline personality disorder. | Treatment: Schema focused therapy<br><br>Duration/Intensity: 36-month programme; twice weekly sessions (50 minutes).<br><br>Comparator: Active comparator: Transference focused therapy<br><br>Service setting: Standalone outpatient intervention | Sample Size: 86.<br><br>Demographics: 80/86 female; SFT group mean age 31.70 (SD=8.89), TFP group 29.45 (SD=6.47); no ethnicity data provided.<br><br>Diagnoses: Primary BPD diagnosis. | Primary outcomes: BPD recovery (BPDSI-IV). Secondary outcomes: Quality of life (WHOQOL); BPD symptoms (BPD-45); symptom severity (SCL-90); self-esteem (SES); actual-ideal self-discrepancy (Miskimins Self-Goal-Other Discrepancy Scale); maladaptive schemas (YSQ); BPD-specific beliefs (BPD); personality functioning (IPO); defence mechanism (DSQ). | Primary outcome: After 3 years of treatment, survival analyses demonstrated that significantly more SFT patients recovered (46% recovered in SFT group vs. 24% in TFP group, RR (relative risk) =2.18; p=.04) or showed clinically significant improvement (66% in SFT group vs. 43% in TFT group, RR=2.33; p=.009) on the Borderline Personality Disorder Severity Index, fourth version. Secondary outcomes: The SFT group also improved more on measures of symptoms and quality of life. |

| | | | | | | |
|---|---|---|---|---|---|---|
| RCT. Specialist/active comparator. | Svartberg et al. 2004 Norway, Canada | To compare the effectiveness of short-term dynamic psychotherapy and cognitive therapy for outpatients with cluster C personality disorders. | Treatment: Dynamic psychotherapy (short term) / Cognitive therapy. Short-term dynamic psychotherapy with the overall goal for previously avoided affects, e.g., sadness/grief or tenderness, to be experienced and expressed adaptively by the patient. Cognitive therapy with the goal to help the patient develop new and more adaptive core beliefs and help the patient to develop more adaptive problem-solving interpersonal behaviours.<br><br>Duration/Intensity: 40-week programme; weekly sessions (50 minutes).<br><br>Comparator: Active comparator (dynamic psychotherapy compared with cognitive therapy)<br><br>Service setting: Standalone outpatient therapy | Sample Size: 51.<br><br>Demographics: ST dynamic psychotherapy group 56.0% female, CT group 44% female; mean age 33.4 (SD=9.7), mean age CT 34.6 (SD=7.9); ethnicities 100% Caucasian.<br><br>Diagnoses: DSM-III-R cluster C or self-defeating PD. | No primary outcome specified. Symptom severity (SCL-90-R GSI); interpersonal problems (IIP), cluster C DSM-III PD (MCMI). | No primary outcome specified. No significant difference found on any outcome between dynamic and cognitive therapy groups. Two years after treatment 54% of dynamic therapy and 42% of cognitive therapy patients had recovered symptomatically. |
| 2. Cognitive and Behavioural treatments vs. Specialist comparators<br>b. Non-randomised experiments and observational studies | | | | | | |
| Quasi-experiment with contemporaneous comparison. Specialist/active comparator. | Chakhssi et al. 2015 The Netherlands | To compare the effectiveness of an ACT programme in a day hospital setting with a day hospital programme including CBT, delivered to people with a personality disorder who have not benefited from previous treatments. | Treatment: ACT<br><br>Duration/Intensity: 26-week programme; twice weekly sessions (360 minutes).<br><br>Comparator: Group based TAU-CBT intervention of the same duration om the same setting (two specialised day hospitals for treatment of people with a personality disorder diagnosis who have relapsed following previous outpatient treatments).<br><br>Service setting: Specialist Day hospital setting for both groups | Sample Size: 81.<br><br>Demographics: 82.7% female; mean age 32.98 (SD=9.94); ethnicity data not provided.<br><br>Diagnoses: DSM-IV PD (SCID-II). Borderline (49.4%); PD NOS (46.9%); avoidant (59.3%); dependent (21.0%); obsessive compulsive (12.4%); antisocial (3.7%) PD | Primary outcome: Personality functioning (SIPP-SF). Secondary outcomes: Psychosocial functioning (OQ-45); psychological flexibility (AAQ); coping styles (UCL); autonomy and social optimism (POS); quality of life (WHOQOL-BREF). | Primary outcome: No significant effect found for group allocation, although there was a substantial improvement from baseline to post-treatment. Similar findings for most secondary measures. |

| | | | | | | |
|---|---|---|---|---|---|---|
| Observational study. Specialist/active comparator. | Horn et al. 2015 The Netherlands | To investigate the effectiveness of different modalities of psychotherapy in patients with PDNOS, i.e., short-term (up to 6 months) and long-term (more than 6 months) outpatient, day hospital, and inpatient psychotherapy. | Treatment: 6 different treatments: Longterm outpatient treatment / Short term outpatient treatment / Long term day hospital / Short term day hospital / Long term inpatient / Short term inpatient (mixed orientation).<br>All treatments varied in theoretical orientations depending on treatment centres, such as psychodynamic (27% of all given treatments), CBT (21% of all given treatments) or an integrative orientation (combining different theoretical frameworks; 52% of all given treatments). Day hospital and inpatient programmes typically consisted of group psychotherapy combined with individual psychotherapy, coaching for social problems, non-verbal or expressive group therapies, discussions about household tasks and living together, community meetings and/or pharmacological treatment.<br><br>Duration/Intensity: Short-term treatments lasted up to 6-months and long-term treatments lasted more than 6-months.<br><br>Outpatient psychotherapy: individual or group psychotherapy sessions, up to 2-sessions per week. Day hospital psychotherapy: 1-session per week. Inpatient psychotherapy: Patients staying at the institutions for 5-days per week.<br><br>Comparator: Naturalistic comparison between six active treatments.<br><br>Service setting: Standalone outpatient interventions | Sample Size: 205.<br><br>Demographics: 72% female; mean age 35.1 (SD=10.3); ethnicity data not provided.<br><br>Diagnoses: PD NOS diagnosis. | Primary outcome: Symptom severity (BSI - Dutch version; GSI). Secondary outcomes: Psychosocial functioning (OQ-45), quality of life (EQ-5D). | Primary outcome: At 60-months after baseline, symptom severity significantly improved across all groups and no significant differences were found between groups with correction for baseline differences. The largest effect was found in short-term day hospital (d=1.42), followed by long-term inpatient (d=1.35), short-term inpatient (d = 1.31), long-term day hospital (d=1.17), long-term outpatient (d=1.14), and lastly short-term outpatient (d=0.91). Some differences were found at earlier time points, with the long-term inpatient psychotherapy group performing less well than other modalities at 12 months. Secondary outcomes: psychosocial functioning and quality of life also improved at 60-months in all groups (except for QoL in short-term day hospital and psychosocial functioning in short-term outpatient and short-term day hospital). |

| | | | | | | |
|---|---|---|---|---|---|---|
| Quasi-experimental design with contemporaneous comparisons (patients involved in choice of treatment). Specialist/active comparator. | Ivaldi et al. 2007 Italy | To compare double setting (individual and group therapy) with individual group therapy for cognitive-evolutionary therapy (DS-CET vs. I-CET). | Treatment: Double-setting cognitive-evolutionary therapy (DS-CET): integrated individual and group therapy which consists of theoretical and methodological contributions from attachment theory, cognitive-behavioural therapy, control mastery theory, interpersonal therapy, and intersubjective group therapy.<br><br>Duration/Intensity: 24-month programme: twice monthly group sessions (120 minutes) + twice monthly individual sessions (60 minutes).<br><br>Comparator: Active comparator (individual cognitive-evolution therapy; I-CET)<br><br>Service setting: Standalone outpatient interventions | Sample Size: 109.<br><br>Demographics: DS-CET group 66% female, I-CET group 38% female; mean age 31.4 DS-CET, 30.4 I-CET; no ethnicity data provided.<br><br>Diagnoses: One of the following DSM-IV diagnoses: 1) BPD, in association or not with other axis I disorders; 2) other cluster B PDs in comorbidity with axis I disorders; 3) transversal comorbidity in axis I (meeting DSM-IV criteria for more than one disorder); or 4) longitudinal comorbidity (persons who along time met the DSM-IV criteria for more than axis I disorders). | No specified primary outcome. Drop-out; global functioning (GAF); symptoms and functioning (BASIS-32); quality of life (QoL-I); self-harming behaviour and substance abuse. | No specified primary outcome. The main data presented are for change from baseline for each treatment. Drop out is significantly higher for the individual CET group than for the double setting group. By 24 months, significant improvements were found for both treatment groups on all outcomes. |
| **2. Cognitive and Behavioural treatments vs. Specialist comparators**<br>c. Uncontrolled intervention development studies | | | | | | |
| Single case experimental design with multiple baselines and crossover between interventions (part of preliminary testing). Specialist/active comparator. | Bentley et al. 2017 USA | To examine the specific effects of mindful emotion awareness training and cognitive reappraisal, two transdiagnostic treatment strategies that target processes underlying self-injurious behaviour. | Treatment: Mindful emotion awareness and cognitive reappraisal - from the Unified Protocol for Transdiagnostic Treatment of Emotional Disorders.<br><br>Duration/Intensity: 2- or 4-week programme.<br><br>Comparator: 4 conditions: 2-week baseline mindful emotion awareness, 4-week baseline mindful emotion awareness, 2-week baseline cognitive reappraisal, and 4-week baseline cognitive reappraisal.<br><br>Service setting: Standalone outpatient intervention | Sample Size: 10.<br>Demographics: 9/10 female; mean age 21.3; ethnicity: 6/10 White, 2/10 Asian, 1/10 multiracial, 1/10 other.<br><br>Diagnoses: DSM-V non-suicidal self-injury (NSSI) disorder. | Primary outcome: non-suicidal self-injury (Ecological momentary assessment). Secondary outcomes: anxiety symptoms and impairment (OASIS); depressive symptoms and impairment (ODSIS), mindfulness (SMQ); emotion regulation (ERQ-R). | Eight out of ten participants reported to demonstrate clinically meaningful reductions in NSSI, 6 in response to one intervention only and two more after an additional intervention was added. Reductions were also reported in measures of symptoms, mindful emotion awareness and cognitive reappraisal skills. |
| **3. Tests of partial/modified Cognitive and Behavioural treatments**<br>a. Randomised Controlled Trials | | | | | | |

| | | | | | | |
|---|---|---|---|---|---|---|
| RCT. Partial/modified. | Salkovskis et al. 1990 UK | To evaluate a cognitively based problem-solving treatment in a population at high risk for repeated self-harm, delivered in the patients' own homes. | Treatment: Cognitive behavioural problem solving - Patients were taught how to identify problems and arrange priorities for problem solving. The next stage was to teach patients how to generate a wide range of solutions and narrowing this down to attainable goals. Next, strategies necessary to work out and implement steps towards realising these goals were considered, together with ways of determining and monitoring success. Emphasis was also placed on the importance of being flexible on the basis of results obtained, then deciding new goals. Homework assignments were also used.<br><br>Duration/Intensity: Programme length 1 month: five treatment sessions (at least 60 minutes).<br><br>Comparator: Treatment as usual (generally discharge to General Practitioner)<br><br>Service setting: Initiated in hospital emergency setting and continued at home | Sample Size: 20.<br><br>Demographics: intervention group 58% female, control group 38% female; intervention mean 26.4 years old (SD=6.0), control mean 28.5 (7.9); no ethnicity data provided.<br><br>Diagnoses: Patients had to fulfil 2/3 of the following: 1) ≥2 previous suicide attempts; 2) antidepressants taken as part of an overdose; 3) patients scored ≥ 4 on scale to predict subsequent suicidal behaviour (Buglass & Horton (1974)). | Primary outcome: Suicidal ideation (BSSI). Secondary outcomes: Tension (POMS); depressive symptoms (BDI); hopelessness (BHI). | Primary outcome: An effect of group was seen on one of the sub-scales of the BSSI, but not the other (Scale 1 $F(1,17)=1.59$, $p>.05$; Scale 2 $F(1,17)=6.08$, $P<.025$). Most change appeared to occur very early in the intervention. Results on secondary outcomes were mixed and analysis reported to be limited by the small sample. |
| Pilot RCT. Partial/modified. | Morey et al. 2010 USA | To conduct a pilot investigation of the effectiveness of Manual Assisted Cognitive Therapy (MACT) as a stand-alone treatment for Borderline Personality Disorder (BPD) with suicidal ideation, and of its enhancement with a Therapeutic Assessment (TA) intervention. | Treatment: Manual Assisted Cognitive Therapy (MACT) plus therapeutic assessment - MACT is a 6-session, manualized therapy that targets deliberate self-harm, incorporating elements of other cognitive-based interventions for BPD. Adaptations were made to the first two-sessions of the TA+MACT condition manual to incorporate the intervention according to Finn's (2007) Therapeutic Assessment model.<br><br>Duration/Intensity: Programme length unclear; 6 sessions.<br><br>Comparator: Manual Assisted Cognitive Therapy (MACT)<br><br>Service setting: Standalone outpatient intervention | Sample Size: 16.<br><br>Demographics: MACT group 75% female, TA+MACT group 88% female; mean age MACT 29.63 (SD=8.7), mean age TA+MACT 32.5 (SD=9.4); ethnicities MACT 88% White, 12% non-white, TA+MACT no ethnicity data provided.<br><br>Diagnoses: BPD diagnosis | No primary outcome specified. BPD (PAI; PDQ-4); suicidal ideation (SUI; SPS). | No primary outcome specified; pilot study not powered to detect differences. Attrition was noted as a problem, with only 44% completing treatment. Most measures showed no statistically significant difference between groups in this small sample. |

| | | | | | | |
|---|---|---|---|---|---|---|
| **3. Tests of partial/modified Cognitive and Behavioural treatments**<br>b. Non-randomised experiments and observational studies | | | | | | |
| Quasi-experiment - crossover design with within patient comparisons. Partial/modified. | Weertman and Arntz 2007 Netherlands | To test if cognitive therapy for treatment of childhood memories by means of imagery with rescripting and historical role plays is an effective method for the treatment of PDs compared to cognitive therapy for present concerns. | Treatment: Cognitive therapy for childhood memories. Therapy consisted of 12 sessions of pre-therapy exploration. Then patients either received 24 sessions focused on the present followed by 24 sessions focused on childhood memories, or the reverse.<br><br>Duration/Intensity: 61 sessions of weekly 1-hour sessions.<br><br>Comparator: Active - Cognitive Therapy focused on present<br><br>Service setting: Standalone outpatient intervention | Sample Size: 21.<br><br>Demographics: 15/21 female; mean age 35.6 (range 20–52); no ethnicity data provided.<br><br>Diagnoses: At least one DSM-IV PD diagnosis, other than borderline, schizotypal, schizoid, antisocial PD, PD NOS. Avoidant (23.8%); paranoid (19%); dependent (9.5%); obsessive-compulsive (33.3%); histrionic (9.5%); narcissistic (4.8%) PD. | No primary outcome specified. Self-esteem (RSES); symptom severity (SCL-90); personality functioning (DPQ; PDBQ); maladaptive schemas (SQ); actual-ideal self-discrepancy (MSGO). | No specified primary outcome. No significant difference was found between treatment phases with a focus on past memories and a focus on the present in a crossover design where each patient received each treatment. Significant improvements were observed over the treatment period as a whole. |
| **4. Tests of Cognitive and Behavioural treatments adapted to specific cohorts**<br>a. Uncontrolled intervention development studies | | | | | | |
| Intervention development and uncontrolled preliminary testing. Adapted for particular groups/context. | Skewes et al. 2015 Australia | To investigate the outcome of ST-g (group schema therapy) delivered in a pilot study for a group of participants with mixed personality disorders in an outpatient university clinic. | Treatment: Group schema therapy - Adapted from group schema cognitive-behavioural therapy protocol (SCBT-g). Adapted model had a strong focus on experiential techniques and mode work for a diagnostically mixed group of personality disorder patients (with a predominant diagnosis of Avoidant personality disorder).<br><br>Duration/Intensity: 5-month programme; weekly sessions (60 minutes).<br><br>Comparator: N/A.<br><br>Service setting: Standalone outpatient intervention | Sample Size: 8.<br><br>Demographics: age range 28-42; no additional demographics provided.<br><br>Diagnoses: At least one DSM-IV TR PD (SCID-II). | Symptom severity (SCL-90-R GSI); maladaptive schemas (YSQ-S2); schema modes (SMI). | Uncontrolled study in which statistical analysis is not presented as numbers are very small. Five of eight participants reported no longer to meet criteria for avoidant personality disorder by the end of follow up. |
| **5. Schema therapy vs. Non-active comparators**<br>a. Randomised Controlled Trials | | | | | | |

| | | | | | | |
|---|---|---|---|---|---|---|
| Randomised Controlled Trial. Non-specialist/inactive comparator. | Bamelis et al. 2014 The Netherlands | To compare the effectiveness of schema therapy with clarification-oriented psychotherapy and with treatment as usual among people diagnosed with cluster C, paranoid, histrionic, or narcissistic personality disordered. | Treatment: Schema therapy<br><br>Duration/Intensity: 24-month programme; 40 weekly sessions first 12 months followed by 10 booster sessions in second 12 months.<br><br>Comparator: Treatment as usual, N=135; clarification-oriented psychotherapy, N=41<br><br>Service setting: Standalone outpatient intervention | Sample Size: 323.<br><br>Demographics: Schema therapy 45.5% male, COP 43.95% male, TAU 41% male; mean age Schema therapy 37.57 (SD=9.69), COP 39.20 (SD=9.37), TAU 38.06 (SD=9.63); no ethnicity data provided.<br><br>Diagnoses: Primary DSM-IV diagnosis of avoidant, dependent, obsessive-compulsive, paranoid, histrionic, or narcissistic PD (SCID-II). | Primary outcome measure: recovery from personality disorder (SCID II). Secondary outcomes: axis I mood and anxiety disorders (SCID I; SCID II); global and social and occupational functioning (global assessment of functioning scale); symptom severity (SCL-90); social functioning (WSAS); actual-ideal self-discrepancy (Miskimins Self-Goal-Other Discrepancy Scale); quality of life (WHOQOL). | Primary outcome: Schema therapy was dominant over treatment as usual, with a significantly greater proportion of recovered patients in this group than in the treatment as usual group (odds ratio for recovery in schema therapy group vs. TAU 4.073 (95% CI 1.774–9.350), p=0.002). Schema therapy was also associated with better outcomes than clarification-focused therapy (odds ratio 2.916 (95% CI 1.043–8.157, p=0.041)). Significant differences were not reported between clarification-focused therapy and treatment as usual. For secondary outcomes, Global assessment of Functioning and Social and Occupational Functioning Assessment Scale were significantly higher for schema therapy recipients, but significant differences were not found for symptom measures, Work and Social Adjustment Scale or WHO Quality of Life Assessment Scale. |
| 5. Schema therapy vs non-active comparators<br>b. Uncontrolled intervention development studies | | | | | | |
| Single case experimental design with multiple measurement timepoints. Non-specialist/inactive comparator. | Videler et al. 2018 Netherlands | To test the effectiveness of schema therapy for personality disorders in older adults. | Treatment: Schema therapy (In the treatment phases, ST, according to the methods described by Young et al. (2003), was provided. In the CBT phase, underlying EMS were targeted by cognitive and behavioural techniques. The experiential phase started with the use of experiential techniques such as imagery rescripting and chair work)<br><br>Duration/Intensity: Program length unclear; 40 sessions followed by 10 booster sessions during a 6 month follow up period.<br><br>Comparator: N/A<br><br>Service setting: Standalone outpatient intervention | Sample Size: 8.<br><br>Demographics: 6/8 female; 60 years or older: mean age 69.3 (SD=3.8); no ethnicity data provided.<br><br>Diagnoses: Primary DSM-IV diagnosis of a cluster C PD or PD NOS with cluster C traits (SCID-II). Avoidant (n=3); PD NOS (n=3); obsessive-compulsive (n=1); dependent (n=1) PD. | Primary outcome: Strength of idiosyncratic beliefs (visual scale). Secondary outcomes: DSM PD diagnosis (Dutch SCID-II); symptom severity (SCL-90); idiosyncratic target complaints (Likert scale); quality of life (WHOQOL-BREF); maladaptive schemas (YSQ). | Multiple scores on measures are individually presented for all seven participants. Primary outcomes: large changes found in dysfunctional core beliefs during the treatment for all but one participant. Secondary outcomes: large improvements were also observed on other outcomes for most participants. At the end of treatment, none of the seven still met criteria for a PD diagnosis. |

| | | | | | | |
|---|---|---|---|---|---|---|
| Intervention development and uncontrolled preliminary testing. Non-specialist/inactive comparator. | Fassbinder et al. 2016 USA | To investigate whether a Group Schema Therapy programme can be implemented in a German University outpatient treatment centre under routine mental health care conditions and whether is effective even in patients with high BPD severity, high comorbidity, and a history of frequent hospitalization. | Treatment: Schema therapy<br><br>Duration/Intensity: 12-month program; weekly group sessions (100 minutes) + weekly individual sessions (60 minutes).<br><br>Comparator: N/A - pre and post<br><br>Service setting: Standalone outpatient intervention | Sample Size: 10. Demographics: 100% female; mean age 35 (SD=13); no ethnicity data provided. Diagnoses: Primary BPD diagnosis (SCID-II). High level of comorbidities (Affective disorder (100%); anxiety disorders (90%); PTSD (70%) | Primary outcome: BPD severity (BPDSI-IV). Secondary outcomes: BPD symptoms (BPD-40); symptom severity (BSI); global functioning (GAF); social and occupational functioning (SOFAS; WSAS); quality of life (WHO QOL-short; EQ-5D); happiness (1-item happiness question); maladaptive schemas (SMI; YSQ); days of hospitalisation. | Uncontrolled feasibility study. Primary outcome: A significant reduction in the overall severity of BPD-symptoms with a large ES occurred at the end of treatment and at 3-year follow-up. Improvements over time were also seen on most other outcome measures. |
| Intervention development and uncontrolled preliminary testing. Non-specialist/inactive comparator. | Dickhaut and Arntz. 2014 The Netherlands | To conduct a pilot investigation of the feasibility and outcomes of combining group and individual modalities in schema therapy. | Treatment: Schema therapy combining group and individual modalities<br><br>Duration/Intensity: 24-month program; weekly group sessions (90 minutes) + weekly individual sessions (60 minutes) for at least the first year.<br><br>Comparator: NA - pre and post<br><br>Service setting: Community mental health centre | Sample Size: 18.<br><br>Demographics: 100% female; mean age 28.5 (SD=8.7); no ethnicity data provided.<br><br>Diagnoses: Primary DSM-IV BPD diagnosis (SCID-II) | Primary outcome: BPD symptoms (BPDSI-IV). Secondary outcomes: Quality of life WHOQOL; EQ-5D); Happiness (1-item question validated); maladaptive schemas (YSQ; SMI). | This was an uncontrolled pilot study not powered to find a significant effect. Primary outcome: At 24 months, at the end of treatment, the recovery rate in terms of participant no longer meeting PD criteria was 14 out of 18 (77.4% (95%CI 45.9, 93.3). Improvements were also seen on other secondary outcomes. |
| Single case series for preliminary testing of intervention. Non-specialist/inactive comparator. | Nordahl et al. 2005 Norway | To evaluate the effectiveness of Young's schema therapy with a limited number of patients with primarily a diagnosis of BPD. | Treatment: Schema therapy - therapy involved 5 steps: 1) to develop a schema mode formulation of the patient. 2) to bond with the patient through re-parenting. 3) work on interpersonal coping skills. 4) enhance problem-solving. 5) gradual termination of schema therapy<br><br>Duration/Intensity: Average program length 22 months; weekly sessions (60 minutes).<br><br>Comparator: N/A - pre post study<br><br>Service setting: Standalone outpatient intervention | Sample Size: 6.<br><br>Demographics: 100% female; age range 19-42; no ethnicity data provided.<br><br>Diagnoses: DSM-IV BPD diagnosis | No primary outcome specified. Symptom severity (SCL-90R); depressive symptoms (BDI); anxiety symptoms (BAI); interpersonal problems (IIP); maladaptive schemas (YSQ); general functioning (GAF). | No primary outcome - report on very small case series in a pilot study of schema therapy. Five out of six patients were reported to have made large improvements in symptoms, maladaptive schemas and general functioning, such that three of the six patients did not fulfil the criteria of DSM-IV BPD at post-treatment, and the rest fulfilled the requirements to a lesser extent than before treatment. |

| 6. Tests of partial/modified Schema therapy<br>a. Randomised Controlled Trials | | | | | | |
|---|---|---|---|---|---|---|
| RCT. Partial/modified. | Nadort et al. 2009 Netherlands | To evaluate the success of implementing outpatient schema focused therapy for people with BPD in routine mental health care, and to test whether outcomes are improved by out of hours crisis support by therapists. | Treatment: Schema therapy (ST) plus phone support - Central to ST is the assumption of 5 schema modes specific for BPD. Schema modes are sets of schemas expressed in pervasive patterns of thinking, feeling, and behaving. Change is achieved through a range of behavioural, cognitive, and experiential techniques.<br><br>Duration/Intensity: 36-month program; twice weekly sessions (45 minutes) in first 12 months followed by weekly sessions (45 minutes) for remaining 24 months.<br><br>Comparator: Schema therapy without phone support<br><br>Service setting: Standalone outpatient intervention | Sample Size: 62. Demographics: ST+phone support group 96.9% female, ST group 96.7% female; mean age ST+phone support 31.8 (SD = 9.2, ST mean age 32.1 (SD=9.1); ethnicities ST+phone support 88% White, 12% Non-white, ST group no ethnicity data provided.<br><br>Diagnoses: DSM-IV BPD diagnosis | Primary outcome: BPD criteria (BPDSI-IV). Secondary outcomes: Quality of life (EQ-5D; WHOQOL); BDP symptoms (BPD-47); symptom severity (SCL-90); maladaptive schemas (YSQ L2). | Primary outcome measure: No significant difference was found at 1.5 years from baseline in recovery from BPD (42% of the patients had recovered with added phone support, 43% without added phone support). Secondary outcome measures did not indicate a significant value to added phone support on any measure. |

#### Appendix 4 – Mentalisation Based Therapy and Psychodynamic Therapy

1. MBT vs. Non-active comparators
    a. Randomised Controlled Trials ........................................................ p. 65
    b. Non-randomised experiments and observational studies ........... p. 67
2. MBT vs. specialist comparators
    a. Randomised Controlled Trials ........................................................ p. 70
    b. Non-randomised experiments and observational studies ........... p. 72
3. Tests of Partial/modified MBT treatments
    a. Randomised Controlled Trials ........................................................ p. 75
4. Psychodynamic therapy treatments vs. non-active comparators
    a. Randomised Controlled Trials ........................................................ p. 76
    b. Non-randomised experiments and observational studies ........... p. 79
    c. Uncontrolled intervention development studies ......................... p. 91
5. Psychodynamic therapy treatments vs. specialist comparators
    a. Randomised Controlled Trials ........................................................ p. 91
    b. Non-randomised experiments and observational studies ........... p. 95
6. Tests of Psychodynamic therapy treatments with different modes of delivery
    a. Non-randomised experiments and observational studies ........... p. 98
7. Tests of Psychodynamic therapy treatments adapted to settings/cohorts
    a. Randomised Controlled Trials ........................................................ p. 103
    b. Non-randomised experiments and observational studies ........... p. 104

| Study design and comparator | Paper | Aim | Treatment details | Sample details | Outcomes | Main findings |
|---|---|---|---|---|---|---|
| **1. MBT vs. non-active comparators**<br>a. Randomised Controlled Trials | | | | | | |
| RCT. Non-specialist/inactive comparator. | Khabir et al. 2018 Iran | To investigate and compare clinical outcomes of DBT and MBT for people with BPD in an Iranian setting. | Treatment:<br>DBT – DBT based group therapy.<br>MBT – MBT based group therapy.<br><br>Duration/Intensity: Programme length unclear; twice weekly sessions (120 minutes).<br><br>Comparator: Medication only.<br><br>Service setting: Standalone outpatient intervention. | Sample Size: 51 (treatment completers N=36).<br><br>Demographics: 25/36 female; mean age 22.61 (only 18-27); no ethnicity data provided.<br><br>Diagnoses: BPD diagnosis. | Primary outcome: BPD symptoms (BPDSI-IV). Secondary outcomes: Anxiety symptoms (BAI), depression symptoms (BDI-II). | Primary outcome: Both treatments were more effective than the control treatment (involving medication only (p=.0001)) in reducing BPD symptoms, but no difference was found between MBT and DBT (p=.4). Similar patterns were seen at follow-up two months after the end of treatment and for secondary outcomes. |
| RCT. Non-specialist/inactive comparator. | Bateman et al. 2008 UK | To investigate whether gains were maintained from a psychoanalytically oriented partial hospitalization programme 8 years after the inception of the 18-month programme. | Treatment: MBT day hospital<br><br>Duration/Intensity: 18-month programme.<br><br>Comparator: Standard psychiatric outpatient care with medication, community support from MH nurses, and periods of partial hospital and inpatient treatment as necessary.<br><br>Service setting: Specialist Day service (compared with generic community services including day care) | Sample Size: 41.<br><br>Demographics: Partially hospitalised/control: Age 30.3(5.86), 33.3(6.60), female-13 (68), 9 (47).<br><br>Diagnoses: DSM-III BPD diagnosis (SCID-II; DIB-R, cut off 7+). | Primary outcomes: number of suicide attempts (medical records); service use (medical records: days of hospitalisation, nr of emergency room visits; further psychiatric outpatient treatment; further therapy; further assertive outreach treatment; years on antidepressants; years on antipsychotics; years on mood stabilisers; 3/+ drugs). Secondary outcomes: symptom severity (ZAN-BPD); global functioning (GAF). | Primary outcome: 23% of experimental group patients made at least one suicide attempt compared with 74% of controls (d=1.4; 95% CI 1.3 to 1.5; p=0.00004). Multiple other outcomes were also reported as better in the experimental group. At end of the follow-up, 13% of the experimental group BPD, vs 87% of TAU met criteria for a personality disorder diagnosis and use of psychiatric outpatient services and medication were lower and global functioning and vocational status better than in the experimental than the control group. |

| | | | | | | |
|---|---|---|---|---|---|---|
| RCT.<br>Non-specialist/inactive comparator. | Bateman and Fonagy 2001 UK | To determine whether the gains made by patients with borderline personality disorder following a psychoanalytically oriented partial hospitalization programme, in comparison to standard psychiatric care, were maintained over an 18-monthfollow-up period. | Treatment: Psychoanalytically Orientated Partial Hospitalisation (prototype for MBT)<br><br>Duration/Intensity: programme length unclear; weekly individual psychotherapy + 3 times a week group therapy (60 minutes) + weekly expressive therapy (60 minutes) + weekly community meeting (60 minutes)<br><br>Comparator: Standard psychiatric care- regular psychiatric review, inpatient care where needed with discharge to a non-psychoanalytic day setting focused on problem solving (72% average length of stay of 6m), outpatient and community follow-up at least every 2 weeks.<br><br>Service setting: Specialist Day service (compared with generic community services including day care) | Sample Size: 44.<br><br>Demographics: Partially hospitalised 68% female, control 47% female; partially hospitalised mean age 30.3 (SD= 5.86), control mean age 33.3 (SD =6.60); no ethnicity data reported.<br><br>Diagnoses: DSM-III BPD diagnosis (SCID-II; DIB-R, cut off 7+). | No primary outcome specified. Suicide and self-damaging acts (SSHI; medical records); hospital admission and length of stay (hospital data; medical records); symptom severity (SCL-90-R); depressive symptoms (BDI); anxiety symptoms (STAI); social functioning (SAS-R; IIP). | No primary outcome specified. Significantly more experimental than control participants had refrained from self-harm or from attempting suicide, and fewer experimental group participants had been admitted over the 18 months following discharge from the study treatment. There were also significant group effects on symptoms and on social functioning. |

| | | | | | | |
|---|---|---|---|---|---|---|
| RCT.<br>Non-specialist/inactive comparator. | Bateman et al. 1999 UK | To compare the effectiveness of psychoanalytically oriented partial hospitalization with standard psychiatric care for patients with borderline personality disorder. | Treatment: Psychoanalytically Orientated Partial Hospitalisation (prototype for MBT)<br><br>Duration/Intensity: programme length unclear; weekly individual psychotherapy + 3 times a week group therapy (60 minutes) + weekly expressive therapy (60 minutes) + weekly community meeting (60 minutes)<br><br>Comparator: Standard psychiatric care- regular psychiatric review, inpatient care where needed with discharge to a non-psychoanalytic day setting focused on problem solving (72% average length of stay of 6m), outpatient and community follow-up at least every 2 weeks.<br><br>Service setting: Specialist Day service (compared with generic community services including day care) | Sample Size: 38.<br><br>Demographics: Partially hospitalised 68% female, control 47% female; partially hospitalised mean age 30.3 (SD= 5.86), control mean age 33.3 (SD =6.60); no ethnicity data reported.<br><br>Diagnoses: DSM-III BPD diagnosis (SCID-II; DIB-R, cut off 7+). | No primary outcome specified. Suicide and self-damaging acts (SSHI; medical records); hospital admission and length of stay (hospital data; medical records); symptom severity (SCL-90-R); depressive symptoms (BDI); anxiety symptoms (STAI); social functioning (SAS-R; IIP). | No primary outcome specified. Greater statistically improvements reported for experimental group than for control in parasuicidal behaviour, medication use, depression, anxiety, global severity, and social adjustment. |
| 1. MBT vs. non-active comparators<br>b. Non-randomised experiments and observational studies | | | | | | |
| Quasi-experiment with pre-post comparison.<br>Non-specialist/inactive comparator. | Beattie et al. 2019 Ireland | To investigate the feasibility of mentalization-based treatment (MBT) for patients with personality disorder in a non-specialist setting. | Treatment: MBT group<br><br>Duration/Intensity: 21–27-month programme; 12 sessions group mentalisation psychoeducation followed by weekly group sessions (75 minutes) + weekly individual sessions (50 minutes).<br><br>Comparator: N/A – pre-post<br><br>Service setting: Generic community mental health service | Sample Size: 8.<br><br>Demographics: 100% female; mean age 42.25(S.D.=7.92); no ethnicity data provided.<br><br>Diagnoses: PD diagnosis. | No primary outcome specified. Interpersonal problems (IIP-64); social functioning (WSAS); symptom severity (SCL-90-R); psychological wellbeing (SOS-10); personality functioning (MCMI-III). | No primary outcome specified. There were insufficient numbers in this feasibility study to run meaningful statistical analyses, but some reductions in symptoms and improvements in function were noted over the treatment period. |

| | | | | | | |
|---|---|---|---|---|---|---|
| Quasi-experimental with pre-post comparison. Non-specialist/inactive comparator. | Carrera et al. 2018 Italy | To investigate whether an MBT path could be implemented in an Italian public mental health setting, and (in the second cohort) to examine change in mentalization in greater detail. | Treatment: Oriented mentalization-based treatment<br><br>Duration/Intensity: Programme length unclear; 45 individual sessions (50 minutes) + 45 group sessions (90 minutes).<br><br>Comparator: N/A - pre-post<br><br>Service setting: Public mental health centre | Sample Size: 15.<br>Demographics: 11/15 female; age and ethnicity data not provided.<br>Diagnoses: DSM-V BPD diagnosis. | No primary outcome specified. Symptom severity (SCL-90-R); global health functioning (HONS); personality status (SCID-II); global functioning (GAF); service impact and service costs (PES, folder data). | Uncontrolled study with no specified primary outcome measure. Significant improvements reported from baseline to end of treatment and follow-up on clinical and social problems (HoNOS), global functioning (GAF) and some measures of symptoms and personality features. |
| Observational study with comparison with a historical cohort. Non-specialist/inactive comparator. | Kvarstein et al. 2015 Norway | To investigate whether MBT, as implemented in a Norwegian specialist treatment unit, has been more effective for BPD patients than the traditional psychodynamic treatment programme delivered before MBT was introduced? | Treatment: Mentalization-based treatment (MBT). The MBT followed guidelines (Bateman & Fonagy, 2006) and manuals for individual (Karterud & Bateman, 2010), psychoeducational (Karterud & Bateman, 2011), and group MBT (Karterud, 2012). Three sections of MBT treatment: 1) individual MBT, 2) MBT psychoeducational group 3) MBT dynamic group<br><br>Duration/Intensity: Up to 36 months programme; 12 months weekly individual MBT + 12 psychoeducational group meetings + weekly dynamic groups followed by 12 months fortnightly individual MBT + continued group meetings followed by 12 months every third week individual MBT + continued group meetings.<br><br>Comparator: Psychodynamic treatment programme (Treatment as routinely provided before the introduction of MBT)<br><br>Service setting: Specialist Day service | Sample Size: 345.<br><br>Demographics: psychodynamic treatment (n = 281), 83% female; mean age 30 (SD=7); MBT treatment (n = 64), 84% female; mean age 26 (SD=6); ethnicity data not provided.<br><br>Diagnoses: BPD diagnosis. Axis II comorbidities: Paranoid, obsessive-compulsive, dependent, schizoid, and narcissistic PD. | No primary outcome specified. Duration of treatment; symptom severity (BSI-18); interpersonal problems (CIP); global functioning (GAF). | No primary outcome specified. Greater improvements were reported over treatment in symptom distress, and interpersonal, global, and occupational functioning for the MBT programme than for the traditional psychodynamic programme delivered before the MBT programme was initiated in 2018. Large reductions in suicidal/self-harming acts, hospital admissions, and use of medication were found following both treatments: significant differences were not reported. |

| | | | | | | |
|---|---|---|---|---|---|---|
| Observational study with pre-post comparison. Non-specialist/inactive comparator. | Löf et al. 2018 Sweden | To observe outcomes of implementing MBT in a psychiatric outpatient setting in Sweden. | Treatment: MBT - MBT was conducted according to the treatment manual developed by Bateman and Fonagy. Patients were offered individual sessions with psychotherapists and groups sessions (6 - 8 participants).<br><br>Duration/Intensity: 18-month programme; 9-12 introductory psychoeducation followed by weekly individual sessions + twice a week MBT group sessions + twice a week expressive group sessions. From August 2008 expressive group sessions removed. From June 2009 changed to one session each of individual and group.<br><br>Comparator: N/A – pre-post study<br><br>Service setting: Standalone outpatient intervention | Sample Size: 97.<br><br>Demographics: 89.3% female; mean age 30.4 years (SD = 7.7);<br>ethnicity data not provided.<br><br>Diagnoses: DSM-IV and ICD-10 BPD diagnosis (SCID-II; ZAN-BPD). | Primary outcome: General symptoms (KABOSS-S). Secondary outcomes: Suicidality (SUAS-S); symptom severity (SLC-90-R); self-harm (DSHI-9); alexithymia (TAS-20); self-image (SASB). | Uncontrolled pre-/post study describing change over time only. Primary outcome: Borderline symptomatology (KABOSS-S) improved significantly over treatment and up to follow-up 18 months after baseline (d=0.79, p<.001 from mixed linear model). There were also significant improvements in suicidality, alexithymia, self-image, and general symptoms. Severe patients improved as much as less severe patients, even though they had worse symptoms at baseline. |
| Quasi-experiment with pre-post comparison. Non-specialist/inactive comparator. | Bales et al . 2012 The Netherlands | To investigate feasibility and outcomes of delivering manualized day hospital MBT in a cohort with severe MBT in the Netherlands. | Treatment: Day hospital delivering Mentalization-Based Treatment (MBT)<br><br>Duration/Intensity: 18-month initial day hospital programme; 5 days a week (270 minutes). Followed by 18-month group therapy.<br><br>Comparator: N/A - pre-post<br><br>Service setting: Specialist Day hospital | Sample Size: 45.<br><br>Demographics: 71.1% female; mean age 30.1 (SD = 6.5); no ethnicity data provided.<br><br>Diagnoses: DSM-IV BPD diagnosis (SCID-II). | No primary outcome specified. Treatment commitment (measure unclear); symptom distress (SCL-90-R GSI); depressive symptoms (BDI); social and interpersonal functioning (IIP-C); personality functioning (SIPP-118; BPDSI); quality of life (EQ-5D); suicide attempts and self-harming behaviour (SSHI); service use (measure unclear). | No primary outcome specified. Quality of life, general symptom distress, depression severity, borderline symptomatology, interpersonal functioning, and social role functioning all improved within 18 months of starting treatment. No suicides occurred during treatment in the study population, but one patient died by suicide four months after dropping out of treatment. |

| | | | | | | |
|---|---|---|---|---|---|---|
| Natural experiment with pre post comparison. Non-specialist/inactive comparator. | Petersen et al. 2010 Denmark | To analyse the effectiveness of long-term mentalization-oriented outpatient group therapeutic intervention in a sample of patients who initially received a short-term day hospital treatment. | Treatment: Mentalisation-orientated group therapy - patients who had completed 5 months of the day hospital treatment in Peterson et al (2008). A psychodynamic group therapy based on promoting mentalisation.<br><br>Duration/Intensity: Programme length unclear; weekly group therapy (30 minutes).<br><br>Comparator: N/A<br><br>Service setting: Specialist Day service followed by standalone outpatient care | Sample Size: 22.<br><br>Demographics: 100% female; mean age 28.5 (SD=6.1); no ethnicity data provided.<br><br>Diagnoses: 1) DSM-IV PD diagnosis; and 2) GAF score <50. | Primary outcomes: The Personality Severity Index Score (PSI); interpersonal problems (IIP-C). Secondary outcomes: Symptom severity (SCL-90-R GSI); general functioning (GAF); Clark's Personal and Social Adjustment Scale (CPSAS), time spent unemployed, service use. | Uncontrolled study with comparisons only over time. Primary outcomes: There were significant improvements following outpatient in personality traits (p=.02; β=−0.0036), interpersonal problems (p=.02; β=−0.0042) and global severity of symptoms (SCL-90 GSI) was significantly reduced (p=.02; β=−0.0043). Low rates of hospitalisation and emergency service use continued from the initial day hospital treatment programme through and beyond the outpatient therapy programme, and months of unemployment per year diminished progressively through out-patient treatment and follow-up. |
| **2. MBT vs. Specialist comparators**<br>a. Randomised Controlled Trials | | | | | | |
| RCT. Specialist/active comparator. | Laurenssen et al. 2018 Netherlands | To compare outcomes of MBT offered in a day hospital setting with a well-established specialist treatment as usual service. | Treatment: Day Hospital MBT (MBT-DH) consists of a highly structured day hospitalization programme offering an MBT programme.<br><br>Duration/Intensity: 18-month programme; 5 days a week (6 hours).<br><br>Comparator: Specialist TAU (S-TAU) involving evidence-based psychotherapy depending on the needs of the patient. Inpatient treatment is offered to some of this group in a hospital ward offering individual and group therapy.<br><br>Service setting: Specialist Day services | Sample Size: 95.<br><br>Demographics: MBT-DH 78% female, S-TAU 81% female; MBT-DH mean age 34.00 (SD=9.38), S-TAU 34.00 (SD=10.62); no ethnicity data provided.<br><br>Diagnoses: DSM-IV BPD diagnosis (SCID-II). | Primary outcome: BPD symptoms (BPDSI: PAI-BOR). Secondary outcomes: Symptom severity (SCID-I; SCID-II; SCL-90 GSI); depressive symptoms (BDI); interpersonal problems (IIP-64); quality of life (EQ-5D-3L). | Primary outcome: No significant difference was found between MBT-and S-TAU on borderline symptom severity at 18 months (coefficient 3.43 (95% CI 3.72, 10.57; p>.05 on mixed effect). Other outcomes were also not significantly different between groups, with substantial improvements from baseline on most measures. MBT-DH had a lower dropout rate than S-TAU (9% vs. 34%). |

| | | | | | | |
|---|---|---|---|---|---|---|
| RCT. Specialist/active comparator. | Jørgensen et al. 2014 (FU to Jørgensen et al. 2013) Denmark | To investigate the 18-months follow-up symptom and interpersonal functioning outcomes of a randomised controlled trial comparing MBT for BPD to supportive group therapy. | Treatment: MBT (combined individual and group)- In the combined MBT, the focus of attention was the patient's relationship with the therapist and with other people, including other group members. In accordance with the MBT treatment manual, the overall aim of the treatment was to develop the patient's ability to mentalize (establish a mentalizing stance, resembling the concept of de-centring in cognitive therapy) and develop more adaptive interpersonal behaviours by working through chains of interpersonal events and emotions, using mentalizing functional analysis, stop-stand-and-rewind techniques, etc.<br><br>Duration/Intensity: 24-month programme: 18 months of weekly individual sessions (45 minutes) + 18-20 months of weekly group sessions (90 minutes) starting 3 months after individual therapy. Comparator: biweekly group therapy (1.5h).<br><br>Comparator: Supportive group therapy (SP) - focused primarily on the individual in the group.<br><br>Service setting: Specialist PD service | Sample Size: 111.<br><br>Demographics: MBT group 96% female, comparator group 95% female; MBT mean age 29.2 (SD=6.1), comparator mean age 29.0 (SD=6.4); no ethnicity data provided.<br><br>Diagnoses: BPD diagnosis; excluding antisocial or paranoid PDs. | No primary outcome specified. Symptom severity (SLC-90-R GSI); depressive symptoms (BDI-II); anxiety symptoms (STAI; BAI); social functioning (SAS-SR); interpersonal problems (IIP); global functioning (GAF). | No specified primary outcomes. 58 patients completed 2 years of treatment. There were no between-group differences on self-reported measures of depression, anxiety, interpersonal and general functioning, with both groups making large improvements. Only therapist-rated level of functioning was significantly higher in the MBT group, whereas change in self-rated social functioning was higher in the supportive treatment group. More than three quarters of patients did not meet diagnostic criteria for BPD at 18-month follow-up in both groups (78% MBT; 80% SP). |

| | | | | | | |
|---|---|---|---|---|---|---|
| RCT (single site). Specialist/active comparator. | Jørgensen et al. 2013 Denmark | To investigate effectiveness of MBT for BPD compared to supportive group therapy for symptom severity and social functioning. | Treatment: MBT (combined individual and group)- In the combined MBT, the focus of attention was the patient's relationship with the therapist and with other people, including other group members. In accordance with the MBT treatment manual, the overall aim of the treatment was to develop the patient's ability to mentalize (establish a mentalizing stance, resembling the concept of de-centring in cognitive therapy) and develop more adaptive interpersonal behaviours by working through chains of interpersonal events and emotions, using mentalizing functional analysis, stop-stand-and-rewind techniques, etc.<br><br>Duration/Intensity: 24-month programme: 18 months of weekly individual sessions (45 minutes) + 18-20 months of weekly group sessions (90 minutes) starting 3 months after individual therapy. Comparator: biweekly group therapy (1.5h).<br><br>Comparator: Supportive group therapy (SP) - focused primarily on the individual in the group.<br><br>Service setting: Specialist PD service | Sample Size: 111.<br><br>Demographics: MBT group 96% female, comparator group 95% female; MBT mean age 29.2 (SD=6.1), mean age comparator 29 (SD=6.4); no ethnicity data provided.<br><br>Diagnoses: BPD diagnosis; excluding antisocial or paranoid PD. At least one personality disorder other than borderline (MBT: 65%; avoidant PD: 22%). | No primary outcome specified. Symptom severity (SLC-90-R GSI); depressive symptoms (BDI-II); anxiety symptoms (STAI; BAI); social functioning (SAS-R); interpersonal problems (IIP); global functioning (GAF). | No primary outcome specified. 58 patients completed 2 years of treatment. There were significant changes for all outcome measures in the MBT group, including general functioning, social functioning, symptoms, and number of diagnostic criteria met for BPD (SCID-II), and most outcomes except of anxiety symptoms (BAI) and general functioning (GAF-F) in the SP group (p<.005). Only GAF showed a significantly higher outcome in the MBT group (GAF-F: F = 8.0, p=.005; GAF-S: F = 12.7, p=.0004). A trend was found for a higher rate of recovery from BPD in the MBT group. Conclusion: The study indicates that both MBT and supportive treatment are highly effective in treating BPD when conducted by a well-trained and experienced psychodynamic staff in a well-organized clinic. |

| | | | | | | |
|---|---|---|---|---|---|---|
| RCT. Specialist/active comparator. | Bateman and Fonagy 2009 UK | To test the hypothesis that patients receiving outpatient MBT would be more likely to desist from para-suicidal behaviour (self-harm and suicide attempts) and require less hospitalization than those offered an outpatient structured protocol of similar intensity but excluding MBT components. | Treatment: MBT<br><br>Duration/Intensity: 18-month programme; weekly sessions<br><br>Comparator: Structured Clinical Management of Borderline Personality Disorder (SCM) - manual developed to reflect best clinical practice, including regular individual and group sessions were offered with appointments every 3 months for psychiatric review.<br><br>Service setting: Outpatient treatment offered in specialist personality disorder centres | Sample Size: 107.<br><br>Demographics: MBT 80.3% female, SCM- 79.4% female; mean age MBT 31.3 (7.6.), SCM 30.9 (SD=7.9); ethnicities MBT White British/European 76.1%, SCM 68.3%, MBT Black African/ Afro-Caribbean 15.5%, SCM 20.6%, MBT Other Chinese/Turkish/Pakistani/ 8.5%, SCM 11.1% .<br><br>Diagnoses: 1) DSM-IV BPD diagnosis; and 2) suicide attempt or episode of life-threatening self-harm within last 6 months. | Primary outcome: severe parasuicidal behaviour, including suicide attempt, life-threatening self-harm, hospital admission (medical records). Secondary outcomes: Global functioning (GAF); symptom severity (SCL-90-R); depressive symptoms (BDI); social functioning (SAS-R; IIP-C); medication use (medical records). | Primary outcome: The MBT group were significantly more likely than the SCM group to have experienced six months free of suicidal behaviours, severe self-injurious behaviours, and hospitalization (73% vs. 43%; x2=11.5, df=1, p<.0007; relative risk=1.7, 95% CI 1.23, 2.35). Secondary outcomes: Measures of symptoms and symptom-related distress, and of interpersonal and social functioning improved in both groups, but significantly more so in the MBT group. |
| **2. MBT vs. Specialist comparators**<br>b. Non-randomised experiments and observational studies | | | | | | |
| Natural experiment with contemporaneous control. Specialist/active comparator. | Barnicot et al. 2019 UK | To investigate whether clinical outcomes at 12 months in naturalistic personality disorder treatment settings differ between people receiving DBT and those receiving MBT. | Treatment: DBT and MBT<br><br>Duration/Intensity: DBT: 12-month programme; 4 weekly sessions. MBT: 18-month programme; 2 weekly/fortnightly sessions + initial short-term psychoeducation.<br><br>Comparator: Mentalisation-based therapy- 18-month period, weekly or fortnightly individual therapy and weekly group therapy. They also provided a short-term group programme which involves weekly groups delivered over a 10-week period<br><br>Service setting: Specialist PD services | Sample Size: 90.<br><br>Demographics: 72% female; mean age 31.0 (SD=13.0); ethnicities: White 64%, black and minority 36%.<br><br>Diagnoses: DSM-IV BPD diagnosis (SCID-II). | No primary outcome specified. Crisis service use: A&E and psychiatric hospital admissions; self-harm (SASII); BPD symptom severity (BEST); emotion regulation (DERS); dissociation (Dissociative Experience Scale); interpersonal problems (SIDES-SR). | No primary outcome specified. Patients receiving DBT were significantly less likely to complete at least 12 months of treatment than those receiving MBT (completion rate 42% vs. 72%), but this was no longer significant after adjusting for baseline differences. At 12 months follow up, groups did not differ in adjusted or unadjusted comparisons of number of incidents of self-harm, BPD severity, emotional dysregulation, relationships with others or dissociation. In unadjusted models, participants receiving DBT reported a significantly steeper decline over time in incidents of self-harm and in emotional dysregulation than participants receiving MBT, remaining significant after adjusting for confounders. |

| | | | | | | |
|---|---|---|---|---|---|---|
| Quasi-experiment with contemporaneous control (matched control design). Specialist/active comparator. | Bales et al. 2015 The Netherlands | To make a naturalistic comparison between the benefits of day hospital MBT and a variety of forms of specialist treatment received by matched controls. | Treatment: Day hospital delivering Mentalization-Based Treatment (MBT)<br><br>Duration/Intensity: 18-month initial day hospital programme; 5 days a week (270 minutes). Followed by 18-month group therapy.<br><br>Comparator: Comparison group received a variety of other types of inpatient, day patient and outpatient specialist treatment for personality disorder as available in the Netherlands, with wide variations in length and intensity.<br><br>Service setting: Specialist Day hospital setting for experimental group; variety of settings for control group | Sample Size: 204.<br><br>Demographics: 69% female MBT, 82% female OPT, 86% female OPT; mean age 30.0 (SD= 6.17) MBT, 30.3 (SD=7.76) OPT, 30.4 (SD=7.93) OPT; no ethnicity data provided.<br><br>Diagnoses: DSM-IV BPD diagnosis (SCID-II; SIDP-IV). | No primary outcome specified. Symptom severity (BSI); personality functioning (SIPP-118). | No primary outcome specified. Participants in both conditions improved at 36 months on all outcome indices. Statistically significant differences favouring the MBT group were reported at 18 months and 36 months for improvement in psychiatric symptoms and in domains of personality functioning, but not in relational functioning. |
| Natural experiment with pre-post comparison. Specialist/active comparator. | Jones et al. 2013a UK | To investigate effects on bed use of over 2 years three models of specialist care for personality disorder incorporating MBT and therapeutic principles and an open access group programme. | Treatment: Same as Jones et al (2012)<br><br>Duration/Intensity: 3-day MBT programme: weekly individual sessions 2-day MBT programme: fortnightly individual sessions SUN programme: open-access to 4 groups per week.<br><br>Comparator: N/A<br><br>Service setting: Specialist personality disorder service | Sample Size: 72.<br><br>Demographics: 75% female; mean age 39 (SD=12.30); ethnicities White 86.11%, Black 1.39%, Mixed race 4.17%, Other 8.33%.<br><br>Diagnoses: PD diagnosis (SCID-II/SAPAS). | Primary outcome: Bed use (clinical records). | The authors reported that numbers in each component treatment group were too small at this stage for meaningful comparisons. Overall evaluation of the service found significant reductions in bed use at 18-months (p<.001, effect size=.067) and 24-months (p<.001, effect size=.068) after starting treatment. |
| Natural experiment with pre-post comparison. Specialist/active comparator. | Jones et al. 2013b UK | To investigate effects on clinical outcomes of three models of specialist care for personality disorder incorporating MBT and therapeutic principles and an open access group programme. | Treatment: Same as Jones et al (2012)<br><br>Duration/Intensity: 3-day MBT programme: weekly individual sessions 2-day MBT programme: fortnightly individual sessions SUN programme: open-access to 4 groups per week.<br><br>Comparator: N/A<br><br>Service setting: Specialist personality disorder service | Sample Size: 72.<br><br>Demographics: 75% female; mean age 39 (SD=12.30); ethnicities White 86.11%, Black 1.39%, Mixed race 4.17%, Other 8.33%.<br><br>Diagnoses: PD diagnosis (SCID-II/SAPAS). | No primary outcome specified. Depression (BDI-II; PHQ-9); anxiety (STAI; GAD-7); social adjustment (SAS-SR); interpersonal problems (IIP), self-esteem (Rosenberg self-esteem scale); QoL (EQ-5D); global health functioning (HoNOS); global functioning (GAF); satisfaction with treatment (CSQ). | No primary outcome specified. Data collected mainly related to very small numbers of participants (<15) and focused on change over time during the treatment period. Those who attended the MBT programmes had improved significantly on both the GAF and HoNOS as well in the total scores on the brief symptom inventory (BSI) but gains on most other outcomes did not reach statistical significance for this small group. Good client satisfaction was reported for the SUN project. |

| | | | | | | |
|---|---|---|---|---|---|---|
| Natural experiment with contemporaneous comparisons. Specialist/active comparator. | Jones et al. 2012 UK | To investigate effects on bed use of three models of specialist care for personality disorder incorporating MBT and therapeutic principles and an open access group programme. | Treatment: Personality disorder service that uses two psychoanalytical models: mentalisation-based treatment (MBT) and the service user network (SUN). MBT programmes adopted therapeutic community principles and included individual and group therapy. The SUN project uses therapeutic community principles alongside coping process and psychoanalytical models and includes 4 open access groups per week, from which service users are not discharged. Duration/Intensity: 3-day MBT programme: weekly individual sessions 2-day MBT programme: fortnightly individual sessions SUN programme: open-access to 4 groups per week. Comparator: Comparison between three models of active treatment Service setting: Specialist personality disorder service | Sample Size: 72. Demographics: 75% female; mean age 39 (SD=12.30); ethnicities White 86.11%, Black 1.39%, Mixed race 4.17%, Other 8.33%. Diagnoses: PD diagnosis (SCID-II/SAPAS) | Primary outcome: Bed use (clinical records). | Across the service (MBT programmes and SUN) bed use was significantly reduced from pre-baseline levels both (p<.001, r=-0.415) and 12-months after starting treatment (p=.013, r=-0.293). Comparisons between groups were made more difficult by some patients attending both MBT and SUN programmes, especially at 12 months, but the number of bed days used by SUN attendees 6 months after starting treatment (median 0, IQR=0) did not differ significantly from bed days used by patients in the MBT programmes (median 0, IQR=4, U=248.00, p=.169, r=70.194). There was some significant evidence for lower bed use at 6 months in the MBT 3 day than the MBT 2-day programme, but the reverse was observed at 12 months. |

3. Tests of partial/modified MBT treatments
    a. Randomised Controlled Trials

| | | | | | | |
|---|---|---|---|---|---|---|
| RCT. Modified. | Smits et al. 2020 The Netherlands | To compare the efficacy of MBT-DH and MBT-IOP 18 months after start of treatment. MBT-DH was hypothesised to be superior to MBT-IOP because of its higher treatment intensity. | Treatment: MBT day hospital - MBT focuses on improving capacity for mentalising in patients with BPD. Mentalising is thought to play a key role in affect regulation and interpersonal relationships. Treatment components and features in MBT-DH and MBT-IOP (control) are generally very similar, but the intensity of group therapy differs markedly: MBT-IOP involves two group therapy sessions per week, whereas MBT-DH entails a day hospital programme 5 days per week, with nine group therapy sessions per week.<br><br>Duration/Intensity: Max 18-month programme; 9 group therapy sessions per week across 5 days.<br><br>Comparator: Mentalisation-based treatment in intensive out-patient (MBT-IOP): "MBT-IOP involves two group therapy sessions per week.<br><br>Service setting: Specialist PD services/ Day hospital (compared with intensive standalone outpatient intervention) | Sample Size: 114.<br><br>Demographics: MBT-DH 59/70 female, MBT-IOP 35/44 female; mean age MBT-DH 31.1, MBT-IOP mean age 29.9; no ethnicity data provided.<br><br>Diagnoses: DSM-IV PD diagnosis (SCID-II). | Primary outcome: Symptom severity (BIS GSI). Secondary outcomes: BPD symptom severity (PAI-BOR); personality functioning (SIPP); interpersonal problems (IIP); quality of life (EQ-5D); suicide attempts and self-harm (SSHI). | Primary Outcome: There was no evidence for a differential rate of change between the two groups (β=−0.06; 95% CI −0.19, 0.07; z=−0.88; p= 0.377). The between-group effect size of Cohen's d= 0.34 indicated that MBT-DH was not superior to MBT-IOP in terms of improvements in symptom severity based on the a priori specified Cohen's d ≥0.5 margin. Large improvements were made over time in both groups from start of treatment to 18 months. Significant differences were not found on most other outcome measures. |

4. Psychodynamic therapy treatments vs. Non-active comparators
    a. Randomised Controlled Trials

| | | | | | | |
|---|---|---|---|---|---|---|
| RCT. Non-specialist/inactive comparator. | Reneses et al. 2013 Spain | To test the hypothesis that combined treatment with Psychic Representation-focused Psychotherapy plus CT (Conventional Treatment) is more effective than CT in borderline personality disorders in decreasing global severity of the symptoms. | Treatment: Psychic representation focused psychotherapy - a novel time limited manualized psychodynamic psychotherapy. PRFP is based on classical psychoanalytic principals and on characteristics per se of brief psychotherapies. In addition to these principles, PRFP adds work focused on distorted psychic representations and their link with the corresponding affects and emotions.<br><br>Duration/Intensity: 20-week programme; weekly sessions (45 minutes).<br><br>Comparator: TAU - control group only received conventional treatment without additional specialist psychotherapy for six months.<br><br>Service setting: Standalone outpatient intervention | Sample Size: 44.<br><br>Demographics: 70.5% female; age range 18-50; no ethnicity data provided.<br><br>Diagnoses: DSM-IV-TR BPD diagnosis (SCID-II). | Primary outcomes: Symptom severity (SCL-90-R); impulsiveness (BIS); social functioning (SASS). Secondary outcomes: BPD symptoms (ZAN-BPD); symptom severity (CGI); depressive symptoms and suicidal intentionality (MADRS); anxiety symptoms (STAI); self-esteem (SES). | Primary outcomes at the end of treatment for the first 44 participants (the study is published at a point when data has not yet been collected for all participants): There was a substantial decrease in global severity of the symptoms in the experimental group compared to the TAU group when measured with the SCL-90 (p=.016, d= 0.78), as well as in Barratt impulsivity score (p=.009 d=0.61) and Social Adaptation Scale (p=.0001 d=0.80). |
| RCT. Non-specialist/inactive comparator. | Abbass et al. 2008 Canada | To compare Intensive dynamic short-term psychotherapy for PD to treatment as usual. | Treatment: Intensive short-term dynamic psychotherapy (ISTDP) - An intensive emotion-focused psychodynamic therapy with an explicit focus on handling resistance in treatment<br><br>Duration/Intensity: Programme length unclear; weekly sessions (60 minutes). The therapist and patient mutually decided upon termination.<br><br>Comparator: Waitlist. Prior to treatment, the control group received treatment as usual involving monthly meetings with the site coordinator which were designed as supportive psychiatric follow-ups.<br><br>Service setting: Standalone outpatient intervention | Sample Size: 27.<br><br>Demographics: 59% female; between the ages of 18 and 70; no ethnicity data provided.<br><br>Diagnoses: DSM-IV PD diagnosis Borderline (44.4%); obsessive compulsive (37%); avoidant (33.3%) PD. | Primary outcomes: Symptom severity (BSI); interpersonal problems (IIP). Secondary outcomes: Personality status; global functioning (GAF); social and occupational functioning (GAF-SO); employment status; number of working hours per week. | Primary outcomes: Treatment group scores were significantly better than control on the primary outcomes at the end of treatment follow up: treatment group mean 0.51 (SD=0.43) on BSI vs. control group 1.10 (SD=0.69), t=2.71, p=.02; treatment group mean on IIP 0.67 (SD=0.66) vs. 1.11 (SD=0.57) on IIP, t=2.08, p=.048. Better end of treatment scores for experimental than control at p<.05 level of significance also reported for all secondary outcomes (GAF and GAF-SO, employment status, number of working hours per week). In long-term follow-up (average of 2.1 years post treatment), the group as a whole (including treated controls) showed significant improvements in all domains and total number of personality disorder diagnoses reduced from 48 to 8. |

| | | | | | | |
|---|---|---|---|---|---|---|
| RCT. Non-specialist/inactive comparator. | Emmelkamp et al. 2006 The Netherlands | To evaluate the comparative effectiveness of brief dynamic therapy and cognitive–behavioural therapy for patients with avoidant personality disorder as their primary problem. | Treatment: CBT or Brief Dynamic therapy (comparison between these two therapies and waitlist control)<br><br>Duration/Intensity:<br>CBT: 6-month programme; 20 weekly individual sessions (45 minutes).<br>Brief Dynamic therapy: 6-month programme; 20 weekly individual sessions (45 minutes).<br><br>Comparator: Waitlist<br><br>Service setting: Standalone outpatient intervention | Sample Size: 62.<br>Demographics: 32/female; mean age 34.3 (SD=8.9); ethnicity data not provided.<br>Diagnoses: Avoidant PD (SCID–II). | Primary outcomes. PD status (SCID–II); dysfunctional borderline beliefs (PDBQ); anxiety symptoms (LWASQ); social phobia (SPAI). | Primary outcomes: Post treatment, CBT was significantly superior to the control condition on primary outcome measures PDBQ avoidant sub-scale (F(1,52)=7.39, p=.01) and Avoidance Scale (F(1,46)=5.39, p=.02). No significant difference was found between BDT and control. CBT was significantly superior to BDT on all primary outcome measures: PDBQ avoidant sub-scale (F(1,51)=5.92, p=.02), LWASQ (F(1,51)=5.69, p=.02), SPAI social phobia sub-scale (F(1,51)=2.98, P=0.09) and Avoidance Scale (F(1,45)=5.25, p=.03), and on the generalisation measure PDBQ obsessive–compulsive sub-scale (F(1,51)=10.84, p=.002). On none of the measures was BDT superior to CBT. Results were maintained at follow up. |
| RCT. Non-specialist/inactive comparator. | Vinnars et al. 2005 Sweden | To compare manualized support-expressive dynamic psychotherapy with community-delivered non-manualized psychodynamic therapy for outpatients with personality disorders. | Treatment: "Supportive-expressive psychotherapy - comprised 40 weekly sessions and followed Luborsky's treatment manuals and other guidelines for dynamic therapy<br><br>Duration/Intensity: 40-week programme; weekly sessions.<br><br>Comparator: Community-delivered Psychodynamic Therapy - not manualised<br><br>Service setting: Standalone outpatient intervention | Sample Size: 156.<br><br>Demographics: 31.4% male; mean age 35.1 (SD=10.3); no ethnicity data provided.<br><br>Diagnoses: At least one DSM-IV PD diagnosis or a diagnosis of passive-aggressive or depressive PD. Avoidant (34.6%); dependent (9.6%); obsessive-compulsive (18.6%); passive-aggressive (11.5%); depressive (36.5%); paranoid (17.3%); schizoid (4.5%); schizotypal (1.3%); histrionic (1.9%); narcissistic (5.1%); borderline (24.4%); and antisocial (7.7%) PD; PD NOS (16.7%). | No primary outcome specified. DSM-IV PD diagnosis; general functioning (GAF); symptom severity (SCL-90). | No primary outcome specified. No significant differences were found between treatment groups on any outcomes. Large improvements were observed in both groups: at the posttreatment assessment, 38 patients (33.6%) did not fulfil the criteria for a personality disorder diagnosis. At the follow-up assessment, 58 patients (46.8%) did not meet the criteria. |

| | | | | | | |
|---|---|---|---|---|---|---|
| RCT. Non-specialist/inactive comparator. | Winston et al. 1994 USA | To compare the results of two forms of short-term psychotherapy and of a waiting list control condition in patients with personality disorders. | Treatment: Brief adaptive psychotherapy (identification of maladaptive pattern and its elucidation in past and present relationships) / Dynamic psychotherapy -short term (confronting defensive behaviour and eliciting affect in an interpersonal context). Two active treatment and one control condition.<br><br>Duration/Intensity: Average programme length 40 weeks; weekly sessions.<br><br>Comparator: Waitlist (waiting on average 14,9 weeks (SD=6.2))<br><br>Service setting: Standalone outpatient intervention | Sample Size: 81.<br><br>Demographics: 48/81 female; mean age 40.8 (range= 23-61); no ethnicity data provided.<br><br>Diagnoses: DSM-III-R PD diagnosis (SCID-II) other than paranoid, schizoid, schizotypal, narcissistic, and borderline PDs. Cluster C (44%); cluster B (22%); cluster A (4%); PD NOS with cluster C features (23%); PD NOS with cluster B features (1%); PD NOS with cluster C and B features (5%). | No primary outcome specified. Target complaints (PTC); symptom severity (SCL-90-R); social functioning (SAS). | No primary outcome specified. For each outcome at the end of treatment (Target complaints, Global symptom severity and social functioning), improvement was significantly greater in the two treatment groups than in the control group (where there was little improvement). No significant differences were found between the two active treatment conditions at the end of treatment or at follow-up. |
| RCT. Non-specialist/inactive comparator. | Piper et al. 1993 Canada | To compare the effectiveness of psychiatric day treatment with a waiting list control for people with affective or personality disorders causing long term social disruption. | Treatment: Day Treatment Programme in Hospital - no specific guidelines - days start with large psychotherapy group. Throughout the rest of the day therapists lead small groups utilising different techniques such as role-play, televised feedback, peer government, life skills, training in communication, and daily living skills.<br><br>Duration/Intensity: 18-week programme; 5 times a week (7 hours).<br><br>Comparator: Delayed-treatment control - whilst waiting for the treatment participants were invited to attend a weekly supportive outpatient group<br><br>Service setting: Specialist Day service | Sample Size: 120.<br><br>Demographics: 80/120 female; mean age 32.7 (SD=9.1); no ethnicity data provided.<br><br>Diagnoses: Patients with long-term psychiatric difficulties that disrupted familial, social, and work functioning. | No primary outcome specified. Social functioning (SAS); emotional reliance (IDI); interpersonal functioning (SIB); number of and satisfaction with friends (People in Your Life Questionnaire); attachment (AQ); symptom severity (SCL-90); mood level (Mood Survey); life satisfaction (1-item); self-esteem (SES); defensive functioning (DSQ). | No primary outcome specified. Seventeen outcome variables included of which the treatment group had better outcomes on 7 out of the 17 variables: social dysfunction, family dysfunction, interpersonal behaviour, mood level, life satisfaction, self-esteem, and severity of disturbance. Advantages of treatment were maintained at post-treatment and follow-up assessment. |
| **5. Psychodynamic therapy treatments vs. Non-active comparators**<br>b. Non-randomised experiments and observational studies | | | | | | |

| | | | | | | |
|---|---|---|---|---|---|---|
| Natural experiment with contemporaneous comparisons. Non-specialist/inactive comparator. | Chiesa et al. 2020 (sample overlap with Chiesa et al. 2002, 2004, 2006, 2009, 2017) UK | To compare the degree of change in reflective functioning (RF) in a sample of patients diagnosed with PD treated in specialist psychodynamic and non-specialist settings. | Treatment: Mixed residential and community-based step-down psychosocial treatment, and residential-only psychosocial treatment<br><br>Duration/Intensity:<br>Residential treatment: 12-month programme; twice weekly individual therapy + twice weekly group therapy. Step-down programme: 30-month programme; 6 months residential treatment followed by 24 months community based psychosocial treatment.<br><br>Comparator: General psychiatric care in generic mental health services<br><br>Service setting: Specialist PD service compared with generic acute care | Sample Size: 143.<br><br>Demographics: RT-CBP group 81.3% female, RT group 74.4% female, GP 62.5%; RT-CBP mean age 33.19, RT 31.18, GP 34.55; ethnicities White 100%.<br><br>Diagnoses: DSM-IV PD (SCID-II). BPD (65%) and other PD (35%). | Primary outcome: Childhood experiences and caregiver relationship (AAI). Secondary outcomes: Symptom severity (SCL-90-R); social functioning (SAS); general mental health (GAS). | Primary outcome: A significant difference was found between the three treatments in RF level of change between intake and 2-year follow-up, F(2, 8.5)=20.47, p<.001. A large effect size between RT-CBP and GP (g=1.54), a medium to large effect size between RT and GP (g=.81), and a medium effect size between RT-CBP and RT (g=.65) were found: overall RT-CBP was associated with the best outcomes in reflective functioning. Across the three secondary outcome dimensions (SAS, GAS, GSI) at 24 months follow-up, RT-CBP and RT were superior to GP. |
| Natural experiment with pre-post comparison. Non-specialist/inactive comparator. | Kealy et al. 2019 Canada | To investigate outcomes of a psychotherapy evening group programme for people with personality disorder diagnoses or personality dysfunction, investigating factors associated with the alleviation of distress related to participants' main goals for therapy. | Treatment: Psychodynamic group therapy (intensive evening programme)<br><br>Duration/Intensity: 18-week programme; 5 weekly sessions (240 minutes).<br><br>Comparator: None<br><br>Service setting: Standalone outpatient intervention | Sample Size: 81.<br><br>Demographics: 69.1% female; mean age 37.7 (SD=10); no ethnicity data provided.<br><br>Diagnoses: 1) PD diagnosis; or 2) significant traits of personality dysfunction (SCID-II). Avoidant (35.8%); obsessive-compulsive (25.9%); borderline (23.5%); paranoid (9.9%); antisocial (4.9%); schizoid (3.7%); schizotypal (2.5%); histrionic (2.5%); narcissistic (1.2%); dependent (1.2%) PD; PD NOS (1.2%). | No primary outcome specified. Symptom severity (BSI-53; SCL-90-R GSI). Secondary outcomes: life satisfaction; achievement of objectives for treatment; therapeutic alliance; group cohesion (GCQ-S). | Uncontrolled study in which only change over time is measured. No primary outcome specified. Statistically significant changes reported in symptom severity, life satisfaction and the distress associated with the difficulties that participants have identified as their main therapy targets. |

| | | | | | | |
|---|---|---|---|---|---|---|
| Natural experiment with pre-post comparison. Non-specialist/inactive comparator. | Joyce et al. 2017 Canada | To investigate the effectiveness of psychodynamic group therapy for improving interpersonal functioning, the relevance of such change to future social functioning, and the influence of early group processes on this change. | Treatment: Psychodynamically focused treatment - Psychodynamic group psychotherapy with the objective of increasing the individual's personal, social, and emotional well-being with a view to more effective functioning in the community. No individual therapy was offered.<br><br>Duration/Intensity: 18-week programme; 5 weekly sessions (240 minutes).<br><br>Comparator: None<br><br>Service setting: Standalone outpatient intervention | Sample Size: 75.<br><br>Demographics: 70.7% female; mean age 37.6; no ethnicity data provided.<br><br>Diagnoses: DSM-IV PD diagnosis or significant personality dysfunction (SCID-II). Concurrent axis I disorders (93.3%). Avoidant (36.0%); obsessive-compulsive (25.3%); borderline (22.7%); paranoid (10.7%); antisocial (5.3%); schizoid (2.7%); schizotypal (2.7%); histrionic (2.7%); narcissistic (1.3%); dependent (1.3%) PD; PD NOS (1.3%). | Primary outcome: Interpersonal problems (IIP-64). | Uncontrolled study in which only change over time was measured. Primary outcome: There was a moderate improvement to interpersonal problems post-treatment (IIP-64 - $F(2, 72)=77.62$, $p<.01$). This was significantly associated with better social functioning at 6 months follow-up. |
| Observational study with pre post comparison. Non-specialist/inactive comparator. | Kvarstein et al. 2017 Norway | The study aims to investigate longitudinal outcomes of outcome psychodynamic group psychotherapy, including variations associated with gender, age, PD-severity and PD-type. | Treatment: Psychodynamic psychotherapy groups - The psychotherapy groups were established between 2002 and 2004. The approach was modified group analysis (non-manualized). New members were admitted when places were vacant (eight patients per group).<br><br>Duration/Intensity: Mean programme length 18 months; weekly sessions (90 minutes).<br><br>Comparator: N/A - observational design<br><br>Service setting: Standalone outpatient intervention | Sample Size: 103.<br><br>Demographics: 40% male; 51% under 39 years old; ethnicity data not provided.<br><br>Diagnoses: PD diagnosis (SCID-II; MINI). Avoidant (45%); borderline (31%); PD NOS (18%); paranoid (16%); dependent (12%); obsessive-compulsive (10%); antisocial and narcissistic (each 3%); and schizotypal (2%) PD. | Primary outcomes: Symptom severity (SCL-90-R GSI); interpersonal problems (IIP-C). Secondary outcomes: Group climate (GCQ-S); therapeutic alliance (TA); therapy experience (self-report). | Uncontrolled study of change over time during therapy. Primary outcomes: Improvements over the course of group therapy were significant for all outcomes investigated. People with a borderline PD diagnosis had shorter treatment duration, more dropouts, and poorer outcomes than for other personality disorder diagnoses. |

| | | | | | | |
|---|---|---|---|---|---|---|
| Natural experiment with contemporaneous comparison. Non-specialist/inactive comparator. | Gregory et al. 2016 USA | To examine the effectiveness of DDP and DBT in real-world settings. | Treatment: Comparison between Dialectical behaviour therapy (DBT); Dynamic deconstructive psychotherapy (DDP)<br><br>Duration/Intensity:<br>DBT: weekly individual sessions (60 minutes) + weekly group sessions (120 minutes).<br>DDP: 12-month programme; weekly individual sessions.<br><br>Comparator: Treatment as usual (TAU)<br><br>Service setting: Standalone outpatient intervention | Sample Size: 68.<br><br>Demographics: DDP group 85% female, 84% DBT group female, 69% TAU female; mean age 28.0 (SD= 11.7) DDP, 36.6 (SD=10.2) DBT, 29.3 (SD=11.5) TAU; ethnicities Caucasian 89% DDP, 84% DBT, 94% TAU.<br><br>Diagnoses: BPD diagnosis. | Primary outcome: BPD symptoms (BEST). Secondary outcomes: Axis I diagnosis (PDSQ); depressive symptoms (BDI); social and occupational impairment (SDS); suicidal ideation and parasuicidal behaviour (SBQ). | Primary outcome: Attrition from DBT was high and DDP obtained better mean BEST score after 12 months of treatment than DBT (d=0.53, p=.042). DDP performed better than TAU control but DBT did not. Both were associated with significant improvements over time on BEST. Greater improvements were reported for DDP than for DBT for depression, disability, and self-harm, but not suicide attempts. |
| Quasi-experiment with pre- post comparison. Non-specialist/inactive comparator. | Stevenson et al. 2015 Australia | To evaluate the effectiveness of conversational model of therapy in treating patients with treatment resistant depression with comorbid personality disorders and histories of early childhood trauma. | Treatment: Conversational model (CM) of psychodynamic psychotherapy, which has a relational and systematic approach; the "aim of therapy is maturational" with the "generation of the 'Self'" being the central aim.<br><br>Duration/Intensity: 12-month programme; twice weekly sessions (50 minutes). Comparator: N/A<br><br>Service setting: Standalone outpatient intervention | Sample Size: 44.<br><br>Demographics: 70.4% female; age 18-55 years; no ethnicity data provided.<br><br>Diagnoses: Treatment-resistant depression (TRD) with comorbid personality disorders and histories of early childhood trauma: 1) HAM-D and BDI scores >20; and 2) resistance to several pharmacotherapies, therapeutic augmentation and in some cases electroconvulsive therapy (ECT). ≤5 criteria for BPD (63.6%); PDs from 1 to 2 clusters (41%); and PDs from all three clusters (19%). | No primary outcome specified. Depressive symptoms (BDI-II; HAM-D); general functioning (GAF); childhood trauma/history, abuse or neglect (CTQ); self-esteem (SES); BPD diagnosis (DIB-R). | Uncontrolled study in which comparisons are between time points and there are no specified primary outcomes. Significant improvements were observed on all outcome measures in the course of treatment. |

| | | | | | | |
|---|---|---|---|---|---|---|
| Natural experiment with pre-post comparison. Non-specialist/inactive comparator. | Joyce et al. 2013 Canada | To investigate effectiveness of psychodynamic group therapy for interpersonal problems, and to explore defence style as a predictor of this outcome. | Treatment: Psychodynamically focused treatment - Psychodynamic group psychotherapy with the objective of increasing the individual's personal, social, and emotional well-being with a view to more effective functioning in the community. No individual therapy is offered.<br><br>Duration/Intensity: 18-week programme; 5 days a week (7 hours on 4 days and 3.5 hours on one day).<br><br>Comparator: None<br><br>Service setting: Specialist Day service | Sample Size: 32.<br><br>Demographics: 59.4% female; mean age 41.6 (SD=10.8); no ethnicity data provided.<br><br>Diagnoses: Poor interpersonal functioning. PD diagnoses: Borderline (62.5%); narcissistic (37.5%); obsessive-compulsive (15.6%); avoidant (6.3%); dependent (3.1%) PD; and PD NOS (12.5%). | Primary outcome: Interpersonal problems (IIP-C). | Uncontrolled study in which only change over time was measured. Primary outcome: There was a significant improvement to social functioning by treatment end (F(8, 24)=6.93, p<.01) and of medium size (partial eta2=.70). |
| Natural experiment with pre-post comparison, and additional analyses comparing concurrent groups. Non-specialist/inactive comparator. | Berghout et al. 2012 The Netherlands | To investigate changes in general symptoms, depression, anxiety, and interpersonal problems during the first 2 years of long-term psychoanalytic psychotherapy (PP) and psychoanalysis (PA). | Treatment: Long term psychoanalysis (PA) or psychoanalytic psychotherapy (PP)<br><br>Duration/Intensity:<br>PA: Average programme length 3.9 years; 3-5 weekly sessions (60 minutes)<br>PP: Average programme length 6.5 years; 1-2 weekly sessions (60 minutes)<br><br>Comparator: no-intervention (pre-post)<br><br>Service setting: Standalone outpatient intervention | Sample Size: 113.<br><br>Demographics: 71% female; mean age 34 (SD = 8.0); no ethnicity data provided.<br><br>Diagnoses: PD diagnosis (85%): PD NOS (39%); dependent (15%); and avoidant (12%) PD. Mood disorders (50%): Dysthymic (31%) and anxiety disorders (12%). | No primary outcome specified. Symptom severity (SCL-90-R); depressive symptoms (BDI-II); anxiety symptoms (STAI); interpersonal problems (IIP-64). | No primary outcome specified. Most analyses are of change over time, but some comparisons are made between psychoanalysis and psychoanalytic psychotherapy. Groups did not differ during the first 2 years of treatment on any measure except interpersonal functioning intrusiveness sub-scale, where PP showed significantly more improvement than PA participants. In each group, mixed results for change in outcome measures over time, with more change on symptomatic measures than interpersonal and social functioning. |

| | | | | | | |
|---|---|---|---|---|---|---|
| Natural experiment with contemporaneous control (at different stage of treatment pathway). Non-specialist/inactive comparator. | Berghout and Zevalkink 2009 The Netherlands | To investigate the clinical impact of long-term psychoanalytic treatment by comparing symptoms and personality between groups in different phases of treatment (before, during, after, at follow-up). | Treatment: Long term psychoanalytic treatment<br><br>Duration/Intensity: Programmes lasting more than 12 months; 25+ sessions.<br><br>Comparator: Comparisons made between recipients of psychotherapy at different phases of treatment - the pre-treatment cohort (n=64): just started long-term psychoanalytic treatment, during-treatment cohort (n=49): 1 year into treatment; end-of-treatment cohort (n=67): just finished (approximately 3 months after treatment termination) long-term psychoanalytic treatment, follow-up cohort (n=51): already finished their treatment 2 years ago.<br><br>Service setting: Standalone outpatient intervention | Sample Size: 231.<br><br>Demographics: 73% female; mean age 36 (SD=8.4); no ethnicity data provided.<br><br>Diagnoses: PD diagnosis (73%). | No primary outcome specified. Symptom severity (SCL-90); depressive symptoms (BDI-II); anxiety symptoms (STAI); interpersonal problems (IIP-64); personality functioning (MMPI-2, Rorschach-CS). | No primary outcome specified. Significantly lower numbers of clinical cases, and lower symptom and higher social functioning scores following than prior to treatment. |
| Quasi-experiment with pre-post comparison (comparison with an inpatient programme also reported). Non-specialist/inactive comparator. | Chiesa et al. 2009 (sample overlap with Chiesa et al. 2002, 2004, 2006,2017, 2020) UK | To describe key features of a community-based psychodynamic programme and its outcomes over a 12-year period. | Treatment: Community based psychodynamic programme<br><br>Duration/Intensity: 12-month programme; twice weekly individual therapy + 5 meetings/week with unit staff + four community meetings/week + one weekly small group psychotherapy + 4 times a week structured programme of activities.<br><br>Comparator: N/A - pre-post<br><br>Service setting: Specialist PD service | Sample Size: 116.<br><br>Demographics: 72% female; mean age 33.9 (SD=9.1); ethnicities 82% Caucasian.<br><br>Diagnoses: DSM-IV PD diagnosis. Borderline (59%); avoidant (47%); dependent (41%); depressive (35%); paranoid (27%); passive-aggressive (24%); histrionic (13%); narcissistic (13%); schizotypal (12%); schizoid (9%); and antisocial (8%) PD. | No primary outcome specified. Symptom severity (BSI); self-mutilation, suicide attempts, and hospital admissions (Cassel Community Adjustment Questionnaire). | No primary outcome specified. The number of patients who self-mutilated, attempted suicide, and were hospitalized at least once before admission to the programme dropped significantly at 12 and 24 months in the community-based treatment sample, and there was also a significant fall in General Severity Index. Findings also suggested better outcomes for the community-based programme than the residential inpatient programme. |

| | | | | | | |
|---|---|---|---|---|---|---|
| Natural experiment with pre-post comparison. Non-specialist/inactive comparator. | Joyce et al. 2009 Canada | To investigate outcomes of an intensive day hospital programme, and to identify predictors of outcome within the sample. | Treatment: Psychodynamically focused treatment group - An insight- oriented, psychodynamic group therapy programme, in which therapist practice is informed by the range of object relations theories.<br><br>Duration/Intensity: 18-week programme; 5 weekly sessions (240 minutes).<br><br>Comparator: None<br><br>Service setting: Specialist Day service | Sample Size: 107.<br><br>Demographics: 63.6% female; mean age 37.4 (SD=9.9); no ethnicity data provided.<br><br>Diagnoses: Problematic personal and interpersonal functioning. Cluster A (14%); cluster B (38.4%); cluster C (50.5%); PD NOS (3.7%); no axis II diagnosis, but traits present (24.3%). | No primary outcome specified. Symptom severity (SCL-90-R GSI); depressive symptoms (BDI-II); anxiety symptoms (STAI); interpersonal problems (IIP); social functioning (SAS-R). | No primary outcomes specified. Results at follow up showed significant improvement in the sample as a whole. , with often large effect sizes across symptom severity and social functioning outcomes. |
| Quasi-experimental design with pre-post comparison. Non-specialist/inactive comparator. | Gerull et al. 2008 Australia | To assess changes in perceived quality of relationships with partners and children of 24 patients diagnosed with Borderline Personality Disorder (BPD) after 12 months of treatment with the Conversational Model (CM). | Treatment: Treatment with the Conversational Model (CM)<br><br>Duration/Intensity: 12-month programme; twice weekly sessions (50 minutes).<br><br>Comparator: Treatment as usual (TAU) (waitlist)<br><br>Service setting: Standalone outpatient intervention | Sample Size: 45.<br><br>Demographics: 17/45 female; CM group mean age 26.9 (SD = 5.4), TAU group 27.7 (SD = 5.1); no ethnicity data provided.<br><br>Diagnoses: BPD diagnosis. | Primary outcome: Social functioning (SAS–SR). | Primary outcome: Significant effects for time x treatment group, suggesting a positive effect from CM were found for relationship with partner ratings on the SAS-SR (Wilks' lambda=.738, $F(1,40)=14.22$, $p=.001$) and for relationships with children (Wilks' Lambda=.823, $F(1,43)=9.26$, $p=.004$). Ratings for overall quality of relationships within the family group did not show a significant effect for the group. |
| Observational study with contemporaneous comparison. Non-specialist/inactive comparator. | Korner et al (sample overlap with Meares and Stevenson 1992). 2008 Australia | To investigate the role of duration on the outcomes of the conversational model. | Treatment: Conversational model - a psychodynamic model focusing on development of self-reflection, and the interplay between the self and the social environment.<br><br>Duration/Intensity: 24-month programme.<br><br>Comparator: 12-months of conversational model treatment and 12-months follow-up.<br><br>Service setting: Specialist PD service | Sample Size: 59.<br><br>Demographics: 100% female; mean age 29.39; no ethnicity data provided.<br><br>Diagnoses: BPD diagnosis. | Primary outcome: Depressive symptoms (Zung Depression scale). Secondary outcome: BPD diagnosis (DIB-R). | Primary outcome: A group which had received two years of therapy was compared with a previous treatment group that had received one year. Evidence was found of a time-group effect on depression score ($F(1,54)=5.55$ $p=.022$)). There was a steady improvement in the two-year treatment group over the entire duration of treatment and a rapid improvement after the first year in the one-year treatment group, but no change in the second year. Secondary outcome: There was also a significant interaction between time and group for DSM BPD criteria ($F(2, 65)=5.548$, $p=.006$), with the two-year group showing improvements over two years and the one-year group only during one year. |

| | | | | | | |
|---|---|---|---|---|---|---|
| Natural experiment with contemporaneous comparisons. Non-specialist/inactive comparator. | Petersen et al. 2008 Denmark | To compare the effectiveness of a specialized short-term psychotherapeutic day treatment programme with a treatment as usual (TAU) for personality-disordered patients on a waiting list in a Danish clinical setting. | Treatment: Specialised Psychotherapeutic Day Treatment Programme: Patients received: 1) twice weekly psychodynamic small-group and large-group therapy; 2) weekly cognitive group therapy, body awareness group therapy, psycho-educational group and music or art group therapy; 3) individual psychotherapy; 4) a key person helping patients meet regularly for therapy, usually contacting the patients by phone when they failed to attend treatment and encouraging the patients to meet with community workers; 5) when needed patients received a medication review by the consulting psychiatrist. Upon termination, all patients were encouraged to continue treatment in outpatient group.<br><br>Duration/Intensity: 5-month programme; weekly psychotherapy (60 minutes).<br><br>Comparator: TAU - waitlist (mean 10.5 months) during the waiting time, low intensity contacts average 1 session per month<br><br>Service setting: Specialist Day service for PD | Sample Size: 66.<br><br>Demographics: 86.8% female; mean age 27.4; no ethnicity data provided.<br><br>Diagnoses: PD diagnosis. | No primary outcome specified. DSM-III-R axis II diagnoses (SCID II); ICD-10 axis I diagnoses (PSE); symptom severity (SCL-90-R GSI); personality symptom severity (SCL-90-R PSI); social functioning (CPSAS); interpersonal problems (IIP-C); target complaint (TC); general functioning (GAF); suicidal acts (self-reported). | No primary outcome specified. The day treatment programme showed significantly greater benefits in reducing symptoms of acute illness (hospitalizations in acute ward, psychiatric hospitalisations, and suicide attempts), in stabilizing the psychosocial functioning (GAF, CPSAS) and in reducing complaints that lead to treatment (TC) than the TAU condition. |
| Natural experiment with pre post comparison. Non-specialist/inactive comparator. | Jørgensen and Kjolbye 2007 Denmark | To investigate the effectiveness of a long-term psychoanalytic psychotherapy for BPD. | Treatment: Psychoanalytically oriented psychotherapy<br><br>Duration/Intensity: 2-year programme; 12 months of weekly individual psychotherapy, 22 months weekly group analytic therapy (90 minutes), and 2 months weekly group psychoeducation.<br><br>Comparator: N/A<br><br>Service setting: Standalone outpatient intervention | Sample Size: 19.<br><br>Demographics: 84% female; mean age 28.3 (SD=5.5; range 21-50); ethnicity data not provided.<br><br>Diagnoses: DSM-IV-TR BPD diagnosis. One or more PDs other than BPD (32%). | No primary outcome specified. Symptom severity and general level of functioning (SCL-90-R GSI); depressive symptoms (BDI); anxiety symptoms (BAI) | Uncontrolled design in which only change over time was measured. No primary outcome specified. Statistically significant positive changes were observed in levels of anxiety, depression and general level of functioning/symptom severity over a 15-month treatment period. |

| | | | | | | |
|---|---|---|---|---|---|---|
| Natural experiment with both contemporaneous and historic comparison groups.<br>Non-specialist/inactive comparator. | Korner et al. 2006 Australia | To evaluate outcomes of an outpatient therapy using the conversational model, compared to TAU under naturalistic conditions, and compared with a historic cohort treated in a similar way. | Treatment: Conversational model<br><br>Duration/Intensity: 12-month programme.<br><br>Comparator: TAU waiting list and a historic cohort treated in a similar way<br><br>Service setting: Specialist PD service | Sample Size: 60.<br><br>Demographics: intervention group 17/29 female control group 16/31 female; intervention mean age 27.9 (SD=5.9), control 29.7 years old (SD=6.1); no ethnicity data provided.<br><br>Diagnoses: BPD diagnosis. | No primary outcome specified. BPD symptom severity (DIB-R); global functioning (GAS); self-harm, medical and hospital emergency visits (self-report, friends, relatives, hospital records, health insurance data). | No primary outcome specified. There was a significantly greater reduction in symptom severity and improvement in general functioning in the treatment compared to the TAU group at 12 months. Self-harm episodes and hospital emergency contacts reduced in the treatment group but increased in the TAU group over the study period. There was no reduction in medical contacts. Outcomes for the treatment group are similar to a historic cohort receiving similar care several years before. |
| Natural experiment with pre-post comparison and comparison with a historic cohort of people with BPD diagnosis.<br>Non-specialist/inactive comparator. | Stevenson et al. 2005 Australia | To investigate whether psychotherapy for borderline personality disorder has a lasting effect, focusing on the clinical outcome five years after treatment ended. | Treatment: Psychotherapy based on the "Conversational model" of Hobson, with the emphasis to the restoration of the developmental pathway. A main feature of the therapeutic approach, involving empathic representation, is seen as potentiating the emergence of reflective function.<br><br>Duration/Intensity: 12-month programme; twice weekly psychotherapy (60 minutes).<br><br>Comparator: Treatment as usual. Progress projections were made regarding the course of BPD over time in a cohort of people attending the same clinic.<br><br>Service setting: Hospital setting | Sample Size: 30.<br><br>Demographics: 63.3% female; mean age 34.7; no ethnicity data provided.<br><br>Diagnoses: DSM-III BPD diagnosis. | No primary outcomes specified. DSM-III BPD diagnosis; hospital admissions, time as inpatient; visits to a medical facility each month, drug use self-destructive behaviour and outwardly directed violence, time away from work; symptom severity (Cornell Index). | No primary outcomes specified. Comparisons between the pre-treatment period, post-treatment, 2 years, and 5 years after the beginning of treatment indicated significant reductions on most measures including depression, suicidality, borderline symptoms, self-harm and violence and inpatient and medical service use. 40% of participants no longer met DSM-III criteria for BPD at 5 years. Comparisons are made with the clinical course of a cohort previously assessed in the same clinic to explore whether changes seen in the treatment group would be expected over time without treatment, and the conclusion is drawn that the treatment group have improved much more than the natural course of BPD would suggest. |

| | | | | | | |
|---|---|---|---|---|---|---|
| Natural experiment (pre-post comparison). Non-specialist/inactive comparator. | Wilberg et al. 2003 Norway | To evaluate outcomes from a psychodynamic outpatient therapy programme, delivered following day treatment for patients with personality disorders. | Treatment: Outpatient group therapy, mainly psychodynamic<br><br>Duration/Intensity: 18-week programme; intensive day treatments.<br><br>Comparator: N/A<br><br>Service setting: Standalone outpatient therapy | Sample Size: 187.<br><br>Demographics: 73% female; mean age 34 (SD=8); no ethnicity data provided.<br><br>Diagnoses: PD diagnosis (86%): Avoidant (48%); borderline (28%); dependent (17%); unspecified (17%); paranoid (10%); obsessive-compulsive (9%); narcissistic (2%); histrionic (2%); antisocial (1%); schizotypal (1%) PD. | No primary outcome specified. Measures included Diagnostic Interviews (SCID-II; SCID-I); general functioning (GAF); symptom severity (SCL-90-R GSI); interpersonal problems (IIP-C); benefit from outpatient group psychotherapy (self-report). | No primary outcome specified. Significant changes were made and maintained for GAF, GSI and CIP across the combined day and outpatient treatment programme. Overall, 50 to 78 percent of the total sample were reliably improved on GAF, GSI, and CIP during the combined treatment period, whereas 3 to 11 percent were reliably deteriorated. Reliable change was GAF>5.8, GSI>.29, CIP>.43 for the total sample. Further significant improvements occurred on these measures during the outpatient phase, but these were relatively small in magnitude. |
| Observational study with pre-post comparison. Non-specialist/inactive comparator. | Lorentzen et al. 2002 Norway | To assess effectiveness of long-term, analytic group psychotherapy as it is carried out under "real life" clinical conditions. | Treatment: Long term analytic group psychotherapy - 6-8 people per group. The approach to treatment was group analysis (Foulkes, 1986), which deemphasizes the importance of the therapist and encourages the whole group to be active in the treatment of the individual. It is similar to a psychoanalytic approach with a focus on intrapsychic and inter-personal events.<br><br>Duration/Intensity: Minimum 6-month programme length; weekly group sessions (90 minutes).<br><br>Comparator: N/A - pre-post<br><br>Service setting: Standalone outpatient | Sample Size: 69.<br><br>Demographics: 53.6% female; mean age 36 (range 21-54); ethnicities 90% Caucasian.<br><br>Diagnoses: DSM-III-R axis I diagnosis (97%) and axis II (47%). | No primary outcome specified. General functioning (GAF); interpersonal problems (IIP); symptom severity (SCL-90 GSI). | No primary outcome specified. Significant changes between the beginning and end of therapy were found on each outcome measure. |
| Quasi-experimental with pre-post comparison. Non-specialist/inactive comparator. | Cookson et al. 2001 UK | To examine the effectiveness of the specialist psychotherapeutic treatment of borderline and other severe personality disorders | Treatment: Psychodynamic psychotherapy<br><br>Duration/Intensity: 1 year programme; weekly (or twice weekly for severe cases) sessions (50 minutes).<br><br>Comparator: N/A – pre-post<br><br>Service setting: Standalone outpatient intervention | Sample Size: 43.<br><br>Demographics: 36/43 female; mean age 28 (SD =6.2); no ethnicity data provided.<br><br>Diagnoses: DSM-IV BPD diagnosis (PDQ-4). | No primary outcome specified. PD diagnosis (PDQ-4); symptom severity (BSI); BPD symptoms (BSI); self-harm impulsivity (MIS). | Pilot study with no specified primary outcome. At all time points (at Assessment, 3 Month Follow-up, 13 Month Follow-up, and 20 Month Follow-up) there were significant differences between the scores at the different time points for three of the measures: The Borderline Syndrome Index, the Personality Diagnostic Questionnaire and the Brief Symptom Inventory. There was a large difference from baseline to 3 months in the Multi-Impulsivity Scale, maintained thereafter. Only the three-month differences remain significant after applying the Bonferroni correction for multiple testing. |

| | | | | | | |
|---|---|---|---|---|---|---|
| Observational study with contemporaneous comparisons. Non-specialist/inactive comparator. | Meares et al. 1999 Australia | The aim of this study is to compare the clinical outcome of patients with borderline personality disorder (BPD) who had received outpatient psychotherapy for 1 year with BPD patients who received no formal psychotherapy for the same period. | Treatment: Interpersonal psychodynamic therapy (also known as Conversational Model) - Individual treatment model was consistent with, and an elaboration of the Conversational Model of Hobson. The Conversational Model has been manualised as 'interpersonal-psychodynamic' psychotherapy (IP). The model is based on the idea that borderline personality disorder is a consequence of a disruption in the development of the self.<br><br>Duration/Intensity: 12-month programme; twice weekly individual therapy (60 minutes).<br><br>Comparator: Waitlist comparison. During waiting, they had usual treatments (e.g. psychotherapy, cognitive therapy, pharmacotherapy) - all had been referred by psychiatrists<br><br>Service setting: Standalone outpatient intervention delivered to people referred from generic mental health services | Sample Size: 60.<br><br>Demographics: gender info not reported; mean age treatment 29.4 (SD = 7.9), mean age control 32.9 (SD = 7.8); no ethnicity data provided.<br><br>Diagnoses: DSM-III BPD diagnosis. | Primary outcome: Number of DSM-III BPD criteria. | Primary outcome: After adjusting for DSM at time 0, the DSM scores of individuals in the treatment group decreased by an average of 4.78 more than subjects in the control group (p=.0007), over the 12-month period. 30% of the treatment group, but none of the control group, had ceased to meet the criteria for BPD. |
| Quasi-experiment with pre-post comparison. Non-specialist/inactive comparator. | Barber et al. 1997 USA | To investigate change in the course of supportive-expressive therapy for people with Avoidant Personality Disorder and Obsessive-Compulsive Personality Disorder, examining overall change over time and comparing the two disorders. | Treatment: Supportive-expressive psychotherapy<br><br>Duration/Intensity: 16-month programme; 52 weekly sessions.<br><br>Comparator: N/A - pre-post<br><br>Service setting: Standalone outpatient intervention | Sample Size: 38.<br><br>Demographics: 50% female; mean age 37 (SD = 12.99); no ethnicity data reported.<br><br>Diagnoses: DSM-III-R diagnosis of avoidant (36.2%) and obsessive-compulsive (36.8%) PD. | No primary outcome specified. Depressive symptoms (SIGH-D; BDI); anxiety symptoms (HARS-IG; BAI); therapeutic alliance (CALPAS); global functioning (GAF); personality functioning (WISPI); interpersonal functioning (IIP). | No primary outcome specified. By the end of treatment, 39% of AVPD participants still retained their diagnosis while only 15% of OCPD did so. Both groups improved significantly during treatment on measures of personality disorders, depression, anxiety, general functioning, and interpersonal problems. Therapeutic alliance improved in people with AVPD but not OCPD. |

| | | | | | | |
|---|---|---|---|---|---|---|
| Observational study. Non-specialist/inactive comparator. | Monsen et al. 1995 (same sample as Monsen et al. 1995b) Norway | To examine the long-term outcome of a newly developed intensive psychotherapeutic outpatient programme, 5 years after the end of therapy. | Treatment: Psychodynamic treatment - Object relations theory and psychodynamic self-psychology-based approach: "A model of therapeutic intervention focused on affect which provides an opportunity for greater specification of processes of affective change in psychotherapy.<br><br>Duration/Intensity: Average programme length was 25.4 months (SD=12.9)<br><br>Comparator: N/A<br><br>Service setting: Specialist PD outpatient clinic. | Sample Size: 25.<br><br>Demographics: 76% female; mean age 28.6 (SD=7.4); no ethnicity data provided.<br><br>Diagnoses: Severe mental illness within the range of PDs and psychoses. DSM-III PD diagnosis (92%); axis I diagnosis (96%). | No primary outcome specified. Affect consciousness (semi-structured interview constructed for this study); mental health diagnoses (SCID-I and SCID-II; MMPI); symptom severity and psychosocial outcomes (HSRS; SCL-90-R); global functioning (GAF). | Uncontrolled study in which only change over time is measured, no specified primary outcome. At termination of therapy, statistically significant improvements were found in symptom severity (DSM-III; MMPI), affect consciousness, and capacity for relationships. The reduction in axis II diagnoses was 72%. These patterns were reported to be generally stable from the end of treatment to 5-year follow-up. |
| Natural experiment with pre post comparison. Non-specialist/inactive comparator. | Monsen et al. 1995b (same sample as Monsen et al. 1995) Norway | To examine the long-term functional outcomes of a newly developed intensive psychotherapeutic outpatient programme, 5 years after the end of therapy. | Treatment: Psychodynamic treatment. Object relations theory and psychodynamic self-psychology-based approach: "A model of therapeutic intervention focused on affect which provides an opportunity for greater specification of processes of affective change in psychotherapy.<br><br>Duration/Intensity: Average programme length was 25.4 months (SD=12.9)<br><br>Comparator: N/A<br><br>Service setting: Standalone outpatient intervention | Sample Size: 25.<br><br>Demographics: 76% female, mean age 28.6 (SD=7.4, range 20-58), no ethnicity data provided.<br><br>Diagnoses: Axis II PD (92%); and axis I diagnosis (96%). | No primary outcome specified. Social adjustment: social network, education, occupation, income, housing conditions, use of health and social services (questionnaire, unspecified); capacity for intimacy (semi-structured interview constructed for this study); neurotic discomfort and identity diffusion (MMPI). | Uncontrolled design in which only change over time was measured. No primary outcome specified. There were significant improvements in markers of personality disturbances and symptoms over the full study period, from start of therapy to 5-year follow-up. Improvements were described on multiple measures of social adjustment including education and self-support, complexity of work, monthly income, and housing conditions, as well as health and social service use during the study period. Participants were more likely to be married/cohabiting rather than single, and closeness of contact with friends and capacity for intimacy were significantly improved, but frequency of contact and relationship with family did not improve. |

| | | | | | | |
|---|---|---|---|---|---|---|
| Natural experiment with pre-post comparison. Non-specialist/inactive comparator. | Karterud et al. 1992 Norway | To investigate if a mixed sample of people with personality disorder diagnoses respond to psychodynamic day hospital treatment. | Treatment: Psychodynamic orientated day hospital treatment - Group psychotherapy was conducted by two stable co-therapists. Half of the patients attended the art therapy group. The other half attended the body awareness group.<br><br>Duration/Intensity: Programme length unclear; 3 times a week group therapy (60 minutes) + weekly individual therapy (60-120 minutes) + weekly occupational meetings (60-180 minutes).<br><br>Comparator: None<br><br>Service setting: Specialist Day hospital | Sample Size: 97.<br><br>Demographics: 71% female; mean age 35.7 (S =9.5); no ethnicity data provided.<br><br>Diagnoses: DSM-III-R PD diagnosis (76.3%): schizotypal (13.4%); borderline (35.1%); cluster C only (18.6%); mixed (6.2%); schizoid (2.1%); and narcissistic (0.1%) PD. | No primary outcome specified. Treatment milieu (WAS); symptom severity (SCL-90-R GSI); overall mental health (HSR); medication use. | Uncontrolled study in which only change over time was measured. No primary outcome specified. Two patients made suicidal attempts during treatment. The level of medication was moderate, and 58% of the patients were drug-free at discharge. Treatment results at discharge, measured by SCL-90 and Health Sickness Rating Scale, varied by diagnostic group, but all groups showed substantial improvements. |
| Quasi-experiment with pre-post comparison. Non-specialist/inactive comparator. | Meares and Stevenson. 1992 Australia | To evaluate the effectiveness of engaging outpatients with BPD in a programme of psychotherapy. | Treatment: Individual outpatient psychotherapy "based on psychology of self", "based on the notion that borderline personality disorder is a consequence of a disruption in the development of the self". The aim is maturational.<br><br>Duration/Intensity: 12-month programme; twice weekly sessions.<br><br>Comparator: N/A<br><br>Service setting: Standalone outpatient intervention (with back-up from inpatient service if needed) | Sample Size: 48.<br><br>Demographics: 63.3% female; mean age 29.4 (SD=7.9); no ethnicity data provided.<br><br>Diagnoses: 1) DSM-III BPD diagnosis; and 2) persisting social dysfunction (e.g., unemployment for more than 12 months, absence of severely dysfunctional interpersonal relationships, antisocial behaviour). | No primary outcome specified. DSM-III criteria; self-rated symptoms (Cornell Index); amount of time away from work, service use, drug use, self-destructive behaviour and outwardly directed violence, hospital admissions and time spent as an inpatient (patient, friends or relatives, medical records, referral sources). | Uncontrolled study with comparisons between time points and no specified primary outcome. Significant improvements reported on all behavioural measures, including self-harm, violence, drug use, employment, and service use in the one year after treatment compared with the one year before. 40% no longer met criteria for BPD at follow-up, and significant drops were also seen on symptom ratings. Early study conducted in the context of a wide-spread view that BPD could not be treated. |

5. Psychodynamic therapy treatments vs. Non-active comparators

c. Uncontrolled intervention development studies

| | | | | | | |
|---|---|---|---|---|---|---|
| Intervention development/uncontrolled preliminary testing. Non-specialist/inactive comparator. | Clarkin et al. 2001 USA | To conduct an initial feasibility study, preparing to examine the effectiveness of transference-focused psychotherapy for BPD | Treatment: Transference Focused Psychotherapy<br><br>Duration/Intensity: 12-month programme.<br><br>Comparator: N/A – pre-post<br><br>Service setting: Standalone outpatient intervention | Sample Size: 23.<br><br>Demographics: 100% female; mean age 32.7 (SD=7.52); ethnicities 76.5% Caucasian, 23.5% Hispanic.<br><br>Diagnoses: 1) Five or more DSM-IV BPD criteria (SCID-II); and 2) at least two incidents of suicidal or self-injurious behaviour. Comorbid axis II disorders: Narcissistic (82%), paranoid (76%), obsessive compulsive (71%), and avoidant personality disorder (65%) PD. | No primary outcome specified. Suicidal and parasuicidal behaviour (PHI); ER visits, number, and length of psychiatric hospitalisations (THI); global functioning (GAF). | Uncontrolled feasibility study with no clear primary outcome and not based on a power calculation. Mixture of outcomes reported, suggesting statistically significant improvements over time on some variables including parasuicide and service utilisation. |
| 6. Psychodynamic therapy treatments vs. Specialist comparators<br>a. Randomised Controlled Trials | | | | | | |
| RCT. Specialist/active comparator. | Berthoud et al. 2017 Switzerland | To examine the impact of adding MOTR to a 10 session General Psychiatric Management intervention. | Treatment: General psychiatric management with added use of motive-oriented therapeutic relationship (MOTR) intervention<br><br>Duration/Intensity: 10-week programme; weekly sessions.<br><br>Comparator: Manual-based psychiatric-psychodynamic 10-session version of general psychiatric management (GPM), a borderline-specific treatment, 1 per week<br><br>Service setting: Standalone outpatient intervention | Sample Size: 50.<br><br>Demographics: GPM group 88% female, MOTR group 68% female; mean age GPM 31.04 (SD= 9.79), MOTR 32.20 (SD= 8.96); no ethnicity data provided.<br><br>Diagnoses: DSM-III BPD (SCID-II). | No primary outcome specified. Psychosocial functioning, including symptom distress, interpersonal relations, and social role functioning (OQ-45); BPD symptoms (BSL-23). | No primary outcome specified. Addition of MOTR (experimental group) was associated with less symptom distress, but no significant differences in interpersonal relations, social functioning, or borderline symptoms. An increase in emotional variability was also observed in the experimental group. |

| | | | | | | |
|---|---|---|---|---|---|---|
| RCT. Specialist/active comparator. | Kramer et al. 2017 Switzerland | To investigate outcomes at a follow-up point of a previously reported randomised controlled trial which compared 10-session GPM plus MOTR to GPM-only (Kramer et al., 2014). | Treatment: Motive orientated therapeutic relationship (MOTR) plus general psychiatric management (GPM). The MOTR method includes a set of therapeutic relationship heuristics and intervention strategies.<br><br>Duration/Intensity: 3-month programme; 10 sessions.<br><br>Comparator: General psychiatric management (GPM)<br><br>Service setting: Standalone outpatient intervention | Sample Size: 99.<br><br>Demographics: 69% female; mean age 32.2 years (SD=10.6); ethnicities 85% Caucasian.<br><br>Diagnoses: DSM-IV BPD diagnosis. | Primary outcome: Progress in psychotherapy, including symptomatic level, interpersonal relationships, social role subscales (OQ-45). Secondary outcomes: Number of psychiatric inpatient hospitalisations, number of visits at emergency services (unclear). | Primary outcome: 40 patients with available OQ-45 data showed significant improvement at the 3-6-months follow-up compared to intake (F(1,39)=12.06, p<.001) as well as sustained treatment effects from discharge to follow-up (F(1, 39)=0.90, p<.35). This did not differ between treatment conditions (F(1, 39)=1.07, p=.31). Secondary outcome: The total number of days in inpatient treatment and total number of crisis consultations during the 12-month follow-up did not differ between the two groups. The MOTOR group was more likely to engage in outpatient psychotherapy during follow-up (x2(1)=5.25, p=.02). |
| RCT. Specialist/active comparator. | Kramer et al. 2014 Switzerland | To investigate the effect of motive orientated therapeutic relationship added to General Psychiatric Management on symptoms and patient-therapist collaboration for people with a diagnosis of BPD. | Treatment: Motive orientated therapeutic relationship (MOTR) plus general psychiatric management (GPM). The MOTR method includes a set of therapeutic relationship heuristics and intervention strategies.<br><br>Duration/Intensity: 3-month programme; 10 sessions.<br><br>Comparator: General psychiatric management (GPM)<br><br>Service setting: Standalone outpatient intervention | Sample Size: 85.<br><br>Demographics: 51/74 female; mean age 32.1 (range 18-65); no ethnicity data reported.<br><br>Diagnoses: DSM-IV BPD diagnosis. | Primary outcome: Progress in psychotherapy, including symptomatic level, interpersonal relationships, social role subscales (OQ-45). Secondary outcomes: Interpersonal functioning (IIP); BPD symptoms (BSL-23); therapeutic alliance (WAI-short version). | Primary outcome: There was a main between-group effect (condition × time) on the total score on the OQ-45 (F(1, 73)=7.25, p<.02), suggesting a positive impact of treatment on symptoms for add-on MOTR. Time effect ANOVAs demonstrated that participants in both groups improved on all outcomes from start to end of treatment. There were no significant group differences in interpersonal problems (IIP) and BPD symptom severity (BSL-23). |
| RCT (pilot). Specialist/active comparator. | Kramer et al. 2011 Switzerland | To investigate the effects of motive-oriented therapeutic relationship (MOTR) compared to TAU in early-phase treatment with people with a BPD diagnosis on outcome, therapeutic alliance, and session impact. | Treatment: Motive-orientated therapeutic relationship and plan analysis plus TAU<br><br>Duration/Intensity: 10 sessions<br><br>Comparator: General psychiatric management (GPM)<br><br>Service setting: Standalone outpatient intervention | Sample Size: 25.<br><br>Demographics: 77% female; mean age 30.72 (SD=10.59; 19–55); ethnicity data not provided.<br><br>Diagnoses: Main BPD diagnosis. | Primary outcome: Psychosocial outcome, including symptomatic, level, interpersonal relationships, and social role (OQ-45). Secondary outcomes: Therapeutic alliance (WAI-short version); therapeutic impact of one session (BPSR-P). | Primary outcome: There was no between-group difference for overall therapeutic outcome (OQ-45: F(1, 23)=1.28; p=.21), but the MOTR group showed significantly greater improvement on the interpersonal problems subscale compared to the TAU group (OQ-45: F(1, 23)=4.53, p<.05). Secondary outcomes: Patient's ratings of therapeutic alliance were significantly more improved in the MOTR group (WAI). There was no between-group difference for therapist's ratings. No between-group difference was found for patient's overall session experience (BPSR-P). |

| | | | | | | |
|---|---|---|---|---|---|---|
| RCT. Specialist/active comparator. | Kallestad et al. 2010 (same sample as Svartberg et al. 2004) Norway | To investigate long-term effectiveness of short-term dynamic psychotherapy (STDP) and cognitive therapy (CT) for reducing symptom severity in Cluster C personality disorders. To explore the role of insight in both STDP and CT for Cluster C personality disorders. | Treatment: Dynamic psychotherapy (short term) - McCullough's Short Term Dynamic Psychotherapy model, which is based on Malan's (1979) triangle of conflict.<br><br>Duration/Intensity: 40-week programme; weekly sessions (50 minutes).<br><br>Comparator: Active comparator (Dynamic psychotherapy compared to Cognitive Therapy (CT))<br><br>Service setting: Standalone outpatient intervention | Sample Size: 49.<br><br>Demographics: no demographics provided.<br><br>Diagnoses: DSM-III cluster C PD diagnosis. | Primary outcomes: Symptom severity (SCL-90-R); interpersonal problems (IIP). | Primary outcomes: No statistically significant differences between the two treatment groups for symptom severity or interpersonal problems. No further details given. However, levels of insight increased significantly for those who received STDP but not CT at follow-up. |
| RCT. Specialist/active comparator. | Clarkin et al. 2007 USA | To compare three-year-long outpatient treatments for borderline personality disorder: dialectical behaviour therapy, transference-focused psychotherapy, and a dynamic supportive treatment. | Treatment: Transference focused therapy / dialectical behaviour therapy / Supportive treatment<br><br>Duration/Intensity: Transference focused therapy: 12 months programme; two individual weekly sessions DBT: 12 months program; a weekly individual and group session and available telephone consultation Supportive treatment: 12 months programme; one weekly session with additional sessions as needed.<br><br>Comparator: Active comparators<br><br>Service setting: Standalone outpatient interventions | Sample Size: 90.<br><br>Demographics: 92.2% female; mean age 30.9 (SD=7.85); ethnicities 67.8% Caucasian, 10% African American, 8.9% Hispanic, 5.6% Asian, 7.8% Other.<br><br>Diagnoses: DSM-IV BPD diagnosis (SCID-II). | Primary outcomes: Suicidality (MOAS); aggression (AIAQ); impulsivity (BIS-11). Secondary outcomes: depressive symptoms (BDI); global functioning (GAF); social functioning (SAS). | Primary outcomes: Both transference-focused psychotherapy and dialectical behaviour therapy were significantly associated with improvement in suicidality. Only transference-focused psychotherapy and supportive treatment were associated with improvement in anger. Transference- focused psychotherapy and supportive treatment were each associated with improvement in facets of impulsivity. Regarding secondary outcomes, all treatments were associated with improvements in depression, anxiety, global functioning, and social functioning. Only transference-focused psychotherapy was significantly predictive of change in irritability and verbal and direct assault. The authors suggest transference-focused psychotherapy may result in impacts on a wider range of outcomes than other treatment conditions. |

| | | | | | | |
|---|---|---|---|---|---|---|
| RCT. Specialist/active comparator. | Svartberg et al. 2004 (same sample as Kallestad et al. 2010) Norway, Canada | To compare the effectiveness of short-term dynamic psychotherapy and cognitive therapy for outpatients with cluster C personality disorders. | Treatment: Dynamic psychotherapy (short term) / Cognitive therapy. Short-term dynamic psychotherapy with the overall goal for previously avoided affects, e.g., sadness/grief or tenderness, to be experienced and expressed adaptively by the patient; Cognitive therapy with the goal to help the patient develop new and more adaptive core beliefs and help the patient to develop more adaptive problem-solving interpersonal behaviours<br><br>Duration/Intensity: 40-week programme; weekly sessions (50 minutes).<br><br>Comparator: active comparator (dynamic psychotherapy compared with cognitive therapy)<br><br>Service setting: Standalone outpatient therapy | Sample Size: 50.<br><br>Demographics: 25/50 female; mean age 34; ethnicity 100% Caucasian.<br><br>Diagnoses: DSM-III-R cluster C or self-defeating PD. | No primary outcome specified. Symptom severity (SCL-90-R GSI); interpersonal problems (IIP); personality functioning (MCMI). | No primary outcome specified. No significant difference found on any outcome between dynamic and cognitive therapy groups. Two years after treatment 54% of dynamic therapy and 42% of cognitive therapy patients had recovered symptomatically. |
| RCT. Specialist/active comparator. | Hellerstein et al. 1998 USA | To report preliminary results of BSP for a sample with primarily Cluster C Axis II disorders. | Treatment: Brief supportive psychotherapy (BSP) - emphasis on building self-esteem, reducing anxiety, and enhancing coping mechanisms.<br><br>Duration/Intensity: Programme length unclear; 30-40 sessions.<br><br>Comparator: Active comparator (short-term dynamic psychotherapy; STDP)<br><br>Service setting: Standalone outpatient intervention (both arms) | Sample Size: 49.<br><br>Demographics: 55.1% female; mean age 41.3 (SD=11.1); ethnicities White 91.8%.<br><br>Diagnoses: PD diagnosis (SCID-II) | No primary outcome specified. Symptom severity (SCL-90-R GSI), interpersonal problems (IIP); major presenting problems (PTC). | No primary outcomes specified. No statistically significant difference between groups on any of outcomes (significant improvements over time in both groups on most outcomes). |

#### 6. Psychodynamic therapy treatments vs. Specialist comparators

##### b. Non-randomised experiments and observational studies

| | | | | | | |
|---|---|---|---|---|---|---|
| Pre-post - secondary analysis of two randomized controlled studies. Specialist/active comparator. | Penzenstadler et al. 2018 Switzerland | The aim of the study is to compare the impact of a 10-session version of a GPM treatment on patients with BPD and patients with BPD and a co-morbid SUD concerning treatment process and outcome. | Treatment: General Psychiatric Management (GPM) for patients with BPD – short-term treatment programme based on the principles of good psychiatric management for BPD. Treatment conducted according to GPM treatment manual principles, 1) establishment of a reliable psychiatric diagnoses and communicating that to the patient, 2) synthesis of the psychiatric anamnesis, 3) identification of treatment focus, 4) definition of objectives, 5) working on treatment-interfering problems, and 6) formulation of core conflictual themes.<br><br>Duration/Intensity: 10-week programme, weekly sessions (60 minutes).<br><br>Comparator: General Psychiatric Management (GPM) for patients with BPD and co-morbid substance misuse disorder<br><br>Service setting: Standalone outpatient intervention | Sample Size: 99.<br><br>Demographics: 69% female; mean age 32.2; ethnicities 85% Caucasian.<br><br>Diagnoses: DSM-IV PD diagnosis (SCID-II), with or without comorbid substance abuse disorder (SUD). | No primary outcome specified. General mental health, symptom severity, interpersonal relationships, social role (OQ-45); interpersonal problems (IIP); BPD symptom severity (BSL-23). | No primary outcome specified. Significant reductions in borderline symptoms were found over the course of treatment for patients both with and without comorbid substance misuse disorders, with no significant differences between the groups on this or other outcomes. The authors suggest that General Psychiatric Management may remain effective in the context of substance misuse disorder. |

| | | | | | | |
|---|---|---|---|---|---|---|
| Natural experimental design with contemporaneous comparisons between different therapeutic approaches. Specialist/active comparator. | Van Manen et al. 2015 The Netherlands | To investigate the "matching hypothesis" that participant who are defined as being "low" on "psychological strengths or ego-adaptive capacities" will experience better outcomes with stabilising psychotherapies and participants who are defined as being "high" on these traits will experience better outcomes with destabilising psychotherapies. | Treatment: Range of therapies, assessing the extent to which they are stabilising (orientated towards acceptance of and support for coping with difficulties faced through experiences of complex emotional needs) or destabilising (change oriented, supporting replacement of maladaptive patterns of emotion, cognition, and behaviour with more adaptive ones, for example through interpretation, confrontation and clarification).<br><br>Duration/Intensity: Destabilising: Average programme length 7.6 months Stabilising: Average programme length 11.7 months<br><br>Comparator: Comparisons are made between therapeutic approaches that are to varying degrees stabilising or destabilising<br><br>Service setting: Inpatient, day patient and outpatient settings | Sample Size: 735.<br><br>Demographics: 69.9% female; mean age 33.7 (SD=9.7); no ethnicity data provided.<br><br>Diagnoses: Primary DSM-IV PD diagnosis. Cluster A (8.2%); cluster B (24.9 %); cluster C (38.9%); and PDNOS (28.0%). | Primary outcomes: Symptom severity (BSI GSI); interpersonal relations and social role functioning subscale (OQ-45). | Predictors of outcomes related to characteristics of therapies and patients are explored. Destabilising treatments are found to be associated with significantly better outcomes, as are great psychological strengths among patients, but an interaction is not found between patient strengths and the extent to which the therapy is destabilising. |

| | | | | | | |
|---|---|---|---|---|---|---|
| Quasi-experiment with contemporaneous comparisons. Specialist/active comparator. | Sachdeva et al. 2013 USA | To use a quasi-randomized design to compare naturalistic twelve-month outcomes of two manual-based treatments for BPD—DBT and DDP—in the real-world setting of a university clinic. | Treatment: Dynamic deconstructive psychotherapy (DDP) - a novel therapy for BPD in which sessions involve elaborating the sequence of recent episodes of interpersonal encounters and maladaptive behaviour (evoking autobiographical memory), identifying, and differentiating specific emotions associated with these episodes, and integrating different ways of making meaning of them<br><br>Duration/Intensity: 12-month programme; weekly individual sessions (60 minutes).<br><br>Comparator: DBT and TAU. DBT - DBT therapists attempt to teach skills such as mindfulness, emotion regulation, and distress tolerance in a skills group and then problem-solve with patients in individual sessions. TAU - unstructured psychotherapy, ranging from cognitive-behavioural to psychodynamic to eclectic.<br><br>Service setting: Standalone outpatient therapy | Sample Size: 71.<br><br>Demographics: DDP group 85% female, DBT group 84% female; DDP mean age 28 (range 18-58), DBT mean age 36.6 (range 18-58); no ethnicity data provided.<br><br>Diagnoses: DSM-IV BPD (SCID-II; IAP). | Primary outcome: PD symptoms (BEST). Secondary outcomes: Depressive symptoms (BDI); social and occupational impairment (SDS); suicidal behaviours (SBQ); DSM-IV PDs (SCID-II); alcohol and drug use (IAP). | Primary outcome: Significantly greater improvement is reported for DDP than DBT (d=0.27, but the control group did significantly worse than either. Clients receiving DDP displayed statistically significant reduction in the number of episodes of self-harm over twelve months, but the other two treatment groups did not. Reduction in self-harm was significantly greater for DDP than for DBT. |
| Natural experiment with contemporaneous controls. Specialist/active comparator. | Wilberg et al. 1998 Norway | The study evaluated the effectiveness of adding a therapy group programme to a period of specialist day service treatment for people with personality disorders. | Treatment: 18 weekday treatment followed by Group psychotherapy (combination of analytical oriented and CBT groups)<br><br>Duration/Intensity: Average programme length 12 months; weekly sessions (90 minutes).<br><br>Comparator: 18 weekday treatment not followed by group psychoanalytic therapy<br><br>Service setting: Specialist Day service (both groups) followed by standalone outpatient intervention (experimental group) | Sample Size: 43.<br><br>Demographics: 77% female; mean age 31(SD=8); no ethnicity data provided.<br><br>Diagnoses: BPD diagnosis. | No primary outcome specified. Psychosocial outcomes (HSRS; SCID-I and -II); employment, social contact, suicide attempts and treatment; symptom severity (SCL-90-R GSI). | No primary outcome specified. The group receiving group therapy following the day hospital group had significantly better global outcome scores on the Health Sickness Rating Scale at treatment discharge and at follow-up than the group not receiving group therapy, and GSI symptom severity scores were also significantly lower at follow-up, but not at treatment discharge, for the group receiving group therapy. |
| 7. Tests of Psychodynamic therapy treatments with different modes of delivery<br>a. Non-randomised experiments and observational studies | | | | | | |

| | | | | | | |
|---|---|---|---|---|---|---|
| Natural experiment with contemporaneous comparisons. Different modes of delivery. | Chiesa et al. 2017 (sample overlap with Chiesa et al. 2002, 2004 2006, 2009, 2020) UK | To evaluate the clinical effectiveness of the three specialist programmes offered to a PD population, including outpatient, residential and stepdown models. | Treatment: Community-based psychosocial treatment and step-down psychosocial treatment (two active treatment groups)<br><br>Duration/Intensity: Community based psychosocial programme: 24-month programme with up to 12-month extension; twice weekly group therapy + twice weekly outreach psychosocial nursing + family and couple therapy as required. Residential treatment: 1-month programme; twice weekly individual therapy + twice weekly group therapy. Step-down programme: 30-month programme; 6 months residential treatment followed by 24 months community based psychosocial treatment.<br><br>Comparator: Active comparator - residential treatment<br><br>Service setting: Specialist PD service | Sample Size: 162.<br><br>Demographics: 77.2% female; mean age 34.1 (SD=8.9); ethnicity data not provided.<br><br>Diagnoses: DSM-IV PD (SCID-II). More than one PD (87%). | No primary outcome specified. Symptom severity (BSI); self-harm and suicide attempts (Cassel Community Adjustment Questionnaire; SSHI). | No primary outcome specified. A significant interaction between treatment model and time was found for psychiatric distress, favouring CBP and RT-CBP compared to RT at 48-month follow-up. CBP and RT-CBP were also found to significantly reduce impulsive behaviour (deliberate self-injury and suicide attempt) compared to RT. Severity of presentation was not found to be a significant predictor of outcome. Long-term RT showed no advantage over long-term CBP, either as stand-alone or as step-down treatment. |

| | | | | | | |
|---|---|---|---|---|---|---|
| Observational study. Different modes of delivery. | Horn et al. 2015 The Netherlands | To investigate the effectiveness of different modalities of psychotherapy in patients with PDNOS, i.e., short-term (up to 6 months) and long-term (more than 6 months) outpatient, day hospital, and inpatient psychotherapy. | Treatment: 6 different treatments: Long term outpatient treatment / Short term outpatient treatment / Long term day hospital / Short term day hospital / Long term inpatient / Short term inpatient (mixed orientation). All treatments varied in theoretical orientations depending on treatment centres, such as psychodynamic (27% of all given treatments), CBT (21% of all given treatments) or an integrative orientation (combining different theoretical frameworks; 52% of all given treatments). Day hospital and inpatient programmes typically consisted of group psychotherapy combined with individual psychotherapy, coaching for social problems, non-verbal or expressive group therapies, discussions about household tasks and living together, community meetings and/or pharmacological treatment.<br><br>Duration/Intensity: Short-term treatments lasted up to 6-months and long-term treatments lasted more than 6-months. Outpatient psychotherapy: individual or group psychotherapy sessions, up to 2-sessions per week. Day hospital psychotherapy: 1-session per week. Inpatient psychotherapy: Patients staying at the institutions for 5-days per week.<br><br>Comparator: Naturalistic comparison between six active treatments<br><br>Service setting: Standalone outpatient interventions | Sample Size: 205.<br><br>Demographics: 72% female; mean age 35.1 (SD=10.3); ethnicity data not provided.<br><br>Diagnoses: PD NOS diagnosis. | Primary outcome: Symptom severity (BSI - Dutch version; GSI). Secondary outcomes: Psychosocial functioning (OQ-45), quality of life (EQ-5D). | Primary outcome: At 60-months after baseline, symptom severity significantly improved across all groups and no significant differences were found between groups with correction for baseline differences. The largest effect was found in short-term day hospital (d=1.42), followed by long-term inpatient (d=1.35), short-term inpatient (d=1.31), long-term day hospital (d=1.17), long-term outpatient (d=1.14), and lastly short-term outpatient (d=0.91). Some differences were found at earlier time points, with the long-term inpatient psychotherapy group performing less well than other modalities at 12 months. Secondary outcomes: psychosocial functioning and quality of life also improved at 60-months in all groups (except for QoL in short-term day hospital and psychosocial functioning in short-term outpatient and short-term day hospital). |

| | | | | | | |
|---|---|---|---|---|---|---|
| Natural experiment with contemporaneous comparisons. Different modes of delivery. | Bartak et al. 2011 The Netherlands | To compare the effectiveness of different psychotherapeutic settings for patients with cluster B personality disorders, including outpatient, day patient and inpatient models of treatment. | Treatment: Day hospital (orientation not specified) / Outpatient / Inpatient<br><br>Duration/Intensity: Outpatient treatment group: mean programme length 14.5 months; 2 weekly sessions Day hospital treatment group: at least one morning/afternoon a week Inpatient treatment group: mean programme length 9.1 months; stayed at institutions 5 days a week<br><br>Comparator: Three different modalities of treatment (inpatient, day service and outpatient) were compared with one another<br><br>Service setting: Mental health centres offering a range of treatment modalities | Sample Size: 207.<br><br>Demographics: 71% female; mean age 31.3 (SD 8.5); no ethnicity data provided.<br><br>Diagnoses: Significant DSM-IV personality pathology (SCID-II Dutch version). Borderline PD (77.3%); narcissistic PD (22.7%); histrionic PD (12.6%); and antisocial PD (8.7%). | Primary outcome: general psychiatric symptomatology (BSI - Dutch version). Secondary outcomes: Psychosocial functioning (OQ-45: interpersonal relations and social role functioning subscales); health-related quality of life (EQ-5D). | In the 18 m after baseline, patients in all settings made large and statistically significant improvements on symptom severity, the primary outcome measure, with no statistically significant differences between settings (outpatient vs. day beta=0.11, 95% CI: -0.17, 0.40; p=.44; outpatient vs. inpatient beta=0.30; 95% CI: -0.01, 0.60; p=.059; day vs. inpatient p=.018; 95% CI: -0.06, 0.42; p=.14). Similarly for other measures, all groups in all settings improved but none was significantly better than the others. |
| Natural experiment with contemporaneous comparisons. Different modes of delivery. | Bartak et al. 2010 The Netherlands | To compare the effectiveness of 5 treatment modalities for patients with cluster C personality disorders in terms of psychiatric symptoms, psychosocial functioning, and quality of life. | Treatment: Day hospital (orientation not specified): 5 treatment modalities- Long-term outpatient; short-term day hospital; long-term day hospital; short-term inpatient; long-term inpatient<br><br>Duration/Intensity:<br>Outpatient treatment group: 6+ month programme; 2 weekly sessions Short-term day hospital treatment group: up to 6-month programme; at least one morning/afternoon a week Long-term day hospital treatment group: 6+ month programme; at least one morning/afternoon a week.<br>Short-term inpatient treatment group: Up to 6-month programme; stayed at institutions 5 days a week<br>Long-term inpatient treatment group: 6+ month programme; stayed at institutions 5 days a week.<br><br>Comparator: 5 different modalities of psychotherapeutic treatment in different settings and over different durations<br><br>Service setting: Mental health centres offering a range of treatment modalities | Sample Size: 371.<br><br>Demographics: 70.4% female; mean age 33.5 (SD 9.5); no ethnicity data provided.<br><br>Diagnoses: Significant DSM-IV personality pathology (SCID-II Dutch version). Cluster C PD, with no co-morbid cluster A or B PD (66.6%); combination of cluster C PD and cluster B PD (23.7%); combination of cluster C PD and cluster A PD (4%); combination of cluster C PD and both cluster A and B PD (5.7%); avoidant PD (63.3%); obsessive-compulsive PD (49.3%); dependent PD (22.6%) | Primary outcome: General psychiatric symptomatology (BSI - Dutch version). Secondary outcomes: Psychosocial functioning (OQ-45: interpersonal relations and social role functioning subscales); health-related quality of life (EQ-5D). | In the 18 m after baseline, patients in all settings made large and statistically significant improvements on symptom severity, the primary outcome measure. In a multi-level modelling analysis with propensity score adjustment, a short-term inpatient group improved more than long-term outpatient, short- and long-term-day patient and long-term inpatient treatment modalities, a difference reaching statistical significance in the model (beta=0.38, p=.0059, 95% CI 0.11, 0.6). Similar differential improvements were observed for quality of life, social role functioning and interpersonal relationships. Other modalities did not differ. |

| | | | | | | |
|---|---|---|---|---|---|---|
| Natural experiment with contemporaneous comparison. Different modes of delivery. | Chiesa et al. 2004 (sample overlap with Chiesa et al. 2002, 2006, 2009, 2017, 2020) UK | To compare a step-down model with brief inpatient treatment followed by a specialist outpatient programme with a specialist inpatient programme and generic community mental health service care. | Treatment: Step-down psychosocial treatment involving shorter-term inpatient stay followed by longer-term outpatient and community treatment<br><br>Duration/Intensity: Inpatient programme: 12-month programme; twice weekly individual psychotherapy + 5 meetings/week with unit staff + twice weekly community meetings + weekly small group psychotherapy + structured programme of activities + psychotropic medication. Step down programme: 6-month admission (as above) followed by 12-18 months outpatient therapy + outreach nursing; twice weekly small group analytic psychotherapy + twice weekly individual and group meetings in the community + active networking with care workers.<br><br>Comparator: Active comparator (long-term residential psychosocial/psychoanalytic treatment) & treatment as usual (general psychiatric comparison)<br><br>Service setting: Specialist PD service | Sample Size: 143.<br><br>Demographics: Step-down group 77.8% female, long-term inpatient group 77.6% female, TAU 65.4% female; mean age step down 32.4, mean age long term inpatient 31.5, mean TAU 34.5; no ethnicity data provided.<br><br>Diagnoses: At least one PD diagnosis. More than two DSM PDs (70%); and concurrent axis I diagnosis (83%). | No primary outcome specified. Symptom severity (SCL-90-R); social functioning (SAS); general functioning (GAS); self-harm, inpatient and outpatient service use (SSHI). | No primary outcome specified. Results are reported for 143 participants. At 24 months follow-up, 24 patients (53%) in the step-down group scored below the cut-off point for symptom severity (the criterion for a clinically relevant change), compared with only seven (14%) and six (12%) in the inpatient and community psychiatric groups, respectively, a statistically significant difference ($F(3, 137) = 23.42$, $p<.0001$). Both symptom severity and number of symptoms reported decreased significantly most sharply in the step-down programme. Patients in the step-down programme also showed better social adaptation and global functioning, and less self-mutilation or suicidal attempts. |

| | | | | | | |
|---|---|---|---|---|---|---|
| Natural experiment with contemporaneous comparison. Different modes of delivery. | Chiesa et al. 2002 (sample overlap with Chiesa et al. 2004, 2006, 2009, 2017, 2020) UK and Norway | To investigate 1) whether different programmes systematically recruit different types of patients, 2) whether there are wide variations in outcome across the different units, and 3) whether there are differences in cost–benefit. | Treatment: Psychoanalytic therapy at two-day hospitals: Psychodynamic group therapy with cognitive behavioural group therapy at Ulleval hospital / Individual and group psychoanalytic therapy at Halliwick hospital.<br><br>Duration/Intensity: Psychoanalytic group therapy with CBT group: 18-week programme; 5 days a week 5 hours of therapy Psychoanalytic individual and group therapy group: 18-month programme; 3 times a week group therapy + one a week individual therapy + twice weekly large group sessions.<br><br>Comparator:<br>Active comparator - In-patient individual psychoanalytic therapy / inpatient intensive socio-therapeutic programme at The Cassel hospital.<br><br>Service setting: Specialist Day hospitals compared with a specialist inpatient service | Sample Size: 243.<br><br>Demographics: Ulleval group 75% female, Halliwick group 59% female, Cassel group 72% female; Ulleval mean age 33 (SD=8), Halliwick 31 (SD=7), Cassel 31 (SD=8); no ethnicity data provided.<br><br>Diagnoses: Severe PD (SCID-II). Schizotypal (14%); paranoid (54%); schizoid (4%); borderline (71%); narcissistic (13%); antisocial (6%); histrionic (8%); avoidant (47%); dependent (39%); obsessive-compulsive (28%); passive-aggressive (25%); and self-defeating (46%) PD. | No primary outcome specified. Symptom severity (SCL-90-R); social functioning (SAS); overall mental health (GAS); global functioning (GAF). | No primary outcome specified. After controlling for admission scores, no significant differences between sites were found either in symptom or social adjustment scores at discharge. No significant differences were found in reliable change in the three sites for symptom severity and social adjustment. However, in-patient treatment at The Cassel had the highest treatment cost. |
| 8. Tests of Psychodynamic Therapy treatments adapted to specific settings/cohorts<br>a. Randomised Controlled Trials | | | | | | |

| | | | | | | |
|---|---|---|---|---|---|---|
| RCT. Adapted for specific group/context. | Gregory et al. 2010 USA | To evaluate whether treatment effects observed in a 12-month randomised controlled trial comparing dynamic deconstructive psychotherapy (DDP) with TAU involving a variety of approaches were sustained over an 18-months follow-up in a group with both a BPD diagnosis and alcohol use problems. | Treatment: Dynamic deconstructive psychotherapy (DDP, psychodynamic psychotherapy) added to TAU and alcohol rehabilitation<br><br>Duration/Intensity: 12–18-month programme; weekly sessions (60 minutes).<br><br>Comparator: Treatment as usual (TAU) including a variety of therapeutic approaches and alcohol rehabilitation<br><br>Service setting: Standalone outpatient intervention (both groups usually participating in alcohol rehabilitation) | Sample Size: 30.<br><br>Demographics: 79% female; mean age 29.2 (SD=8.2); ethnicities 88% White.<br><br>Diagnoses: 1) DSM-IV BPD; and 2) active alcohol abuse or dependence. | Primary outcomes: Clinically meaningful improvement in BPD severity (BEST). Secondary outcomes: depressive symptoms (BDI); dissociative symptoms (DES); social support (SPS). parasuicide behaviour (LPC); Alcohol misuse (ASI); institutional care (THI). | Primary outcomes: Significantly more DDP participants showed clinically meaningful improvement by 12 months (BEST: x2 = 7.73, OR = 16, p=.005), which was maintained during the 18-month follow-up. Secondary outcomes: DDP participants showed significantly greater improvements over the treatment and follow-up period for BPD symptoms and for depression compared with for TAU, and they also showed significantly more improvement in parasuicide behaviours and recreational drug use. There were no between-group differences regarding dissociation, heavy drinking days, perceived social support, or days employed. |
| RCT. Adapted for particular groups/context. | Gregory et al. 2008 USA | To describe the results of a 12-month controlled study that assesses the feasibility, tolerability, and efficacy of a manual-based psychodynamic psychotherapy for persons with co-occurring BPD and alcohol use disorder. | Treatment: Dynamic deconstructive psychotherapy (DDP, psychodynamic psychotherapy) added to TAU and alcohol rehabilitation.<br><br>Duration/Intensity: 12–18-month programme with 1h weekly sessions.<br><br>Comparator: Treatment as usual (TAU) including a variety of therapeutic approaches and alcohol rehabilitation<br><br>Service setting: Standalone outpatient intervention | Sample Size: 30.<br><br>Demographics: 80% female; mean age 28.7 (SD=7.7); ethnicities Caucasian 90%.<br><br>Diagnoses: 1) DSM-IV BPD; and 2) active alcohol abuse or dependence. | Primary outcomes: Parasuicide behaviour (LPC); Alcohol misuse (ASI); Institutional care (THI). Secondary outcomes: Depressive symptoms (BDI); dissociative symptoms (DES); social support (The Social Provisions Scale (SPS)); BPD symptom severity (BEST). | Primary outcomes: DDP participants showed statistically significant improvement in parasuicide behaviour, (ARR=.21; 95% CI .20, .54), alcohol misuse (ARR = .14; 95% CI .25, .49), and institutional care (ARR=.12; 95% CI, .22, .46) compared to TAU. Secondary outcomes: DDP patients also showed improvements in depression (BDI: F(2, 28) = 4.22, p<.05), perceived social support (SPS: F(2, 28) = 4.36, p<.05) and core symptoms of BPD (BEST: F(2, 28) = 4.32, p<.05) compared to TAU and treatment retention was 67% to 73%. The results support the feasibility, tolerability, and efficacy of DDP for the co-occurring subgroup. Such treatment effects were maintained at 30 months follow-up. |
| 8. Tests of Psychodynamic Therapy treatments adapted to specific settings/cohorts<br>b. Non-randomised experiments and observational studies | | | | | | |

| | | | | | | |
|---|---|---|---|---|---|---|
| Quasi-experiment with contemporaneous comparison.<br>Adapted for particular groups/context. | Ridolfi et al. 2019 Italy | To assess the impact of a 6-session psychoeducational group (PEG) intervention for borderline personality disorder (BPD) in an underserved community-based outpatient setting. | Treatment: Intervention group - A psychoeducational programme based on General Psychiatric Management (GPM). Material from the GPM handbook was used to develop the group programme.<br><br>Duration/Intensity: 6-week programme; weekly sessions (90 minutes).<br><br>Comparator: Wait-list control - they participated in PEGs at the study's completion<br><br>Service setting: Standalone outpatient intervention | Sample Size: 96.<br><br>Demographics: 21/48 intervention female; mean age 35; no ethnicity data provided.<br><br>Diagnoses: DSM-IV BPD diagnosis (SCID-II) | No primary outcome specified. PD diagnosis and criteria (SCID-II; MSI-BPD; ZAN-BPD). | No primary outcome specified. Improvements were greater for the treatment group on all types of BPD symptom except for ZAN-BPD impulsivity rating. Benefits remained stable during 2-month follow-up. |

#### Appendix 5 – Other Therapies


1. Mixed therapeutic modalities vs. non-active comparators
    a. Randomised Controlled Trials .......................................................... p. 106
    b. Non-randomised experiments and observational studies .......... p. 108
2. Mixed therapeutic modalities vs. specialist comparators
    a. Randomised Controlled Trials .......................................................... p. 112
3. Other individual therapy vs. non-active comparators
    a. Randomised Controlled Trials .......................................................... p. 112
    b. Non-randomised experiments and observational studies .......... p. 115
4. Other individual therapy vs. active comparators
    a. Randomised Controlled Trials .......................................................... p. 116
5. Social-interpersonal and functional therapies vs. non-active comparators
    a. Randomised Controlled Trials .......................................................... p. 117
6. Social-interpersonal and functional therapies vs. active comparators
    a. Randomised Controlled Trials .......................................................... p. 119
7. Self-management and care planning vs. self-management
    a. Randomised Controlled Trials .......................................................... p. 120
8. Self-management and care planning vs. different ways of organising services
    a. Randomised Controlled Trials .......................................................... p. 121
    b. Non-randomised experiments and observational studies .......... p. 122
9. Novel mental health service model vs day hospital
    a. Randomised Controlled Trials .......................................................... p. 124
10. Novel mental health service model vs established generic or specialist mental health services
    a. Randomised Controlled Trials .......................................................... p. 127
    b. Non-randomised experiments and observational studies .......... p. 129
    c. Uncontrolled intervention development studies ......................... p. 132


| Study design and comparator | Paper | Aim | Treatment details | Sample details | Outcomes | Main findings |
|---|---|---|---|---|---|---|
| 1. Mixed therapeutic modalities vs. Non-active comparators<br>a. Randomised Controlled Trials | | | | | | |

| | | | | | | |
|---|---|---|---|---|---|---|
| RCT. Non-specialist/inactive comparator. | Leppänen et al. 2016 Finland | (1) To create a structured treatment model which is easily applicable to the public community mental health care system in Finland and (2) to evaluate its effectiveness in comparison to treatment as usual | Treatment: A new Community Treatment by Experts model was created (including elements of elements of ST and DBT) - The content of each individual therapy sessions was determined individually between the therapist and the patient. The psycho-educational group's manual was available for the therapists.<br><br>Duration/Intensity: 12-month programme; weekly individual therapy (45-60 minutes) + weekly psychoeducational group sessions (90 minutes).<br><br>Comparator: Treatment as usual (TAU) group to receive treatment that would normally be offered, in accordance with the current treatment practices of Oulu city mental health care services<br><br>Service setting: Generic community mental health services (experimental group receive therapy added to this generic care) | Sample Size: 71.<br><br>Demographics: CTBE group 84.2% female, TAU group 87.5% female; mean age CTBE 31.9 (SD=8.3), TAU 32.3 (SD=8.8); no ethnicity data provided.<br><br>Diagnoses: BPD diagnosis (SCID-II) | Primary outcome: BPD symptoms (BPDSI-IV). Secondary outcome: health-related quality of life (HRQoL). | Primary outcome: After 1 year no significant difference was found between groups in total severity of borderline symptoms (($t(49)=-1.24, p=.220$). Secondary outcomes: improvements were seen on more BPDSI-IV sub-scales for the CTBE group than the control group, and their total quality of life score was also significantly better. |
| RCT. Non-specialist/inactive comparator. | Amianto et al. 2011 Italy | To evaluate the efficacy of adding Sequential Brief Adlerian Psychodynamic Psychotherapy (SB-APP) to Supervised Team Management (STM) in BPD treatment compared to STM alone in a naturalistic group of "heavy" MHS users with BPD. Effectiveness was evaluated 6 times along a two-year follow-up. | Treatment: Psychodynamic psychotherapy added to Supervised Team Management consisting of (a) medication (b) unstructured support focused on relationships and social functioning (c) rehabilitative interventions<br><br>Duration/Intensity: 40-week programme; weekly sessions.<br><br>Comparator: Treatment as usual - Supervised Team Management only<br><br>Service setting: Experimental and control group provided with Supervised Team Management by a multidisciplinary community team who received training in managing borderline personality disorder | Sample Size: 35.<br><br>Demographics: 17/35 female; mean age 40; no ethnicity data provided.<br><br>Diagnoses: DSM-IV-TR BPD diagnosis. | No primary outcome specified. Symptom severity (SCL-90-R; CGI; CGI-M; STAXI); working alliance (WAI-S). | No defined primary outcome. Improvements from baseline reported in both groups with Supervised Team Management at multiple domains and time points. Some evidence reported of greater improvement in experimental group on disturbed relationships, impulsivity, suicidality/self-damaging behaviours, chronic feelings of emptiness, and working alliance, but not on other scales and sub-scales. |

| | | | | | | |
|---|---|---|---|---|---|---|
| RCT. Non-specialist/inactive comparator. | Doering et al. 2010 Austria | To compare transference-focused psychotherapy with treatment as usual by experienced community psychotherapists | Treatment: Transference focused therapy<br><br>Duration/Intensity: 12-month programme; twice weekly sessions (50 minutes).<br><br>Comparator: Treatment as usual delivered by experienced community psychotherapists<br><br>Service setting: Standalone outpatient interventions | Sample Size: 104.<br><br>Demographics: 100% female; mean age 27.46 (SD=6.8); no ethnicity data provided.<br><br>Diagnoses: DSM-IV BPD diagnosis | Primary outcomes: Treatment dropouts, suicide attempts (CISSB, PHI). Secondary outcomes: BPD diagnosis (SCID–I and –II); general functioning (GAF); depressive symptoms (BDI); anxiety symptoms (STAI); symptom severity (BSI); inpatient admission (CRTHI); personality functioning (STIPO). | Primary outcome: Significantly fewer participants dropped out of the transference-focused psychotherapy group (38.5% v. 67.3%; chi squared=8.683, df= 1,p=.003) and also significantly fewer attempted suicide (d = 0.8, P = 0.009). Secondary outcomes: Transference-focused psychotherapy was also significantly superior in the domains of borderline symptomatology, psychosocial functioning. personality organisation, numbers no longer meeting personality disorder criteria, and psychiatric in-patient admissions. Both groups improved significantly in the domains of depression and anxiety and the transference-focused psychotherapy group in general psychopathology, all without significant group differences. Self-harming behaviour did not change in either group. |
| 1. Mixed therapeutic modalities vs. Non-active comparators<br>b. Non-randomised experiments and observational studies | | | | | | |

| | | | | | | |
|---|---|---|---|---|---|---|
| Observational study with pre-post comparison. Non-specialist/inactive comparator. | Savard et al. 2019 Canada | To report on the effectiveness of a time-limited day-hospital crisis treatment for personality disorders (PDs) in a naturalistic setting. | Treatment: Day hospital (orientation mixed: psychodynamic/DBT) - Individual and group therapy, which focus on crisis resolution and rehabilitation. Four thematic groups focusing on resolving crises and interpersonal conflicts, reducing symptoms, and fostering insight are offered in a predetermined sequence. The Monday group emphasizes motivation and stages of change and encourages participants to elaborate specific objectives for the week. The Tuesday group focuses on interpersonal problems using a psychodynamic approach. The Wednesday group is based on a DBT approach and addresses different topics each week (introduction to personality disorders, distress tolerance, managing emotions, problems resolution, defence mechanisms, and cognitive distortions). Finally, the Thursday group, called "expressive group," is an art therapy group following guidelines described by Johns and Karterud. Staff members have 2 meetings per week to discuss new referrals, therapeutic needs, and the clinical evolution of every patient.<br><br>Duration/Intensity: 6-week programme; weekly individual therapy (30 minutes) + group therapy (7 hours a week).<br><br>Comparator: N/A<br><br>Service setting: Specialist Day hospital | Sample Size: 260.<br><br>Demographics: 78% female; age distribution 18-24 years 23.8%, 25-30 years 22.7%, 31-40 years 26.2%, 41-50 17.7%, 51+ years 9.6%; no ethnicity data provided.<br><br>Diagnoses: PD diagnosis. BPD (68.2%); cluster B features (14%); narcissistic (7.4%); mixed (5.8%); dependent and histrionic (both 1.6%); obsessive-compulsive (1.2%); and schizotypal (0.4%) PD. | Primary outcome: Symptom distress, interpersonal relations and social functioning (OQ-45). | Uncontrolled study in which comparisons are between timepoints. Patients significantly improved during treatment on the total OQ-45.2 scale (d=0.78; 95% CI 0.60, 0.98; p<.001) and its 3 subscales. Reliable change was observed for 55% of patients for the total scale. |

| | | | | | | |
|---|---|---|---|---|---|---|
| Observational study with pre-post comparison. Non-specialist/inactive comparator. | Lana et al. 2015 Spain | To assess effectiveness of a 6-month day programme involving a mixture of therapeutic approaches in reducing repeated and/or extended hospitalisations and recurrent Emergency Room visits; and (b) to determine if this benefit is maintained in the mid-long term during the three-year follow-up. | Treatment: Integrated day therapeutic programme (mixed treatment approach). The programme, which takes place from Monday to Friday, comprises several weekly group interventions: (a)skill training group (2.5h), based on dialectic behaviour therapy (DBT); (b) relationship therapy (1.5h), supported by mentalisation-based treatment (MBT); (c) stress management group(2h); (d) psychoeducational group (1.5h); (e) individual therapy once a week, support psychodynamic psychotherapy or DBT, depending on the therapist's approach. Additionally, as frequently as needed by each patient: (f) medication review; (g) nursing consultation; and (h) telephone consultation.<br><br>Duration/Intensity: 6-month programme; weekly group skills training (150 minutes) + weekly relationship therapy (90 minutes) + weekly stress management group (120 minutes) + weekly psychoeducation group (90 minutes) + individual therapy.<br><br>Comparator: N/A pre-post<br><br>Service setting: Specialist Day service | Sample Size: 51.<br><br>Demographics: 61% female; mean age 33. 4 (SD=9.2); ethnicity data not provided.<br><br>Diagnoses: 1) DSM-IV BPD diagnosis; or 2) DSM-IV PD diagnosis with self-harm, suicidal or impulsive behaviour in at least two areas (expenses, sex, substance abuse, careless driving, food binges). BPD (78.4%); PD NOS (5.9%); dependent (3.9%); paranoid (3.9%); schizotypal (3.9%); narcissistic (2.0%); and avoidant (2.0%) PD. | Primary outcome: Number and duration of hospital admissions. | Uncontrolled study in which comparisons are over time. The percentage of patients to hospital over the preceding 6 months significantly decreased from 62.7%, in the 6months prior to the programme start (T0), to 19.6% after 6 months of treatment (T1), and this reduction remained stable 3 years after baseline (B (SE)=-2.20 (0.43), 95% CI −3.04, −1.37, p<.0001). Similarly significant reductions were also found in number of admissions, bed days and use of emergency services. |

| | | | | | | |
|---|---|---|---|---|---|---|
| Natural experiment with pre post comparison. Non-specialist/inactive comparator. | Nysæter et al. 2009 Norway | 1) To assess the long-term effectiveness and the natural course of non-manualised psychotherapy for patients with BPD, and 2) to investigate the relationship between the working alliance, patient and therapist characteristics, and attrition in a naturalistic course of psychotherapy for patients with BPD. | Treatment: Non-manualised psychotherapy - Therapists determine the frames of psychotherapy they wish to implement, i.e., duration, frequency, regularity and type of therapy, which is representative of the organisation of the Scandinavian mental healthcare system.<br><br>Duration/Intensity: Programme length open ended; 1-3 times a week individual sessions (60 minutes).<br><br>Comparator: N/A - pre post study<br><br>Service setting: Standalone outpatient intervention | Sample Size: 32.<br><br>Demographics: 81% female; mean age 28.9 (range 20-43); no ethnicity data provided.<br><br>Diagnoses: DSM-IV BPD diagnosis (SCID-II) | Primary outcome: BPD criteria (SCID-II). Secondary outcomes: Symptom severity (SCL-90-R GSI); interpersonal problems (IIP-64); general functioning (GAF); working alliance (WAI). | Primary Outcome: Intent-to-treat analyses found large effect sizes was found for change in BPD criteria from admission to discharge (d=1.21), and from admission to follow-up (d=1.53). Twenty out of 23 no longer met DSM-IV criteria for BPD at the discharge assessment. Secondary outcomes: There were significant improvements from baseline in all outcomes at discharge and 2-year follow-up. |
| Observational study. Non-specialist/inactive comparator. | Halsteinli et al. 2008 Norway | To explore the relationship between staff related variables and patient outcome in day treatment programmes for patients with PDs. | Treatment: Day treatment programmes: The treatment is based on group therapies consisting of a mixture of psychodynamic and cognitive behavioural groups.<br><br>Duration/Intensity: Programme length unclear; 6.5 to 25.5 hours per week (mean = 15.4, SD = 4.2).<br><br>Comparator: N/A - uncontrolled observational study<br><br>Service setting: Specialist Day treatment centres | Sample Size: 1574.<br><br>Demographics: 73.2% female; mean age 35.1 (SD = 9.1); ethnicity data not provided.<br><br>Diagnoses: Variety of PD diagnoses. Cluster A and B patients (31%). | Primary outcome: Psychosocial functioning (GAF). | Findings were reported on the contribution of staff and service variables to patient outcomes on the GAF, examined through multilevel modelling. 12% of variation in outcomes was at treatment unit level, with a higher proportion of nurses/other college-educated staff associated with better outcome. A small association was found between centres offering more hours of therapy per week and higher GAF, and a university-linked unit achieved better outcomes than others. |

| | | | | | | |
|---|---|---|---|---|---|---|
| Natural experiment with pre-post comparison. Non-specialist/inactive comparator. | Karterud et al. 2003 Norway | To investigate outcomes of time-limited day treatment programmes for patients with personality disorder are effective when implemented on a large scale in routine settings. | Treatment: Day treatment programme (mixed: CBT & psychodynamic) - The treatment programmes are based on group therapies, and they typically consist of a mixture of psychodynamic and cognitive-behavioural groups (Karterud et al., 1998). The treatment follows principles that are considered in contemporary psychiatry as appropriate therapy for patients with PD.<br><br>Duration/Intensity: Programme length unclear; 8-16.5 hours per week follower by post-discharge weekly group sessions for maximum of 3.5 years.<br><br>Comparator: None<br><br>Service setting: Specialist Day hospital | Sample Size: 1244.<br><br>Demographics: 72% female; mean age 34 (SD=9); no ethnicity data provided.<br><br>Diagnoses: At least one DSM-III-R or DSM-IV PD diagnosis (81%): Schizotypal (1.8%); schizoid (0.7%); paranoid (12.5%); antisocial (0.7%); borderline (22.1%); narcissistic (0.9%); histrionic (0.4%); avoidant (20.3%); obsessive-compulsive (3.2%); dependent (3.1%) PD; PD NOS (15.4%). Diagnosis deferred (2.1%); no PD (16.7%). | No primary outcome specified. Global functioning (GAF); symptom severity (SCL-90-R); interpersonal problems (CIP); treatment milieu (WAS); quality of life, work functioning, parasuicidal behaviour (self-report). | Uncontrolled study in which only change over time was measured. No primary outcome specified. Statistically significant improvements were observed by 1 year follow-up for all main outcomes for completers of the programme. 22% of those who started the programme dropped out and showed much smaller improvements than completers. Rates of suicide and self-harm were very low among completers. |
| Natural experiment with pre post comparison. Non-specialist/inactive comparator. | Wilberg et al. 1999 Norway | The study evaluated the outcomes of a specialist day treatment programme for people with personality disorders. | Treatment: 18 weekday hospital treatment, including analytically and cognitively oriented groups<br><br>Duration/Intensity: 18-week programme.<br><br>Comparator: N/A<br><br>Service setting: Specialist Day service | Sample Size: 96.<br><br>Demographics: 76% female; mean age 33 (SD=8); no ethnicity data provided.<br><br>Diagnoses: One or more PD diagnoses (85%). Avoidant (39%); borderline (35%); PD NOS (19%); dependent (16%); and paranoid (8%) PD. Axis I disorder (99%). | No primary outcome specified. DSM-III-R and DSM-IV PD diagnosis (LEAD); general functioning (GAF); DSM-III-R Axis I diagnosis (SCID-I); symptom severity (SCL-90-R GSI); interpersonal problems (IIP-C); symptoms and work functioning (therapist form); suicide (National Death Register). | No primary outcome specified. Uncontrolled study in which comparisons are between time points. GAF, GSI and IIP-C score are reported to have improved significantly during treatment, with a further significant improvement in GAF from end of treatment to one year follow-up and GSI and IIP-C remaining similar. 64% of those who completed day hospital treatment continued to outpatient group therapy. |

2. Mixed therapeutic modalities vs. Specialist comparators

a. Randomised Controlled Trials

| | | | | | | |
|---|---|---|---|---|---|---|
| RCT. Specialist/active comparator. | Kvarstein et al. 2013 Norway | Firstly, to compare costs and clinical gains for PD patients randomly allocated to two different formats of psychotherapy: (1) An intensive, day hospital-based treatment in a step-down format and (2) individual psychotherapy in specialist outpatient practice. Secondly, to specifically investigate the differences associated with two frequent PD subgroups, borderline and avoidant PD. | Treatment: Three phase step-down treatment: (1) Day hospital programme with psychodynamic and CBT group therapies. Followed by (2) outpatient therapy consisting of individual and group therapy, and (3) outpatient group psychotherapy only. All delivered by experienced staff.<br><br>Duration/Intensity: 48-month programme; 18 weeks day hospital followed by 30 months outpatient individual and group psychotherapy followed by 12 months outpatient group psychotherapy only.<br><br>Comparator: Outpatient active comparator: Any therapeutic methods in accordance with each therapist's preferred practice. No limitations on therapy duration, intensity or use of other services. Majority were psychodynamic and psychoanalytic treatment approaches.<br><br>Service setting: Specialist day service compared with standalone outpatient care | Sample Size: 107.<br><br>Demographics: 76% female; mean age 31 (SD = 7); ethnicity data not provided.<br><br>Diagnoses: PD diagnosis with focus on BPD (Intervention: 47%; TAU: 46%) and Avoidant PD (Intervention: 45%; TAU: 35%). Axis II comorbidities: PD NOS, paranoid, obsessive-compulsive, dependent, schizoid, and narcissistic PD. | No primary outcome specified. Global functioning (GAF); health service costs. | No primary outcome specified. The costs of step-down treatment were higher than those of outpatient treatment, but these high costs were compensated by considerably lower costs of other health services. In the sample as a whole, no significant difference was found in cost-effectiveness between stepdown day treatment and outpatient treatment. However, costs and clinical gains depended on the type of PD. For borderline PD patients, cost-effectiveness did not differ by treatment condition. Health service costs declined during the trial and functioning improved to mild impairment levels (GAF > 60). For avoidant PD patients, considerable supplementary health costs were incurred during the intervention, but clinical improvements were superior to the step-down condition. |
| 3. Other individual therapy vs. Non-active comparators<br>a. Randomised Controlled Trials | | | | | | |
| RCT. Non-specialist/inactive comparator. | Haeyen et al. 2018 The Netherlands | To evaluate the effects of an art therapy intervention on psychological functioning of patients with a PD. | Treatment: Art therapy - art sessions and assignments to improve mindfulness, self-validation, emotion regulation skills, interpersonal functioning and insight, and comprehension, with a reflection at the end of each session.<br><br>Duration/Intensity: 10-week programme; weekly sessions (90 minutes).<br><br>Comparator: Waitlist<br><br>Service setting: Standalone outpatient intervention | Sample Size: 74.<br><br>Demographics: 71.1% treatment group female, 69.4% control group female; mean age treatment 36.82 (SD=8.92), control 38.14 (SD=11.97); no ethnicity data provided.<br><br>Diagnoses: Primary diagnosis of at least one PD cluster B and/or C; or a PD not otherwise specified. | No primary outcome specified. Psychological flexibility (AAQ-II); psychosocial functioning (OQ45); cognitive schemas (SMI). | End of treatment, and follow-up (15-weeks from baseline). At end of treatment, there was a significant improvement in psychological flexibility (AAQ-II total score, global subjective mental functioning (OQ45 total score) (Cohen's d=-1.24, 95% CI -1.81, -.68; p<.001), and most of the SMI modes (vulnerable child, angry child, engaged child, impulsive child, compliant surrender, detached protector, self-aggrandizer, punitive parent, demanding parent, happy child, and healthy adult). At follow-up (15-weeks from baseline), effects were reported as maintained for all outcome variables. |

| | | | | | | |
|---|---|---|---|---|---|---|
| RCT. Specialist/active comparator AND non-specialist or TAU or inactive comparator. | Andreoli et al. 2016 Switzerland | (a) To determine whether outpatient psychotherapy targeting to abandonment experiences and fears can reduce suicidality and improve outcome in borderline patients referred to the emergency room with major depressive disorder and self-destructive behaviour severe enough to require medical/ surgical treatment and a brief psychiatric hospitalization (b) to compare delivery of abandonment psychotherapy by specialist psychotherapists and by nurses. | Treatment: Abandonment psychotherapy - a twice weekly cognitive/psychodynamic manualised psychotherapy targeting abandonment fears that may be triggers to crises. Comparison groups were (1) delivery by trained psychotherapists (2) delivery by nurses (3) treatment as usual control.<br><br>Duration/Intensity:<br>Delivery by trained psychotherapists: 3-month programme; twice weekly + intensive treatment as usual.<br>Delivery by nurses: 3-month programme; twice weekly + intensive treatment as usual.<br><br>Comparator: All groups including control received intensive treatment as usual in a psychiatric crisis intervention unit, including intensive nurse visits following crisis, weekly clinical review by psychiatrist, group therapy, social worker support, access to night hospitalisation, 24-hour emergency response and family intervention.<br><br>Service setting: Specialist crisis intervention unit (both groups) | Sample Size: 170.<br><br>Demographics: 84.1% female; mean age 31.9 (SD= 10.1); no ethnicity data provided.<br><br>Diagnoses: 1) DSM-IV MDD and 2) DSM-IV BPD. | Primary outcomes: suicidal relapse and hospitalisation. Secondary outcomes: overall mental health (GAS); symptom severity (CGI); depressive symptoms (HDRS); psychosocial outcomes (HSRS - modified version). | Primary outcomes: Suicidality- participants who received either form of AP had fewer episodes of suicidal relapse (AP-P vs. TAU: Pearson $\chi 2=8.09$, df=1, p=.004; AP-N vs. TAU: Pearson $\chi 2=9.33$, df=1, p=.002). They also had increased survival to suicidal crisis relapse compared to patients assigned to TAU (AP-P vs. TAU log-rank test: Mantel $\chi 2=7.63$, df=1, p=.006; AP-N vs. TAU log-rank test: Mantel $\chi 2=9.87$, df=1, p=.002). Those who received AP were also less likely to be hospitalised as an inpatient than those assigned to TAU (AP-P vs. Tau Pearson $\chi 2=6.34$, df=1, p=.012; AP-N vs. TAU: Pearson $\chi 2=6.34$, df=1, p=.012). Recipients of AP also showed significantly greater improvement on suicidal ideation, global functioning, symptom severity and depression diagnosis. Similar outcomes were reported for the two modes of delivery of AP (nurse and psychotherapists). |

| | | | | | | |
|---|---|---|---|---|---|---|
| RCT. Non-specialist/inactive comparator. | Leirvåg et al. 2010 Norway | To compare outcomes of Body-Awareness Group Therapy with more traditional psychodynamic psychotherapy for people seen as having severe PDs as follow-on treatment from day hospitals. | Treatment: Body awareness group therapy (BAGT)<br><br>Duration/Intensity: 18 weekday treatment programme followed by Body awareness group therapy: 25-month programme; weekly sessions (120 minutes).<br><br>Comparator: Group Psychotherapy (PGT)<br><br>Service setting: Specialist Day services | Sample Size: 50.<br><br>Demographics: 100% female; mean age 35 (SD=6); ethnicity data not provided.<br><br>Diagnoses: Patients treated for severe PD. DSM-III-R axis II diagnoses (SCID-II): Paranoid (BAGT: 10%; PGT: 3%); borderline (BAGT: 19%; PGT: 21%); histrionic (BAGT: 10%; PGT: 10%); avoidant (BAGT: 48%; PGT: 38%); obsessive-compulsive (BAGT: 19%; PGT: 7%); and dependent (BAGT: 14%; PGT: 24%) PD; PD NOS (BAGT: 19%; PGT: 10%); No PD (BAGT: 14%; PGT: 10%). | No primary outcome specified. Global functioning (GAF); symptom severity (SCl-90 GSI); interpersonal problems (CIP); benefits from day treatment and outpatient group therapy (self-report); group climate (GCQ). | No clearly specified primary outcome. The magnitude of improvement change during therapy was significantly greater for global functioning, symptom distress and interpersonal distress for the BAGT group. High ratings were reported for satisfaction with therapy and group climate for the BAGT group. |
| RCT. Non-specialist/inactive comparator. | Winston et al. 1994 USA | To compare the results of two forms of short-term psychotherapy and of a waiting list control condition in patients with personality disorders. | Treatment: Brief adaptive psychotherapy (identification of maladaptive pattern and its elucidation in past and present relationships) / Dynamic psychotherapy - short term (confronting defensive behaviour and eliciting affect in an interpersonal context). Two active treatment and one control condition.<br><br>Duration/Intensity: Average programme length 40 weeks; weekly sessions.<br><br>Comparator: Waitlist (waiting on average 14,9 weeks (SD=6.2))<br><br>Service setting: Standalone outpatient intervention | Sample Size: 81.<br><br>Demographics: 48/81 female; mean age 40.8 (range= 23-61); no ethnicity data provided.<br><br>Diagnoses: DSM-III-R PD diagnosis (SCID-II) other than paranoid, schizoid, schizotypal, narcissistic, and borderline PDs. Cluster C (44%); cluster B (22%); cluster A (4%); PD NOS with cluster C features (23%); PD NOS with cluster B features (1%); PD NOS with cluster C and B features (5%). | No primary outcome specified. Target complaints (PTC); symptom severity (SCL-90-R); social functioning (SAS). | No primary outcome specified. For each outcome at the end of treatment (Target complaints, Global symptom severity and social functioning), improvement was significantly greater in the two treatment groups than in the control group (where there was little improvement). No significant differences were found between the two active treatment conditions at the end of treatment or at follow-up. |

| | | | | | | |
|---|---|---|---|---|---|---|
| RCT (pilot). Non-specialist/inactive comparator. | Zanarini et al. 2008 USA | Whether prompt psychoeducation after diagnosis of BPD leads to a decline in core symptoms and improvement in psychosocial functioning. | Treatment: Psychoeducation group (latest information on the following aspects of BPD: aetiology, phenomenology, co-occurring disorders, treatment options, and longitudinal course). Workshop took place within a week of diagnostic disclosure.<br><br>Duration/Intensity: 1 session workshop.<br><br>Comparator: Wait-list (participated in the psychoeducation workshop at the end of this 12-week study)<br><br>Service setting: Standalone outpatient intervention | Sample Size: 50.<br><br>Demographics: 100% female; mean age 19.3 (SD=1.4); ethnicities 33/50 White.<br><br>Diagnoses: DIB-R and DSM-IV criteria for BPD. | Primary outcomes: BPD symptom severity (ZAN-BPD); social and occupational impairment (SDS). | Primary outcomes: No significant differences were found between immediate and delayed psychoeducation groups in total severity of BPD scores (z-score for group: −0.414, p=0.679), although significant differences were found on two BPD sub-scales. Of note, there was a large decline for the sample as a whole in severity of BPD symptoms in the 12 weeks following diagnosis from a baseline high in the moderate range (score of 10–18) to an endpoint score in the mild range (score of 1–9) of the ZAN-BPD. |
| 3. Other individual therapy vs. Non-active comparators<br>b. Non-randomised experiments and observational studies | | | | | | |
| Quasi-experimental with pre-post comparison. Non-specialist/inactive comparator. | Cameron et al. 2018 USA | To investigate whether a programme designed to improve emotional regulation skills in adults who experienced Adverse Childhood Experiences is associated with improvements in psychological well-being, physical health indices, and quality of life from the programme's onset to after its completion 12 weeks later, and to explore whether faith-based and secular versions have different outcomes. | Treatment: Emotion regulation skills (Adverse Childhood Experiences Overcomers programme); faith based or secular (dependent on personal beliefs and preference). Faith based version had biblical messages and content, secular version quoted from philosophers and scientists.<br><br>Duration/Intensity: 12-week programme; weekly group sessions (120 minutes).<br><br>Comparator: N/A - pre and post<br><br>Service setting: Standalone outpatient treatment - participants recruited through the media | Sample Size: 92.<br><br>Demographics: 70/92 female; mean age 47.06 (SD= 14.52); ethnicities White 59%, Hispanic 36%, Other 5%.<br><br>Diagnoses: Community based sample with adverse childhood experiences (ACEs). Two or more ACEs (85%); 4 or more ACEs (54%); and 7 or more ACEs (33%). | No primary outcome specified. Adverse childhood experiences (Ace Score Calculator); emotional suppressions (CECS); rumination (RRQ); cognitive reappraisal (ERQ); mindfulness (MAAS); resilience (ER 89); self-efficacy (GSE); perceived stress (PSS); mood states (mDES); depressive symptoms (CES-D); quality of life (SF36); symptom load (Health appraisal questionnaire - adapted); sick days. | Uncontrolled study with no specified primary outcome. Significant improvements reported from pre-test to post-test in all facets of emotion regulation, psychological resilience, mental well-being and physical symptoms and illness, and in some facets of quality of life (p< .001). The faith-based and secular versions of the programme yielded comparable improvements in well-being. |
| 4. Other individual therapy vs. Active comparators<br>a. Randomised Controlled Trials | | | | | | |

| | | | | | | |
|---|---|---|---|---|---|---|
| RCT. Specialist/active comparator. | Clarkin et al. 2007 USA | To compare three yearlong outpatient treatments for borderline personality disorder: dialectical behaviour therapy, transference-focused psychotherapy, and a dynamic supportive treatment | Treatment: Transference focused therapy / DBT / Supportive treatment<br><br>Duration/Intensity:<br>Transference focused therapy: 12 months programme; two individual weekly sessions.<br>DBT: 12 months programme; a weekly individual and group session and available telephone consultation.<br>Supportive treatment: 12 months programme; one weekly session with additional sessions as needed.<br><br>Comparator: Compared to each other<br><br>Service setting: Standalone outpatient interventions | Sample Size: 90.<br><br>Demographics: 92.2% female; mean age 30.9 (SD=7.85); ethnicities 67.8% Caucasian, 10% African American, 8.9% Hispanic, 5.6% Asian, 7.8% Other.<br><br>Diagnoses: DSM-IV BPD diagnosis (SCID-II). | Primary outcomes: Suicidality (MOAS); aggression (AIAQ); impulsivity (BIS-11). Secondary outcomes: depressive symptoms (BDI); global functioning (GAF); social functioning (SAS). | Regarding primary outcomes, both transference-focused psychotherapy and dialectical behaviour therapy were significantly associated with improvement in suicidality. Only transference-focused psychotherapy and supportive treatment were associated with improvement in anger. Transference- focused psychotherapy and supportive treatment were each associated with improvement in facets of impulsivity. Regarding secondary outcomes, all treatments were associated with improvements in depression, anxiety, global functioning and social functioning. Only transference-focused psychotherapy was significantly predictive of change in irritability and verbal and direct assault. The authors suggest transference-focused psychotherapy may result in impacts on a wider range of outcomes than other treatment conditions. |
| RCT. Specialist/active comparator AND non-specialist or TAU or inactive comparator. | Andreoli et al. 2016 Switzerland | (a) To determine whether outpatient psychotherapy targeting to abandonment experiences and fears can reduce suicidality and improve outcome in borderline patients referred to the emergency room with major depressive disorder and self-destructive behaviour severe enough to require medical/ surgical treatment and a brief psychiatric hospitalization (b) to compare delivery of abandonment psychotherapy by specialist psychotherapists and by nurses. | Treatment: Abandonment psychotherapy - a twice weekly cognitive/psychodynamic manualised psychotherapy targeting abandonment fears that may be triggers to crises. Comparison groups were (1) delivery by trained psychotherapists (2) delivery by nurses (3) treatment as usual control.<br><br>Duration/Intensity:<br>Delivery by trained psychotherapists: 3-month programme; twice weekly + intensive treatment as usual.<br>Delivery by nurses: 3-month programme; twice weekly + intensive treatment as usual.<br><br>Comparator: All groups including control received intensive treatment as usual in a psychiatric crisis intervention unit, including intensive nurse visits following crisis, weekly clinical review by psychiatrist, group therapy, social worker support, access to night hospitalisation, 24-hour emergency response and family intervention.<br><br>Service setting: Specialist crisis intervention unit (both groups) | Sample Size: 170.<br><br>Demographics: 84.1% female; mean age 31.9 (SD= 10.1); no ethnicity data provided.<br><br>Diagnoses: 1) DSM-IV MDD and 2) DSM-IV BPD. | Primary outcomes: suicidal relapse and hospitalisation. Secondary outcomes: overall mental health (GAS); symptom severity (CGI); depressive symptoms (HDRS); psychosocial outcomes (HSRS - modified version). | Primary outcomes: Suicidality- participants who received either form of AP had fewer episodes of suicidal relapse (AP-P vs. TAU: Pearson $\chi2=8.09$, df=1, p=.004; AP-N vs. TAU: Pearson $\chi2=9.33$, df=1, p=.002). They also had increased survival to suicidal crisis relapse compared to patients assigned to TAU (AP-P vs. TAU log-rank test: Mantel $\chi2=7.63$, df=1, p=.006; AP-N vs. TAU log-rank test: Mantel $\chi2=9.87$, df=1, p=.002). Those who received AP were also less likely to be hospitalised as an inpatient than those assigned to TAU (AP-P vs. Tau Pearson $\chi2=6.34$, df=1, p=.012; AP-N vs. TAU: Pearson $\chi2=6.34$, df=1, p=.012). Recipients of AP also showed significantly greater improvement on suicidal ideation, global functioning, symptom severity and depression diagnosis. Similar outcomes were reported for the two modes of delivery of AP (nurse and psychotherapists). |

| 5. Social-interpersonal and functional therapies vs. Non-active comparators<br>a. Randomised Controlled Trials | | | | | | |
|---|---|---|---|---|---|---|
| RCT. Non-specialist/inactive comparator. | Pascual et al. 2015 Spain | Evaluate the efficacy of a cognitive rehabilitation group therapy as compared to a psychoeducational group intervention in participants with BPD on psychosocial functioning. | Treatment: Cognitive rehabilitation group - Cognitive Rehabilitation (CR): consists of group sessions with exercises addressing neurocognitive issues related to sustained attention, processing speed, memory, and executive functioning. The whole programme aimed at getting new strategies to improve functional adaptation, thus tasks were carried out in the clinical setting and at home. Some homework tasks were based on their daily life difficulties and problems.<br><br>Duration/Intensity: 16-week programme; twice weekly group sessions (120 minutes).<br><br>Comparator: The psychoeducational intervention consisted of 16 weekly group sessions. This therapy aimed at improving awareness of illness, interpersonal abilities, family balance, therapeutical adherence, emotional management in frustrating situations, problem solving, and lifestyle regularity. During this intervention, no homework tasks were required.<br><br>Service setting: Standalone outpatient intervention | Sample Size: 70.<br><br>Demographics: 74% female; mean age 32.4 (range 18 to 45); no ethnicity data provided.<br><br>Diagnoses: DSM-IV-TR BPD diagnosis (SCID-II; DIB-R). | Primary outcome: Functioning (FAST). Secondary outcomes: BPD symptom severity (CGI-BPD; BSL-23); general functioning (GAF); anxiety symptoms (HARS); depressive symptom severity (MADRS); impulsiveness (BIS); neuropsychological assessment (neuropsychological battery). | The primary outcome of the trial (FAST) did not show main effects of group, group x time or number of sessions. There was a significant main effect of time for both treatments (baseline, post-treatment, and 6-month follow-up assessments) [F(2, 40.04)=6.34, p=.004]. Posthoc analyses showed that CR was the intervention which showed greater improvement in the FAST (p=.018). Secondary outcomes: The psychoeducational intervention showed a significant enhancement of depressive symptoms and attention functioning. Results suggest that the general functional improvement observed in BPD is independent of clinical and neuropsychological changes. |

| | | | | | | |
|---|---|---|---|---|---|---|
| RCT. Non-specialist/inactive comparator. | Huband et al. 2007 UK | To determine the effectiveness of a problem-solving intervention for adults with personality disorder in the community under conditions resembling routine clinical practice. | Treatment: Brief psychoeducation plus problem-solving therapy: Group problem-solving therapy aims to improve social competence by teaching how to discover solutions to problems in living and individual psychoeducation about personality disorder and the nature of their own diagnosis.<br><br>Duration/Intensity: Programme length unclear; 3 individual psychoeducation sessions (60 minutes) followed by 16 weekly problem-solving groups (120 minutes). Additional fortnightly or less frequent support sessions on request.<br><br>Comparator: Waitlist<br><br>Service setting: Standalone outpatient intervention | Sample Size: 176.<br><br>Demographics: intervention group 48% male, control group 49% male; mean age intervention 36.2 (SD=9.69), control 36.2 (SD=9.31); no ethnicity data provided.<br><br>Diagnoses: At least one DSM–IV PD | Primary outcomes: Social problem-solving ability (SPSI-R); social functioning (SFQ). Secondary outcomes: Anger (STAXI-2); impulsiveness (BIS); shame (ESS); dissociation (DES). | Primary outcomes: Those in the intervention group had significantly better social problem-solving skills (d=0.56, p<.001, 95% CI 1.21, 2.97) and higher overall social functioning (d=-0.25, p=.03, 95% CI -1.99, -0.18) at end point (mean=24 weeks after randomisation). Secondary outcomes: No significant changes across groups, except lower anger expression in the intervention group at end point. |
| RCT. Non-specialist/inactive comparator. | Munroe-Blum et al. 1995 Canada | To compare short-term manualized interpersonal group psychotherapy with individual open-ended psychodynamic psychotherapy for people with borderline personality disorder. | Treatment: Interpersonal group psychotherapy - manual guided group using interpersonal psychotherapy<br><br>Duration/Intensity: Programme length unclear; 25 weekly sessions (90 minutes) + 5 twice a week sessions (90 minutes).<br><br>Comparator: TAU - Individual dynamic psychotherapy: The comparison treatment model, individual dynamic psychotherapy consisted of open-ended, individual, dynamic psychotherapy. Although this is a "treatment-as-usual" comparison, there were none the less several controls.<br><br>Service setting: Standalone outpatient intervention | Sample Size: 110.<br><br>Demographics: 81% female, age range 18 to 52 years, no ethnicity data provided.<br><br>Diagnoses: BPD diagnosis (DIB) | No primary outcome specified. Social dysfunctional behaviours (OBI); social functioning (SAS); depressive symptoms (BDI); symptomatology (HSCL-90). | No primary outcome specified. No statistically significant differences in outcome variables between experimental group treatment and individual TAU control at end of treatment and 12-month follow-up, though both groups resulted in statistically significant improvements for all outcomes at end of treatment and follow-up. Lower costs and thus greater potential cost-effectiveness were reported for the experimental group therapy. |
| 6. Social-interpersonal and functional therapies vs. Active comparators<br>a. Randomised Controlled Trials | | | | | | |

| | | | | | | |
|---|---|---|---|---|---|---|
| Pilot RCT (Proof of concept feasibility trial with stepped-wedge design (randomised). Specialist/active comparator. | Di Simplicio et al. 2020 UK | (1) To assess the feasibility of recruitment and delivery of a brief psychological intervention, FIT, to reduce self-harm in a community sample of young people aged 16–25, and (2) to investigate effects at 3 and 6 months on the self-harm frequency, self-harm severity, and self-efficacy for control over self-harm, comparing usual care (UC) plus FIT that was delivered either immediately (Immediate FIT) or after 3 months (Delayed FIT). (3) to explore whether retention in therapy and change in the self-harm frequency after FIT were associated with participants' baseline characteristics. | Treatment: Functional Imagery Training (FIT) plus usual care (UC) (Immediate FIT)<br><br>Duration/Intensity: 8-week programme; 2 face to face sessions (90 minutes) + 5 phone support calls (15-30 minutes).<br><br>Comparator: Active comparator: Delayed FIT (where only UC was received over the initial 3 months)<br><br>Service setting: Standalone outpatient intervention | Sample Size: 38.<br><br>Demographics: 31/38 female; mean age 19.7 immediate FIT, 19.2 delayed FIT; ethnicities 36/38 White, 1/38 Asian, 1/10 mixed—White and Black Caribbean.<br><br>Diagnoses: At least two episodes of self-harm in the previous 3 months. | Primary outcome: Self-harm frequency and severity (1-item question). Secondary outcomes: Self-efficacy for control of self-harm (SEC); mental health services use (clinical records). | This was a feasibility trial not powered to find a significant effect. Primary outcome: A significant main effect of time was found for number of self-harm episodes but no statistically significant difference between treatment groups (time: F=5.36, p=.006, g2=0.11; time x intervention: F = 0.94, p=.40, g2=.022). Numbers of self-harm episodes, mental health service use and maximum severity all reduced over time in both groups, with no very severe episodes. |

| | | | | | | |
|---|---|---|---|---|---|---|
| RCT. Specialist/active comparator. | Stravynski et al. 1994 Canada | to test the efficacy of social skills training as developed for avoidant personality disorder, comparing sessions in the clinic only with sessions in real life settings. | Treatment: Social skills training (SST) in-vivo - four sessions held in the hospital and the last four sessions took place in real-life situations in the community (e.g., shopping centre, museum, restaurant). SST consisted of a sequence of behaviour modification techniques aimed at the development and the building up of pre-determined targeted skills.<br><br>Duration/Intensity: 8-week programme; weekly sessions (90 minutes) followed by a 6-month follow up of monthly sessions.<br><br>Comparator: active comparator (SST in the clinic/hospital only)<br><br>Service setting: Standalone outpatient intervention | Sample Size: 31.<br><br>Demographics: Intervention group 47.1% female, control group 35.7% female; mean age intervention 31 (range 18-59), mean age control 32 (range 18-55); no ethnicity data provided.<br><br>Diagnoses: DSM-III avoidant PD. | No primary outcome specified: Social target performance (diary); social avoidance and distress (SAD); social activity (SSQ); depression (BDI); anxiety (HARS); personality functioning (MMPI); anxiety (STAI); maladjustment (SSIAM); global performance and distress (behavioural assessment). | No primary outcome specified. No significant difference found between group receiving skills training in vivo and group in clinic only. Significant improvements found in both over the treatment period. |
| **7. Self-management and care planning vs. Self-management**<br>a. Randomised Controlled Trials | | | | | | |
| RCT. Self-management and care planning. | De Saeger et al. 2014 the Netherlands | To benchmark the efficacy of TA among patients with severe personality pathology awaiting an already assigned course of treatment. | Treatment: Therapeutic assessment (TA)<br><br>Duration/Intensity: Programme length unclear; 4 sessions.<br><br>Comparator: Active comparator: Structured goal-focused pre-treatment intervention (GFPTI)<br><br>Service setting: Specialist PD service (both groups on the waiting list for specialist PD service) | Sample Size: 74.<br><br>Demographics: 60.8% female; mean age 39 (SD=10.13); ethnicity 100% White.<br><br>Diagnoses: Any personality disorder. One or more PD diagnoses (55.5%): Avoidant (25.9%); PD NOS (16.7%); borderline (7.4%); obsessive–compulsive (5.6%) PD. | No primary outcome specified. Treatment readiness (AQ; EFTS); therapeutic alliance (HAq–II); demoralisation (RCdem); symptom severity (BSI); satisfaction with treatment (CSQ8). | No clearly identified primary outcome. TA resulted in higher expectancy for treatment outcome than did GFPTI, and there were also benefits for perception of personal progress and patients' (but not therapists') perception of working alliance and for overall satisfaction. No difference was found in demoralization or global symptom severity. |

| | | | | | | |
|---|---|---|---|---|---|---|
| RCT (pilot). Self-management and care planning. | Borschmann et al. 2013 UK | To examine the feasibility of recruiting and retaining adults with borderline personality disorder to a pilot RCT investigating the potential efficacy and cost-effectiveness of using a joint crisis plan. | Treatment: Enhanced CMHT / Joint Crisis Planning<br><br>Duration/Intensity: Single joint crisis planning meeting (60 minutes) + standard care.<br><br>Comparator: Participants in both groups continued to receive standard care from their treating CMHT. This included, as a part of the care programme approach (CPA), the provision for service users to receive written copies of their care plan, including a brief 'crisis contingency plan', in addition to regular contact with a care coordinator or allocated member of the clinical team.<br><br>Service setting: Generic community mental health teams | Sample Size: 88.<br><br>Demographics: 19.3% male; mean age 35.8 (SD= 11.6); ethnicities White 73.9%, Asian 1.1%, Black 10.2%, Mixed 8%, Other 6.8%.<br><br>Diagnoses: 1) DSM-IV BPD diagnosis (SCID-II); and 2) self-harm events in past 12 months | Primary outcome: Self-harming behaviour. Secondary outcomes: Depressive and anxiety symptoms (HADS); engagement and satisfaction with services (CSQ; SES); perceived coercion (TES); working alliance (WAI); quality of life (EQ-5D); social functioning (WSAS); well-being (WEMWBS); resource use (AD-SUS). | Primary outcome: this was a pilot study not powered to find an effect, but intention-to-treat analysis revealed no significant differences in the proportion of participants who reported self-harming (odds ratio (OR=1.9, 95% CI 0.53, 6.5, p=.33) nor were significant differences reported in frequency of self-harming behaviour or any other secondary outcome measures or costs. |
| 8. Self-management and care planning vs. different ways of organising services<br>a. Randomised Controlled Trials | | | | | | |
| RCT. Different ways of organising services [case management]. | Ranger et al. 2009 UK | To assess the effectiveness of nidotherapy added to assertive outreach in a group of people with severe mental illness and a comorbid personality disturbance. | Treatment: Nidotherapy enhanced assertive outreach - collaborative treatment involving the systematic assessment and modification of the environment to minimise the impact of any form of mental disorder on the individual or on society involved a combination of environmental analysis, articulation of the patient's needs at a physical, social, and personal environmental level, and setting of targets.<br><br>Duration/Intensity: 12-month programme; up to 15 sessions.<br><br>Comparator: Control group TAU (standard assertive outreach)<br><br>Service setting: Generic assertive outreach team (intensive community mental health care) - both arms | Sample Size: 52.<br><br>Demographics: 17/52 female; no additional demographic provided.<br><br>Diagnoses: 1) Severe mental illness; and 2) comorbid ICD-10 PD or personality difficulty (PAS-I). | Primary outcome: Duration of psychiatric admissions. Secondary outcomes: Service costs; clinical symptomatology (BPRS); social functioning (SFQ-KW); service costs (SFSUS). | Primary outcome: There was no difference between the treatment groups after 18 months in number of admissions (p=.91) or duration of bed use (p=.258). Secondary outcomes: Clinical symptoms, social functioning and engagement all showed somewhat greater improvement in the Nidotherapy enhanced (Active) group but this was small and not significant. |
| 8. Self-management and care planning vs. different ways of organising services<br>b. Non-randomised experiments and observational studies | | | | | | |

| | | | | | | |
|---|---|---|---|---|---|---|
| Observational study with pre-post comparison. Different ways of organising services [case management]. | Graham et al. 2019 UK | To assess the impact of establishing a specialized community personality disorder team on out of area placements, local hospital admissions and out of hours crisis contacts for service users with borderline personality disorder. | Treatment: Specialist PD case management team aiming to resettle people in local area who were previously residing in Out of Area specialist inpatient and residential placements, and to support them through an intensive case management approach.<br><br>Duration/Intensity: 12-month programme before evaluation.<br><br>Comparator: N/A – pre-post<br><br>Service setting: Specialist PD team | Sample Size: 37.<br><br>Demographics: 35/37 female; mean age 30; no ethnicity data provided.<br><br>Diagnoses: PD diagnosis. | No primary outcome specified. Service use (unclear). | No primary outcome specified. All service users in out of area placements were repatriated to live in the community locally (100%), and there was a statistically significant decrease in inpatient admissions (80%), as well as in bed days in hospital as well as a smaller but statistically significant increase in out of hours community crisis contacts. |
| Observational study (including a contemporaneous comparison). Different ways of organising services [case management]. | Solberg et al. 2018 USA | To investigate whether Collaborative Case Management is more effective than usual primary care in management of major depressive disorder among people who also meet criteria for a PD. | Treatment: Collaborative care management (CCM)- an approach to people identified as having depression in primary care which involves allocation of a care manager and regular review by a psychiatrist - the aim is to manage patients' care and ensure evidence-based guidelines are followed.<br><br>Duration/Intensity: Not recorded.<br><br>Comparator: TAU usual US primary care for depression without the allocation of a care manager.<br><br>Service setting: US Primary care services for depression | Sample Size: 9614.<br><br>Demographics: 72% female; age≥18; no ethnicity data provided.<br><br>Diagnoses: 1) Primary care patients with clinical diagnosis of MDD; and 2) a PHQ-9 score ≥10; with and without PD diagnosis. | Primary outcome: Remission of depression (PHQ-9 score <5); persistent depressive symptoms (PHQ-9 score ≥10). | Primary outcome: Rate of remission of depression in the context of a PD was lower by 11.5% for people receiving Usual Care than Collaborative Clinical Management 6 months after diagnosis (Odds of remission adjusted for baseline variables 0.369 (95% CI 0.201, 0.676, p=.001). Criteria for persistent depressive symptoms were also more likely still to be met in the UC group. |

| | | | | | | |
|---|---|---|---|---|---|---|
| Quasi-experiment with contemporaneous comparisons. Different ways of organising services [case management]. | Stringer et al. 2013 Netherlands | To describe the feasibility of a collaborative care programme delivered to people with BPD who are not currently able to engage in psychotherapy and make a preliminary assessment of its outcomes compared with usual care. | Treatment: Collaborative care programme (CCP), which aims "to increase shared decision making and enhancement of self-management skills of chronically ill patients and optimize continuity and coordination of care"; Nurses function as collaborative care managers, and thus are responsible for both proper implementation and optimal organisation of treatment. CCP "was developed to improve quality of care for patients with severe borderline or NOS personality disorders within a CMHC setting". Framework established for collaborative care planning with patients and carers for people not currently able to engage in psychotherapy, including psychoeducation and problem-solving elements.<br><br>Duration/Intensity: 9-month programme.<br><br>Comparator: Treatment as usual<br><br>Service setting: Generic community mental health teams | Sample Size: 30.<br><br>Demographics: intervention group 93.8% female, control group 80% female; mean age 42.9 (SD=11.7), mean age control 44.5 (SD=8.7). No ethnicity data provided.<br><br>Diagnoses: Main DSM-IV-TR BPD or PD NOS diagnosis; 2) score of 15≤ on BPDSI; and 3) received psychiatric care for at least two years. Main diagnosis: BPD (intervention: 75.0%; control: 10%); and PD NOS (intervention: 25%; control: 30%).koon | Primary outcomes: Quality of life (MANSA); BPD symptom severity (BPDSI). | Pilot study not powered to detect significant differences. Reduction in severity of borderline symptoms was significantly greater in the treatment than the control group, with 50% of cases falling below the cut off for a BPD diagnosis at the end of follow-up. Number of contacts with mental health services also reduced. Changes on other outcomes were not significant. |
| 9. Novel mental health service model vs Day hospital<br>a. Randomised Controlled Trials | | | | | | |

| | | | | | | |
|---|---|---|---|---|---|---|
| RCT.<br>Day hospital vs outpatient comparator. | Antonsen et al. 2017 (overlapping sample Arnevik et al. 2009). Norway | To compare over 6-year period outcomes for an experimental condition involving day hospital followed by outpatient care with an outpatient therapy programme among a sub-sample of trial participants with a BPD diagnosis. | Treatment: Specialist Day hospital followed by outpatient therapy - mixed modes of therapy (mixed: CBT & psychodynamic)<br><br>Duration/Intensity: Intervention: initial 18-week programme; 3-4 days a week. Followed by a) a 4-year programme; weekly group therapy of 1.5-hour duration and b) a 2.5-year programme of weekly individual therapy. Average treatment duration for BPD patients was 28 months (SD=16) and average number of treatment session was 94 (SD=81).<br><br>Comparator: Average treatment duration was 24 months (SD=21) and average number of therapy sessions was 60 (SD=66).<br><br>Comparator: Outpatient individual psychotherapy (OIP)<br><br>Service setting: Specialist Day hospital (experimental group) followed by outpatient intervention compared with standalone outpatient intervention only | Sample Size: 52.<br><br>Demographics: 85% female; mean age 29 (SD=6.7); no ethnicity data provided.<br><br>Diagnoses: BPD diagnosis; excluding schizotypal or antisocial PD. Comorbid PDs: avoidant (33%); paranoid (15%); obsessive-compulsive (12%); dependent (10%); and narcissistic (2%) PD. | No primary outcome specified: Symptom severity (SCL-90-R GSI); depressive symptoms (BDI); global functioning (GAF); social and occupational functioning (WSAS); quality of life (QoL scale); interpersonal problems (CIP); personality functioning (SIPP-118). Secondary outcomes: diagnostic status (SCID-II); self-harm, suicidal thoughts, and suicide attempts (self-report). | No primary outcome specified. The group receiving initial day hospital treatment showed significantly greater reductions of symptom distress, greater improvements in self-control and identity across the 6 years, as well as a greater improvement in psychosocial functioning between 3- and 6-year follow-up. No differences were found in interpersonal functioning, depression, quality of life, or self-harm and suicidal thoughts. Only 10% in the step-down group and 7% in the outpatient group met the diagnostic criteria for BPD at six-year follow-up, with no between-group differences. |

| | | | | | | |
|---|---|---|---|---|---|---|
| RCT.<br>Day hospital vs outpatient comparator. | Antonsen et al. 2014 (FU to Arnevik et al 2009) Norway | To compare over a 6-year period outcomes from an experimental group receiving initial step-down day hospital programme with a control group receiving an outpatient therapy programme. | Treatment: Step-down day hospital (mixed: CBT & psychodynamic)<br><br>Duration/Intensity:<br>Intervention: initial 18-week programme; 3-4 days a week. Followed by a) a 4-year programme; weekly group therapy of 1.5-hour duration and b) a 2.5-year programme of weekly individual therapy.<br>Comparator: average duration was 24 months (SD=20), and the average number of consultations was 56 (SD=56.7).<br><br>Comparator: Outpatient individual psychotherapy (OIP)<br><br>Service setting: Specialist Day hospital (experimental group) compared with standalone outpatient intervention) | Sample Size: 113.<br><br>Demographics: 75% female; mean age 31 (SD=7.3); no ethnicity data provided.<br><br>Diagnoses: Diagnosis of PD other than antisocial. BPD (46%) and avoidant (41%). | Primary outcomes: Symptom distress (SCL-90-R); depressive symptoms (BDI); general functioning (GAF); social and occupational functioning (WSAS); quality of life (QoL), interpersonal problems (CIP); personality functioning (SIPP-118). Secondary outcomes: PD diagnostic status (SCID-II); self-harm, suicidal thoughts, and suicide attempts (self-report). | Primary outcomes: There were no statistically significant differences between groups at the 6-year follow-up for the primary outcome variables (GAF (p=.52), WSAS (p=.47), CIP (p=.96), GSI (p=.38), BDI (p=.47), and QoL (p=.27). Both groups improved on all outcome variables between baseline and 6 years. A significantly greater improvement in psychosocial functioning (GAF: p<.0001; WSAS: p=.001) between 3 and 6 years was reported for the treatment group. Regarding secondary measures, there was no significant differences in numbers of diagnoses, self-harm and suicide attempts or suicidality. |
| RCT.<br>Day hospital vs outpatient comparator. | Gullestad et al. 2012 (FU to Arnevik et al 2009) Norway | To compare the two treatments modalities on a wide range of clinical measures, including symptom distress, interpersonal functioning, psychosocial functioning, quality of life, and axis I and II diagnoses. | Treatment: Step-down day hospital (mixed: CBT & psychodynamic)<br><br>Duration/Intensity: 48-month programme; 18-week hospital treatment with 4 times a week group therapy followed by weekly group therapy (90 minutes) up to 48 months + weekly individual therapy up to 30 months.<br><br>Comparator: Treatment as usual (outpatient individual psychotherapy)<br><br>Service setting: Specialist Day hospital compared with standalone outpatient intervention | Sample Size: 113.<br><br>Demographics: 75% female; mean age: 31 (SD=7.3); no ethnicity data reported.<br><br>Diagnoses: PD diagnosis | Primary outcomes: Symptom severity (SCL-90); depressive symptoms (BDI); interpersonal problems (CIP); global functioning (GAF); social and occupational functioning (WSAS); subjective quality of life. Secondary outcomes: Self-harm, suicidal thoughts, and suicide attempts (self-report). | Multiple primary outcomes specified. At 37 month follow up, a statistically significant interaction was found between group and time 8-36 months on GAF (estimate: 0.34, SE=0.096, 95% CI 0.15, 0.53, p<.001), suggesting patients had improved more in the outpatient than the step-down treatment. Statistically significant differences between groups were not found on other primary and secondary outcome measures. |

| | | | | | | |
|---|---|---|---|---|---|---|
| RCT.<br>Day hospital vs outpatient comparator. | Arnevik et al. 2010 (FU to Arnevik et al 2009) Norway | To compare outcomes from an experimental group receiving initial step-down day hospital programme with a control group receiving an outpatient therapy programme over an 18-month period. | Treatment: Specialist Day hospital (mixed: CBT & psychodynamic) followed by outpatient<br><br>Duration/Intensity:<br>Step-down: 18 weeks short-term day hospital, group therapies 3-4 days/week.<br>Outpatient individual psychotherapy (max 2.5 years) and weekly 1.5h group psychotherapy (max 4 years).<br>Comparator: Mean number of sessions received at 18-month follow-up was 29 (SD=11) for individual and 21 (SD=11) for group therapy.<br><br>Comparator: Outpatient individual psychotherapy (OIP)<br><br>Service setting: Specialist Day hospital (experimental group) compared with standalone outpatient intervention. | Sample Size: 114.<br><br>Demographics: 74% female; mean age 31 (SD=7.4); no ethnicity data provided.<br><br>Diagnoses: DSM-IV PD diagnosis, other than antisocial and schizotypal personality disorder. Borderline (46%); avoidant (40%); PD NOS (21%); paranoid (15%); obsessive-compulsive (9%); dependent (7%); narcissistic (2%); and schizoid (1%) PD. | No primary outcome specified. Self-injury, suicidal thoughts, and suicide attempts (self-report); symptom severity (SCL-90-R); depressive symptoms (BDI); hopelessness (BSH); quality of life (10-point scale); interpersonal problems (CIP); global functioning (GAF); personality functioning (SIPP-118); self-esteem (ISE). | No primary outcome specified. Self-injuries, suicidal thoughts, and suicide attempts declined in both groups at 8- and 18-months follow-up (numbers of incidents were reported to be too small to test for significance). The outpatient group showed significantly greater improvements in self-esteem and interpersonal problems. Both groups tended to improve from baseline on most other outcomes, with no between-group differences (no test of statistical significance reported). |
| RCT.<br>Day hospital vs outpatient comparator. | Arnevik et al. 2009 Norway | To compare outcomes from an experimental group receiving initial step-down day hospital programme with a control group receiving an outpatient therapy programme over an 8-month period. | Treatment: Step-down day hospital (mixed: CBT & psychodynamic)<br><br>Duration/Intensity: Step-down: 18 weeks day hospital psychotherapy (DHP), followed by weekly outpatient and individual and group psychotherapy.<br><br>Comparator: Mean duration of received therapy at 8-month follow-up was 4.5 months (SD=2.6).<br><br>Comparator: Outpatient individual psychotherapy (OIP). The mean duration of received therapy at follow-up was 4.5 months (SD=2.6)<br><br>Service setting: Specialist day hospital (experimental group) compared with standalone outpatient intervention | Sample Size: 114.<br><br>Demographics: 74% female; mean age 31 years (SD=7.4); no ethnicity data provided.<br><br>Diagnoses: DSM-IV PD diagnosis other than antisocial and schizotypal personality disorder. Borderline (46%); avoidant (40%); PD NOS (21%); paranoid (15%); obsessive-compulsive (9%); dependent (7%); narcissistic (2%); and schizoid (1%) PD. | No primary outcomes specified. Self-injury, suicidal thoughts, and suicide attempts (self-report); symptom distress (SCL-90-R); depressive symptoms (BDI); hopelessness (BSH); quality of life (10-point scale); interpersonal problems (CIP); general functioning (GAF); personality functioning (SIPP-118). | No primary outcome specified. There were no significant between-group differences on outcomes including suicidal ideation and attempts, symptoms, and social functioning over 18 months. Both groups improved on clinical measures from baseline (F=6.80, p<.001), with no between-group differences (F=0.43, p=.83). |

10. Novel mental health service model vs established generic or specialist mental health services

a. Randomised Controlled Trials

| | | | | | | |
|---|---|---|---|---|---|---|
| Cluster RCT (two clusters). Established generic or specialist mental health services [stepped care]. | Grenyer et al. 2018 Australia | To examine whether implementing a stepped care model of psychological therapy reduces demand on hospital units by people with personality disorder, in a cluster randomized controlled trial. | Treatment: Stepped care psychological therapy - service wide<br><br>Duration/Intensity: Up to 37-month programme; initial triage to stepped care followed by 1 month of weekly contact followed by up to 36 months standard care.<br><br>Comparator: Treatment as usual (TAU)<br><br>Service setting: Community mental health team or outpatient clinician (sometimes with substantial waiting lists) | Sample Size: 642.<br><br>Demographics: 46% intervention group female, 55.4% TAU group female; mean age 36.85 (SD = 13.11); no ethnicity data provided.<br><br>Diagnoses: ICD-10 PD diagnosis. | Primary outcomes: Number and length of inpatient stays; number of emergency department presentations. | Primary outcomes: An interaction was found between time and study site for total bed days ($F(1,640)=4.301, p=.038$), suggesting an effect from the intervention in reducing bed days. Patients in the intervention site were also reported to be 1.28 times more likely (95% CI= 1.17, 1.40; $\chi 2= 19.980, p=.000$) to have a reduction in A & E attendance from baseline than control site participants. Direct cost savings for implementing the approach was estimated at USD$ 2,720 per patient per year. |

| | | | | | | |
|---|---|---|---|---|---|---|
| RCT. Established generic or specialist mental health services [therapeutic community]. | Pearce et al. 2017 UK | To obtain randomised controlled trial regarding the effectiveness of democratic therapeutic communities in treating personality disorder. | Treatment: Democratic therapeutic community - Attendance at a DTC prep group meeting for up to a year. After a minimum of 3 months' attendance participants able to join the DTC via democratic selection process (members and staff vote). DTC treatment consists of structured and unstructured group therapy, following 1) democratisation (shared decision-making), 2) permissiveness (range of behaviour tolerated, 3) reality confrontation (members challenge and feedback to one another around behaviour), 4) communalism (shared living), 5) a culture of enquiry (questioning events is encourage), 6) milieu approach (all activities therapeutic)<br><br>Duration/Intensity: 3–12-month preparatory programme; weekly group meetings (120 minutes) followed by up to 18-month programme; 5-15 hours of mixed group therapy a week.<br><br>Comparator: Treatment as usual (TAU) – TAU participants offered three sessions of joint crisis planning by clinician. Other elements of TAU varied by patient needs and were delivered by non-specialist services<br><br>Service setting: Specialist service (therapeutic community) for treatment group, generic services for control | Sample Size: 121.<br><br>Demographics: 72.7% female; mean age 32.91 (SD=10.17); ethnicities 94.2 White, 3.3% White other, 2.5% Black and ethnic minority.<br><br>Diagnoses: PD diagnosis (SCID-II). | Primary outcome: Days of inpatient psychiatric treatment. Secondary outcomes: General health (GHQ); social functioning (SFQ); self-harm or aggressive behaviour (MOAS); treatment satisfaction (CSQ); suicidal acts and acts of self-harm (self-report); service use (self-report). | Primary outcome: Although fewer people in the active intervention arm had an admission to hospital 12 months after randomisation, numbers of admissions were low overall and the difference was not statistically significant (difference 11.4%, 95% CI –10.1, 31.6%). Secondary outcomes: DTC showed significant advantages over TAU in aggression and self-harm measured by the Modified Overt Aggression Scale, and satisfaction with treatment, measured by the Client Satisfaction Questionnaire. There were no significant differences in other outcomes between those randomised to DTC and TAU. |

#### 10. Novel mental health service model vs established generic or specialist mental health services

b. Non-randomised experiments and observational studies

| | | | | | | |
|---|---|---|---|---|---|---|
| Observational study. Established generic or specialist mental health services [stepped care]. | Huxley et al. 2019 Australia | To examine the effectiveness of the intervention in reducing individual mental health symptoms and improving quality of life. | Treatment: Integrated brief intervention (Stepped Care Service Wide Model; same treatment as Grenyer et al 2018): brief intervention delivered immediately after a period of acute care, followed by referrals and escalations in care determined using clinical judgement of clinicians and consultation with treatment team.<br><br>Duration/Intensity: Programme length unclear; 4 weekly sessions (50 minutes).<br><br>Comparator: N/A - observational design<br><br>Service setting: Brief intervention clinic introduced to community mental health service pathway as part of stepped care approach | Sample Size: 67.<br><br>Demographics: 75.39% female; mean age 31.54 (SD=13.40); no ethnicity data provided.<br><br>Diagnoses: PD diagnosis. | No specified primary outcome. Distress (MHI-5); BPD symptom severity and deliberate self-harm/suicide (MSI-BPD); suicidal ideation and quality of life (one item). | No specified primary outcome. Uncontrolled study examining change over time. At end of treatment all outcomes saw significant improvements over the treatment period, including total DSM-V symptoms, distress (MHI-5), quality of life, BPD symptoms and suicidal ideation. An accompanying study in the same paper describes treatment pathways for a larger cohort. |
| Observational study with contemporaneous comparison - choice of treatment based on clinical judgement. Established generic or specialist mental health services [stepped care]. | Laporte et al. 2018 Canada | To examine the clinical outcomes of treatment in a stepped care model including a short-term therapy clinic, comparing them with treatment in a clinic offering extended care. | Treatment: Short term stepped care model - initial referral is to rapid treatment in this setting with individual therapy and group therapy modalities. Group sessions make use of psychoeducation and group process to develop better emotion regulation, better interpersonal skills, and decreased impulsivity. Similar principles can be used in individual therapy. Short term treatment over 12 weeks, followed by referral to extended treatment clinic if indicated.<br><br>Duration/Intensity: 12-week programme; weekly individual and group therapy.<br><br>Comparator: Extended clinic. 6-month blocks, with a maximum of 2 years, de-pending on patients' progress. Similar therapies provided to short term stepped care model.<br><br>Service setting: Standalone outpatient therapies | Sample Size: 615.<br><br>Demographics: Short term model 92% female, comparison 89% female; mean age 27.1, (SD=7.8), comparison 36.1 (SD=10.4); ethnicity data not provided.<br><br>Diagnoses: BPD diagnosis (Short term model: 100%; Extended care model: 86%) | No primary outcome specified. Impulsivity (BIS-II); self-esteem (SES); depressive symptoms (BDI); emotional regulation (DERS); symptom severity (SCL-90-R); alcohol and drug use (ASI); self-harm and suicide attempts (SHBQ). | In both treatment groups, main comparisons were with baseline in the same treatment conditions rather than between ST and EC clinics. In both settings there were significant reductions in all symptoms over the treatment period, with the exception of drug and alcohol misuse which reduced significantly only in the Extended Clinic. In the Short-Term Clinic, the rate of premature termination (29%) was similar to many other studies. The number of dropout or early discharge was higher in the Extended Clinic. The authors conclude that substantial gains are made by many through a short-term clinic programme. |

| | | | | | | |
|---|---|---|---|---|---|---|
| Natural experiment with pre-post comparison. Established generic or specialist mental health services [therapeutic community]. | Barr et al. 2010 UK | The study aimed to clarify whether one-day therapeutic communities can be effective for people with personality disorder. | Treatment: One day a week therapeutic communities<br><br>Duration/Intensity: 12-month programme; weekly therapeutic communities.<br><br>Comparator: N/A – pre-post<br><br>Service setting: Specialist PD service (therapeutic community) | Sample Size: 20.<br><br>Demographics: 17/20 (85%) female; mean age 35.15 (SD=12.13); no ethnicity data provided.<br><br>Diagnoses: PD diagnosis. Avoidant PD (100%), depressive PD (95%), schizotypal and obsessive-compulsive PD (both 74%), paranoid PD (68%), borderline and negativistic PD (both 63%). | No primary outcome specified. Personality diagnosis (PDQ-4); clinical severity (CORE-OM; TAG); social functioning (SFQ); self-harm (DSHI); BPD symptoms (ZAN-BPD); emergency hospital attendances (SUSI); cost-offset (calculated by the Personal Social Services Research Unit). | No primary outcome specified. Over the course of treatment, significant improvements were observed in ratings of symptoms (CORE-OM) social functioning (SFQ) and risk/severity (TAG), but differences did not reach statistical significance on self-harm, service use or other included measures. |
| Observational study with pre-post comparisons. Established generic or specialist mental health services [support groups]. | Miller and Crawford 2010 United Kingdom | To describe a new open access community service for people with personality disorder and to explore interim service utilisation and outcomes. | Treatment: Peer support network (SUN) - community-based open access support groups for people with personality disorder. It aimed to help people develop effective ways of coping, reduce emergencies and improve access to appropriate service<br><br>Duration/Intensity: 8-month programme; 3-4 times weekly group sessions (150 minutes).<br><br>Comparator: N/A - pre-post<br><br>Service setting: Standalone open access programme receiving referrals from a range of sources | Sample Size: 171.<br><br>Demographics: 67.8% female; mean age 39.7 (SD=9.6); ethnicities 63.7% White British, 7.6% White other, 5.8% Black and ethnic minorities.<br><br>Diagnoses: "Probable personality disorder" (90%). | No primary outcome specified. Personality status (SAPAS); social functioning (SFQ); service use (records); treatment satisfaction and service impact (10-item questionnaire). | Uncontrolled study in which comparisons are over time, no primary outcome specified. Decreased levels of contact with health and social care services in the period after joining the SUN project were reported and this was particularly marked for 'unplanned' contacts with services and use of in-patient treatment. Higher levels of social functioning in the 6 months following contact with the service than in the prior 6 months were reported, the reduction in SFQ score being both clinically significant and larger than that reported in previous studies of out-patient psychological treatment for people with personality disorder. Good ratings were obtained for satisfaction. 44.3% of questionnaire respondents had left the service, citing other commitments, difficulties with group members, and unhelpfulness of the service as reasons. |

| | | | | | | |
|---|---|---|---|---|---|---|
| Observational study. Established generic or specialist mental health services [specialist PD team]. | Pretorius et al. 2010 UK | To conduct a naturalistic assessment of the acceptability and outcomes of a of specialist community treatment programme for patients with personality disorder, based on a flexible and responsive team approach. | Treatment: The Coventry Community Specialist Personality Disorder Service - a tertiary specialist team with the aim of using the structure of a clinical psychiatric service, designed around the assertive outreach recovery model, to deliver therapy in a variety of settings to individuals with a primary diagnosis of personality disorder. The Coventry service takes the stance that the quality of the therapeutic alliance is one of the most important factors for successful outcome of psychotherapy and that a consistent, structured, focused, non-collusive approach based on the principles of attachment and the recovery model can be containing and therapeutic.<br><br>Duration/Intensity: Not recorded.<br><br>Comparator: N/A<br><br>Service setting: Specialist team (assertive outreach principles) | Sample Size: 183.<br><br>Demographics: age range 18-65; additional demographic characteristics not reported.<br><br>Diagnoses: Primary PD diagnosis. | No primary outcome specified. Service use; personality status (PDQ-4; SAPAS); global distress (CORE); social avoidance and distress (SADS); social functioning (SFQ). | Uncontrolled measurement of changes over time, no primary outcome specified. For a cohort for whom data were available (95 clients), an 82% reduction in days in hospital for the 3 years after being accepted by the service compared with the 3 years before (p=.000). Small but statistically significant improvement over time in self-rated outcomes for a sub-sample completing measures. |
| 10. Novel mental health service model vs established generic or specialist mental health services<br>c. Uncontrolled intervention development studies | | | | | | |

| | | | | | | |
|---|---|---|---|---|---|---|
| Intervention development and uncontrolled preliminary testing. Established generic or specialist mental health services [therapeutic community]. | Scott et al. 2010 UK | Evaluate a pilot mini therapeutic community service for older adults diagnosable with personality disorder. | Treatment: Abingdon Group Therapeutic treatment model treatment group - Facilitated within a democratic mini therapeutic community (TC) framework (Pearce & Haigh, 2008). Its ethos is one of recovery and is underpinned by the premise that PD is treatable, and, with appropriate psychotherapeutic interventions, the associated morbidity can be reduced to such an extent that people can resume a functional and rewarding life. The specific psychotherapy model is based on the integrative programme of diagnosis and treatment in the Wallingford Group. A weekly three-hour integrative large group based on democratic TC principles with a maximum of 14 members.<br><br>Duration/Intensity: 12-month programme; weekly group session (180 minutes); additional access to support beyond this.<br><br>Comparator: N/A<br><br>Service setting: Mental health services for older adult | Sample Size: 9.<br><br>Demographics: Older adults (65+), no gender or age data available.<br><br>Diagnoses: PD diagnosis. | No primary outcome specified. PD diagnosis (SAPAS); service use (SUSI); social functioning (SFQ); self-harm (SHI); distress (MHI-5). | Uncontrolled study with no statistical analysis as numbers are very small. Of 23 referrals, 9 entered and 4 completed this group. Some evidence is suggested that for those who participated, costs of treatment were reduced over a year. Statistical analyses were not carried out due to small numbers. |
